# *Trans*-eQTLs reveal the architecture of human gene regulatory networks

**DOI:** 10.64898/2026.02.04.26343575

**Authors:** C.A. Robert Warmerdam, Harm-Jan Westra, Adriaan van der Graaf, Marc Jan Bonder, Patrick Deelen, Tijs van Lieshout, Anke Jannie Landman, Mihkel Jesse, Benjamin J. Strober, Toni Boltz, Sandra Lapinska, Sini Nagpal, Manke Xie, Darwin Tay, Holger Kirsten, Haroon Naeem, Venket Raghavan, Aiman Farzeen, Alexander Teumer, Marie-Julie Favé, Elodie Persyn, Alex Tokolyi, René Pool, Jouke Jan Hottenga, Ruth D. Rodríguez, María Rivas-Torrubia, Binisha Hamal Mishra, Brandon Pierce, Lin Tong, Qingbo S. Wang, Takanori Hasegawa, Surya B. Chhetri, Diptavo Dutta, Stefan Weiss, Théo Dupuis, Leo-Pekka Lyytikäinen, Pashupati P. Mishra, Andrew R. Wood, Katie L. Burnham, Jia Wen, Evans Cheruiyot, Collins K. Boahen, Rick Jansen, Knut Krohn, Joachim Thiery, IMI DIRECT Consortium, Kensuke Daida, J. Raphael Gibbs, Joost Verlouw, Viktorija Kukushkina, Reedik Mägi, John Budde, Matt Johnson, Jessie Sanford, Eline Slagboom, Frank Beutner, Markus Loeffler, PRECISESADS Clinical Consortium, Javier Martin, Emma Raitoharju, Muhammad G. Kibriya, Farzana Jasmine, Matthias Nauck, The HELIOS Study Team, Juan Carlos Souto, Angel Martinez-Perez, M. Arfan Ikram, Juha Mykkänen, Katja Pahkala, Suvi P. Rovio, Laura M. Raffield, Marleen van Greevenbroek, Carla J.H van der Kallen, Casper G. Schalkwijk, Jan Veldink, Krista Freimann, Martijn Vochteloo, Anoek Kooijmans, Lilia Ouadah, Annique Claringbould, Yunfeng Huang, Julien Bryois, Ellen A. Tsai, Matthias Heinig, Monique G.P. van der Wijst, Estonian Biobank Research Team, sc-eQTLGen Consortium, Vinod Kumar, Peter M. Visscher, Allan F. McRae, Grant W. Montgomery, Yun Li, Julian C. Knight, Andrew Singleton, Luigi Ferrucci, Timothy M. Frayling, Andrew Brown, José Manuel Soria, Uwe Völker, Alexis Battle, Yukinori Okada, Koichi Fukunaga, Ho Namkoong, Habibul Ahsan, Terho Lehtimäki, Mika Kähönen, Olli T. Raitakari, Marta E. Alarcón-Riquelme, Guillermo Barturen, Brenda W.J.H Penninx, Dirk S. Paul, Michael Inouye, Philip Awadalla, Yun Ju Sung, Carlos Cruchaga, Joyce van Meurs, Holger Prokisch, Annette Peters, Christian Gieger, Cornelis Blauwendraat, Younes Mokrab, Emma E. Davenport, Markus Scholz, Marie Loh, John Chambers, Greg Gibson, Dorret I. Boomsma, Ana Viñuela, Bogdan Pasaniuc, Roel Ophoff, Alkes L. Price, Kaur Alasoo, Zoltan Kutalik, Tõnu Esko, Lude H. Franke, Urmo Võsa

## Abstract

Many non-coding variants influence complex traits and diseases through gene regulation, yet the mechanisms linking these variants to downstream biology remain poorly understood. Here, we present *eQTLGen Phase 2*, a comprehensive genome-wide analysis of gene expression quantitative trait loci (eQTLs) in 43,301 blood samples from 52 datasets. Beyond local *cis*-effects, this sample size enabled the first systematic mapping of *trans*-eQTLs at scale. We identify *cis*-eQTLs for nearly all expressed genes (94.7%) and *trans*-eQTLs for over half (56.2%). Second, by colocalizing *cis*-eQTLs with *trans*-eQTLs, we infer a directed gene regulatory network comprising 47,554 directed gene regulatory relationships. These networks reveal how genetic perturbations in upstream regulators produce dose-dependent downstream effects, supported by Perturb-seq and ChIP-seq data. Third, integrating this network with 87 genome-wide association studies allows us to systematically prioritize trait-relevant pathways and candidate genes. Variants exerting both *cis*– and *trans*-effects are markedly more likely to colocalize with trait associations than *cis*-only variants, delineating a subset of functionally active *cis*-eQTLs from a large group with limited downstream impact. This distinction provides a conceptual framework for identifying regulatory variants that truly mediate complex trait biology. Together, these results provide a publicly available resource of *cis*– and *trans*-eQTLs and an in vivo scaffold for human gene-regulatory networks, elucidating how propagation of *cis*-effects modulates complex disease.

## Introduction

Non-coding genetic variants play a central role in human disease by modulating gene regulation rather than protein sequence^1,2^. Expression quantitative trait locus (eQTL) studies systematically map these regulatory variants and link them to their target genes, thereby providing mechanistic insights into genome-wide association study (GWAS) signals^3–6^. To date, most eQTL studies have focused on *cis*-acting effects in which the genetic variant and gene are nearby. As *cis*-effects are relatively strong, they can be detected using modest sample sizes. However, most gene regulation occurs within interconnected networks where *trans*-acting variants propagate regulatory effects across distant genes and chromosomes^7^. Characterizing these *trans*-eQTLs is therefore crucial to understand how local regulatory changes translate into global transcriptional programs and complex trait variation^8–13^.

Despite their biological importance, *trans*-eQTLs have been difficult to study due to their small effect sizes. In our previous *eQTLGen* analysis of 31,684 blood samples^6^, we conducted genome-wide *cis*-eQTL mapping and targeted *trans*-eQTL analyses to 10,317 GWAS-associated variants. While this design revealed many disease-relevant regulatory mechanisms, it provided only a partial view of the *trans*-regulatory landscape. Here, we present *eQTLGen Phase 2*, a comprehensive expansion of our earlier work that increases the sample size to 43,301 individuals from 52 datasets, increasing statistical power, and extends *trans*-eQTL discovery to a genome-wide scale (**Figure 1a**). Leveraging an encoded meta-analysis approach that preserves privacy and maximizes computational efficiency^14^, we performed a genome-wide eQTL analysis, followed by fine-mapping of both *cis*– and *trans*-eQTLs, systematic *cis−trans* colocalization, and integration with 87 GWAS traits (**Figure S1)**. This large-scale design allowed us to identify regulatory variants affecting nearly all expressed genes, to reconstruct directed gene regulatory networks, and to distinguish regulatory variants with downstream functional impact from those that appear neutral.

**Figure 1.**
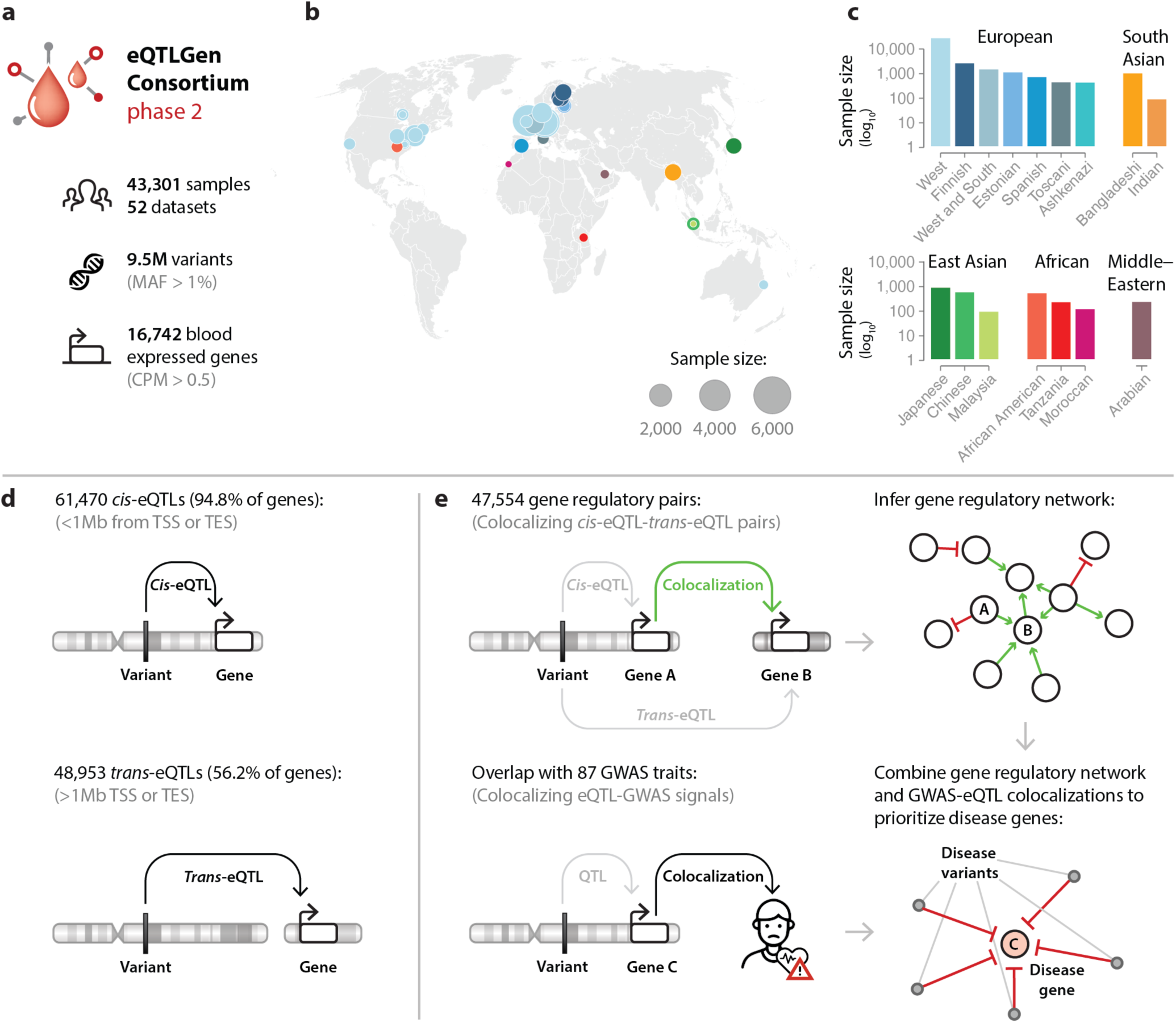
| Overview of the data and analyses. **a**, Overview of the data used for eQTL analyses. **b**, Overview of the cohorts contributing to the analyses. Dot size signifies sample size. Dot color denotes the datasets population label. **c**, Sample sizes of the cohorts from each population. Y-axis is shown in log_10_ scale. **d**, Overview of the different types of eQTLs, including the numbers of each kind found in this study. **e**, Different applications of eQTLGen data.

Our results reveal a pervasive landscape of gene regulation in which 56.2% of all genes are affected by *trans*-acting variants and a subset of *cis*-eQTLs serve as upstream drivers of widespread transcriptional and disease-related effects. We observe that variants with both *cis*-and *trans*-regulatory effects are markedly more likely to be associated with complex traits than variants with only *cis*-effects. This finding suggests that many *cis*-eQTLs do not propagate their effects through gene networks and may be less relevant for disease biology. Altogether, our findings refine the functional interpretation of eQTLs, provide a scalable approach to prioritize regulatory variants with downstream impact, and offer a more mechanistic understanding of how non-coding variation contributes to complex traits.

## Results

### Effect of common genetic variation on blood gene expression

We developed harmonized analysis pipelines to analyze 43,301 individuals from 52 cohorts from several populations (**Figure 1b**,**c**, **Figure S2** and **Table S1**). The genetic similarity between these cohorts recapitulates the clustering that is expected given the geographic origins of the datasets (**Figure S3**). Most participants (38,997, 90.1%) were of European annotated datasets (**Figure 1c**). After quality control (**Methods**), each cohort contributed between 8,635 and 28,896 profiled genes (median 18,346), of which 16,742 were retained for meta-analysis. Between 5.1 and 18.5 million variants per cohort (median 9.8 million) passed per-cohort filters (minor allele frequency (MAF) > 0.01, minor allele count (MAC) > 10), resulting in 9,514,548 variants that were included in the combined meta-analysis (**Figure 1a**, **Figure S4**).

eQTL analysis was conducted using a multivariate linear model for each variant–gene–cohort combination (**Methods**), followed by an inverse-variance weighted meta-analysis of all variant–gene combinations across cohorts. The median genomic inflation factor (median λ = 1.01, range 0.72−1.09, **Figure S5**) indicated well-calibrated results. We identified significant locus–gene combinations by merging the 0.5 Mb genomic regions surrounding each significant variant–gene association per gene and resulting loci varied from 0.5 Mb to 10.8 Mb in size (median = 1.1 Mb, mean = 1.4 Mb). At the genome-wide significance threshold of 5×10^−8^, we identified 57,682 primary locus–gene associations (excluding associations potentially caused by cross-mapping, cross-hybridization artifacts or for which the variants are in the HLA locus, **Methods**). Of these, 15,102 were *cis*-eQTLs (eQTLs for which the variants are located within ±1 Mb of the gene’s transcription start site (TSS) or transcription end site (TES)), and 42,580 were *trans*-eQTLs (eQTLs for which the variants are located >1 Mb from the TSS or TES). We estimate that the P-value threshold of 5×10^−8^ corresponds to an FDR < 1% for *cis*-eQTLs and ≈ 4.2% for *trans*-eQTLs (**Supplementary Note 1**).

To identify independent signals beyond primary eQTLs and prioritize variants most likely to be causal, we applied statistical fine-mapping (**Methods**). This approach detects multiple signals within the same locus and prioritizes variants by their probability of causality rather than by marginal association strength. Using the 57,682 significant locus–gene combinations, fine-mapping identified at least one credible set (CS) for 53,240 (92.4%) of them (log Bayes factor (LBF) > 2, eQTL P-value < 1×10⁻^5^, **Methods**). In total, fine-mapping detected 61,100 *cis*-eQTLs and 44,869 *trans*-eQTLs, encompassing 14,768 (92.4%) *cis*-eGenes and 8,867 (53.0%) *trans*-eGenes. These associations correspond to 42,166 independent *cis*-signals and 6,224 independent *trans*-signals (**Methods**). In the remainder of this work, we define eQTLs as significant or fine-mapped variant–gene associations.

We observed 12,750 *cis*-eQTL loci with multiple CSs, corresponding to 79.8% of genes and 84.2% of fine-mapped loci (mean CSs per locus = 4.03). For *trans*-eQTL loci, the average number of CSs per fine-mapped locus was 1.04, which is expected given the smaller effect sizes of *trans*-eQTLs compared to *cis*-eQTLs. The average CS size was 6.73 variants for *cis*-eQTL loci and 42.7 for *trans*-eQTL loci. Fine-mapping prioritized a single most likely causal variant (posterior inclusion probability (PIP) > 0.9) in 35,983 CSs (33.9%). Among all CSs, 58,181 (54.9%) contained at least one indel, with an indel being the most probable causal variant (PIP > 0.9) in 10,384 eQTLs (9.8%). Additionally, 1,578 CSs included at least one structural variant (SV), and an SV was identified as the most likely causal variant in 76 eQTLs (0.07%). Overall, 75.8% of the most likely causal eQTL variants (variants with the highest PIP or lead variants of a CS) were SNPs, followed by indels (24.0%) and SVs (0.2%).

For 4,371 loci (7.6%), the fine-mapping analysis did not result in a CS. For these loci, we included the eQTL variant with the highest absolute Z-score (eQTL P-value < 5×10⁻^8^). In total, we identified 61,470 *cis*-eQTLs and 48,953 *trans*-eQTLs, corresponding to *cis*-eQTLs for 15,146 genes (94.8% of all genes tested for *cis*-eQTLs) and *trans*-eQTLs for 9,406 genes (56.2% of all genes tested) (**Figure S5** and **Table S2**).

As expected, *cis*-effects are stronger than *trans*-effects (median |β| across the lead *cis*– and *trans*-eQTLs per gene = 0.171, 0.049; P-value < 2.2×10^−308^, Mann-Whitney U test, **Figure 2a**). For both *cis*– and *trans*-eQTLs the absolute betas of the most significant eQTL per gene were negatively correlated to loss-of-function (LoF) Z-scores^15,16^ (the standardized deviation between the observed and expected number of LoF variants). This indicates that genes with stronger eQTLs tend to be less evolutionary constrained (*cis*-eQTL P-value = 3.9×10^−96^, *trans*-eQTL P-value = 1.1×10^−37^; **Figure 2a**). Genes without detectable *cis*-eQTLs were typically lowly expressed (Mann-Whitney U test against expression deciles, P-value = 9.5×10^−16^) or more evolutionarily constrained (Mann-Whitney U test against predicted LoF intolerance (pLI), P-value = 7.5×10^−4^). For abundantly expressed genes (top 10^th^ expression decile), we observed a strong correlation between the number of CSs and the LOEUF score^15^ (a metric for a gene’s intolerance to LoF variation) of the corresponding genes (**Figure 2b**,**c****, Figure S6** and **Table S3**). This indicates that genes under stronger evolutionary constraint (lower LOEUF) tolerate fewer independent regulatory variants, in accordance with previous observations^6,17^. Thus, besides affecting coding variation, evolutionary constraint also appears to limit the accumulation of regulatory variation. While we observed a similar pattern for *trans*-eQTLs, the effect for those eQTLs is less pronounced (**Figure S7** and **Table S3**). We hypothesize that relatively more *trans*-eQTLs are allowed for constrained genes because *trans*-eQTLs are very context-dependent^18^ and generally have smaller effect sizes than *cis*-eQTLs. Consequently, *trans*-eQTLs are likely subject to lower evolutionary pressure than *cis*-eQTLs, similar to how GWAS signals tend to prioritize context– and trait-specific variants^19^.

**Figure 2.**
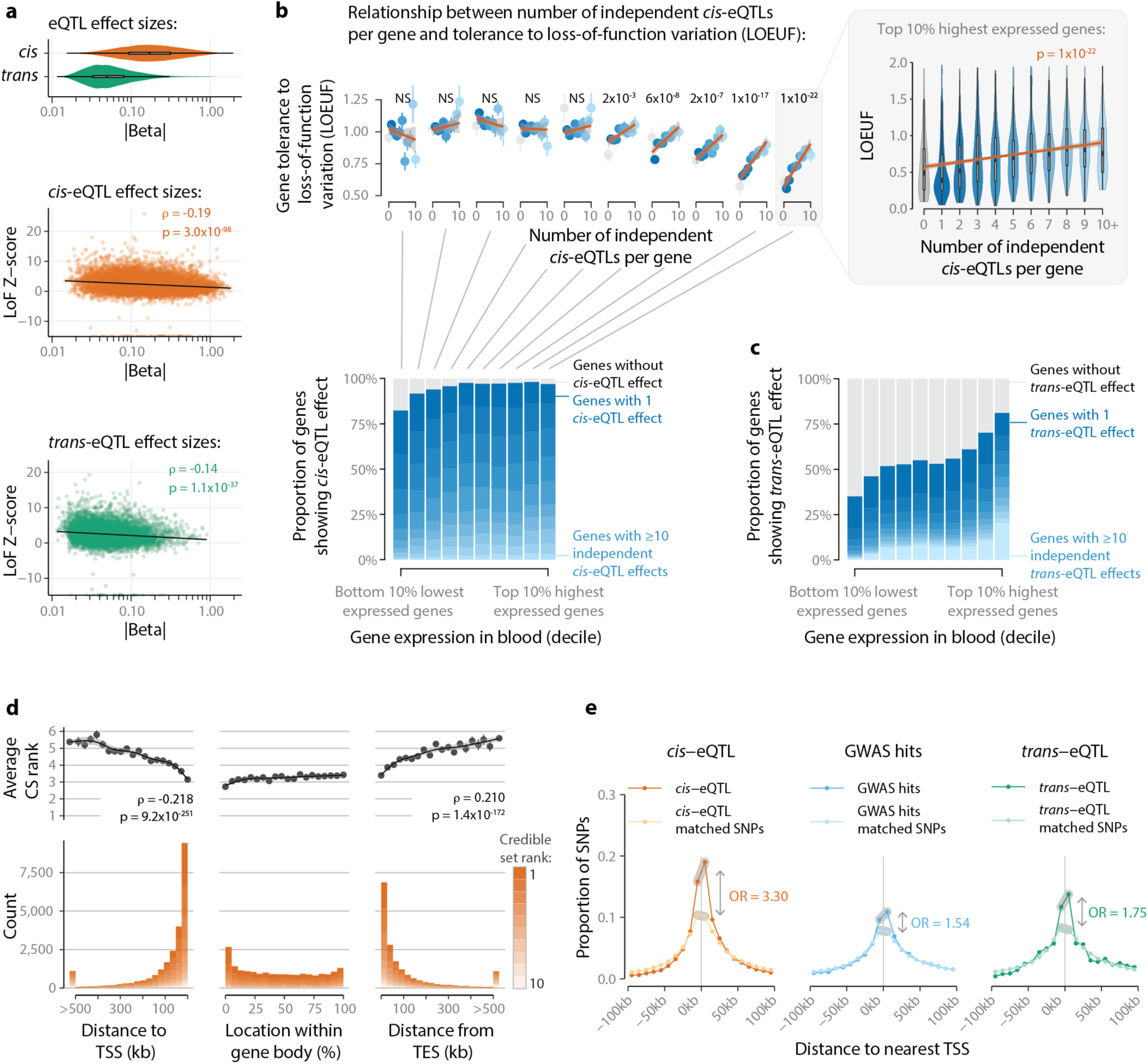
| Overview of *cis*– and *trans*-eQTLs. **a**, Top panel shows the difference in absolute beta between *cis*– and *trans*-eQTLs. Bottom panels show the relationship between the LoF Z-score and absolute beta of the lead eQTL per gene. **b**, Relationship between the number of *cis*-eQTL credible sets (CSs) per gene, evolutionary constraint, and gene expression levels categorized in the 10 gene expression deciles. Top panel shows the relationship between the number of independent *cis*-eQTL CSs and LOEUF scores, with a P-value indicating whether the visualized Spearman correlation is significant. Spearman correlations are calculated on all individual values per gene, while means (± standard errors) are presented across CS numbers. The right panel shows a violin plot of the same relationship for the top 10% highest expressed bins. The bottom panel shows the proportion of independent *cis*-eQTL CSs per gene expression decile. **c**, The proportion of independent *trans*-eQTL CSs per gene expression decile. **d**, The distribution of all *cis*-eQTL CSs compared to the gene body, normalized across all genes. CS rank represents the significance of each *cis*-eQTL with respect to all other *cis*-eQTLs per gene, with the most significant *cis*-eQTL per gene assigned rank 1 and less significant eQTLs assigned lower ranks. Top panel shows the average CS rank across the window, along with Spearman correlation estimates. Bottom panel shows the raw count of *cis*-eQTL CSs. **e**, The comparative distribution of *cis*-eQTLs, GWAS hits, and *trans*-eQTLs around the nearest TSS. Odds ratios (ORs) reflect the enrichment of these SNPs within 10kb either side of the nearest TSS compared to matched control SNPs.

We next investigated several other eQTL characteristics. To do so, we defined a set of randomly sampled background variants that match our eQTL variants based on MAF, linkage disequilibrium (LD), and TSS density. Indels were modestly enriched among eQTL variants compared to control variants (odds ratio (OR) = 1.04, P-value = 4.2×10^−5^, two-sided Fisher’s exact test), and SVs were substantially enriched (OR = 1.64, P-value = 1.6×10^−7^, two-sided Fisher’s exact test). As these SVs could reflect gene copy number alterations, this enrichment is expected based on the previously observed global dosage sensitivity of gene expression^20^.

Meanwhile, SNPs were depleted (OR = 0.96, P-value = 5.6×10^−6^, two-sided Fisher’s exact test; **Figure S8**). We applied the ENSEMBL Variant Effect Predictor^21^ to identify if specific variant annotations were enriched among the identified eQTLs. Among all *cis*-eQTL variants, we observed an enrichment of 19 variant effect predictions and a depletion of three variant effect predictions (Bonferroni-adjusted threshold = 8.7×10^−4^, two-sided Fisher’s exact tests, **Table S4**). Overall, *cis*-eQTL variants showed strong exclusive enrichment for several variant categories related to the length of the transcript, including frameshift, splice donor region, inframe insertion, and inframe deletion variants. These types of variants could result in differences in the length of the actionable transcript between genotype groups, which could manifest as a gene-level eQTL effect. Among all *trans*-eQTL variants, we observed an enrichment of seven consequences and a depletion of three (Bonferroni-adjusted α = 8.7×10^−4^, two-sided Fisher’s exact tests). Missense variants showed the strongest enrichment among *trans*-eQTL variants (OR = 3.8, P-value = 1.4×10^−56^), suggesting that some *trans*-eQTL variants may work on protein level by affecting protein folding or the DNA binding affinity of a putative upstream transcription factor (TF) or regulator.

We also observed that most *cis*-eQTL variants are positioned within the linked eGene (33.4%), while 21.4% are positioned 50kb upstream of the gene’s TSS and 15.4% are positioned 50kb upstream of the gene’s TES (**Figure 2d**). For each gene, we ranked the fine-mapped *cis*-eQTLs based on their significance, with the most significant assigned rank 1 and less significant ones receiving ascending (lower) ranks. In general, stronger *cis*-eQTL variants tended to be positioned closer to the gene body compared to weaker *cis*-eQTL variants (**Figure 2d**). Comparing the ranks across different locations relative to the gene body revealed that the most significant eQTLs per gene are for intragenic variants (mean CS rank 3.2), followed by those within 50 kb of a gene TSS (mean CS rank 3.3) and those within 50 kb of a gene TES (mean CS rank 3.5) (**Figure 2d**). Previous studies have shown that GWAS trait−associated variants are typically located further from the nearest TSS than *cis*-eQTL variants^22^. Consistent with this, we observed a similar pattern for the eQTLs we identified. Furthermore, compared to *cis*-eQTL variants, *trans*-eQTL variants are less enriched near the closest TSSs (*cis*-eQTL OR = 3.03, P-value < 2.2×10^−308^; *trans*-eQTL OR = 2.04, P-value < 2.2×10^−308^; P-value for difference in log odds = 1.4×10^−108^), suggesting that *trans*-eQTL variants are more similar to GWAS variants in this respect compared to *cis*-eQTL variants (**Figure 2e**).

### Replication of eQTLs across tissues and cell types

To evaluate the tissue-specificity and potential cellular drivers of the eQTLs identified, we undertook extensive replication analyses. We first quantified the overlap of *cis*– and *trans*-eQTLs with GWAS loci for blood cell−related and non-blood-cell traits to distinguish eQTLs driven by cell-type composition from those reflecting intracellular regulatory effects (**Figure 3a**). We then performed replication analysis across all the large-scale resources available to us, including single-cell eQTL meta-analyses in blood (sc-eQTLGen^23^, 1,323 donors) and brain^24^ (757 donors), bulk brain eQTLs from MetaBrain^13^ (2,759 unique cortex donors), bulk lymphoblastoid cell line (LCL) eQTL meta-analysis^25^ (3,734 samples), and plasma protein QTL (pQTL) data from 34,557 individuals from the Pharma Proteomics Project^26^ (**Figure 3b**, **Figure S9**, **Figure S10**, and **Table S5**).

**Figure 3.**
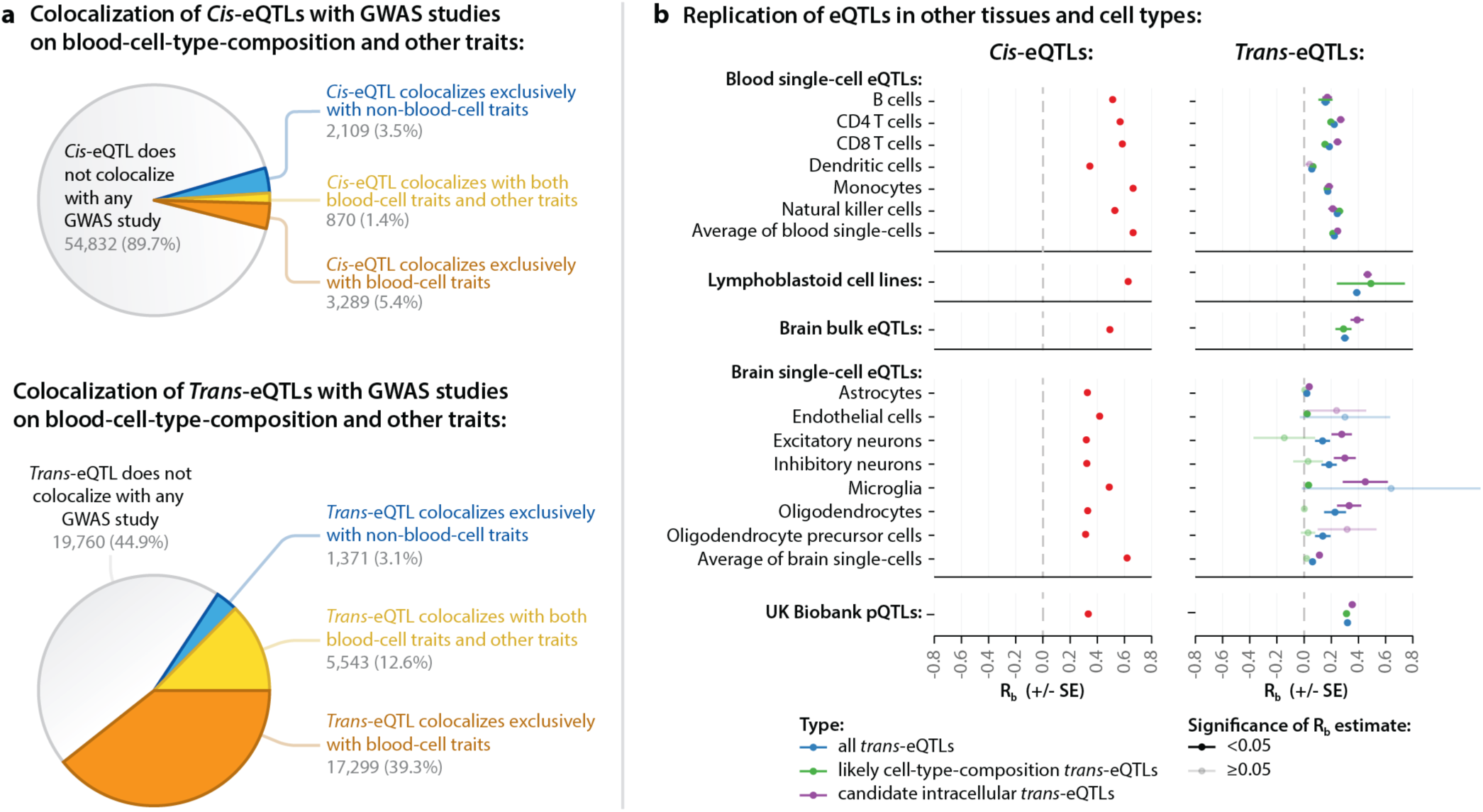
| Replication of *cis*– and *trans*-eQTLs and distinguishing between cell-type composition and intracellular effects. **a**, Fraction of *cis*-eQTLs (top) and *trans-*eQTLs (bottom) colocalizing with GWAS loci for blood-cell-related and non-blood-cell traits. *Cis*-eQTLs predominantly do not colocalize with any trait (89.7%), whereas *trans*-eQTLs show strong enrichment for blood-cell-related traits (39.3% exclusively blood cell, 12.6% both blood-cell and other traits). **b**, Replication of *cis*-eQTLs and *trans*-eQTLs in independent single-cell and bulk eQTL datasets from blood and brain and in the UK Biobank Plasma Proteomics Project pQTL data. *Trans*-eQTLs were further subdivided into likely cell-composition-driven and candidate intracellular subsets based on colocalization with any blood-cell-trait GWAS (colocalization PP4 > 0.8). Each point represents the correlation estimate of effects between the discovery and replication dataset (R_b_). Error bar depicts standard error of R_b_. Alpha level indicates significant replication signal (two-sided R_b_, P < 0.05). R_b_ estimates were consistently higher for *cis*-eQTLs across all cell types and tissues, while intracellular *trans*-eQTLs showed the highest R_b_ estimates in microglia in brain single-cell data. For the analyses in this figure, we excluded the 916 *trans*-eQTLs for which the gene-variant distance was < 5 Mb. Full results, including replication signals in GTEx, are shown in **Figure S9** and **Table S5**. **Figure S11** shows an alternate version of panel a wherein colocalizations are summarized by genetic signal instead of by eQTL.

Among all *cis*-eQTLs, 5.4% overlapped exclusively with blood-cell-related traits, 3.5% exclusively with non-blood-cell traits, and 1.4% with both (posterior probability for colocalization (PP4) > 0.8; **Figure 3a, Methods**). In contrast, *trans*-eQTLs were far more enriched for blood cell–related traits: 39.3% colocalized exclusively with blood-cell traits, 12.6% with both blood-cell and other complex traits, and only 3.1% exclusively with non-blood-cell traits (**Figure 3a**).

When aggregating these results over independent groups of genetic signals rather than variant–gene combinations, we observed that just 14.9% of independent *trans*-signals colocalized with exclusively blood-cell-related traits, 6.9% with both blood-cell and other complex traits, and 4.2 with exclusively non-blood-cell traits (**Figure S11**). These results indicate that a sizable proportion of *trans*-eQTLs likely reflect variation in blood-cell composition between individuals and thus do not represent true intracellular regulatory effects. On a variant level, this proportion was attenuated, reflecting that blood-cell composition loci often influence many genes simultaneously, which inflates their representation when counted at the eQTL level.

*Trans*-eQTL replication is complicated by two major factors: replication dataset sample size and the presence of cell-type-composition effects among the observed *trans*-eQTLs. While bulk eQTL datasets have larger sample sizes than single-cell datasets, bulk datasets cannot distinguish cell-type-composition effects from intracellular effects. We therefore performed replication analyses in both single-cell and bulk datasets from different tissues and cell types. We also stratified *trans*-eQTLs into two subsets using our colocalization results: one set likely to be driven by blood-cell-type composition, and another set more likely to represent intracellular *trans*-regulation (referred to as “candidate intracellular *trans*-eQTLs” below).

We first examined the replication of *cis*– and *trans*-eQTLs in the sc-eQTLGen consortium, an eQTL meta-analysis in single-cell RNA-seq data from six blood-cell types (B cells, CD4⁺ T cells, CD8⁺ T cells, dendritic cells, monocytes, and natural killer cells). Both *cis*– and *trans*-eQTLs showed significant replication signals (two-sided P-value < 0.05, significance test on R_b_ metric; **Figure 3b**, **Methods**) in all cell types, although replication was higher for *cis*-eQTLs (mean R_b_ = 0.54, range 0.34−0.67) and more modest for *trans*-eQTLs (mean R_b_ = 0.18, range 0.06−0.24). Similar to single-cell RNA-seq data from blood cell subtypes, LCLs are also a valuable data for prioritizing likely intracellular *trans*-eQTLs, and we observed relatively high replication in the largest available LCL resource (R_b_ of 0.63 and 0.39 for *cis*– and *trans*-eQTLs, respectively).

We also observed significant replication signal for *cis*– and *trans*-eQTLs derived from bulk brain tissue and from some of the single-cell brain datasets (**Figure 3b**). Among all brain cell types, microglia most closely resemble immune cells, functioning as macrophage-like cells in the brain^27^. Consistent with this role, microglia exhibited the strongest replication signal for “candidate intracellular” *trans*-eQTLs (R_b_ = 0.45, two-sided P = 6.6×10^−3^), despite representing only 4% of total included brain cells. Replication in other glial cell types (astrocytes and oligodendrocytes) and neurons was weaker, consistent with previous observations^13^. While replication power was limited by the available sample sizes, the concordant signals observed across multiple independent datasets suggest that a substantial subset of the *trans*-eQTLs we observe represent reproducible regulatory effects.

In GTEx post-mortem bulk tissues^5^, *cis*-eQTLs showed strong and widespread signal concordance, whereas the full set of *trans*-eQTLs were modestly concordant (**Figure S9, Figure S10**, and **Table S5**). We observed that the “candidate intracellular” subset of *trans*-eQTLs exhibited significantly higher R_b_ values than the full *trans*-eQTL set (two-sided P = 3×10^−8^, paired Wilcoxon test; **Figure S10**). The “candidate intracellular” subset of *trans*-eQTLs showed significant concordance in six tissues (two-sided P < 0.05, significance test on R_b_ metric, R_b_ > 0), while the full set of *trans*-eQTLs showed significant concordance in three tissues (including whole blood). These results suggest that *trans*-eQTLs that do not colocalize with blood-cell-trait GWAS are depleted of cell-type-composition effects and are thus more likely to reflect genuine intracellular regulation.

Finally, both *cis*– and *trans*-eQTLs replicated significantly in plasma pQTL data from the UK Biobank^26^ (R_b_ of 0.33 and 0.32, respectively; **Figure 3b**). Here, the direction of allelic effects was concordant for 70% of replicated *cis*-eQTL effects and 81% of replicated *trans*-eQTL effects (Benjamini-Hochberg false discovery rate (FDR) < 0.05; **Figure S12**), underscoring the biological relevance of the identified *cis* and *trans* links across molecular layers. Together, these analyses demonstrate that *cis*-eQTLs are broadly shared across tissues and cell types, whereas *trans*-eQTLs are more context-specific, with a considerable subset driven by differences in cellular composition.

### Inferring directed gene regulatory networks using *cis*– and *trans*-eQTLs

Having genome-wide summary statistics for all assessed genes allowed us to identify the overlap between *cis*– and *trans*-eQTLs. Identifying this overlap might suggest that the identified eQTL variant impacts the *trans*-eGene through the *cis*-eGene (vertical pleiotropy), allowing us to infer a putative directional relationship between the *cis*-eGene and the *trans*-eGene. To test whether this overlap exists, we performed colocalization analyses between all pairs of *cis*– and *trans*-eQTLs in overlapping genomic regions.

We identified 49,715 colocalization events between *cis*-eQTL and *trans*-eQTL signals (colocalization PP4 ≥ 0.8 and LD R^2^ ≥ 0.8 between lead variants of colocalizing signals; **Methods**, **Table S6**), constituting 3,558 out of 61,100 (6%) fine-mapped *cis*-eQTL CSs and 8,351 out of 43,973 (18%) fine-mapped *trans*-eQTL CSs. By comparing the effect directions of both the *cis*– and *trans*-eQTL CSs we can infer for every colocalizing pair of eQTLs if the *cis*-eGene is likely activating or repressing the downstream *trans*-eGene. For 1,083 gene pairs, multiple eQTL CS pairs colocalized. For 105 (9.7%) of these, the effect directions inferred from the *cis*– and *trans*-eQTL signals were inconsistent between eQTL CS pairs. After removing these *cis*-eGene−*trans*-eGene pairs, we identified 47,554 linked *cis*-eGene−*trans*-eGene pairs (reflecting 2,612 unique *cis*-eGenes and 6,533 *trans*-eGenes; **Figure 4a**). Taking all these links together allows us to infer an interconnected network. We observe that a few highly connected *cis*-eGenes have many downstream effects, while many *cis*-eGenes have a limited number of connections. Meanwhile, there are comparatively few *trans-*eGenes for which we detected many upstream regulators (**Figure 4b**). We observed that the 2,612 unique *cis*-eGenes are strongly enriched for encoding TFs (Gene Ontology Term GO:0043565, P-value = 2×10^−82^, ToppGene hypergeometric test^28^, **Methods**) or transcriptional coregulators (GO:0003712, P-value = 5×10^−7^) or show colocalization with blood-cell traits (P-value = 1×10^−110^; **Figure 4c**). Notably, we observe an increasingly enriched fraction of transcription factors (GO:0043565) among *cis*-eGenes that exhibit multiple colocalizing signals with their corresponding *trans*-eGenes (**Figure 4d**). For 24 *cis*-*trans* gene pairs there are three independent credible sets (LD R^2^<0.5 between lead variants) at the *cis*-eQTL locus that colocalize with three independent credible sets at the corresponding *trans*-eQTL locus (**Table S7**). In all cases, the allele increasing expression of the *cis*-eGene exerted a consistent directional effect on the *trans*-eGene across all three independent signals, either increasing or decreasing the expression in a concordant manner, and 17 out of 19 unique *cis*-eGenes (89%) have known transcription regulatory activity. For example, three independent *cis*-eQTL signals for the *NPAS2* gene colocalize with three independent *trans*-eQTL effects of *NR1D1* (**Figure S13**) with proportional *cis*-eQTL effects observed for both genes (**Figure S13c**). Both genes encode key components of the circadian clock and are known to regulate each other by a regulatory feedback loop in the brain^29^. Together, these observations support a causal, dose-dependent regulatory relationship between such *cis*– and *trans*-eGenes, rather than coincidental linkage or pleiotropic effects.

**Figure 4.**
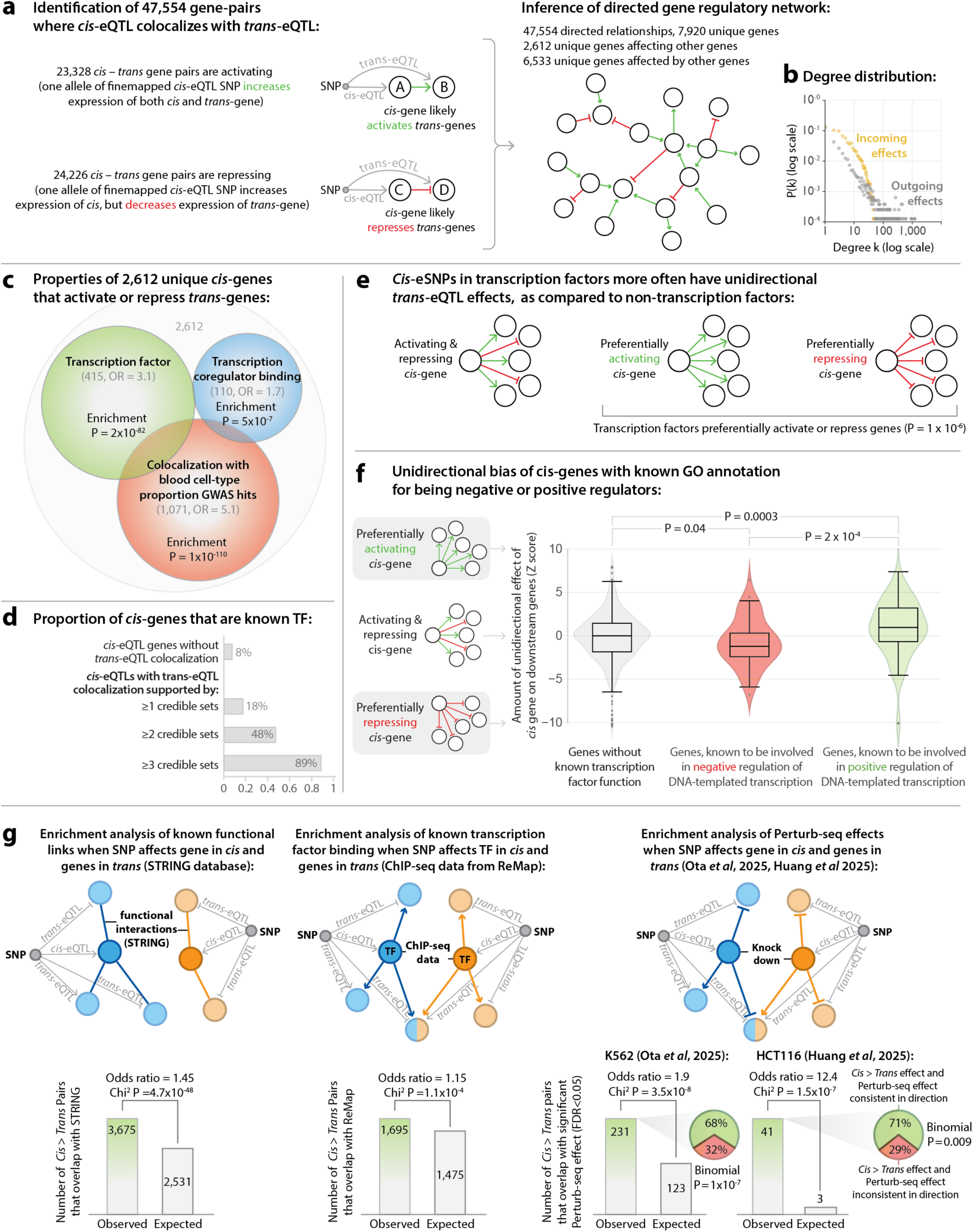
| Inferring directed gene regulatory networks using *cis*– and *trans*-eQTLs. **a**, Overlapping fine-mapped *cis*– and *trans*-eQTLs reveals 47,554 *cis−trans* gene pairs (2,612 unique *cis*-genes and 6,533 *trans*-genes), allowing inference of activating (23,328) and repressing (24,226) relationships. Activating and repressing relationships are indicated by green and red arrows, respectively. **b**, Degree distribution of the inferred gene-regulatory network. Purple dots represent the out-degree distribution, indicating that only a few *cis*-genes regulate many downstream *trans*-genes. Yellow dots represent the in-degree distribution, showing that comparatively few genes have many upstream regulators. The x-axis shows the number of connections for each gene in the network (k), and the y-axis shows the proportion of genes with k connections (P(k)). **c**, *Cis*-genes that regulate *trans*-genes are enriched for transcription factors, transcriptional coregulators, and loci colocalizing with blood-cell-trait GWAS hits. **d**, Fraction of *cis*-eGenes annotated as transcription factors (GO:0043565) as a function of the number of colocalizing eQTL signals with their corresponding *trans*-eGenes. **e**, Transcription factor *cis*-genes more often show unidirectional effects, preferentially activating or repressing *trans*-genes, compared to non-transcription factor *cis*-genes. **f**, Transcription factors that preferentially activate *trans*-genes are enriched for positive regulatory processes, while those that preferentially repress *trans*-genes are enriched for negative regulatory functions. **g**, Inferred regulatory links are supported by known functional interactions from STRING, transcription factor binding from ReMap ChIP-seq data^31^, and gene perturbation effects from Perturb-seq^32,33^, with consistent direction of regulation for most overlapping pairs. Cartoons above each bar chart illustrate what each dataset type tests (left-to-right: undirected links, directed links, and directed links with activating/repressing properties). Blue and orange arrows depict gene links in the two dummy networks.

For 23,328 of the gene pairs (49%), the same allele increased the expression of both the *cis-*eGene and *trans*-eGene, suggesting that the *cis*-gene activates the *trans*-gene (**Figure 4a**). For the other 24,226 gene pairs (51%), the variant’s alleles regulate the *cis*-eGene and *trans*-eGene in opposite directions, suggesting that the *cis*-eGene represses the *trans*-eGene. In addition, *cis*-eGenes that are TFs preferentially show unidirectional activating or repressing effects on colocalizing *trans*-eGenes (**Methods**) when compared to non-TF *cis*-eGenes (P-value = 1×10^−6^, Wilcoxon signed-rank test; **Figure 4e** and **Methods**). We also observed that the directional bias of *cis*-eGenes (quantified by the Z-score from a binomial test, positive values indicating preferential activation and negative values indicating preferential repression), aligns with established gene regulatory roles. Specifically, genes annotated for negative regulation (GO:0045892) more frequently exhibit repressing effects, whereas those associated with positive regulation (GO:0045893) tend to display activating effects. (**Figure 4f** and **Methods**).

We then studied to what extent the putative directional relationships are supported by orthogonal data. For this, we used known functional interactions between gene pairs from STRING^30^ (an aggregated database of protein–protein interactions derived from computational predictions, curated sources, and literature mining), ChIP-seq data from ReMap^31^ (which allowed us to test whether upstream *cis*-eGenes bind near their downstream *trans*-targets), and two recent genome-wide Perturb-seq datasets (experimental mapping of transcriptional changes following perturbation of individual genes) in K562^32^ and HCT116^33^ cells (**Figure 4g**). Comparing the set of inferred putative directional relationships with a matched set of control pairs revealed greater-than-expected overlap across all resources: STRING (P-value = 4.7×10^−48^, two-sided Chi^2^ test), ReMap (P-value = 1.1×10^−4^), the K562 Perturb-seq dataset (P-value = 4×10^−8^) and the HCT116 Perturb-seq dataset (P-value = 2×10^−7^). The relative enrichment was highest for the Perturb-seq datasets (OR = 1.9 and 12.4, respectively). Unlike STRING and ReMap, the Perturb-seq datasets also enabled us to determine to what extent the observed direction of effect (activation or repression) was consistent with experimental perturbation outcomes. We found that this was the case for 68% of the 231 overlapping relationships in the K562 dataset and 71% of 41 overlapping relationships for the HCT116 dataset, considerably more than what would be expected by chance (P-values = 1×10^−7^ and 9×10^−3^ for K562 and HCT116 respectively, two-sided binomial tests). While the *trans*-eQTLs that we link to upstream regulators could, in principle, be mediated by *cis*-eQTLs acting through an intermediate trait^34^ rather than reflecting a direct effect of *cis*-eQTLs, those results suggest that many of the identified gene–gene pairs likely represent biologically interpretable intracellular gene regulation that can be corroborated using different lines of evidence.

### eQTL-informed regulatory networks converge on trait-relevant genes and pathways

To illustrate how directed gene regulatory links inform relevant pathways, we examined which *cis*-eQTLs and *trans*-eQTLs colocalize with complex trait associations. To do so, we conducted comprehensive colocalization analyses with 87 GWAS traits covering immune, metabolic, respiratory, and neuropsychiatric domains (**Table S8**). This analysis identified colocalization for 30,029 (28.3%) eQTL signals, comprising 10.1% of *cis*-eQTLs, 9.6% of the distinct genetic signals for which we detect *cis*-effects, 53.2% of *trans*-eQTLs, and 24.5% of the distinct genetic signals for which we detect *trans*-effects (**Methods**).

Across all loci, the proportion of GWAS signals that colocalized with eQTLs was highly trait-dependent (**Figure 5a, b**). Cell-type-composition and immune-mediated traits showed the strongest overlap, followed by metabolic and neuropsychiatric traits. This is expected given the tissue-specific relevance of blood for blood-cell-type-composition and immune traits and the tissue specificity of *trans*-eQTLs. For individual phenotypes, we observed the largest fractions of GWAS loci colocalizing with at least one eQTL for blood-cell traits, autoimmune diseases (Type I diabetes 47%, Crohn’s disease 47% and eczema 45%), respiratory diseases (45%), and lipid traits (total cholesterol 45% and LDL cholesterol 42%) (**Figure S14, Table S9**). Among the *cis*-eQTLs that colocalized with *trans*-eQTL effects, a substantial proportion (71.9%) also colocalized with at least one GWAS phenotype, suggesting that genetic variants with both *cis-*and *trans-* effects are particularly relevant for complex trait variation.

**Figure 5.**
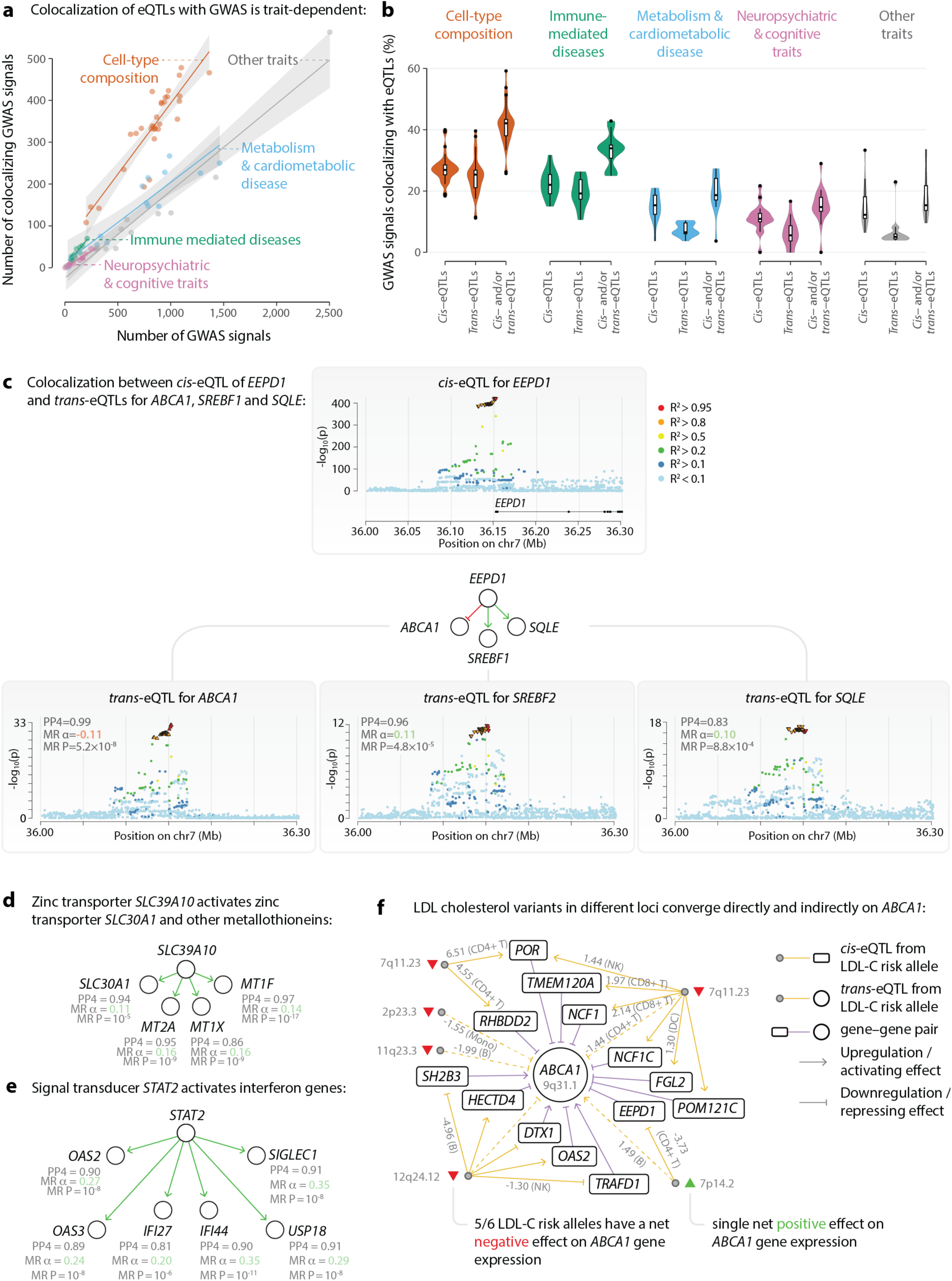
| Colocalized eQTL−GWAS networks reveal directed regulatory cascades and convergence on disease genes. **a**, Trait-specific dependence of eQTL−GWAS colocalization counts, highlighting the strongest overlap for immune and cell-type-composition traits. **b**, Distribution of GWAS signal colocalizations across major trait categories for *cis*– and *trans*-eQTLs. **c**, A *cis*-eQTL for *EEPD1* colocalizes with *trans*-eQTLs for *ABCA1, SREBF1,* and *SQLE*, which are key regulators of lipid metabolism. Colocalization posterior probabilities (PP4) and Mendelian randomization analyses (MR α) support a directional effect from *EEPD1* to its targets. **d**, The product of zinc transporter encoding gene *SLC39A10* regulates *SLC30A1* and the metallothionein-encoding genes *MT1X* and *MT2A* through *trans*-eQTLs, forming a zinc homeostasis network. **e**, The signal transducer encoding gene *STAT2* exerts *trans*-effects on multiple interferon-stimulated genes (*OAS2, OAS3, IFI6*, and *IFI44*), reconstructing the canonical interferon pathway. **f**, Example of LDL cholesterol (LDL-C)−associated variants converging on *ABCA1* through direct *cis*-eQTL-mediated and indirect *trans*-eQTL-mediated regulatory paths. Where available, edges are annotated with the most significant concordant single-cell eQTL Z-score and cell type from sc-eQTLGen^23^.

As an example, a *cis*-eQTL for *EEPD1*, which encodes a protein involved in DNA repair and lipid metabolism, colocalizes with LDL, non-HDL, and total cholesterol GWAS association signals^35^ in this locus (PP4 > 0.8; **Figure S15**). This *cis*-eQTL also colocalizes with *trans*-eQTLs for the genes *ABCA1*, *SREBF1,* and *SQLE* (colocalization PP4 = 0.99, 0.96, and 0.83, respectively; **Figure 5c, Figure S16**), which encode proteins that are key regulators of cholesterol biosynthesis (**Figure 5c**). Importantly, the regulatory function of the protein product of *EEPD1* on the ABCA1 protein has been supported by a functional study^36^. The shared genetic signal suggests that the protein encoded by *EEPD1* modulates lipid metabolism through a molecular cascade that converges on these downstream targets. *Cis*-Mendelian Randomization (MR) analysis shows a downregulating effect of *EEPD1* on *ABCA1* (MR α = – 0.11, two-sided P = 5.2×10^−8^; **Methods** and **Figure 5c**) and up-regulating effects of *EEPD1* on *SREBF2* (MR α = 0.11, two-sided P = 4.8×10^−5^) and *SQLE* (MR α = 0.10, two-sided P = 8.8×10^−4^). Similarly, we observed significant downregulating causal effect estimates from *EEPD1* to LDL (MR α = –0.02, two-sided P = 1.3×10^−97^; **Figure S13**), non-HDL (MR α = –0.02, two-sided P = 2.4×10^−100^), and total cholesterol (MR α = –0.02, two-sided P = 1.8×10^−96^). Together, these findings provide supporting evidence for a directional effect from *EEPD1* to its downstream *trans*-eQTL genes, as well as to lipid levels. This finding is consistent with the known interactions of *EEPD1* acting upstream of lipid homeostasis genes and thereby influencing plasma lipid levels^36,37^.

A second example involves the *SLC39A10* gene, whose protein product, ZIP10, mediates the influx of zinc ions into the cytoplasm^38^. The primary *cis*-eQTL for *SLC39A10* colocalizes with a *trans*-eQTL for *SLC30A1*, a gene encoding for the zinc exporter ZNT1 that reduces cytoplasmic zinc levels^39^. This relationship mirrors the established opposing roles of ZIP10 and ZNT1 in maintaining zinc balance. Furthermore, a quaternary *cis*-signal colocalizes with secondary *trans*-signals for the metallothionein-encoding genes *MT1X, MT2A,* and *MT1F* whose expression patterns are known to respond to cytoplasmic zinc levels^40^. MR analysis linking the *cis*– and *trans*-eQTLs supports a directional effect from *SLC39A10* to *MT1X* (MR α = 0.16, two-sided P-value = 5.4×10^−9^)*, MT2A* (MR α = 0.16, two-sided P-value = 2.9×10^−9^), and *MT1F* (MR α = 0.14, two-sided P-value = 2.4×10^−17^) (**Figure 5d**, **Figure S17**, and **Figure S16**). This is consistent with prior evidence that ZIP10 activity influences metallothionein expression^41^. Moreover, the activating relationship between *SLC39A10*, *MT2A* and *MT1X* is also supported by perturbation experiments in HCT116 cells^33^. Together, these results recapitulate the established biological relationships in cellular zinc homeostasis and provide a candidate molecular framework for interpreting nutrient and metal ion homeostasis in blood.

Finally, the signal transducer protein encoded by the gene *STAT2* emerged as a central upstream regulator with *trans*-effects on six interferon-stimulated genes (*OAS2*, *OAS3*, *IFI27*, *IFI44*, *SIGLEC1* and *USP18*; **Figure 5e**, **Figure S18**, and **Figure S16**). This network recapitulates the canonical interferon signaling pathway, underscoring the biological plausibility of our inferred regulatory links.

Integrating *cis−trans* networks with GWAS data revealed cases where distinct trait-associated variants converge on the same downstream gene with consistent regulatory effects. Specifically, we traced all the paths from GWAS variants to genes with a maximum length of two edges and determined whether the trait-increasing allele upregulated or downregulated each downstream gene. For each gene, we then aggregated the directional effects across all paths to compute a gene prioritization score, defined as the sum of signed effects from individual paths (**Methods**). Across 70 non-blood cell-type-composition traits, we identified 782 genes linked to 37 traits with an absolute prioritization score of at least 2 (Benjamini-Hochberg-corrected P-value < 0.05, two-sided Z-tests; **Table S10**). Of these 782 genes, 382 (48.8%) were consistently upregulated and 400 (51.2%) consistently downregulated by trait-increasing GWAS variants. For example, the expression of *ABCA1*, whose product is the cholesterol efflux pump^42^, is directly and indirectly regulated by six variants associated to LDL cholesterol through both *cis*– and *trans*-mediated paths, and five show a consistently downregulating effect (**Figure 5f** and **Figure S19**). Five of the six paths are further supported by concordant cis-eQTLs with nominal significance in sc-eQTLGen^23^. (**Figure 5f**). The importance of ABCA1 in cholesterol transport is well established, as loss-of-function variants in *ABCA1* can cause tangier disease, which is a rare disorder that is characterized by markedly reduced HDL cholesterol and excessive cellular cholesterol accumulation^43^. Together, these examples demonstrate that large-scale *cis−trans* integration enables reconstruction of disease-relevant regulatory pathways and provides mechanistic insights into how non-coding variants converge on core genes underlying complex traits.

### Distinguishing functionally active *cis*-eQTLs from putative neutral *cis*-eQTLs

To better understand the regulatory architecture of eQTLs, we compared *cis*-eQTLs based on whether they were accompanied by downstream *trans*-effects. We defined *functionally active cis*-eQTLs as variants that both modulate the expression of a nearby gene (*cis*-effect) and exert a *trans*-eQTL effect on a distal gene. In contrast, we defined *putatively neutral cis*-eQTLs as variants that influence a gene in *cis* but are not associated with any detectable downstream *trans*-eQTL effects (**Figure 6a**). Among the 61,470 *cis*-eQTLs, 3,558 (5.8%) are defined as functionally active and 57,912 (94.2%) are considered putatively neutral. Given that *trans*-eQTLs were tested only in blood, *cis*-eQTLs without detectable downstream effects in our study could still exert *trans*-effects in other tissues. Nevertheless, the presence of *cis*-eQTLs in blood without downstream consequences raises questions about their functional relevance in this tissue.

**Figure 6.**
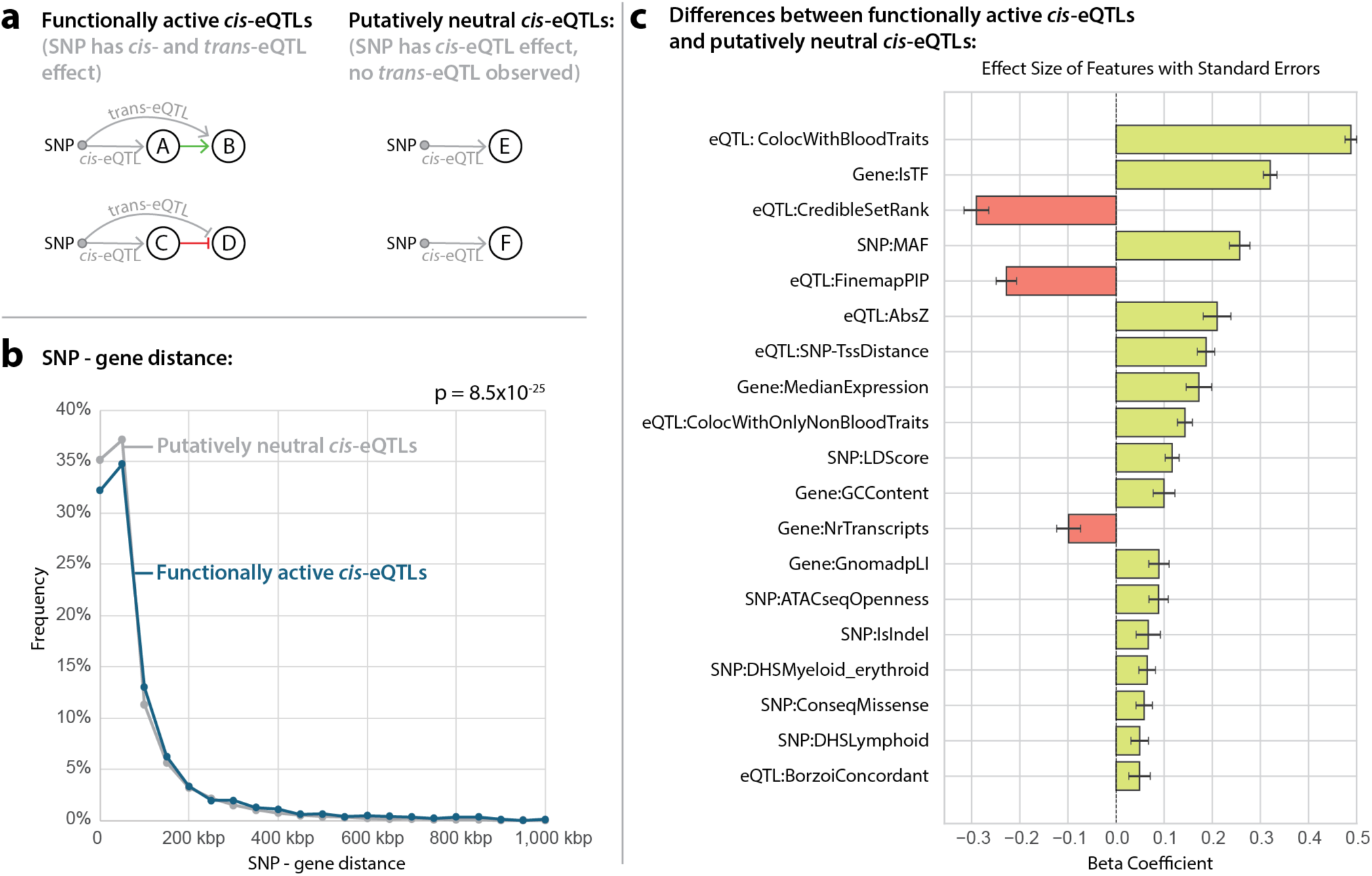
| Distinguishing functionally active from putatively neutral *cis*-eQTLs. **a**, Schematic overview defining “functionally active” *cis*-eQTLs as variants exhibiting both *cis*– and *trans*-eQTL effects and “putatively neutral” *cis*-eQTLs as variants showing only *cis*-effects without detectable downstream *trans* associations. **b**, Functionally active *cis*-eQTLs are typically located further from their target genes compared to neutral *cis*-eQTLs. **c**, Comparison of genomic and regulatory features between active and neutral *cis*-eQTLs. Significant enrichments (logistic regression model) are shown with red and green colors. Red indicates a depletion or a negative correlation for the functionally active *cis*-eQTLs, green colors indicate an enrichment or a positive correlation for the functionally active *cis*-eQTLs. Functionally active *cis*-eQTLs are less often fine-mapped to single variants, have higher linkage disequilibrium scores, and more frequently map to regions of open chromatin. They are also more likely to be missense variants, to affect constrained genes, and to show concordant regulatory direction with sequence-based models. In contrast, neutral *cis*-eQTLs more often occur near the transcription start site, affect genes with many transcript isoforms, and show discordant effects with sequence-based predictions. Together, these results distinguish a subset of *cis*-eQTLs with broader regulatory impact from those with limited or no downstream consequences. All features were standardized to unit variance before estimating beta coefficients. Error bars reflect standard errors of the beta coefficients.

We subsequently collected 46 eQTL features (**Table S11**)^5,15,21,22,25,44–47^ that were informative about either the *cis*-eQTL (e.g., SNP−gene distance), the *cis*-eQTL SNP (e.g., extent of LD), or the *cis*-eQTL gene (e.g., pLI score) and used these as explanatory variables to predict whether a *cis*-eQTL is functionally active or not by using logistic regression. Because colocalization depends on signal strength, we included allele frequency, *cis*-eQTL significance, and median expression of the *cis*-eGene to account for this dependency.

Functionally active *cis*-eQTLs exhibit markedly different genomic and regulatory characteristics compared to putatively neutral ones (**Table S11**). Functionally active *cis*-eQTLs typically map further away from the genes (β = 0.19, two-sided P-value = 8.5×10^−25^; **Figure 6b**) and are significantly more likely to reside in regions of open chromatin, determined by single-cell ATAC-seq data (β = 0.089, P-value = 9.4×10^−6^) and DNA hypersensitivity sites (β = 0.065, P-value = 1.9×10^−4^; **Figure 6c**). However, when functionally active variants map within the *cis*-gene, they are more often missense variants (β = 0.059, P-value = 6.1×10^−4^).

Functionally active *cis*-eQTLs are less often fine-mapped to the resolution of a single variant (β = –0.23, P-value = 1.1×10^−26^) and have higher LD scores (a quantity for the amount of LD a variant has with neighboring variants^48^; β = 0.12, P-value = 5.8×10^−16^). Functionally active *cis*-eQTL genes are also more constrained (as reflected by pLI; β = 0.089, P-value = 2.2×10^−5^) and thus more often exhibit essential fundamental functions in the organism. Notably, they more often encode TFs (β = 0,32, P-value = 9.1×10^−115^), indicating that they more often have a central position in regulatory cascades.

Functionally active *cis*-eQTLs are also significantly more likely to colocalize with GWAS signals, reinforcing their potential regulatory relevance. However, we do note that the colocalizing traits are often blood-cell-type composition traits (β = 0.49, P-value < 2.2×10^−308^), which suggests some of these functionally active *cis*-eQTLs are not resulting in intracellular *trans*-eQTLs but rather show *trans*-eQTLs on genes that are markers for the corresponding cell type. When excluding blood-cell-type-composition GWAS colocalizations, the enrichment of colocalization of functionally active *cis*-eQTLs with GWAS traits is somewhat attenuated but still highly significant (β = 0.14, P-value = 4.7×10^−20^). Of note, 2,085 functionally active *cis*-eQTLs do not show any colocalization with disease-associated GWAS signals but are still strongly enriched for encoding TFs (P-value = 4×10^−63^, two-sided Chi^2^ test).

In contrast, putatively neutral *cis*-eQTLs more often affect genes with a large number of transcript isoforms (β = –0.10, P-value = 7.5×10^−5^), suggesting that some of these neutral *cis*-eQTLs may primarily influence minor isoforms rather than the dominant transcript, resulting in reduced functional impact. Putatively neutral *cis*-eQTLs also more often show opposite allelic effects to predicted differential expression by Borzoi (β = 0.05, P-value = 0.02), suggesting that these variants function less often in the way that is predicted based on the context of the genomic sequence.

To test the robustness of these findings, we performed these analyses again while confining ourselves to a subset of genes on which multiple variants were having an independent *cis*-eQTL effect but with discrepant *trans*-eQTL effects, e.g., a *cis*-eQTL gene with two CSs where one variant is associated with a *trans*-eQTL but the other is not. Similar to the analysis above, we considered those variants with *trans*-eQTLs as functionally active and those without as putatively neutral. We then compared 2,877 functionally active with 3,715 putatively ‘non-functional’ *cis*-eQTLs and redid the logistic regression analysis. Except for the concordance with Borzoi, all previously mentioned enrichments remained significant, with the same direction of effect (**Table S11**), suggesting the robustness of these results.

While 23,316 (52.0%) of the 44,869 fine-mapped *trans*-eQTLs originate from variants that also act in *cis*, roughly 4,720 (75.8%) out of the 6,224 independent *trans*-eQTL variants did not show significant *cis*-effects on nearby genes. These “*trans*-only” variants were strongly depleted for colocalization with complex trait loci compared to variants with both *cis*– and *trans*-effects (OR ≈ 5.0, P-value < 6.0×10^−137^, two-sided Fisher’s exact test). Excluding *trans*-variants associated to blood-cell-composition, we again observed a significant, but somewhat attenuated depletion (OR ≈ 2.9, P-value < 2.9×10^−13^, two-sided Fisher’s exact test). This depletion suggests that many *trans* associations lacking a *cis* mediator might not represent stable intracellular regulatory effects but rather cell (sub-type) composition or other context-dependent effects. Our results thus reinforce that disease-relevant regulatory variants predominantly act through local (*cis*) perturbations that propagate downstream (*trans*), while *trans*-only associations are more difficult to interpret.

Taken together, these findings support a conceptual distinction between regulatory variants that have broader functional consequences and those with limited downstream effects. The enrichment of functionally active *cis*-eQTLs in TFs, enhancers, and GWAS loci suggests that they often serve as upstream regulators in gene networks. Conversely, the origins of putatively neutral *cis*-eQTLs remain more elusive, making their functional impact unclear. These insights highlight the need to reconsider how *cis*-eQTLs are interpreted in genomic studies and suggest that a subset of them may not be biologically meaningful in the context of regulatory networks.

## Discussion

In this study, we conducted the most highly powered and comprehensive eQTL meta-analysis to date and mapped how genetic variation influencing gene expression in human blood across 43,301 individuals from eQTLGen Phase 2. The study is a substantial advance over our earlier release^1^, with a 37% increase in sample size and having genome-wide *trans*-eQTL coverage.

In accordance with recent large-scale studies^6,12,49,50^, we demonstrate that nearly all expressed genes are under genetic control, with 15,146 (94.8% of tested genes) of them showing *cis*-eQTL effects. Similarly, our sample size allowed us to detect non-primary *cis*-eQTL signals for 79.1% of eGenes. For *trans*-eQTLs, we report 42,580 primary associations affecting 9,406 genes (56.2% of those tested) and 48,953 when including non-primary signals, expectedly exceeding previous smaller-scale studies^6,12,49^. The high power and genome-wide scope of our analyses have enabled several key insights into the architecture of human gene regulatory networks that have not been possible to evaluate before.

First, our results show that *trans*-regulation is pervasive and largely mediated by upstream *cis*-regulated genes, which are often TFs. The resulting 47,554 directed regulatory links reconstruct known biological hierarchies and are corroborated by Perturb-seq data, supporting their biological relevance. Given that these findings reflect in vivo causal inferences, they also provide a complementary approach for identifying regulatory relationships in contexts that are challenging to capture with in vitro models. Moreover, the inferred network offers a prioritized set of candidate regulatory interactions that can guide and accelerate targeted in vitro experiments, reducing the need for exhaustive screening.

Second, integration with GWAS data across 87 complex traits reveals that adding genome-wide *trans*-effects to the analyses nearly doubles the proportion of GWAS loci with a potential molecular explanation, underscoring the importance of distal regulation in complex trait etiology. We demonstrate that genome-wide *cis*– and *trans*-eQTL data facilitates the shift from association– to causal-level inference, enables systematic prioritization of trait-related regulatory pathways, and pinpoints biologically interpretable trait-related candidate genes. For example, the outlined cases of *EEPD1*, *SLC39A1*, and *STAT2* illustrate putatively causal links between *cis*-regulators and downstream targets, whereas the regulatory network of variants associated with LDL cholesterol converges on *ABCA1*, *a* well-known gene encoding a cholesterol transporter^42^. Importantly, our approach provides a framework that can be readily expanded to any complex trait.

Third, variants with both *cis*– and *trans*-effects are more likely to colocalize with trait-associated loci than *cis*-only variants. This observation supports a conceptual distinction between functionally active *cis*-eQTLs with regulatory effects on downstream genes and putatively neutral *cis*-eQTLs with limited biological consequence. Functionally active *cis*-eQTLs are enriched for TFs, distal enhancers, and functionally essential genes, whereas putatively neutral *cis*-eQTLs often affect highly spliced or lowly expressed genes. Such distinction between functional and neutral molecular variation is in line with a recent observation, where removing the genetically driven component of protein abundance markedly improved the protein-disease associations^51^, suggesting that a large fraction of genetic regulation of proteins is non-disease-relevant. We speculate that this may also hold for gene expression levels, where we detected propagating systems-level effects for only a subset of *cis*-eQTLs. As such, our results refine our understanding of which eQTLs are more likely to mediate disease mechanisms and have a potential to improve SNP-to-gene linking strategies in the future.

Several hypotheses may explain why some *cis*-eQTLs do not exhibit *trans*-effects. First, post-transcriptional buffering may limit the noise in mRNA levels of upstream *cis*-eGenes^52–54^. Second, eQTLs could exhibit buffering, as tightly linked variants may have opposing effects at different stages of the central dogma^55^. Third, cell-type-specificity is likely to play a role: functionally active and neutral *cis*-eQTLs for the same gene might act in distinct cellular contexts, explaining differences in downstream consequences^18^. Fourth, technical artifacts, such as primer polymorphisms or alignment bias, could also generate spurious *cis*-eQTLs^56^. Finally, multiprotein or multienzyme complexes can buffer the downstream effects of *cis*-eQTLs on individual subunits. Variants that increase the expression of a rate-limiting subunit of a multiprotein complex can relieve a bottleneck, increasing complex activity, and thereby propagate *trans*-effects. In contrast, variants that affect non-limiting subunits, which are already abundant, likely do not alter complex activity and therefore have minimal downstream consequences^57^. Future studies testing these hypotheses will help to clarify how *cis*-eQTLs might influence downstream processes.

Our study has several limitations. First, despite our aim to include diverse ancestries, most samples are still of European descent. This imbalance limits the generalizability of our findings to non-European populations and may cause us to miss the regulatory effects of variants that are rare or absent in European cohorts. Although large multi-ancestry biobanks have recently been established^58,59^, datasets that include both genotype and gene expression measurements across ancestries remain limited. Future efforts should prioritize generating paired genotype and gene expression data from underrepresented populations to enhance the equity and applicability of eQTL resources, as well as to generate knowledge that we may be missing by studying mostly European individuals.

Second, despite our efforts to harmonize analytical pipelines, residual heterogeneity across cohorts, genotyping platforms, and expression quantification methods likely led to conservative discovery and could affect fine-mapping results^60^.

Finally, while we corrected extensively for cell-type composition, some *trans*-eQTLs may still reflect subtle differences in cell proportions. Future studies incorporating high-resolution cell-type-composition data and mixed-model approaches will further improve eQTL resources.

Beyond those limitations, our study provides a valuable bridge between population genetics and data-driven modeling of gene regulation. Foundation models trained on millions of gene expression profiles require high-quality ground-truth relationships between genes to learn meaningful causal structures^61^. The eQTLGen regulatory maps offer precisely this: a large, experimentally grounded set of causal directed edges linking genetic perturbations to transcriptional responses. Integrating these networks into future foundation models will enable more interpretable and mechanistically faithful predictions of gene perturbations, cellular states, and disease mechanisms.

Together, these findings transform our understanding of how genetic variation orchestrates transcriptional networks and delineate which regulatory variants are functionally consequential. To our knowledge, this work provides the largest resource of *cis*– and *trans*-eQTLs to date, establishing a foundation for future genomics studies and offering insights into gene expression regulation and its role in complex disease biology.

## Methods

### Study population

The analysis consisted of 43,301 individuals from 52 individual datasets that had gene expression data available from whole blood or peripheral blood mononuclear cells (PBMC). The majority of contributing datasets were profiled using RNA-seq (37 datasets, N = 26,295, 61% of samples) and genotyped using genotyping arrays (43 datasets, N = 38,528, 89% of samples), with the remaining datasets profiled with expression arrays and whole-genome sequencing (WGS), respectively. Cohort-specific information sections are available in the **Supplementary Information**, and **Table S1** provides an overview.

All cohorts included in this study enrolled participants with informed consent and collected and analyzed data in accordance with ethical and institutional regulations. The information about individual institutional review board approvals is available in the original publications for each cohort and the **Supplementary Information**. Where applicable, data access agreements were signed by the investigators before acquisition of the data, and these state the data usage terms. To protect the privacy of the participants, data access was restricted to the investigators of this study, as defined in those data access agreements. Following the data use agreements, only summary-level data will be made publicly available. The protocol of the eQTLGen phase 2 study was reviewed by the Medical Ethics Review Board (METc) of the University Medical Center Groningen under research registry number 19149, which concluded that it is not a clinical research study with human subjects as meant in the Dutch Medical Research Involving Human Subjects Act (WMO). All procedures performed in studies involving human participants were in accordance with the ethical standards of the institutional and/or national research committee and with the 1964 Helsinki declaration and its later amendments or comparable ethical standards.

### Genotype quality control

Each cohort started with unimputed genotype data in plink.bed or.vcf format. Initial filtering, genotype quality control (QC) and analysis was conducted using R package bigsnpr v1.8.4^62^ (https://privefl.github.io/bigsnpr/index.html) and its interface to PLINK2^63,64^ (https://www.cog-genomics.org/plink/2.0/). Variants were filtered based on a minor allele frequency (MAF) > 0.01, a SNP missingness < 5%, and a Hardy-Weinberg disequilibrium P-value > 1×10^−6^. Samples with > 5% missing variants were removed from all subsequent analyses. If reported sex was available in the genotype.fam files, it was checked against the genetic data using the plink2 –-check-sex argument on the pruned set of SNPs (––indep-pairwise 50 1 0.2), and samples with an X chromosome heterozygosity value F between 0.2 and 0.8 were removed from the analysis. We also checked the samples for heterozygosity (plink2 –-het) and removed those that showed excess heterozygosity (>3 standard deviations (SDs) from the dataset mean). Because our analysis pipeline required non-related samples, the KING sample-relatedness metric^3^ threshold was set to 2^−4.5^ (corresponding to third-degree relatives) and one individual was removed from each pair of related samples.

Genotype samples were projected into the principal component (PCA) space of 1000 Genomes (1000G) p3v5^65,66^ samples, as provided by the bigsnpr package, using the function bed_projectPCA. Prior to the calculation of PCA space on 1000G samples, reference samples showing relatedness (both samples showed KING sample-relatedness < 2^−4.5^) were removed.

For each sample, the average Euclidean distance from that sample to each 1000G superpopulation was calculated based on the first three principal components (PCs). Samples that were genetic outliers or were not confidently assigned to any ancestry (average Euclidean distance to second most similar superpopulation < 2× smaller than to the most similar superpopulation) were removed from further analyses.

To remove individual genetic outliers from the analysis, the first 10 genotype PCs were calculated on the target dataset using the bigsnpr bed_autoSVD function, which removes long-range linkage disequilibrium (LD) regions before calculating PCs. Cohort analysts checked the first two genotype PCs and, if there was a small number of population outliers, removed them by selecting an appropriate threshold of SDs for which PC1 and PC2 could deviate from the mean of PC1 and PC2. Based on the calculated PCs, the sample “outlier-ness” score S^62^ was calculated for each sample, and cohort analysts were asked to assign a suitable S threshold based on the distribution of S values (default S threshold 0.4) and to remove potential outlier samples. Genotype PCs were recalculated after removal of population outliers and used as covariates in the association testing to remove the effect of population stratification.

If the data contained a considerable amount of samples from multiple superpopulations, we asked the cohort analysts to split the data into separate datasets and re-run genotype QC and subsequent analyses on separate subsets.

### Genotype imputation

We used the 1000G 30x WGS reference panel^67^ (all ancestries) to impute the autosomes of all genotype datasets. Genotype .bed files were lifted from hg19 to hg38 using CrossMap v0.4.1^68^. We used GenotypeHarmonizer v1.4.20^69^ to align strands to match the reference genome. Genotype data was converted to .vcf format using PLINK2^63,64^, and the BCFtools v1.10.2^70^ +fixref plugin was used to check if the reference and alternative alleles matched the reference panel. Filters were applied to filter out variants with MAF < 0.01, > 5% missingness, or a Hardy-Weinberg disequilibrium P < 1×10^−6^. Indels were removed, as were duplicate SNPs and variants with missing information about alleles. Only biallelic SNPs were kept, and genotype data was split into chromosomes. Genotype data was prephased using Eagle v2.4.1^71^ (http://data.broadinstitute.org/alkesgroup/Eagle/downloads/) and imputed using Minimac4^72^ (https://genome.sph.umich.edu/wiki/Minimac4). We filtered the imputed data to have a minimum MAF of 0.01. We used Genotype-IO v1.0.6 (https://github.com/molgenis/systemsgenetics/wiki/Genotype-IO) to export SNP QC metrics. These were used in the central site for further variant filtering.

### Gene expression QC and pre-processing

#### RNA-seq datasets

Raw expression matrices for RNA-seq datasets had been constructed by each cohort in their previous research, using variable tools and methods (**Supplementary Information**). These matrices were further filtered to include genes with counts per million (CPM) > 0.5 in at least 1% of the samples. The expression matrices were further normalized using the trimmed mean of M-values (TMM) method^73^ using R package edgeR v3.32.1. Before running association analysis, we forced all genes to be normally distributed by rank-based inverse normal transformation. For all analyses, we used ENSEMBL v106^74^ (ENSEMBL Release 106, 2022) stable gene IDs as the reference while converting. Other gene naming conventions were converted accordingly when necessary.

#### Mapping HGNC symbols to ENSEMBL gene identifiers

Some of the eQTLGen cohorts had expression count matrices available with HGNC gene symbols as gene identifiers. For these cohorts, we replaced the HGNC symbols with ENSEMBL gene identifiers by utilizing the biomaRt R package^75^ to access the ENSEMBL v109 database^74^ (ENSEMBL Release 109, 2022). First, we extracted all ENSEMBL gene identifiers, corresponding HGNC symbols, and external HGNC synonyms from the ENSEMBL database. Next, for each HGNC symbol present in the input data, we queried the overlap with ENSEMBL HGNC symbols and synonyms and mapped them to a corresponding ENSEMBL gene identifier. Some ENSEMBL gene identifiers had multiple corresponding HGNC symbols or synonyms available in the input data, resulting in duplicated ENSEMBL gene identifiers. For these cases, we confined the analysis to the latest ENSEMBL v109 HGNC symbol and discarded other genes mapping to corresponding ENSG ID. If the latest ENSEMBL v109 HGNC symbol for a given ENSG ID was not present in the input data, we confined the analysis to the random HGNC synonym mapping to the corresponding ENSEMBL gene identifier.

#### Array-based datasets

##### Empirical probe matching

For array-based datasets, we used the links from our previous study^12^, which used an empirical probe mapping approach to connect the most suitable probe to each gene that showed expression in the combined BIOS whole-blood expression dataset^76^. For the Illumina HT12v3, HT12v4, HumanRef-8, and Affymetrix U219 arrays, we included up to 13,287, 14,063, 9,230, and 14,220 unique probes, respectively, each corresponding to a single ENSEMBL GENE ID.

##### Illumina arrays

Raw expression matrices for Illumina arrays from HT12v3, HT12v4, and HuRef8 were quantile-normalized using the R package preprocessCore v1.52.1. Before running association analysis, we forced all genes to be normally distributed by rank-based inverse normal transformation.

##### Affymetrix arrays

For the Affymetrix U219 array platform used by the NTR-NESDA cohort, we used an already pre-processed, RMA-normalized expression matrix, as detailed in^6^. Before running association analysis, we forced all genes to be normally distributed by rank-based inverse normal transformation.

##### Gene expression QC

For each cohort, we calculated the first four gene expression PCs on the normalized and log_2_-transformed expression matrix and visualized those. We removed outlier samples that deviated by more than 3 SD from the median of any of the PCs, and expression data was re-preprocessed after outlier samples were removed. We did not apply any strict thresholds for expression levels. Instead, we collected the two sets of per-gene summaries (mean, median, SD, minimum, maximum, number of unique values) calculated on raw and normalized and log_2_-transformed expression data from each cohort and used them to apply per-cohort gene filters in the central analysis site.

##### Sample mix-ups

To check potential sample mix-ups, we visualized sex-specific gene expression by plotting the mean expression of genes encoded from chromosome Y against the expression of the gene *XIST*, which is encoded from chromosome X^77^. Genetic sex calculated by PLINK (v1.9) was then compared against sex-specific gene expression, and the sample mix-ups identified were either solved or removed from further analyses.

### Per-cohort association analyses

Running genome-wide eQTL analyses in consortium settings poses several technical and organizational challenges. Usually, it is not possible to collect primary genotype and gene expression data into a central site and analyze them jointly (mega-analysis) due to privacy concerns, institutional data protection procedures, and legislation. The standard way of conducting GWAS meta-analyses is to perform GWAS for each dataset and then share association summary statistics for meta-analysis at the central site. However, for eQTL analysis, this would mean that each cohort would need to share full summary statistics from >15,000 phenotypes (blood-expressed genes), each of which would be several terabytes of data. This makes the classical meta-analysis approach impractical for consortium-based eQTL meta-analyses. While it is also possible to use a multi-phase approach to identify associated regions and perform locus-wide association analyses in the second phase^78^, this would not yield a complete association profile for each gene, which is desirable for a number of computational approaches that integrate GWAS and QTL data.

To address these challenges, we customized the HASE approach^14^ to prepare genotype, gene expression, and covariate data for each cohort. This method calculates encoded data matrices from raw genotype and gene expression matrices by multiplying those with a random matrix F and the inverse of this matrix F, respectively, and calculates partial derivatives from covariate data. Because the random matrix F is not stored, it is not possible to obtain any information for individual study participants from the encoded matrices. However, it is still possible to use the encoded matrices to calculate association statistics for each variant and gene combination. We used the Python package PyCryptodome (pycryptodomex v3.9.7) for additional cryptographic security when generating the random matrix F and replaced the original sample IDs in the encoded matrices with the cohort name and an index number. This allowed us to circumvent sharing of many very large summary statistic files because the encoded matrices and partial derivative files are much smaller and use an efficient file format (Hierarchical Data Format v5: HDF5).

Each cohort provided the partial derivatives for the following covariates: the first 10 genotype PCs and the first 100 gene expression PCs. Importantly, the HASE method gave us the flexibility to select the most appropriate set of covariates to include in the final encoded meta-analysis in the central site. Each cohort also permuted the links between genotype and expression samples and provided information for one permutation round.

To perform QC in the encoded meta-analysis, each cohort calculated summary statistics for each variant (allele frequency, number of carriers for each genotype, imputation Mach R^2^, Hardy-Weinberg disequilibrium P-value, and whether it was imputed or typed) and for each gene (on normalized and log_2_-transformed expression matrix: minimum, maximum, mean, median, SD, number of unique values).

Per-cohort encoded matrices, partial derivative files, and summary data files for genes and variants were stored and analyzed in the restricted access group in the University of Tartu High Performance Computing Centre^79^.

### High-dimensional encoded meta-analyses

We customized the HASE encoded mega-analysis approach^14^ and used it to conduct high-dimensional genome-wide eQTL meta-analyses. We devised a pipeline that conducts inverse-variance weighted meta-analyses for every gene in a parallelized manner. For each gene, it first runs the HASE encoded analysis on each cohort and extracts the β and standard error (SE) for every variant. It then compares the results with the per-cohort SNP and gene QC metrics and sets those per-cohort associations to missing when a variant fails QC (Mach R2 < 0.4, Hardy-Weinberg equilibrium (HWE) P < 1×10^−6^, MAF < 0.01, or MAC ≤ 10) or when a gene is lowly expressed (fraction of zero values was ≤ 0.8). It then runs inverse-variance meta-analysis on the remaining cohorts and reports the meta-analysis β, SE (β), and the heterogeneity statistic I2. We included four genetic PCs, 20 gene expression PCs, and other technical covariates (e.g. RNA integrity number) where available. Covariates were pruned to prevent any collinearity among them (**Table S12**). For European ancestry datasets, we restricted the analysis to variants where the allele frequency (AF) was within 0.2 of the AF from the European subset of samples in the 1000G 30x WGS reference panel. In an additional step, genes were filtered to include only those available in at least 50% of the cohorts and 50% of the RNA-seq samples. Variants were also filtered for those available in at least 50% of the cohorts.

### Empirical gene−gene co-expression and LD

Since we did not have access to individual-level genotype data from the contributing cohorts, we leveraged summary statistics from a permuted eQTL meta-analysis. These results, generated under the null hypothesis of no association between variants and gene expression, can be represented as a matrix of effect sizes (e.g., β-values or Z-scores) across all variant–gene pairs. Because the data are permuted, any observed associations are expected to reflect random noise rather than true biological signal. We hypothesized that this noise is not entirely independent across variants. Instead, due to LD patterns and gene−gene correlations, the noise may exhibit correlation between variants as well as genes. By analyzing these correlations in the permuted summary statistics, we aimed to approximate the underlying genotype correlation structure (i.e., LD) and gene expression correlations without requiring individual-level data.

To calculate the LD and gene−gene correlations, we continued with our permuted variant−gene summary statistics. To ensure that the association metrics in these summary statistics were independent of sample size, we first converted the β-values and SE to Pearson correlation coefficients *r* using the transformation:

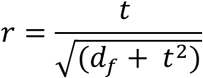

where *t* is the t-statistic and *d_f_* is the degrees of freedom (sample size – 1). This transformation yields a variant−gene correlation matrix that serves as the basis for both the LD and gene−gene correlation analyses.

### Deriving gene−gene correlations

To calculate gene−gene correlations, we selected the 6,268,843 variants that were available for at least 95% of all eQTLGen samples (n = 41,136) and further sub-set these to 28,417 approximately uncorrelated variants using the 1000G 30x WGS reference panel (MAF ≥ 1%, missingness ≤ 1%, HWE P-value > 0.05, spaced apart by at least 1000 bp, exclusion of the HLA region, and in approximate linkage equilibrium using a maximum R^2^ of 0.02 in the 1000G 30x WGS reference data). Using the variant−gene correlation matrix derived from the permuted summary statistics, we treated the 28,417 uncorrelated variants as observations and computed pairwise correlations between all genes. This resulted in an empirical gene−gene correlation matrix that reflects co-expression patterns.

### Deriving in-sample LD estimation

Analogous to the gene−gene correlation analysis, we estimated LD by correlating variants with each other. However, to better utilize the full set of gene−variant associations and reduce redundancy among genes, we first orthogonalized the gene dimension using PCA. We performed PCA on the variant−gene correlation matrix from the permuted data, using only the 28,417 uncorrelated variants. To orthogonalize the observations (genes), we projected the matrix onto the eigenvectors and scaled the resulting PCs by their corresponding singular values. We limited this analysis to the 10,497 genes with an average eQTLGen sample size of at least 90%. The final LD matrix was computed by correlating variants across the rows of the transformed matrix, where the columns now represent orthogonalized PCs. This approach allowed us to estimate in-sample LD using only summary-level data. The idea of using large-scale summary statistics data under the null hypothesis to allow for calculating LD was also recently proposed by others^80^ specifically to circumvent having to share privacy-sensitive genotype data.

Fine-mapping of eQTL effectsSignificant eQTL variants were identified using a genome-wide significance threshold of 5×10^−8^. For each significant variant, we defined a genomic window of ±500 kilobases (Kb). Overlapping windows were merged per gene to form a set of non-overlapping loci. Additionally, loci were combined if the distance between them was ≤ 2 megabases (Mb). For each locus, a derived eQTLGen in-sample LD matrix was constructed and used as an additional input.

Fine-mapping was performed using the SuSiE (Sum of Single Effects) with Regression Summary Statistics (RSS) model, as implemented in SuSieR v0.12.35 R package^81–83^. Within each locus, we included only variants with at least 80% of the maximum sample size in that locus. To ensure convergence, we implemented a two-stage fallback strategy. In our primary attempt, SuSiE RSS was run iteratively across a predefined set of credible set (CS) numbers (10, 5, and 3) with estimate_residual_variance = TRUE. If the model converged and identified at least one non-empty CS, the result was accepted and the loop exited. In our fallback attempt, if no successful result was obtained in the primary attempt, the same loop was repeated with estimate_residual_variance = FALSE. Again, the loop exited upon successful convergence and CS detection. If all attempts failed, the variant with the maximum absolute Z-score was selected as the single lead variant for that locus−gene combination. We note that the lead variants selected by this final fallback procedure have not been included in the colocalization analyses and the interpretations thereof.

### CS filtering and post-processing

We retained CSs where the lead variant (the variant with the highest posterior inclusion probability (PIP)) had a log Bayes factor (LBF) > 2 and a nominal P-value < 1×10^−5^. We removed *trans*-eQTLs CSs that likely suffered from cross-hybridization events. Variants located within the HLA region (chr6:25,000,000-35,000,000) were also removed due to this region’s complex LD structure. Finally, we clumped eQTLs per gene using a maximum pairwise R^2^ threshold of 0.9, prioritizing variants with a higher LBF over those with a lower LBF.

### Removal of cross-hybridization events

Some *trans*-eQTL associations may arise from cross-hybridization artifacts due to sequence similarity, where sequencing reads originating from one gene (the focal gene) are incorrectly mapped to another gene in the case of RNA-seq data or where transcripts of another gene are hybridized to probes of the focal gene in the case of microarrays. If the focal gene is genetically regulated in *cis*, these misalignments can result in the detection of both a true *cis*-eQTL for the focal gene and a spurious *trans*-eQTL for the gene to which the reads were incorrectly mapped. These false positive *trans*-eQTLs driven by technical cross-mapping or hybridization rather than genuine biological regulation must be identified and filtered out to ensure the integrity of downstream analyses^56^.

For this purpose, we generated 35 bp single-end synthetic reads from the ENSEMBL^74^ v106 GRCh38 gene annotation (GTF), using the corresponding human genome reference sequence. To capture exon−exon junctions, additional sequences were created to span these boundaries. A sliding window approach was employed across the exonic regions of each gene, with a 10 bp shift between consecutive windows. This resulted in a staggered set of synthetic reads for each of the *trans*-eGenes, which were compiled into FASTA format.

For each *trans*-eQTL variant, a 10 Mb window centered on the variant position (5 Mb upstream and downstream) was extracted from the reference genome to construct localized reference sequences. These variant-specific reference genomes were indexed to facilitate alignment.

Each *trans*-gene-specific FASTA file was aligned to its corresponding variant reference genome using BWA with permissive alignment parameters to accommodate sequence divergence. This alignment strategy enabled assessment of read-mapping behavior in the context of local genomic variation. Ultimately, *trans*-eQTLs were flagged as cross-mapping effects if on average 5% of the *trans*-gene read bases (averaged over all generated reads) could be mapped within the 10 Mb window surrounding the *trans*-eQTL variant.

### Comparison with eQTLGen phase 1

We obtained eQTLGen phase 1 summary statistics and allele frequencies from eqtlgen.org^6^. Both the complete summary statistics and the QC-filtered, significant subset were harmonized to enable direct comparison with eQTLGen phase 2. To achieve this, we lifted all results to the hg38 genome build and matched variants by chromosome, position, and alleles. Effect directions were swapped when the tested allele in one dataset matched the reference allele in the other dataset. We estimated *β* and SE(*β*) for every eQTL using the equation by Zhu et al., 2016^84^ (See **Correlations between effect sizes**). For each gene, we retrieved all QC-filtered lead *cis*-eQTLs from eQTLGen phase 1 and searched for their counterparts in phase 2. Similarly, we retrieved all QC-filtered *trans*-eQTLs from phase 1. Conversely, we looked up all fine-mapped lead *cis*– and *trans*-eQTLs from phase 2 in phase 1. For all comparisons, we assessed concordance by plotting Z-scores against each other, computing Pearson correlation coefficients and R_b_ metrics. These indicated that concordance is as expected (**Figure S20**).

### Per-cohort effects

To identify if our meta-analysis result was not solely driven by individual cohorts, we reran our implementation of HASE to export summary statistics for all our fine-mapped *cis*– and *trans*-eQTLs on a per-cohort level. For each cohort we assessed concordance to the meta-analyzed results by plotting Z-scores of the meta-analysis against the individual cohort, calculating the percentage of concordant effects and computing linear regression slopes over the Z-scores. In general, concordance was shown to be as expected. Smaller cohorts of underrepresented populations and those that used microarrays data had the lowest concordance. Large European ancestry cohorts in which expression was measured using RNA-sequencing had the largest concordance (**Figure S21** and **Figure S22**).

### Testing the effect of gene expression abundance and evolutionary constraint on eQTL discovery

To assess gene expression abundance, we calculated the median expression per gene within each cohort. This was done at each of the cohorts within the DataQC pipeline described above. Gene ranks across all cohorts were then aggregated using the robust rank aggregation method, as implemented in the RobustRankAggreg R package v1.2.1^85^. Following the ranking, all genes were divided into 10 equally sized bins. For each bin, we quantified the number of CSs associated with the genes it contained, enabling comparison of CS distribution across the gene expression spectrum.

Gene-level constraint metrics were derived from the gnomAD v4 gene constraint dataset^15,16^. For each gene, we selected the constraint values corresponding to the MANE (Matched Annotation from NCBI and EMBL-EBI) transcript, where available. If no MANE transcript was annotated, we used the most extreme constraint value across all transcripts: the maximum for pLI and loss-of-function (LoF) Z-score and the minimum for LOEUF.

To evaluate the relationship between gene constraint and eQTL architecture, we performed the following statistical analyses. The correlation between the number of independent eQTLs per gene and LOEUF was assessed using Spearman’s rank correlation. The association between the presence or absence of any eQTL and pLI was tested using the Mann-Whitney U test. The correlation between the significance of the most significant *cis*– and *trans*-eQTL per gene and the LoF Z-score was evaluated using Pearson’s correlation test.

### Enrichment analyses on eQTL variant properties

Enrichment analyses of basic variant properties (variant consequences and variant classes) were carried out by matching each eQTL variant to 100 randomly sampled control variants from a set with identical transcription start site (TSS) density, an LD score within 0.1 SD of the focal variant (calculated among all variants), and a MAF within 0.02 of the focal variant. TSS distances were calculated using a radius of 500 kb, using the TSS of the genes assessed in eQTLGen phase 2. LD scores were calculated using the 1000G 30X WGS reference panel and LD score regression software^48^. The sampling of control variants was done with replacement. We dropped 246 out of 57,806 *cis*-eQTL variants and 200 out of 20,840 *trans*-eQTL variants from the enrichment analyses because there were fewer than 10 eligible control variants.

#### Variant classes

Variant classes were derived from the 1000G 30X reference dataset. Insertions and deletions were annotated by taking the effect of the alternative allele compared to the reference allele. HGSV structural variant classes^86,87^ were taken from the allele annotations INS, DUP, or DEL.

#### Variant consequences

Variant consequences were predicted for every independent fine-mapped variant using ENSEMBL VEP (cache version v111)^21^, run through the VIP variant interpretation pipeline (v8.3.1)^88^.

#### Distance of eQTLs and GWAS variants to nearest TSS

We replicated the work of Mostafavi et al., 2024 with regards to the comparison of *cis*-eQTLs and GWAS hits in terms of distance to their nearest TSS^22^. We used the annotations and GWAS variants provided by the authors while replacing their *cis*-eQTLs with the *cis*-eQTLs and *trans*-eQTLs identified by the current study.

#### Replication analyses and lookups

##### Blood single-cell eQTL data from the single-cell eQTLGen consortium

The single-cell eQTLGen (sc-eQTLGen) consortium provided replication of the eQTLs we discovered^23^. Specifically, within sc-eQTLGen, 12 PBMC eQTL datasets were harmoniously reprocessed and integrated leveraging a meta-analysis framework. The complete dataset comprises 2,032 individuals post-QC, and analyses within the consortium were performed at the level of the six major PBMC types (CD4+ T cells, CD+ T cells, B cells, dendritic cells, monocytes, and natural killer cells). Details are provided in the sc-eQTLGen manuscript^23^. Here, we replicated all *cis*– and *trans*-eQTLs by taking the lead eQTL variant per CS (largest PIP or the eQTL variant with the largest absolute Z-score for loci in which fine-mapping failed). Replication was performed within the six major cell types, focusing on the 1,323 samples with European descent that were assessed using the 10X technology on whole PBMCs. Within the sc-eQTLGen consortium, genes were deemed expressed if they were non-zero in each of the datasets (per cell type), and SNPs were only considered if they passed imputation QC, had a non-zero genotype count for each genotype in each separate dataset, and a MAF > 1% in the dataset. For the replication, we required that the SNP−gene combination was observed in at least three of the datasets and had an I^2^ < 40%.

##### Brain single-cell eQTL data from the single-cell MetaBrain project

Brain single-nucleus data was collected from 10 different brain cortex datasets in the context of the single-cell MetaBrain project (scMetaBrain)^24^. In short, after QC, 3.8 million nuclei were used to create pseudobulks for eight major cell types (excitatory and inhibitory neurons, microglia, astrocytes, oligodendrocytes and their precursor cells, and endothelial cells) for up to 757 donors, depending on the cell type. Each pseudobulk was consequently normalized and corrected for dataset label indicators and 98 PCs that were calculated over the sample−sample correlation matrix. Genotype data were mostly acquired from WGS datasets, which were processed per genotype dataset. Genotype processing used a similar pipeline to that used for eQTLGen phase 2, with similar filter settings and genotype imputation (based on GRCh38, 1000G 30x WGS). We tested variants with a call-rate > 60% and a HWE P-value > 1×10^−5^. Testing of eQTLGen *cis*– and *trans*-eQTL variant–gene pairs was performed using Spearman correlation for variants with MAF > 1%. Subsequently, *β*s and SE(*β)* were estimated from the Z-scores using the formula from Zhu et al., 2016^84^ (see **Correlations between effect sizes**).

##### Bulk eQTL data from various GTEx tissues

WGS-based genotype data were processed as part of this project. GTEx v8 gene expression matrices for non-blood tissues were downloaded from the GTEx portal website, together with their respective covariate tables, which included sequencing batch labels, PEER factors, and other covariates^5^. For each tissue, the RNA-seq data were corrected for these covariates, and eQTLgen *cis*– and *trans*-eQTL variant–gene pairs were subsequently tested using Spearman’s correlation for variants with MAF > 1%. Subsequently, *β*s and SE(*β)* were estimated from the Z-scores using the formula from Zhu et al., 2016^84^ (see **Correlations between effect sizes**).

##### Brain bulk eQTL data from Metabrain

Bulk brain cortex eQTL data was collected from 14 datasets in the context of the MetaBrain project^13^. In short, RNA-seq datasets were re-downloaded and realigned, and read counts were counted in a harmonized manner. Genotype data was filtered, harmonized, and imputed using 1000G v3p5 and HRC imputation reference panels, excluding indels. For the purpose of replication in this study, RNA-seq data for the brain cortex region with matching genotype data was combined for 2,759 donors, which represented the Cortex-EUR + Cortex-AFR datasets from the MetaBrain manuscript^13^, minus the ENA dataset. RNA-seq data was corrected for four genetic PCs, dataset labels, 20 technical covariates, and 100 expression PCs. We tested variants with a call-rate > 60%, a minimum genotype count of 5, and a HWE P-value > 1×10^−5^. *cis*– and *trans*-eQTL variant–gene pairs were tested using Spearman’s correlation for variants with MAF > 1%. Subsequently, *β*s and SE(*β)* were estimated from the Z-scores using the formula from Zhu et al., 2016^84^ (see **Correlations between effect sizes**).

##### Plasma pQTL data from UK Biobank Pharma Proteomics Project

To replicate our fine-mapped eQTLs among pQTLs, for every eQTL, we extracted the corresponding pQTL summary statistics from the UK Biobank Pharma Proteomics Project^26^. First, we downloaded the discovery pQTL summary statistics data using the Python Synapse client (v4.8.0). All summary statistics were then harmonized to those in the 1000G 30X reference panel as described in the GWAS processing section. After matching, pQTL effect sizes were flipped as needed to ensure alignment of the effect allele with that used in eQTLGen Phase 2. To perform the lookup for the correct gene−protein pairs, we matched our eQTL genes to the corresponding pQTLs based on the ENSEMBL^74^ gene IDs in the olink_protein_map_3k_v1.tsv file downloaded with the pQTL summary statistics.

##### Correlations between effect sizes

To estimate the correlation between eQTLGen eQTLs and the corresponding replication or lookup dataset, we used the R_b_ metric^89^, which is robust to sample size differences between the discovery and replication dataset. We clumped per-gene eQTL lead variants (LD R^2^<0.1; eQTLGen empirical LD), prioritizing eQTLs by (1) LBFs, and (2) absolute Z-scores, to remove partly correlating eQTL effects before calculating R_b_. To calculate a P-value for the R_b_ metric, the Z-score was first calculated by dividing the R_b_ by SE(R_b_) and then squared to calculate the Chi^2^ statistic. The P-value was then derived from the Chi^2^ distribution with one degree of freedom.

The average eQTL effect between different cell types of sc-RNA-seq datasets was calculated by first calculating the replication Z by dividing the replication *β* by SE(*β*). The average replication Z was then calculated over different cell types. Finally, average replication *β* and SE(*β*) were estimated from the Z-score, using following formula from Zhu et al., 2016^84^:

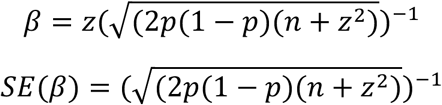

where *p* is the MAF, *n* is the sample size, and *z* is the average Z-score over individual cell types. The MAF used was calculated over eQTLGen samples, and we used fixed sample sizes of 1,323 for the sc-eQTLGen blood dataset and of 757 for the scMetaBrain brain dataset. For these analyses we excluded the 916 trans-eQTLs for which the gene-variant distance was < 5 Mb.

##### Defining independent groups of *cis*– and *trans*-eQTL signals

To enable us to calculate variant-level statistics (such as those presented in **Figure S11**), we performed an exhaustive colocalization analysis, colocalizing all variants overlapping eQTL, *cis* and *trans*, searching for dependance between genetic signals. Here we defined independent sets as exclusively non-colocalizing groups of eQTL signals. This approach allowed us to identify which eQTLs likely originated from the same underlying genetic effect. For each locus, we extracted the LBFs from the output of fine-mapping and used the coloc.bf_bf command from coloc v5.2.3^90–93^ with default settings (p1=1e-4, p2=1e-4, p12=5e-6, overlap.min=0.5, trim_by_posterior=TRUE, prior_weights1=NULL, prior_weights2=NULL) to conduct colocalization analyses. Coloc was only assessed if at least one of the variants in the credible set was kept after SNP overlap. Among all colocalizing credible set (CS) pairs (colocalization PP4 > 0.8), we then identified communities of colocalizing CSs in an exhaustive manner, such that all CSs connected by at least one colocalization were grouped together. This identified 42,166 likely distinct genetic effects underlying *cis*-eQTLs and 6,224 likely distinct genetic effects underlying trans-eQTLs. We refer to these as independent *cis*– and *trans*-signals or variants.

##### Prediction of blood RNA expression of fine-mapped *cis*-eQTLs using Borzoi

To predict the effect of fine-mapped eQTLs using Borzoi, we took the *cis*-eQTL variants in a CS with a PIP ≥ 0.9 as determined by SuSiE, replicating the strategy proposed by Linder et al.^46^. Furthermore, we sub-set this set of fine-mapped *cis*-eQTLs to single base pair substitutions only, as the distribution of predicted effects of indels is not comparable to the distribution of predicted effects of SNPs^94^. We predicted the variant effect (referred to as “diff_score”) as previously described by Linder et al.^46^, specifically predicting the ‘logSED’ (‘sum of expression differences’) score for the RNA blood tracks, averaging over multiple tracks. We defined a variant as concordant with Borzoi when the sign of the prediction matched the sign of the β as observed for the *cis-*eQTL.

#### Inference of directed gene regulatory relationships

##### *Cis−trans* colocalization analysis

We utilized the output of the eQTL fine-mapping to conduct statistical colocalization analyses. Although the *coloc* method was originally devised for independent samples, recent simulations have shown it is also suitable for colocalizing different traits (i.e. different genes) measured from the same individuals^31^. For each *cis*-eQTL locus, we determined overlapping *trans*-eQTL loci and enrolled those in the analyses. For each locus, we extracted the LBFs from the output of fine-mapping and used the coloc.bf_bf command from coloc v5.2.3^90–93^ with default settings (p1=1e-4, p2=1e-4, p12=5e-6, overlap.min=0.5, trim_by_posterior=TRUE, prior_weights1=NULL, prior_weights2=NULL) to conduct colocalization analyses in an iterative manner. Because fine-mapping was done for up to 10 independent signals per locus, the colocalization analyses allowed us to test for pairwise colocalization between all independent signals of two overlapping loci.

Subsequently, we filtered the colocalization result by requiring at least 500 variants in the colocalization analysis and that lead variants of colocalizing signals are in high LD (R^2^ > 0.8, empirical in-sample LD). We further only reported colocalization events that passed the same QC criteria we imposed on the fine-mapped CSs, that survived the cross-mapping filter, and for which the lead variant of the CSs matched the lead variant reported in the colocalization output. We also required that *cis*– and *trans*-eQTLs genes are at least 5 Mb apart.

##### Inference of activating and repressing directed gene−gene relationships

For the colocalizing *cis*– and *trans*-eQTLs, we determined whether the *cis*-gene activated or repressed the *trans*-gene. If the SNP allele that increased the expression of the *cis*-gene also increased the expression of the *trans*-gene, we concluded that the *cis*-gene activated the *trans*-gene. If the SNP allele increased the expression of the *cis*-gene but repressed the expression of the *trans*-gene, we concluded that the *cis*-gene repressed the *trans*-gene. For 682 gene pairs, multiple CSs affected both the *cis*– and *trans*-gene. For these pairs, we checked whether the predicted activating or repressing effect was consistent for the different CSs. This was the case for 614 of the 682 gene pairs (90%). We removed the 68 gene pairs with conflicting directions, yielding a final set of 18,327 activating gene pairs and 20,111 repressing gene pairs, reflecting 7,965 unique genes (2,619 unique genes that affect another gene and 6,542 unique genes that are affected by another gene).

##### Enrichment analysis of properties of *trans*-acting genes

We studied the 2,619 unique genes that were affecting other genes in *trans*. We performed Gene Ontology ‘molecular function’ enrichment analysis using ToppGene^28^, using all genes for which we identified a *cis*-eQTL as a background set. Strong enrichment was observed for genes with known transcription factor binding activity and for genes that are transcriptional coregulators. For the *trans*-acting genes that were affecting multiple genes, we determined whether they were preferentially activating or repressing other genes by conducting a binomial test per *trans*-acting gene. We converted this binomial test P-value to a Z-score. This permitted us to test whether transcription factors that affect multiple *trans*-genes significantly more often preferentially activate or preferentially repress other genes compared to *trans*-acting genes that are not a transcription factor (Wilcoxon test on absolute Z-scores). For the *trans*-acting genes that were preferentially activating or repressing *trans*-genes (binomial test P < 0.05), we used ToppGene to determine enrichment for Gene Ontology ‘biological process’ terms.

##### STRING

The STRING database of physical and functional interactions between proteins integrates multiple sources and assigns a confidence score for the interactions^30^. We downloaded the full network with scored links for selected organism *Homo sapiens* (9606.protein.links.full.v12.0.txt) from the STRING download portal (https://string-db.org/cgi/download; accessed December 11, 2024). We mapped ENSP to ENSG identifiers with the mygene Python package (version 3.2.2)^95^. When the *cis*– and *trans*-genes of a pair were found to code for two proteins with an interaction in the STRING database, the confidence score provided was used.

##### Remap

We used the ReMap 2022 database of transcription-regulator binding sites derived from manually curated ChIP-seq experiments^31^. We downloaded the *Homo sapiens* (hg38) BED file with 68.2 million non-redundant peaks (remap2022_nr_macs2_hg19_v1_0.bed) from the ReMap download page (https://remap.univ-amu.fr/download_page, accessed December 24, 2024). We filtered for binding sites that overlap with regions from 200 bp upstream to 75 bp downstream of the TSS for annotated genes and transcripts located on chromosomes 1−22, X, Y, and MT, using the hg38 annotation file (Homo_sapiens.GRCh38.106.gtf) and the pyranges Python package (version 0.1.2)^96^. We then filtered to retain data obtained in blood-related cell types (K-562, GM12878, THP-1, U937, HL-60, NB4, Jurkat, Raji, Daudi, MOLT-4, Jeko-1, OCI-Ly7, and MM1.S).

##### Processing the Replogle/Ota dataset

We downloaded the Perturb-seq summary statistics (P-value and log fold-change (logFC) matrices) accompanying the preprint by Ota et al. (2025) from Zenodo^32,97^. This data is based on the Replogle et al. (2022)^98^. study using the K562 human myelogenous leukemia cell line. The matrices have 9,866 columns with CRISPRi perturbed genes and 8,428 rows with genes for which the effect on expression was quantified. We mapped *cis*-genes to the perturbed genes of the Perturb-seq dataset and *trans*-genes to affected genes from the Perturb-seq dataset. For combinations present in the Perturb-seq matrices, we extracted the P-value and logFC values for that gene pair. We then corrected for the extracted P-values across all overlapping gene pairs, regardless of their significance using a Benjamini Hochberg correction. We then filtered out the gene pairs with an FDR>0.05.

##### Processing the Atlas/Orion 2025 dataset

To be able to overlap our *cis*−*trans*-gene pairs with a second Perturb-seq dataset, we obtained the X-Atlas/Orion Perturb-seq dataset^33^ and processed it in a similar way as the Ota et al. (2025)^32^ dataset was processed. This Perturb-seq dataset was obtained from experiments in the HCT116 cell line. We first defined a list of expressed genes. Expressed genes were defined according to Huang et al. as “genes that collectively account for 99% of the cp10k (counts per 10,000) counts averaged across cells containing a non-targeting sgRNA pair”^33^. 11,200 genes were identified for the HCT116 perturb-seq dataset.

Because non-targeting guides may still induce subtle perturbation-associated phenotypes, we defined a core control group for the rest of the analysis. For this step, we took all the cells hit by the non-targeting guides. For each non-targeting guide, we compared the gene expression profile of the cells carrying this guide, to a background set made from all the others non-targeting guides cells. After log-normalizing the gene counts, we ran a differential gene expression analysis using Model-based Analysis of Single-cell Transcriptomics (MAST)^99^. MAST is a two-part generalized linear model that models both the probability of a gene being expressed, using a logistic regression model, and the positive expression mean, using a Gaussian linear model. P-values were adjusted using a Benjamini-Hochberg correction. The non-targeting guides with a detectable phenotype, defined as controls that had more than 8 differentially expressed genes, were filtered out. 692 non-targeting guides were retained for our core control set for the rest of the analysis.

Next, we computed a differential gene expression analysis for all the perturbed guides. Following the Replogle et al.^98^ pipeline, guides were defined as eligible if (i) they were expressed within more than 25 cells and (ii) for which each cell expresses at least 500 genes. Because the number of cells were varying across the perturbations, we ran a Limma Voom differential gene expression analysis^100^, a linear model that accounts for library size variabilities. For each perturbation, we selected the 2,000 most variable genes using Seurat flavor v3^101^ and compared log-normalized counts in perturbation cells to those in the core control group. We used the UMI, the batch correction and the percentage of mitochondrial genes as covariates. Finally, we generated two tables, one for the log_2_-fold changes (logFC) and one for the adjusted p-values of the downstream genes per guide. After doing so, 7,470 perturbations remained.

We then used these tables to overlap the *cis*–*trans*-gene pairs, using the same procedure as applied to the Ota et al. study^32^.

##### STRING, Remap, and Perturb-seq enrichment analysis

For each of the gene pairs where a *cis*– and *trans*-eQTL colocalized, we determined whether there was overlap with the STRING, ReMap, and Perturb-seq data. To establish whether we observed more overlap than expected by chance, we first defined a set of positive directed gene−gene relationships (i.e. a *cis*-eQTL gene that colocalizes with a trans-eQTL gene). Because some *cis*-genes were linked to hundreds of *trans*-genes, which could otherwise dominate the signal, we limited the number of pairs per *cis*-gene to a maximum of 10. This resulted in a balanced, diverse set of 10,406 positive pairs representing 2,619 unique *cis*-genes.

We subsequently constructed a carefully matched set of 10,406 negative pairs that preserved the same degree distribution as the positives—each *cis*-gene and *trans*-gene occurred the same number of times as in the positive set, but their pairings were shuffled. The negative set thus contains the same 2,619 unique *cis*-genes present in the positive set. This matching strategy ensures that any difference in overlap between the STRING, ReMap, or Perturb-seq relationships with the positives and negatives is not driven by the fact that certain *cis*– or *trans*-genes are more often present or absent in these resources.

Finally, we determined enrichment of overlap by performing a Chi^2^ test on the fraction of positives that overlap STRING, ReMap, or Perturb-seq and the fraction of negatives that overlap these repositories. In figure 4f, these Chi^2^ P-values and odds ratios are shown, but the overlap of STRING, ReMap, and Perturb-seq relationships shown are based on all 38,438 gene pairs. The expected overlap, shown in figure 4f, is based on the odds ratio we observed in the abovementioned enrichment analysis.

#### eQTL−GWAS colocalization analysis

##### GWAS selection

To enable eQTL−GWAS colocalization analyses, we compiled a set of 87 traits and diseases from multiple sources. We first included 60 traits and diseases reported by Zhang et al.^102^, selected based on clinical relevance (particularly immune-mediated conditions), statistical power (heritability Z-score > 5), and availability of summary statistics. For these traits, we updated the GWAS summary statistics when more recent versions were available. In addition, we incorporated five cardiovascular GWASs, selected as potential positive controls based on their relevance in whole blood. Finally, we included 23 GWASs related to cell-type composition that were used by Vuckovic et al.^103^ to assess the potential influence of cell-type heterogeneity on *trans*-eQTL signals. A list of the traits and GWAS summary statistics is available in **Table S8**. Celiac disease was excluded because the largest GWASs for this trait have a limited number of variants available.

##### GWAS processing

All GWAS summary statistics were harmonized to the GRCh38 (hg38) genome build by performing a LiftOver^104,105^ when necessary. Effect sizes were standardized to either β coefficients or log odds ratios and converted accordingly if originally reported in a different format. Confidence intervals were converted to SE where applicable. Variants were matched to those in the 1000G 30X reference panel based on genomic coordinates and both alleles. Multiallelic indels were excluded if an insertion and a deletion shared the same genomic position and alleles. We note that these variants are not duplicates, they only appear identical due to indel allele normalization. However, we excluded this limited set of variants from the matching process because they can result in incorrect variant matchings. After matching, GWAS effect sizes were flipped as needed to ensure alignment of the effect allele used in eQTLGen Phase 2.

For those GWASs in which an effective sample size was not reported, we calculated it using the equation:

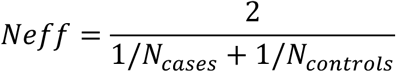

where *N_eff_*, *N_cases_*, and *N_controls_* represent the effective sample size, the number of cases, and the number of controls, respectively.

Fine-mapping was performed using the same locus definitions as for eQTLs, with one exception. To avoid fine-mapping prohibitively large loci, we did not merge loci if their distance was < 2 Mb. Within each locus, we included only variants with at least 80% of the maximum sample size in that locus. We also removed variants with β = 0 or SE(β) = 0 and those whose association Z-score was substantially different from what was expected based on the LD reference. To do the latter, we utilized the function kriging_rss from the SuSieR package, while specifying n as median sample size over the locus. We then extracted the standardized Z-score differences per each variant and converted them to P-values. We removed all variants with a Bonferroni-corrected P-value ≤ 0.05, while using the number of variants in the locus for multiple testing. We estimated the phenotypic variance using the sdY.est function from the coloc v5.2.3 package.

Subsequently, we used the SuSiE model, as implemented in SuSieR v0.12.35 R package^81,82^, to conduct fine-mapping for each GWAS locus, using the derived eQTLGen out-of-sample empirical LD matrix as a reference. Fine-mapping was conducted in iterative fashion across a predefined set of CS numbers (10, 5, and 3) with estimate_residual_variance = FALSE (recommended setting when the out-of-sample LD matrix is used). If the model converged and identified at least one non-empty CS, the result was accepted and the loop exited early. If the model did not converge or no CSs were identified, fine-mapping was performed only on the primary GWAS signal using the finemap.abf function from the coloc v5.2.3^90–93^ package.

Colocalization analyses and post-hoc filtering were done using the same setup as the *cis−trans* colocalization analyses. In addition, we limited ourselves to GWAS signals for which the maximum LBF was at least 2.

### The proportion of GWAS loci explained by eQTLs

To provide an estimate of the proportion of GWAS loci that are explained by the eQTLs identified in this study, we counted all fine-mapped GWAS signals for which there was at least one variant with an LBF > 2 and a P-value < 1×10⁻^5^. Every signal that colocalized to a *cis*-eQTL or *trans*-eQTL was correspondingly annotated. Category definitions are listed in **Table S8**.

### Mendelian randomization analyses

For genomic locus–based Mendelian randomization analyses, we used the pleiotropy-robust method MRLink2^106^, reimplemented in the R programming language. To specify exposure and outcome variables, we intersected tested *cis*-eQTL loci with colocalizing *trans*-eQTL or GWAS loci, using the same locus definitions and QC filters used in the corresponding fine-mapping analyses. Input eQTL and GWAS β and their SEs were normalized to ‘variance explained’ units by the method implemented in the original MRLink2 software^106^. As additional inputs required by the MRLink2 method, we provided the LD matrix empirically derived from eQTLGen, specified the sample size as the median per-variant sample size across the corresponding intersecting eQTL or GWAS loci, and specified the maximal allowed correlation between input variants as R = 0.99.

### Prioritization of core genes for GWAS phenotypes

We developed a gene prioritization framework that integrates *cis-* and *trans*-eQTL data with GWAS summary statistics to identify genes consistently up– or downregulated by disease-associated variants. Our approach constructs a directed gene regulatory network and traces regulatory paths from GWAS variants to genes, followed by a permutation-based statistical test to identify genes with consistent regulatory effects.

We first constructed a directed gene regulatory network using *cis*-eQTL-to-*trans*-eQTL colocalizations as edges linking genes to each other. Next, we integrated all filtered GWAS−eQTL colocalizations as additional edges, linking genes to GWAS associations. All unique colocalizing CSs from GWAS were coded as a GWAS association node. We merged GWAS associations if the lead variants of their colocalizing eQTL signals were correlated with an R^2^ > 0.5.

For all combinations of GWAS association nodes and gene nodes, we computed the shortest path from the GWAS association to the gene using the directed gene regulatory network. To ensure biological plausibility and interpretability, we retained only paths that matched one of the following categories:

● Direct *cis*-eQTL paths: variant → gene (*cis*-eQTL),
● Direct *trans*-eQTL paths: variant → gene (*trans*-eQTL),
● Indirect *trans*-eQTL paths: variant → *cis*-gene → *trans*-gene.

To further refine the set of paths, we applied two key criteria:

● eQTL significance: the variant must exhibit a nominally significant eQTL effect on the target gene (P < 0.05, |Z| > 1.96).
● Directional concordance: the sign of the direct eQTL effect from the variant to the focal gene must match the inferred regulatory direction derived from the sequence of edges in the path.

For each gene, we computed a signed path score by summing the directional effects (+1 for upregulation, –1 for downregulation) across all valid paths. Scores were stratified by path type and path length and then aggregated into a single gene score.

To assess the significance of the observed gene scores, we generated a null distribution using 1,000 permutations:

● For each permutation, we sampled the same number of variant–gene paths per path category from a pool of independent GWAS traits.
● Associations were sampled with replacement.
● For each sampled set, we recalculated gene scores.

We computed empirical Z-scores and two-sided P-values for each gene by comparing the observed gene score to the mean and SD of the permuted scores. Genes with extreme Z-scores were prioritized as likely targets of consistent regulatory influence by disease-associated variants.

## Functionally active and neutral cis-eQTL

We defined the list of ‘functionally active’ *cis*-eQTLs as those *cis*-eQTLs that show colocalization with *trans*-eQTLs (see ‘Inference of directed gene regulatory relationships’). We compared this list of *cis*-eQTLs with a set of *cis*-eQTLs that do not show colocalization with *trans*-eQTLs by fitting a logistic regression model that included significance, MAF, and median gene expression in order not to bias the comparisons. We note that the effect size of a *cis*-eQTL is a function of the *cis*-eQTL significance and MAF and is thus also controlled for in this matching procedure. *Cis*-eQTLs variants that appeared multiple times were made unique by arbitrarily selecting one eQTL for the specific variant. This ultimately results in 54,439 putatively neutral *cis*-eQTLs and 2,869 functionally active *cis*-eQTLs.

We subsequently collected a set of eQTL features that were informative about either the *cis*-eQTL (e.g., SNP−gene distance), the *cis*-eQTL SNP (e.g., extent of LD), or the gene (e.g., pLI score) (**Table S11**). We derived these from Mostafavi et al., 2023^22^, ENSEMBL Variant Effect Predictor^21^, gnomAD^15^, chromatin accessibility in PBMCs (derived using newly generated single-cell multi-ome data, manuscript in preparation)^45^, and replication results from GTEx^5^, sc-eQTLGen^23^, MetaBrain^13^, scMetaBrain^24^ and INTERVAL sQTL data^12^. This permitted us to test whether any of these features described a significant difference between ‘functionally active’ and ‘neutral’ *cis*-eQTLs.

To test the robustness of these findings, we redid the enrichment analyses while confining ourselves to a subset of genes on which multiple SNPs were having an independent *cis*-eQTL effect, but with discrepant *trans*-eQTL effects. In other words, when there are multiple *cis*-eQTL CSs for a gene where one does colocalize with a *trans*-eQTL and the other does not colocalize with that *trans*-gene, there is a discrepancy.

## Code availability

The Nextflow^107^ pipeline for automatic QC, filtering, and reporting is provided here: https://github.com/eQTLGen/DataQC. The Nextflow pipeline for imputation is provided at: https://github.com/eQTLGen/eQTLGenImpute. The Nextflow pipeline for per-cohort data preparations is available at: https://github.com/eQTLGen/PerCohortDataPreparations. The pipeline that performs per-cohort calculations of effect sizes and SE and the inverse-variance meta-analysis is available at: https://github.com/eQTLGen/MetaAnalysis. The pipeline for fine-mapping is available at: https://github.com/eQTLGen/ExtractMetaAnalysisResults. The pipeline for processing and harmonization of external summary statistics is available at: https://github.com/eQTLGen/GwasProcessing. The Nextflow pipeline for *cis−trans* colocalization analysis is available at: https://github.com/eQTLGen/eQTLGenCisTransSuSieColoc. The full pipeline used for processing GWAS summary statistics is available at: https://github.com/eQTLGen/GwasProcessing. The Nextflow pipeline for colocalization analysis between eQTL and GWAS summary statistics is available at: https://github.com/eQTLGen/eQTLGenEqtlGwasSuSieColoc. The code that is used to summarize follow-up analyses, prepare figures and calculate results for this manuscript is available at: https://github.com/eQTLGen/WarmerdamEtAl2026.

## Data availability

Full summary statistics from all discovery analyses will be made publicly available upon acceptance for publication. Our study is comprised of existing human blood eQTL datasets. Data availability of these datasets is included in the **Supplementary Information**. Data generated by or processed for downstream analyses in this study can be found in **Supplementary Tables**. Summary statistics for eQTLs in replication datasets will also be made publicly available upon acceptance, with the exception of unpublished datasets, which will be released upon their respective publications.

## Acknowledgements

P.D. is supported by an NWO ZonMW-VENI Grant (no. 9150161910057).

M. Jesse is supported by the Estonian Research Council (MOB3ERC115).

V.R. was funded by the German Federal Ministry of Education and Research (BMBF) within the framework of the e:Med research and funding concept (grant # 01ZX1906B & 01ZX2206B).

A. Teumer has been funded by the Deutsche Forschungsgemeinschaft (DFG, German Research Foundation) – 542489987.

R.D.R is supported by the Innovative Medicines Initiative grant 3TR (GA#831434).

M.R is supported by the Innovative Medicines Initiative grant 3TR (GA#831434)

D. Dutta is supported by the Intramural Research Program of the National Cancer Institute, National Institutes of Health, US Department of Health and Human Services.

The Wellcome Sanger Institute is supported by core funding from the Wellcome Trust [220540/Z/20/A].

This research was supported in part by the Intramural Research Program of the National Institutes of Health (NIH). The contributions of the NIH author(s) were made as part of their official duties as NIH federal employees, are in compliance with agency policy requirements, and are considered Works of the United States Government. However, the findings and conclusions presented in this paper are those of the author(s) and do not necessarily reflect the views of the NIH or the U.S. Department of Health and Human Services.

P.C.C. is supported by the Innovative Medicines Initiative Joint Undertaking (grant 115565)

J. Martin is supported by the Spanish Ministry of Science and Education. PID2022-13929208-I00

E.R. is supported by the Academy of Finland (Grant number: 338395)

Y.M. and H. Naeem are supported in part by the Qatar National Research Fund (QNRF Awards PPM1-1122-150008) and internal funds from Sidra Medicine.

L.M.R. was supported by R01 AG075884

This work was supported by the Estonian Research Council grant PUT (PSG1028).

This project has received funding from the European Research Council (ERC) under the European Union’s Horizon 2020 research and innovation programme (grant agreement n° 772376 – EScORIAL.

A.C. received funding from NWO (Veni 09150162310207) and Diabetes Fonds (Junior Fellowship 2023.81.006).

M.G.P. vdW received funding from NWO (VIDI 223.041).

P.M.V. receives funding from the European Research Council (ERC) under the European Union’s Horizon Europe research and innovation programme (grant agreement No 101198904)

J.C.K. received funding from the Medical Research Council (MR/V002503/1), Wellcome Trust Investigator Award (204969/Z/16/Z), Wellcome Trust Grants (090532/Z/09/Z and 203141/Z/16/Z) to core facilities Wellcome Centre for Human Genetics, Chinese Academy of Medical Sciences (CAMS) Innovation Fund for Medical Science (CIFMS), China (grant number: 2018-I2M-2-002), and NIHR Oxford Biomedical Research Centre.

T.M.F. is funded by Swiss National Science Foundation grant 10003484 and Novo Nordisk and the Fondation pour la recherche sur le diabete.

A.B. was supported by NIH R35GM139580.

Y.O. was supported by JSPS KAKENHI (25H01057), and AMED (JP24km0405217, JP24ek0109594, JP24ek0410113, JP24kk0305022, JP223fa627001, JP223fa627002, JP223fa627010, JP223fa627011, JP22zf0127008, JP24tm0524002, JP24wm0625504, JP24gm1810011), JST Moonshot R&D (JPMJMS2021, JPMJMS2024), Takeda Science Foundation, Ono Pharmaceutical Foundation for Oncology, Immunology, and Neurology, Bioinformatics Initiative of Osaka University Graduate School of Medicine, Institute for Open and Transdisciplinary Research Initiatives, Center for Infectious Disease Education and Research (CiDER), and Center for Advanced Modality and DDS (CAMaD), Osaka University, RIKEN TRIP initiative (AGIS).

M.E.A. is supported by the Innovative Medicines Initiative Joint Undertaking (grant 115565).

This work was in part supported by core funding from the British Heart Foundation (RG/F/23/110103), NIHR Cambridge Biomedical Research Centre (NIHR203312) [*], BHF Chair Award (CH/12/2/29428), Cambridge BHF Centre of Research Excellence (RE/24/130011), and by Health Data Research UK, which is funded by the UK Medical Research Council, Engineering and Physical Sciences Research Council, Economic and Social Research Council, Department of Health and Social Care (England), Chief Scientist Office of the Scottish Government Health and Social Care Directorates, Health and Social Care Research and Development Division (Welsh Government), Public Health Agency (Northern Ireland), British Heart Foundation and the Wellcome trust. *The views expressed are those of the authors and not necessarily those of the NIHR or the Department of Health and Social Care.

D.I.B. is supported by the KNAW Academy Professor Award (PAH/6635).

R.O. is supported by R01 MH115676. B.Pasaniuc is supported by R01 MH115676.

A.L.P. is supported by U01 HG012009.

K.A. is supported by the Estonian Research Council (MOB3ERC115).

Z.K. was supported by the Swiss National Science Foundation (315230_219587).

This work was supported by the Ministry of Education and Research Centres of Excellence grant TK214 name of CoE. This work was supported by the Estonian Research Council grant PUT (PRG1911). The Project leading to this article has received funding from the European Union’s Horizon 2020 research and innovation-under grant agreement No 101017802.

This work was supported by the Estonian Research Council grant PUT (PRG1291).

U. Võsa was supported by the European Regional Development Fund, the programme Mobilitas Pluss (MOBTP108). The research was conducted using the Estonian Center of Genomics/Roadmap II funded by the Estonian Research Council (project number TT17).

L.H.F. was supported by Dutch Research Council grants (ZonMW-VICI 09150182010019 to L.H.F. and ZonMW LongCOVID grant 10430302110002), European Union Horizon Europe Research and Innovation Program grant 101057553 (LongCovid), a Senior Investigator Grant from the Oncode Institute, a grant from Oncode Accelerator, and a grant from Saxum Volutum (Pericode). L.H.F. is also supported by a public-private partnership with Biogen and Roche.

Meta-analyses and interpretative bioinformatic analyses were performed at the University of Tartu High Performance Computing Center^16^ and at the University of Groningen High Performance Computing Center. We thank Katherine McIntyre for the English editing of our manuscript. The authors wish to acknowledge the contributions of all participating cohorts and study participants.

## Author contributions

Conceptualization: H.W., L.H.F., U.Võsa; Data Curation: C.A.R.W., H.W., P.D., H.Naeem, Y.M., L.H.F., U.Võsa; Formal Analysis: C.A.R.W., H.W., A.vdG., M.J.B., P.D., T.vL., A.J.L., M.Jesse, T.B., S.L., S.N., M.X., D.T., H.K., V.R., A.F., A.Teumer, M.F., E.P., A.Tokolyi, R.P., J.J.H., R.D.R., M.R., B.H., B.Pierce, L.T., Q.S.W., T.H., S.B.C., D.D., S.W., T.D., L.L., P.P.M., A.R.W., K.L.B., J.W., E.C., C.Boahen, H.Naeem, K.Freimann, Y.M., L.H.F., U.Võsa; Funding Acquisition: Z.K., T.E., L.H.F., U.Võsa; Investigation: C.A.R.W., H.W., P.D., H.Naeem, Y.M., L.H.F., U.Võsa; Methodology: C.A.R.W., H.W., A.vdG., M.J.B., P.D., T.vL., A.J.L., M.Jesse, M.H., Z.K., L.H.F., U.Võsa; Project Administration: C.A.R.W., L.H.F., U.Võsa; Resources: T.B., S.L., S.N., M.X., D.T., H.K., V.R., A.F., A.Teumer, M.F., E.P., A.Tokolyi, R.P., J.J.H., R.D.R., M.R., B.H., B.Pierce, L.T., Q.S.W., T.H., S.B.C., D.D., S.W., T.D., L.L., P.P.M., A.R.W., K.L.B., J.W., E.C., C.Boahen, R.J., F.B., K.K., M.Loeffler, J.T., I.D.C., K.D., J.R.G., J.Verlouw, V.Kukushkina, R.M., J.Budde, M.Johnson, J.S., E.S., P.C.C., J.Martin, E.R., M.Kibriya, F.J., H.Naeem, M.N., J.C.S., A.M., M.A.I., J.Mykkänen, K.P., S.P.R., L.M.R., M.vG., C.J.H.vdK., C.G.S., J.Veldink, K.Freimann, M.V., A.K., L.O., A.C., Y.H., J.Bryois, E.A.T., M.G.P..vdW., E.B.R.T., s.C., V.Kumar, P.M.V., A.F.M., G.W.M., Y.L., J.C.K., A.S., L.F., T.M.F., J.M.S., U.Völker, A.Battle, Y.M., Y.O., K.Fukunaga, H.Namkoong, H.A., T.L., M.Kähönen, O.T.R., M.E.A., G.B., B.W.J.H.P., D.S.P., M.I., P.A., Y.J.S., C.C., J.vM., H.P., A.P., C.G., C.Blauwendraat, E.E.D., M.S., M.Loh, J.C., G.G., D.I.B., A.V., A.Brown, B.Pasaniuc, R.O., K.A., T.H.S.T.; Software: C.A.R.W., H.W., A.vdG., M.J.B., P.D., T.vL., A.J.L., M.Jesse, L.H.F., U.Võsa; Supervision: H.W., M.J.B., P.D., B.J.S., V.Kumar, P.M.V., A.F.M., G.W.M., Y.L., J.C.K., A.S., L.F., T.M.F., J.M.S., U.Völker, A.Battle, Y.M., Y.O., K.Fukunaga, H.Namkoong, H.A., T.L., M.Kähönen, O.T.R., M.E.A., G.B., B.W.J.H.P., D.S.P., M.I., P.A., Y.J.S., C.C., J.vM., H.P., A.P., C.G., C.Blauwendraat, E.E.D., M.S., M.Loh, J.C., G.G., D.I.B., A.V., A.Brown, B.Pasaniuc, R.O., A.L.P., K.A., L.H.F., U.Võsa; Validation: C.A.R.W., H.W., M.J.B., P.D., M.V., A.K., L.O., A.C., Y.H., J.Bryois, E.A.T., s.C., K.A., L.H.F., U.Võsa; Visualization: C.A.R.W., L.H.F., U.Võsa; Writing – Original Draft Preparation: C.A.R.W., H.W., A.vdG., M.J.B., P.D., T.vL., A.J.L., L.H.F., U.Võsa; Writing – Review & Editing: C.A.R.W., H.W., M.J.B., P.D., T.vL., B.J.S., S.W., L.L., P.P.M., L.M.R., M.vG., L.O., A.C., T.M.F., D.I.B., A.L.P., K.A., Z.K., L.H.F., U.Võsa.

## Competing interests

Q.S.W. is an employee of Calico Life Sciences LLC. L.M.R. is a consultant for the NHLBI TOPMed Administrative Coordinating Center (through Westat). J.Veldink reports to have sponsored research agreements with Biogen, Eli Lilly, Trace and Astra Zeneca. M.V. is currently a full-time employee of Illumina and owns stock in the company; these interests began after M.V.’s contribution to this work was completed. Y.H. is a full-time employee and holds stock options at Biogen Inc. J.B. is an employee and stockholder of Roche. A.B. is a shareholder in Alphabet Inc, and a founder and equity holder of CellCipher, Inc. M.E.A.R. received honoraries for courses and conferences from GSK, funding project from AstraZeneca that do not influence the work presented here. D.S.P. is an employee and stockholder of AstraZeneca. M.I. is a trustee of the Public Health Genomics (PHG) Foundation, a member of the Scientific Advisory Board of Open Targets, and has research collaborations with AstraZeneca, Nightingale Health and Pfizer which are unrelated to this study. L.H.F. has ongoing collaborations with Biogen and Roche on eQTL mapping in brain. These collaborations did not influence the design, analysis, or interpretation of the current study. All other authors declared no competing interests.

## Notes

### Author Declarations

The eQTLGen phase 2 study was reviewed by the Medical Ethics Review Board (METc) of the University Medical Center Groningen under research registry number 19149, which concluded that it is not a clinical research study with human subjects as meant in the Dutch Medical Research Involving Human Subjects Act (WMO)

