## Supplementary material for "*Trans*-eQTLs reveal the architecture of human gene regulatory networks"

### Table of contents


Table of contents 2
Supplementary Figures 4
Supplementary notes 36
Supplementary note 1: Calculation of false discovery rate (FDR) for *cis*- and *trans*-eQTLs: 36
Cohort Information 37
AMP-PD 37
LIFE-Heart 39
LIFE-Adult 41
Rotterdam Study - RNA-seq 43
PAN 45
NTR 47
Leiden Longevity Study 49
LifeLines DEEP 51
CODAM 53
Fehrmann 54
300TZFG 55
GTEx 57
YFS 59
STRIP 61
SHIP-TREND 63
Rotterdam Study (HT12v4) 65
PRECISEADS 67
OphoffBP 69
NTR NESDA 71
Morocco 72
Jackson Heart Study 73
Japan COVID-19 Task Force 75
INTERVAL 77
HELIOS 80
GAIT-2 83
GAinS 85
EstBB (RNA-seq) 88
EstBB (Illumina HT12v3) 90
IMI DIRECT 92
Knight-ADRC cohort 94
CHDWB 97
CanPath 98
CAD 100
BEST 101
KORA - batch 1 103
KORA - Batch 2 105
InCHIANTI 107
DGN 108
BSGS 110
Consortium Banner Authors 111
Estonian Biobank research team 111
PRECISEADS Clinical Consortium 111
IMI DIRECT Consortium 112
sc-eQTLGen Consortium 113
The HELIOS Study Team 115

### Supplementary Figures

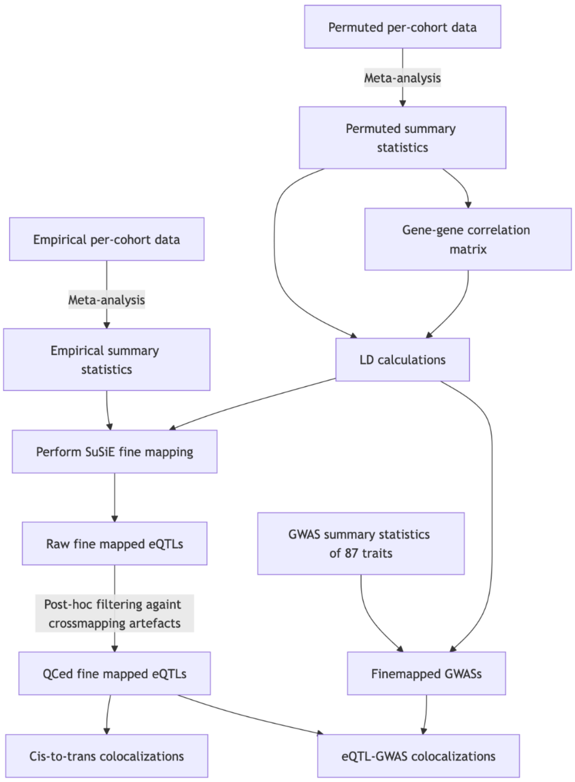


**Figure S1. Overview of the central analysis.** We performed two meta-analyses: one empirical and one permuted. The permuted analysis was used to calculate linkage disequilibrium and gene−gene correlations. These were used to fine-map eQTLs and GWASs. Ultimately, this allowed us to perform a comprehensive colocalization analysis. LD, linkage disequilibrium. GWAS, genome-wide association study. QC, quality control.

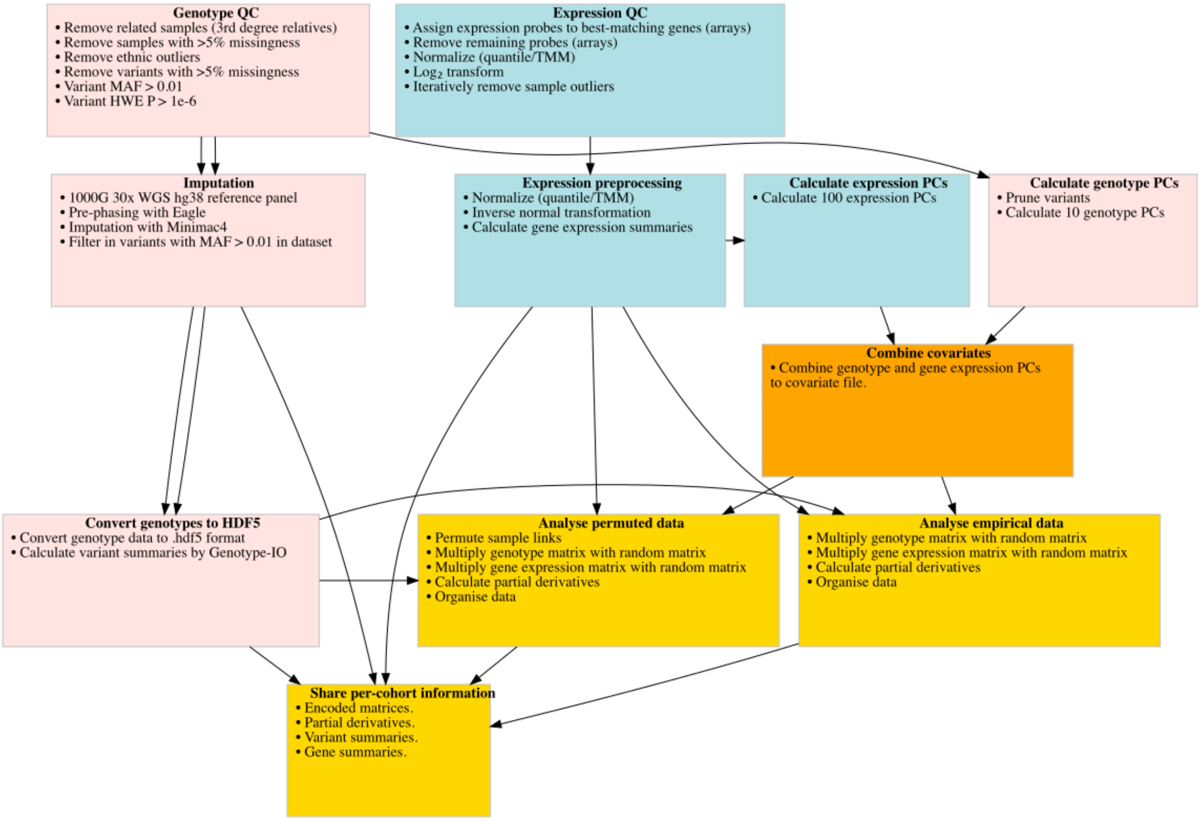


**Figure S2. Overview of the per-cohort analysis pipelines.** Boxes depict the different steps of the per-cohort data processing. QC, quality control. MAF, minor allele frequency. HWE, Hardy-Weinberg equilibrium. TMM, trimmed mean of M-values. PC, principal component. SNP, single nucleotide polymorphism.

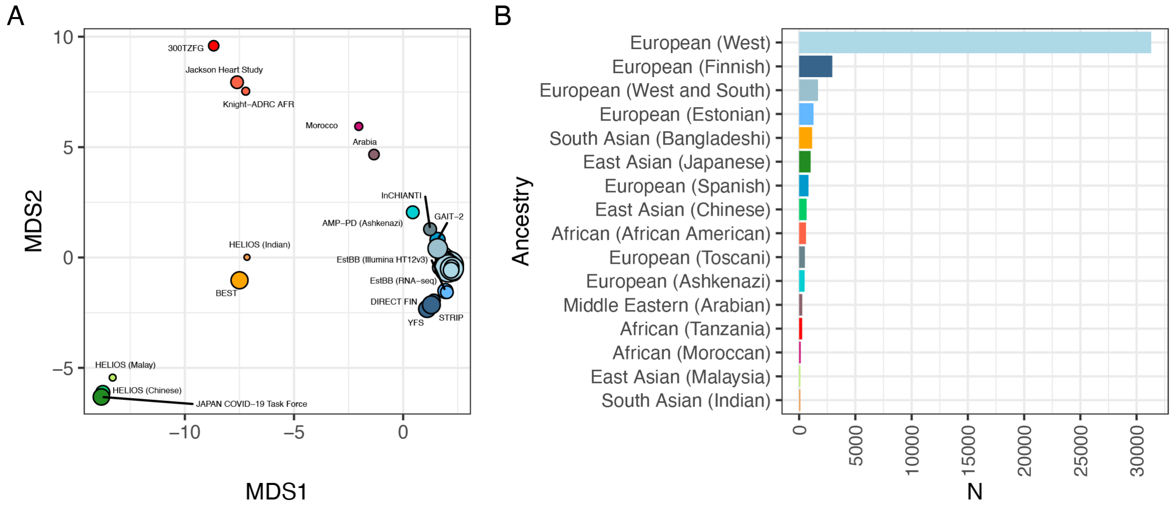


**Figure S3. Genetic distance between the cohorts contributing to eQTLGen phase 2. a**, First two axes of multidimensional scaling, calculated on the pairwise Euclidean distances between allele frequencies of each eQTLGen cohort (9,122 linkage disequilibrium−pruned variants, minor allele frequency > 0.05 in every cohort). Dot size signifies sample size. **b**, Sample sizes of the cohorts from each ancestry.

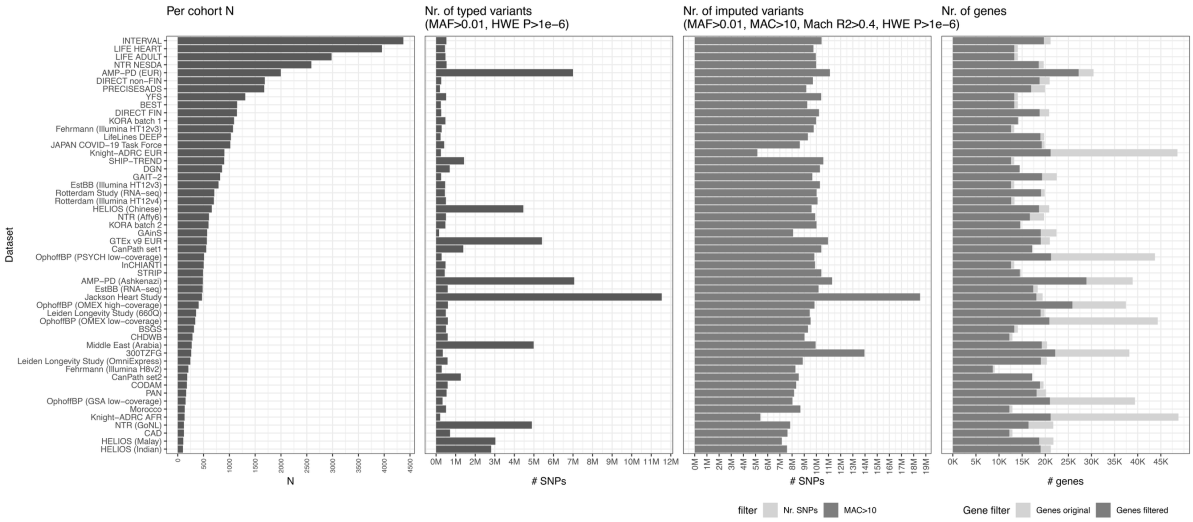


**Figure S4. Overview of dataset features and per-cohort filters.** Variants with a per-dataset minor allele frequency (MAF) > 0.01, minor allele count (MAC) > 10, Mach $R^2 > 0.4$, and Hardy-Weinberg equilibrium (HWE) $P > 1 \times 10^{-6}$ were included in the meta-analyses. Similarly, genes for which the raw expression values were unique for at least 80% of the samples were included to the meta-analyses.

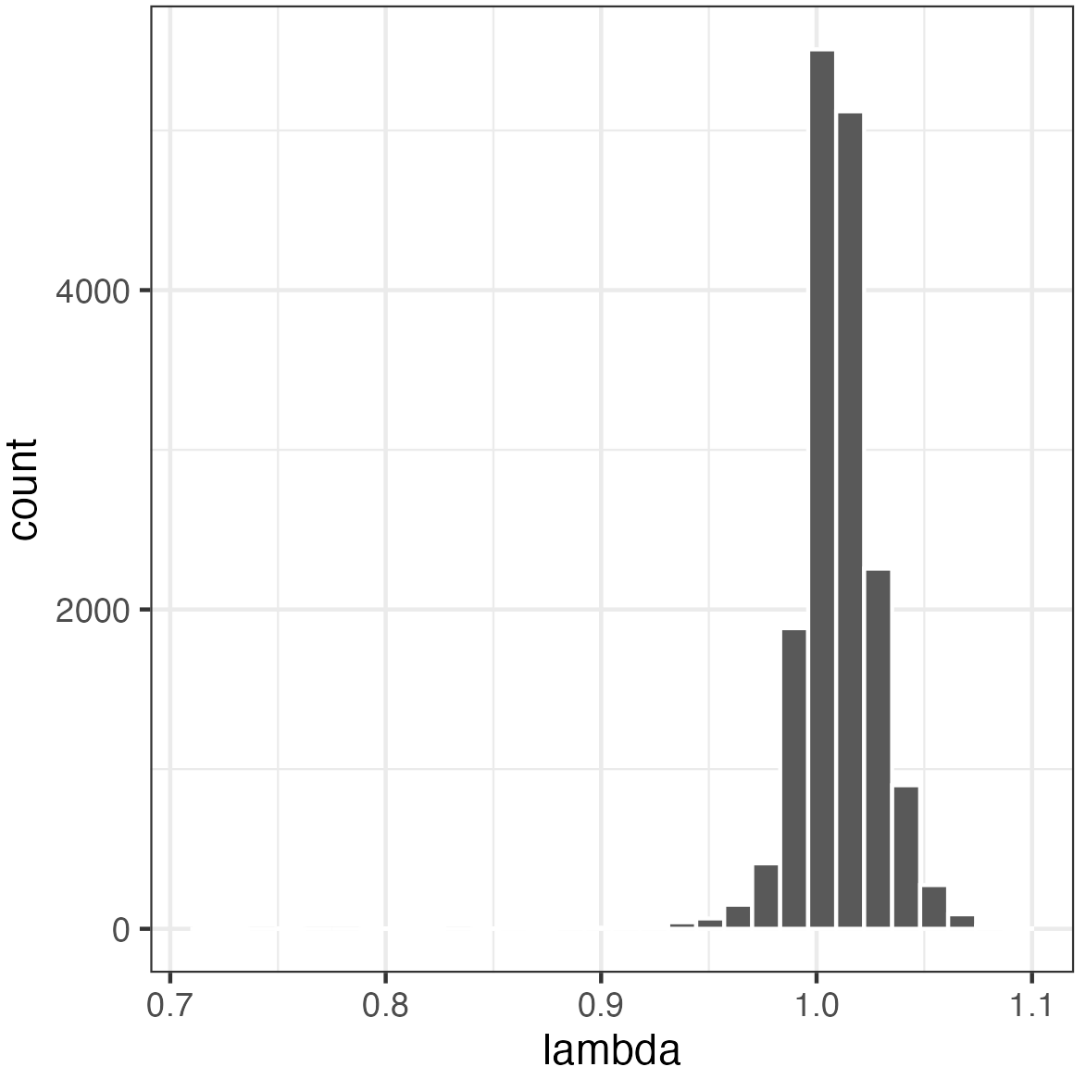


**Figure S5. Histogram with lambda inflation factors for all 16,742 genes in the meta-analysis.**

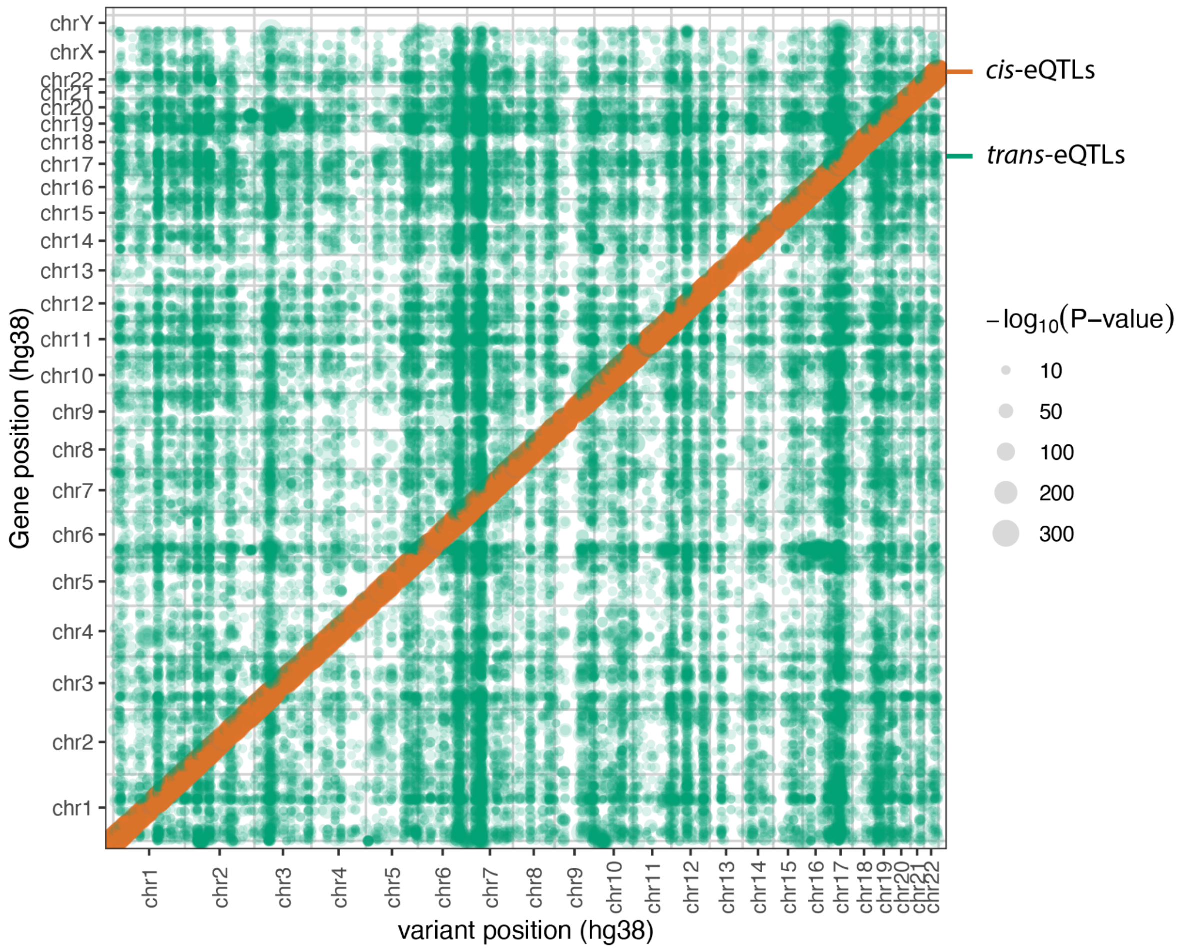


**Figure S5. Dot plot with fine-mapped associations.** Each identified eQTL is mapped according to its genomic position. X-axis indicates the genomic coordinates of the variants. Y-axis indicates the genomic coordinates of the corresponding genes.

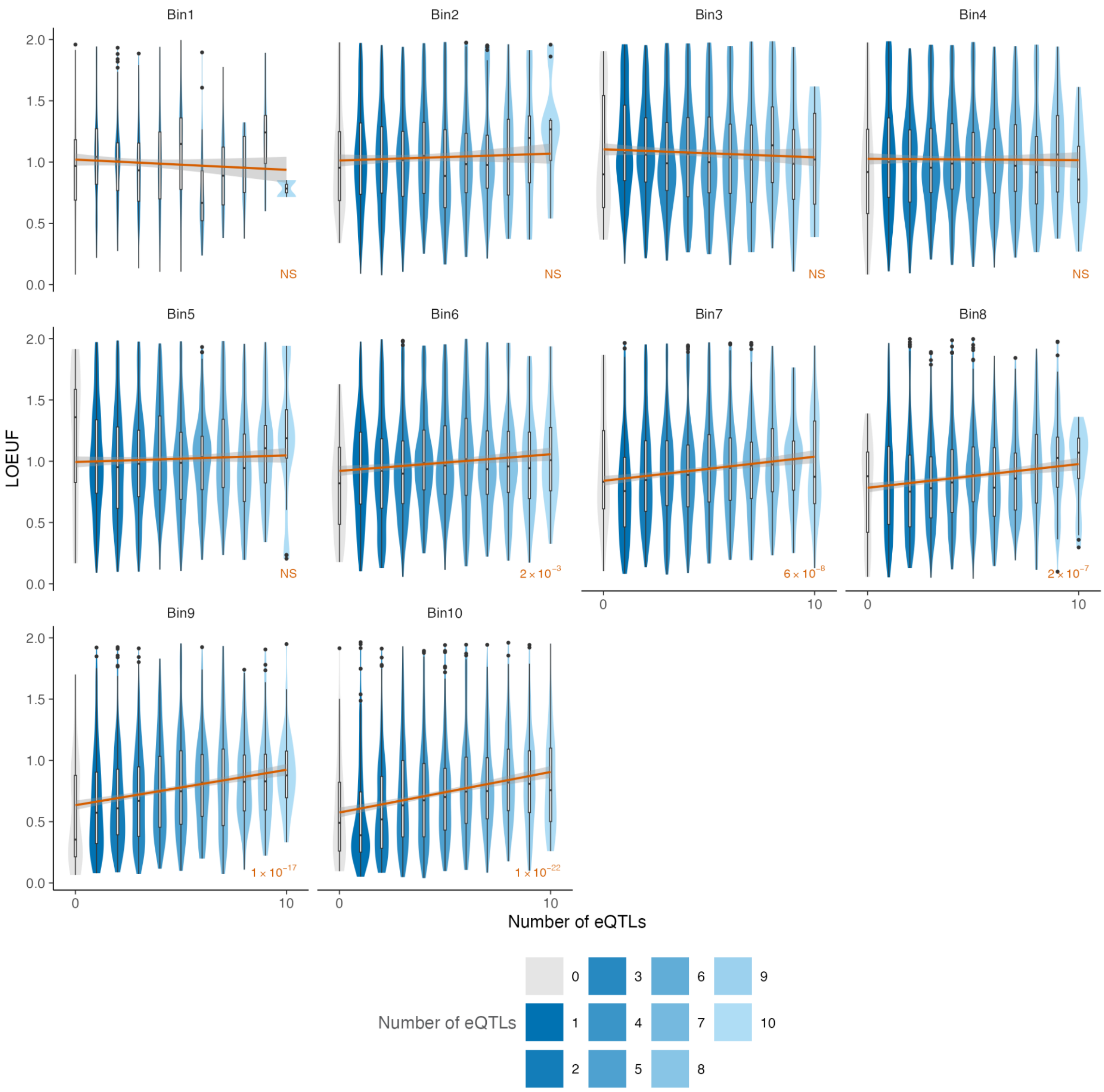


**Figure S6. Correlations between credible set counts and LOEUF across expression deciles (*cis*).** Orange line represents the slope using a simple linear regression model. Orange text indicates the P-values of Spearman correlations for each decile. Bin1 to Bin10 represent the gene expression deciles 1 to 10. The statistical models were run using all data points.

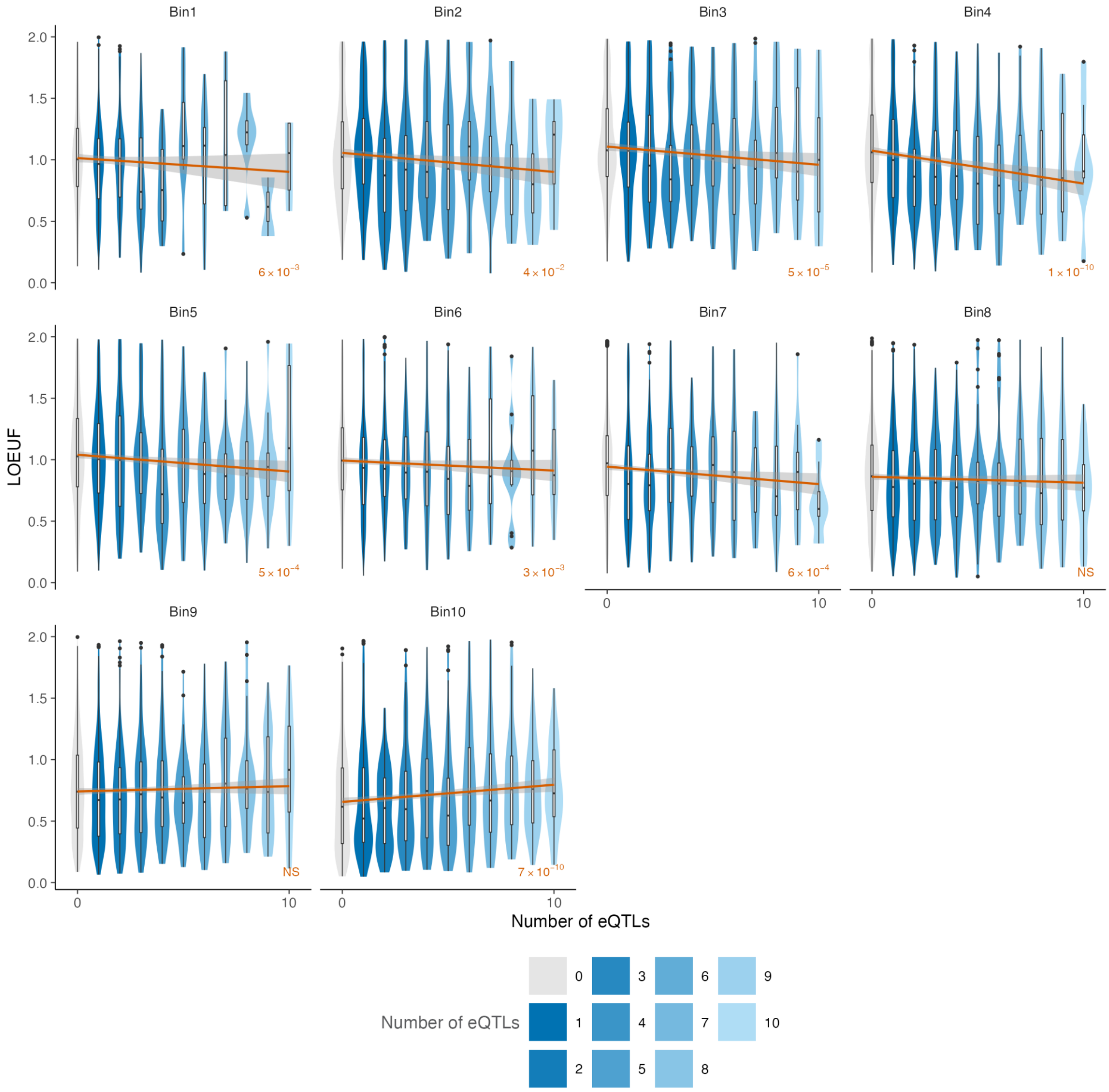


**Figure S7. Correlations between credible set counts and LOEUF across expression deciles (*trans*).** Orange line represents the slope using a simple linear regression model. Orange text indicates P-values of Spearman correlations for each decile. Bin1 to Bin10 represent the gene expression deciles 1 to 10. The statistical models were run using all data points. Only those genes with under 10 *trans*-eQTL credible sets are visualized for clarity.

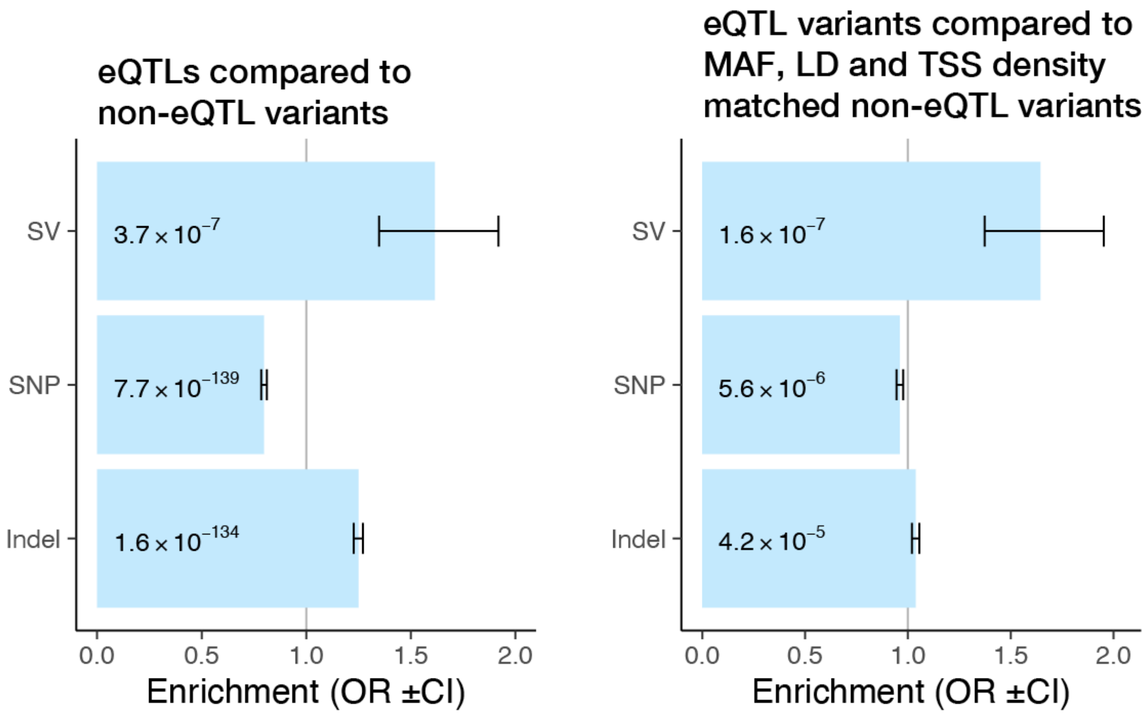


**Figure S8. Enrichments of variant classes among eQTLs.** Left panel shows enrichment of variant classes compared to non-eQTL variants. Right panel shows enrichments of variant classes when compared to non-eQTL variants with matching minor allele frequency (MAF), linkage disequilibrium (LD), and transcription start site (TSS) density. Enrichments were calculated compared to all other variant classes using Fisher's exact test. SV, structural variant. SNP, single nucleotide polymorphism. OR, odds ratio. CI, confidence interval.

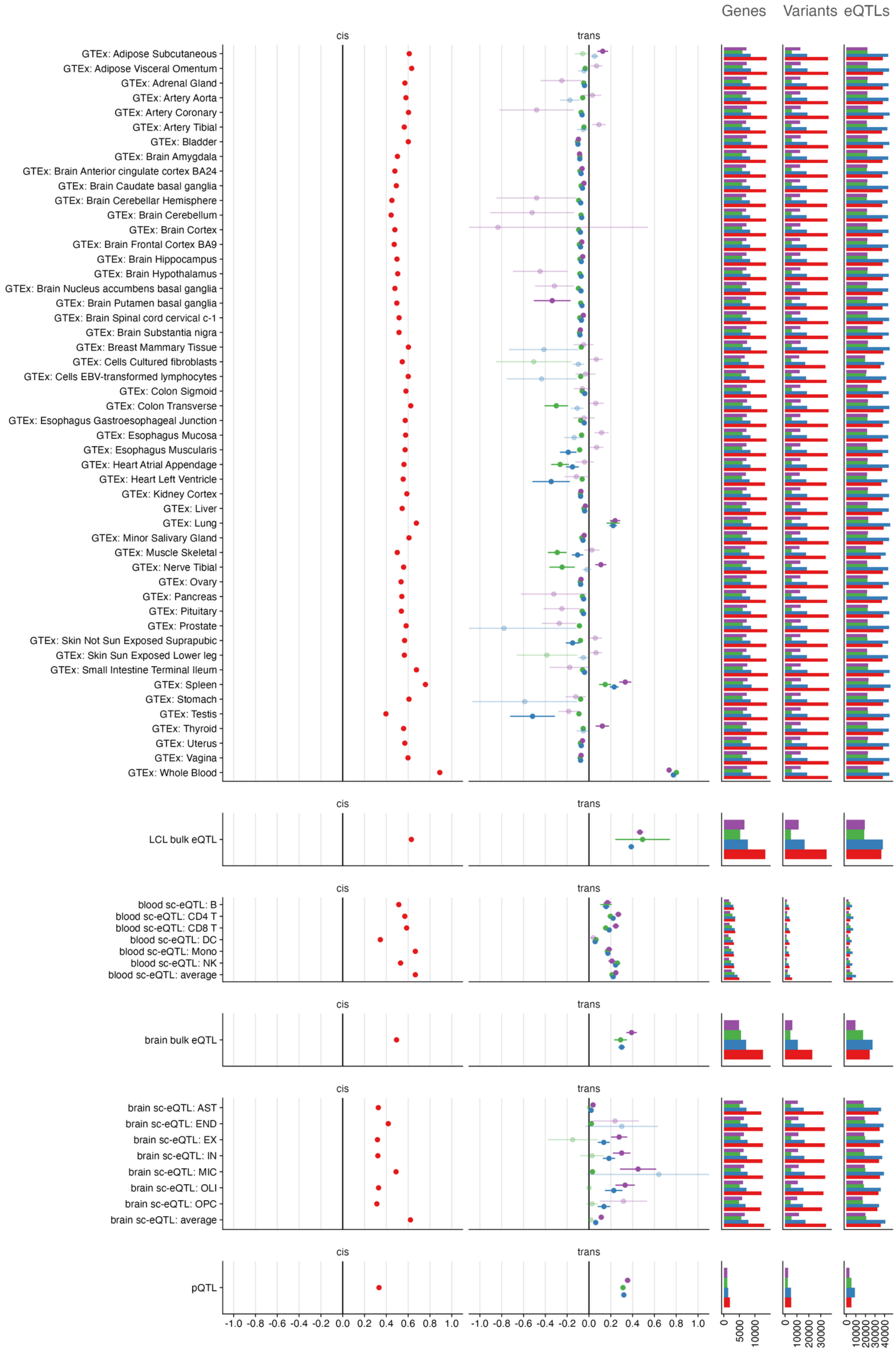


**Figure S9. Overview of the replication signals between eQTLGen and different replication and lookup analyses.** Each point represents the correlation estimate of effects between the discovery and replication dataset ($R_b$). Error bars depict standard error of $R_b$. Increased transparency of the points and error bars indicate non-significant replication signals (two-sided $R_b$ $P \geq 0.05$). For each tissue, the $R_b$ shown is calculated on only *cis*-eQTL effects (red), on all *trans*-eQTL effects (blue), on putative cell-type-composition *trans*-eQTL effects (coloc PP4 > 0.8 with any blood traits) (green), and on candidate intracellular *trans*-eQTL effects (no colocalization with any blood trait) (purple). Rightmost barplots depict the available numbers of genes, variants, and eQTLs (variant–gene combinations), used for calculating $R_b$ for corresponding comparison. LCL, lymphoblastoid cell lines. B, B cells. CD4 T, CD4 T cells. CD8 T, CD8 T cells. DC, dendritic cells. Mono, monocytes. NK, natural killer cells. AST, astrocytes. END, endothelial cells. EX, excitatory neurons. IN, inhibitory neurons. MIC, microglia. OLI, oligodendrocytes. OPC, oligodendrocyte precursor cells.

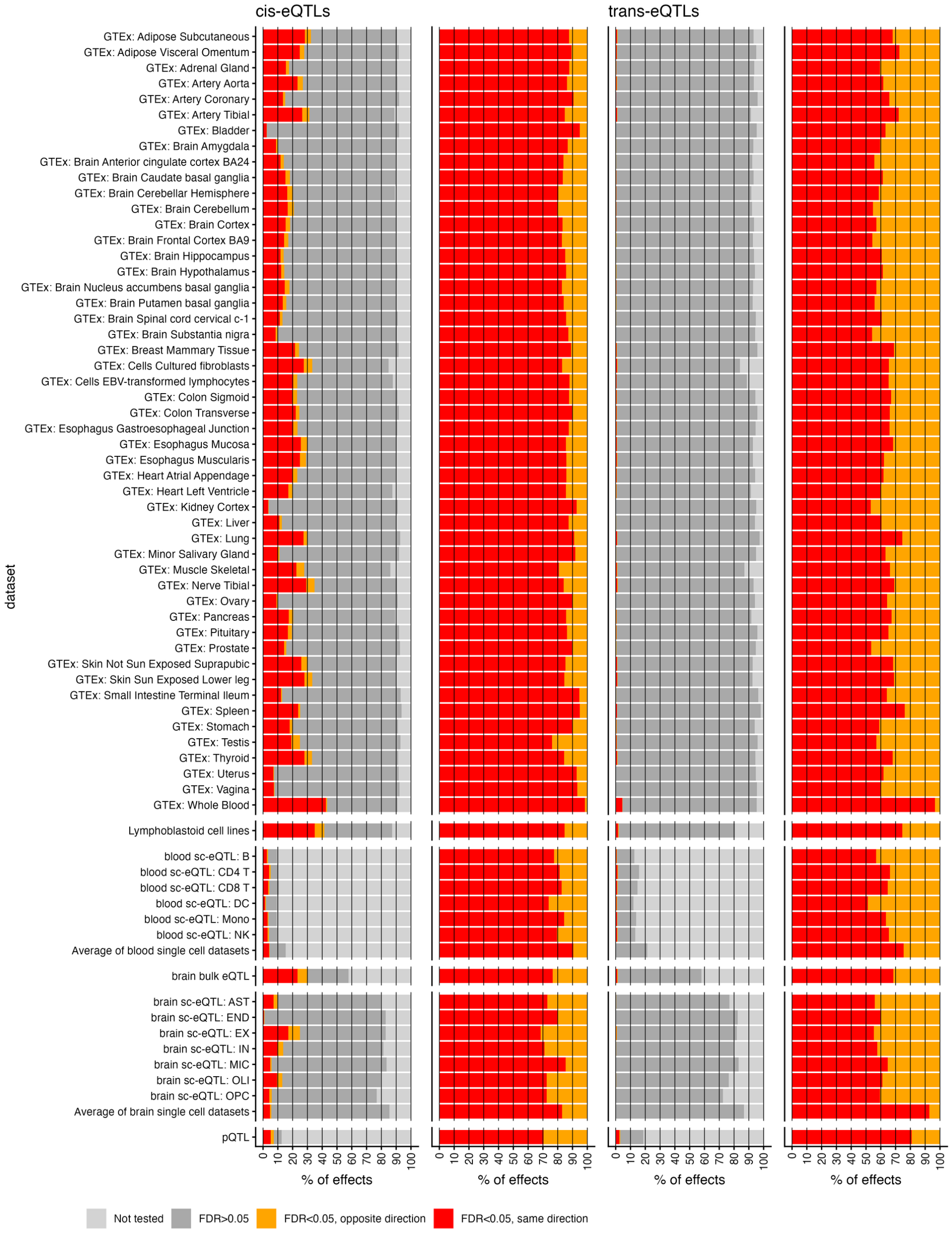


**Figure S10. Allelic concordances between eQTLGen discovery eQTL effects and different replication and lookup analyses in variety of datasets.** Shown are results for *cis*-eQTLs (two left columns) and *trans*-eQTLs (two right columns), depicting the overview of all effects and concordances for false discovery rate (FDR) < 0.05 effects, respectively. Significance of the replication effects was determined by Benjamini-Hochberg FDR. Note that formal replication power for *trans*-eQTL effects was very limited in most replication datasets, and the results in the last column are based on limited number of FDR < 0.05 variants. B, B cells. CD4 T, CD4 T cells. CD8 T, CD8 T cells. DC, dendritic cells. Mono, monocytes. NK, natural killer cells. AST, astrocytes. END, endothelial cells. EX, excitatory neurons. IN, inhibitory neurons. MIC, microglia. OLI, oligodendrocytes. OPC, oligodendrocyte precursor cells.

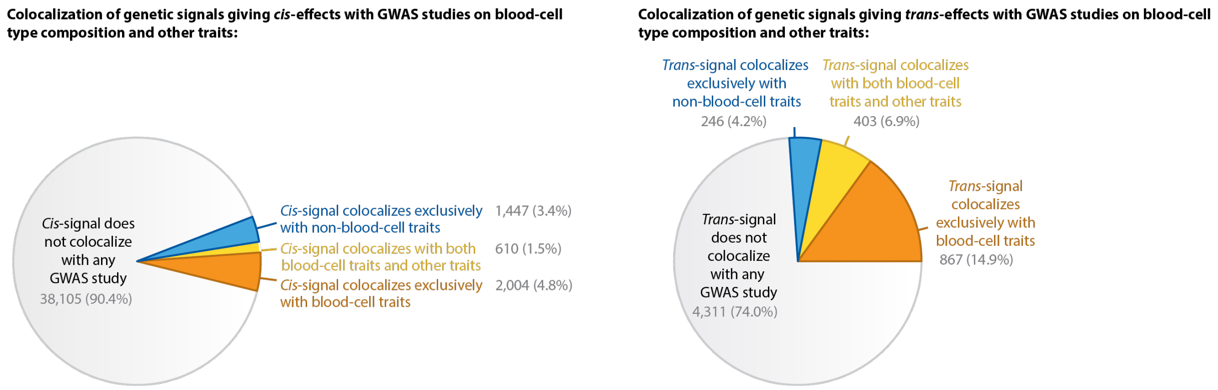


**Figure S11. Colocalizations of genetic signals giving *cis*- and *trans*-effects with GWAS studies on blood-cell-type-composition and other traits.** Alternate version of figure 3a, but on a genetic variant-level instead of an eQTL-level. Sharing of genetic signals was determined through an exhaustive colocalization approach (**Methods**).

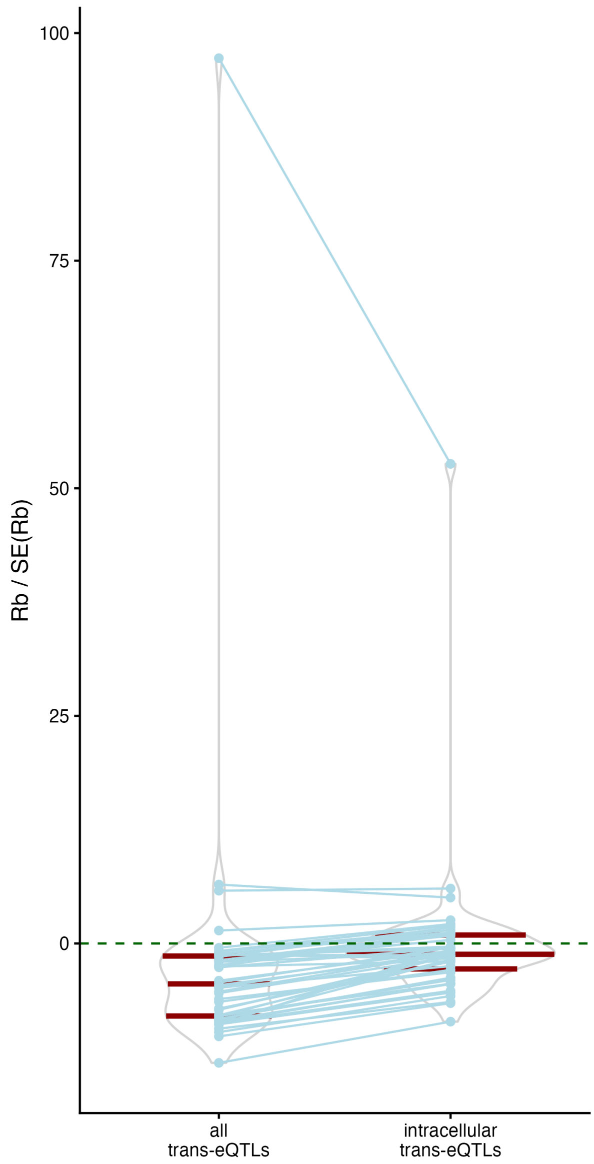


**Figure S12. Comparison of replication signal between eQTLGen and all GTEx tissues for blood cell-type composition and intracellular *trans*-eQTLs.** Each dot signifies the correlation estimate between the discovery and replication dataset ($R_b$ metric) divided by its standard error. Comparison of correlations calculated for all *trans*-eQTLs and putatively intracellular *trans*-eQTLs is shown by the violin plots. Red lines within each violin represent the boundaries of the quantiles in the distributions. The blue lines connect the same GTEx tissue in either analysis. The highest $R_b$ metrics are for GTEx whole blood, which is part of the eQTLGen discovery analysis, and all metrics were calculated by specifying complete sample overlap between discovery and replication datasets. Despite the limited power for *trans*-eQTL replication in GTEx tissues, we observe generally higher correlation between effect sizes for putatively intracellular *trans*-eQTLs (two-sided $P = 3\times10^{-8}$, paired Wilcoxon test) and more tissues with significantly positive correlation ($P < 0.05$, $R_b > 0$).

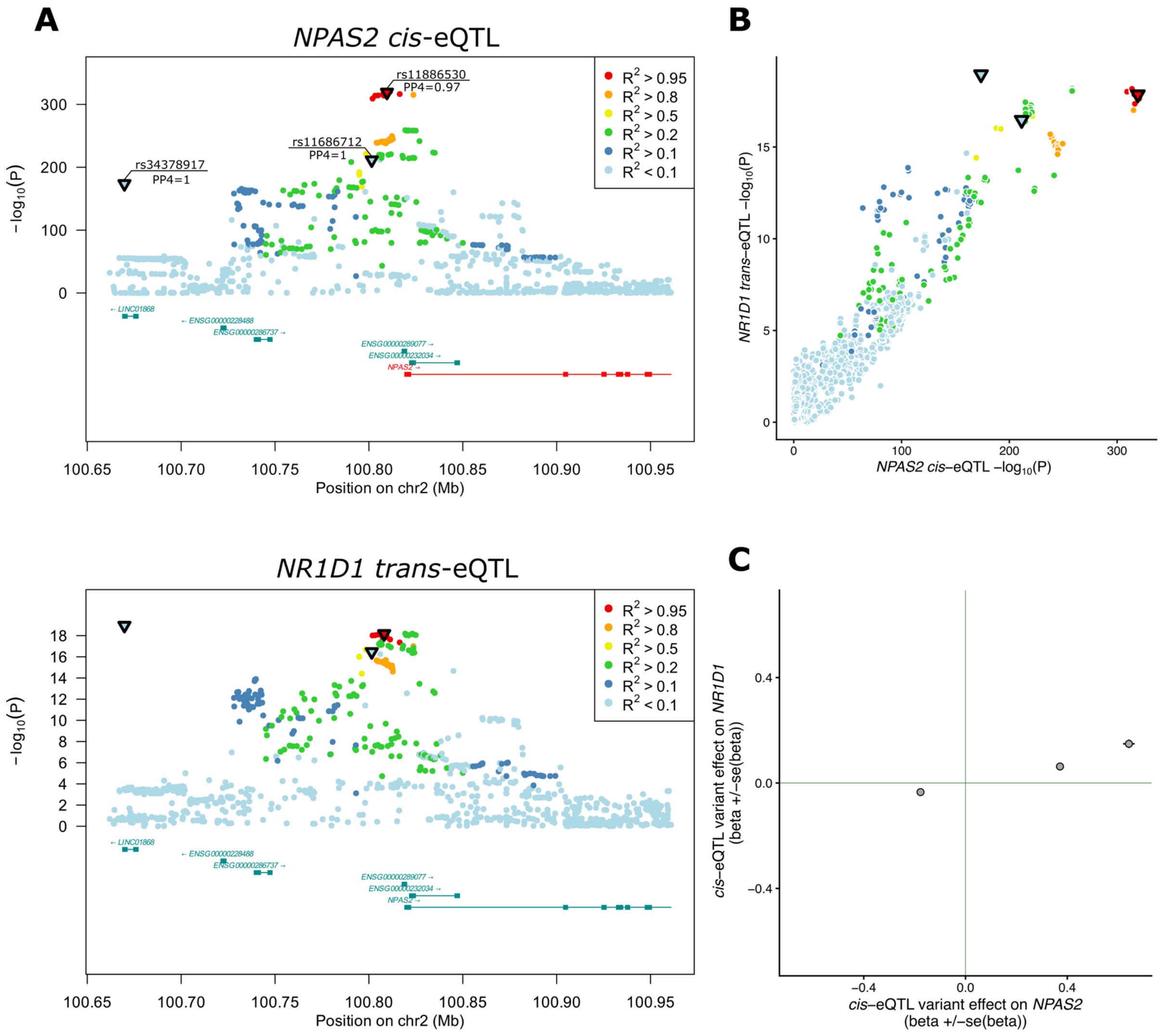


**Figure S13. Example of *cis*-eQTL locus of *NPAS2* where three independent eQTL signals colocalise with three independent *trans*-eQTL signals of *NR1D1*. a,** regional plots of *NPAS2 cis*-eQTL and *NR1D1 trans*-eQTL. *Cis*-eQTL and *trans*-eQTL lead variants for colocalising signals are outlined as inverted triangles and shown is posterior probability of colocalisation (PP4 from coloc). All variants are coloured based on the LD (eQTLGen empirical LD) with *cis*-eQTL lead variant. **b**, Scatterplot of *cis*-eQTL and *trans*-eQTL $log_{10}$(P-values). **c,** Effect of *cis*-eQTL lead variants on *cis*-eGene *NPAS2* and *trans*-eGene *NR1D1*. We observe proportional effect on both, *cis*-eGene and *trans*-eGene.

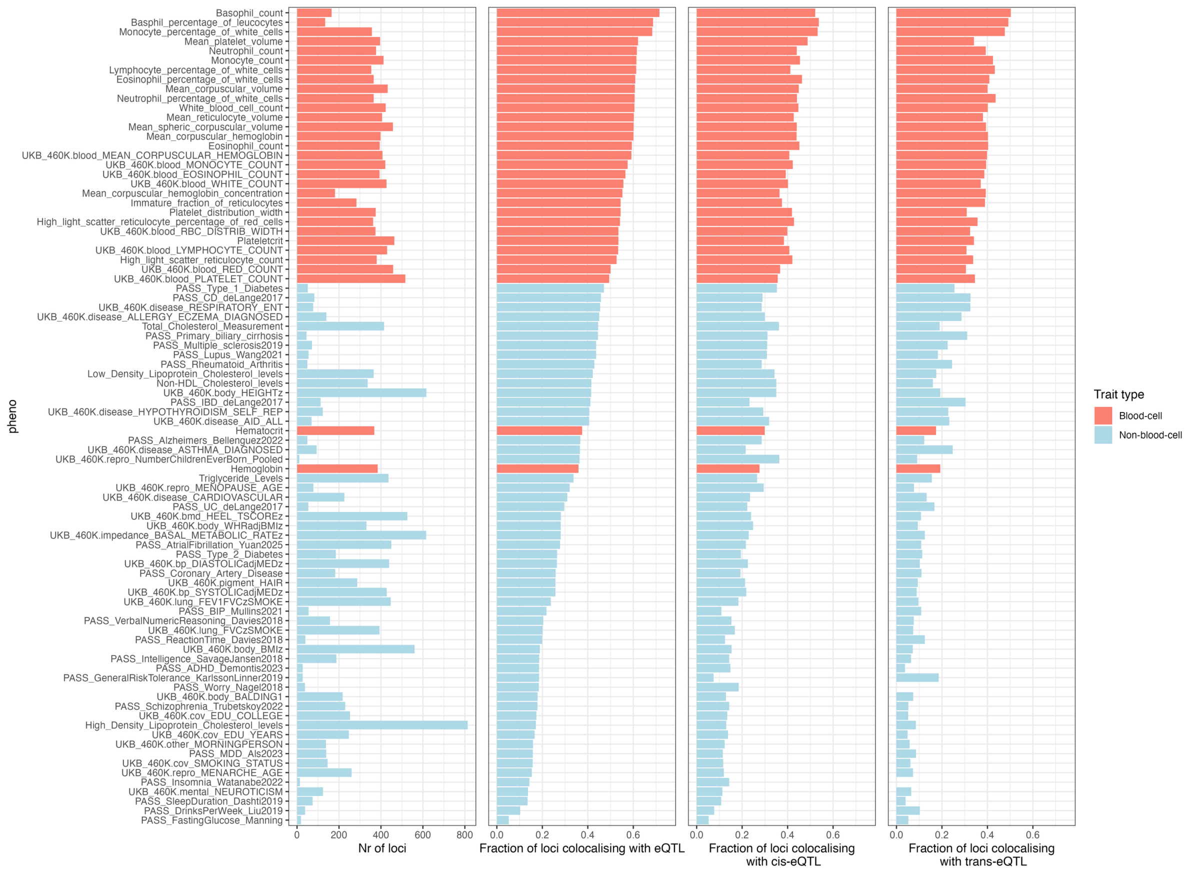


**Figure S14. Overview of per-phenotype colocalization events.** From left to right, the bar plots depict the number of fine-mapped GWAS loci, the fraction of GWAS loci colocalizing with any eQTLGen blood eQTL signal (coloc PP4 > 0.8), the fraction of GWAS loci colocalizing with any eQTLGen *cis*-eQTL signal, and the fraction of loci colocalizing with any eQTLGen *trans*-eQTL signal. Colors signify whether the GWAS trait is associated with blood-cell abundance.

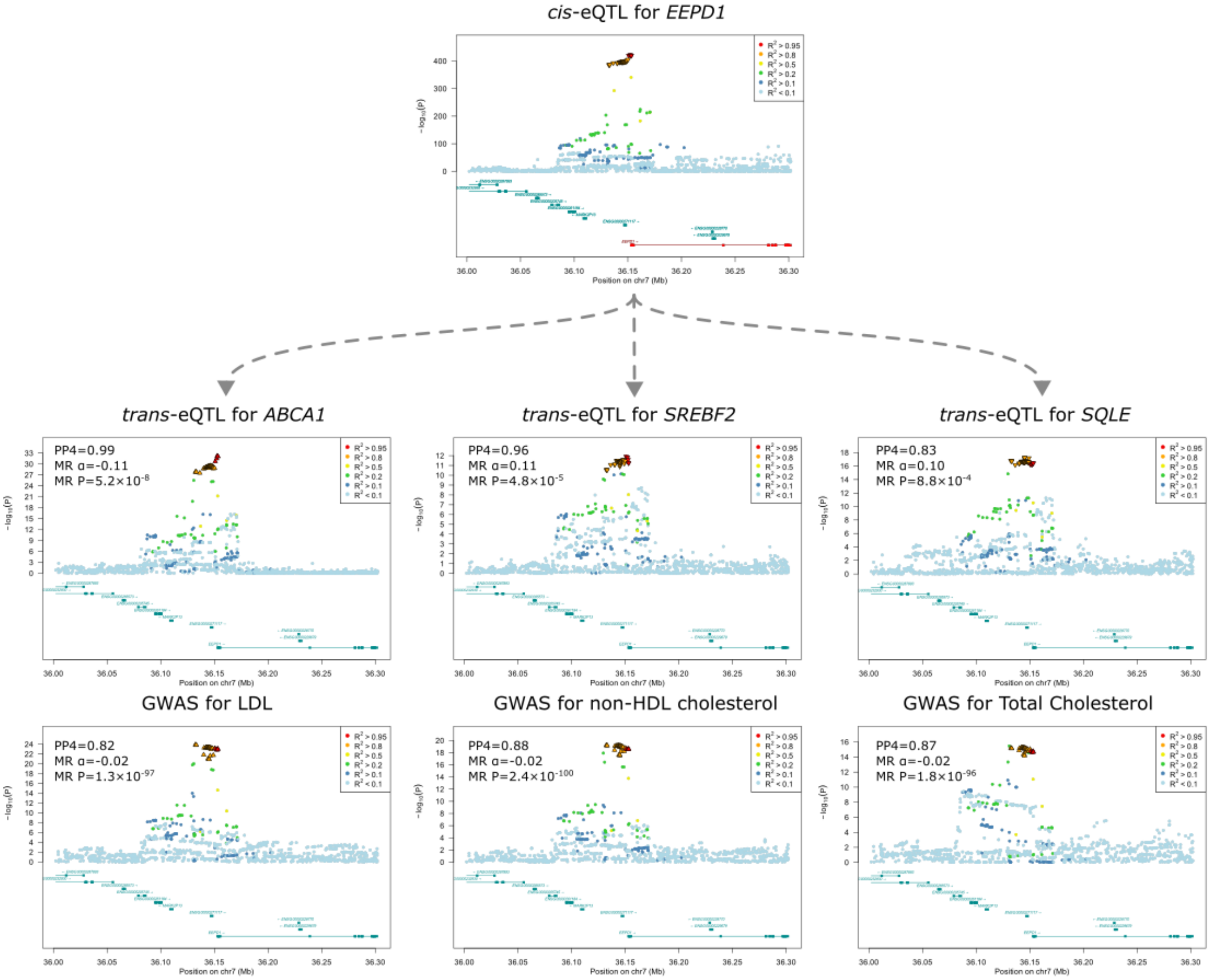


**Figure S15. Regional plots for *EEPD1* locus.** Shown are regional association plots for the *EEPD1 cis*-eQTL, three *trans*-eQTL signals colocalizing with *EEPD1 cis*-eQTL, and colocalizing GWAS signals for LDL, non-LDL cholesterol, and total cholesterol. Symbol color indicates the linkage disequilibrium (LD) $R^2$ relative to the primary *cis*-eQTL lead variant. Black outlines on symbols indicate variants in high LD ($R^2 > 0.8$, eQTLGen empirical LD) with the lead *cis*-eQTL variant, and their shape indicates the eQTL effect of the alternative allele. The *cis*-eQTL for *EEPD1* shows colocalization (colocalization PP4 > 0.8) and a significant Mendelian randomization effect (MR two-sided $P < 0.05$, **Methods**) with *trans*-eQTLs of *ABCA1*, *SREBF1,* and *SQLE*, as well as for the lipid traits shown.

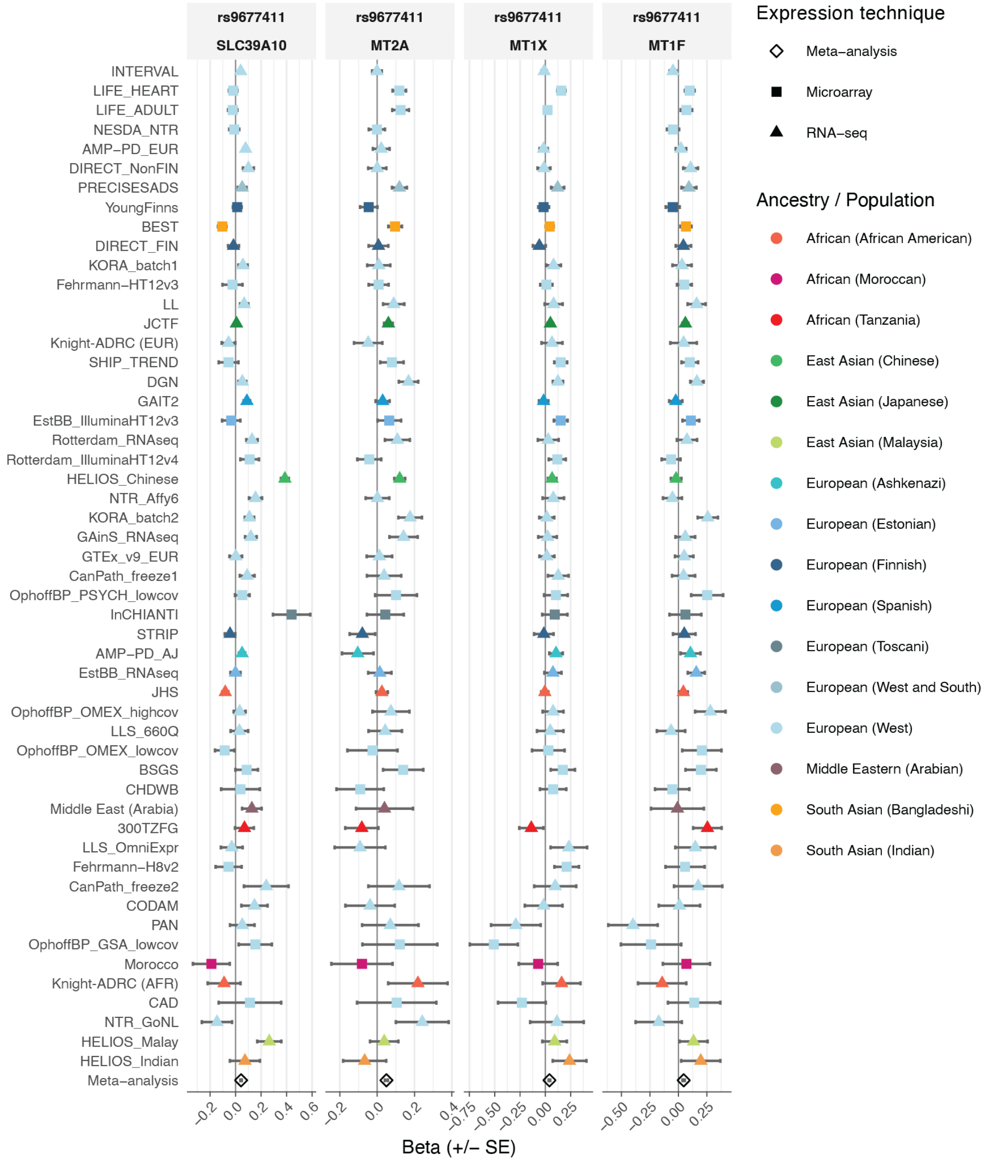
rs9677411
SLC39A10
rs9677411
MT2A
rs9677411
MT1X
rs9677411
MT1F
INTERVAL
LIFE_HEART
LIFE_ADULT
NESDA_NTR
AMP−PD_EUR
DIRECT_NonFIN
PRECISESADS
YoungFinns
BEST
DIRECT_FIN
KORA_batch1
Fehrmann−HT12v3
LL
JCTF
Knight-ADRC (EUR)
SHIP_TREND
DGN
GAIT2
EstBB_IlluminaHT12v3
Rotterdam_RNAseq
Rotterdam_IlluminaHT12v4
HELIOS_Chinese
NTR_Affy6
KORA_batch2
GAinS_RNAseq
GTEx_v9_EUR
CanPath_freeze1
OphoffBP_PSYCH_lowcov
InCHIANTI
STRIP
AMP−PD_AJ
EstBB_RNAseq
JHS
OphoffBP_OMEX_highcov
LLS_660Q
OphoffBP_OMEX_lowcov
BSGS
CHDWB
Middle East (Arabia)
300TZFG
LLS_OmniExpr
Fehrmann−H8v2
CanPath_freeze2
CODAM
PAN
OphoffBP_GSA_lowcov
Morocco
Knight-ADRC (AFR)
CAD
NTR_GoNL
HELIOS_Malay
HELIOS_Indian
Meta−analysis
−0.2 0.0 0.2 0.4 0.6
−0.2 0.0 0.2 0.4
−0.75 −0.50 −0.25 0.00 0.25
−0.50 −0.25 0.00 0.25
Beta (+/− SE)
Expression technique
Meta−analysis
Microarray
RNA−seq
Ancestry / Population
African (African American)
African (Moroccan)
African (Tanzania)
East Asian (Chinese)
East Asian (Japanese)
East Asian (Malaysia)
European (Ashkenazi)
European (Estonian)
European (Finnish)
European (Spanish)
European (Toscani)
European (West and South)
European (West)
Middle Eastern (Arabian)
South Asian (Bangladeshi)
South Asian (Indian)

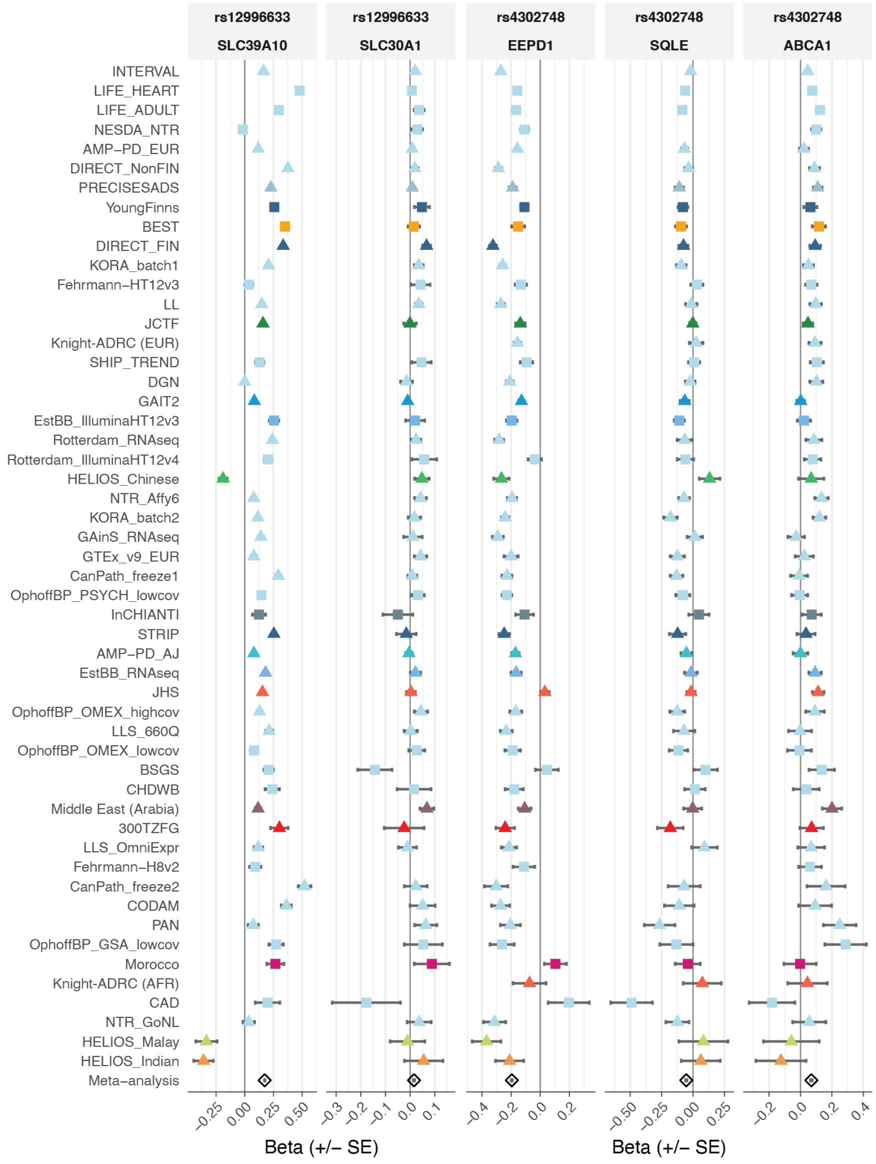
rs12996633
SLC39A10
rs12996633
SLC30A1
rs4302748
EEPD1
rs4302748
SQLE
rs4302748
ABCA1
INTERVAL
LIFE_HEART
LIFE_ADULT
NESDA_NTR
AMP−PD_EUR
DIRECT_NonFIN
PRECISESADS
YoungFinns
BEST
DIRECT_FIN
KORA_batch1
Fehrmann−HT12v3
LL
JCTF
Knight-ADRC (EUR)
SHIP_TREND
DGN
GAIT2
EstBB_IlluminaHT12v3
Rotterdam_RNAseq
Rotterdam_IlluminaHT12v4
HELIOS_Chinese
NTR_Affy6
KORA_batch2
GAinS_RNAseq
GTEx_v9_EUR
CanPath_freeze1
OphoffBP_PSYCH_lowcov
InCHIANTI
STRIP
AMP−PD_AJ
EstBB_RNAseq
JHS
OphoffBP_OMEX_highcov
LLS_660Q
OphoffBP_OMEX_lowcov
BSGS
CHDWB
Middle East (Arabia)
300TZFG
LLS_OmniExpr
Fehrmann−H8v2
CanPath_freeze2
CODAM
PAN
OphoffBP_GSA_lowcov
Morocco
Knight-ADRC (AFR)
CAD
NTR_GoNL
HELIOS_Malay
HELIOS_Indian
Meta−analysis
−0.25 0.00 0.25 0.50
−0.3 −0.2 −0.1 0.0 0.1
−0.4 −0.2 0.0 0.2
−0.50 −0.25 0.00 0.25
−0.2 0.0 0.2 0.4
Beta (+/− SE)
Beta (+/− SE)

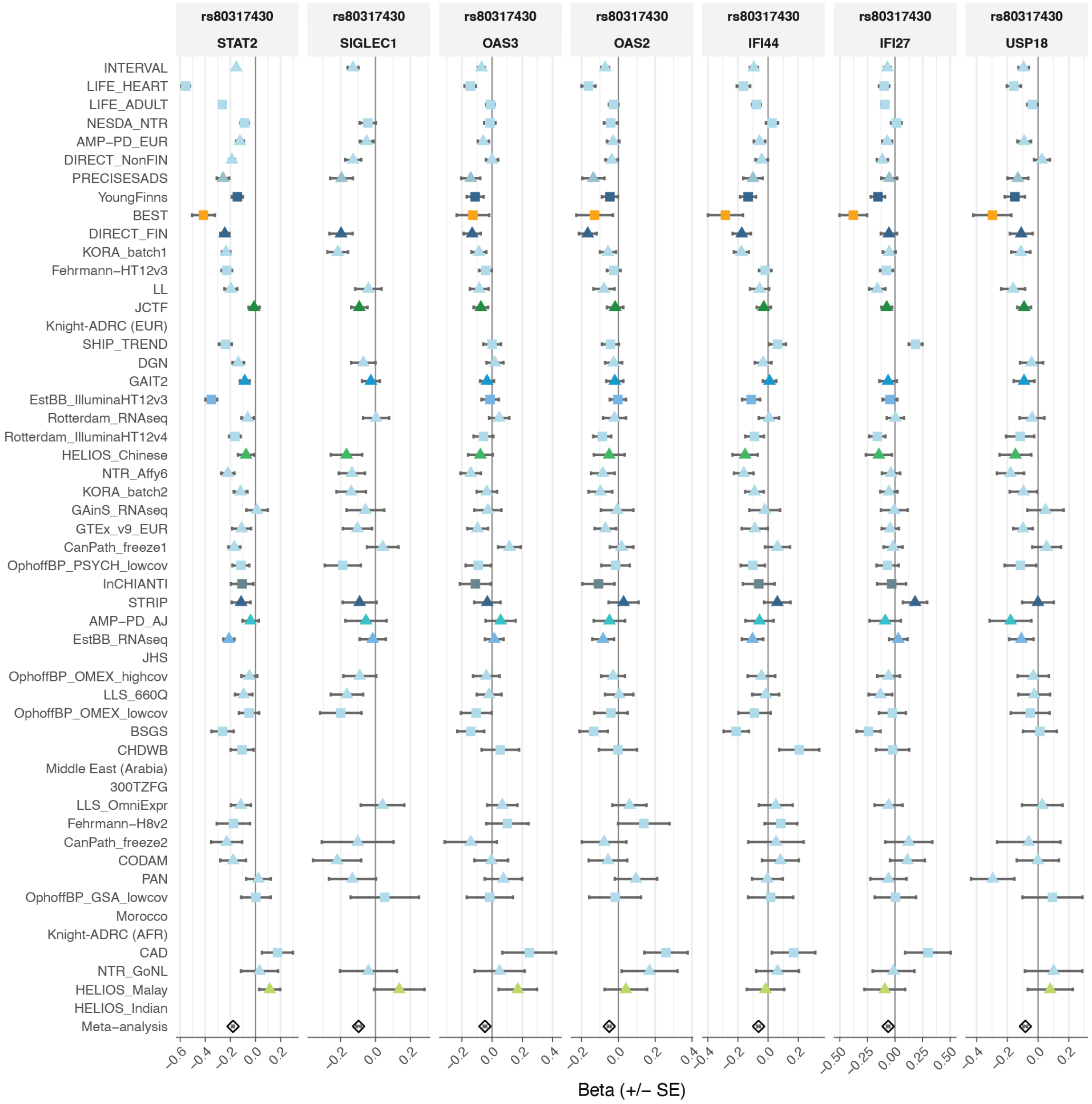


**Figure S16. Forest plots presenting the cross-cohort consistency of eQTLs underlying example gene−gene pairs,** Cohorts are ordered by sample size and colored by ancestry and / or population. Each variant−gene combination represents one of the edges from figure 5c, d, and e.

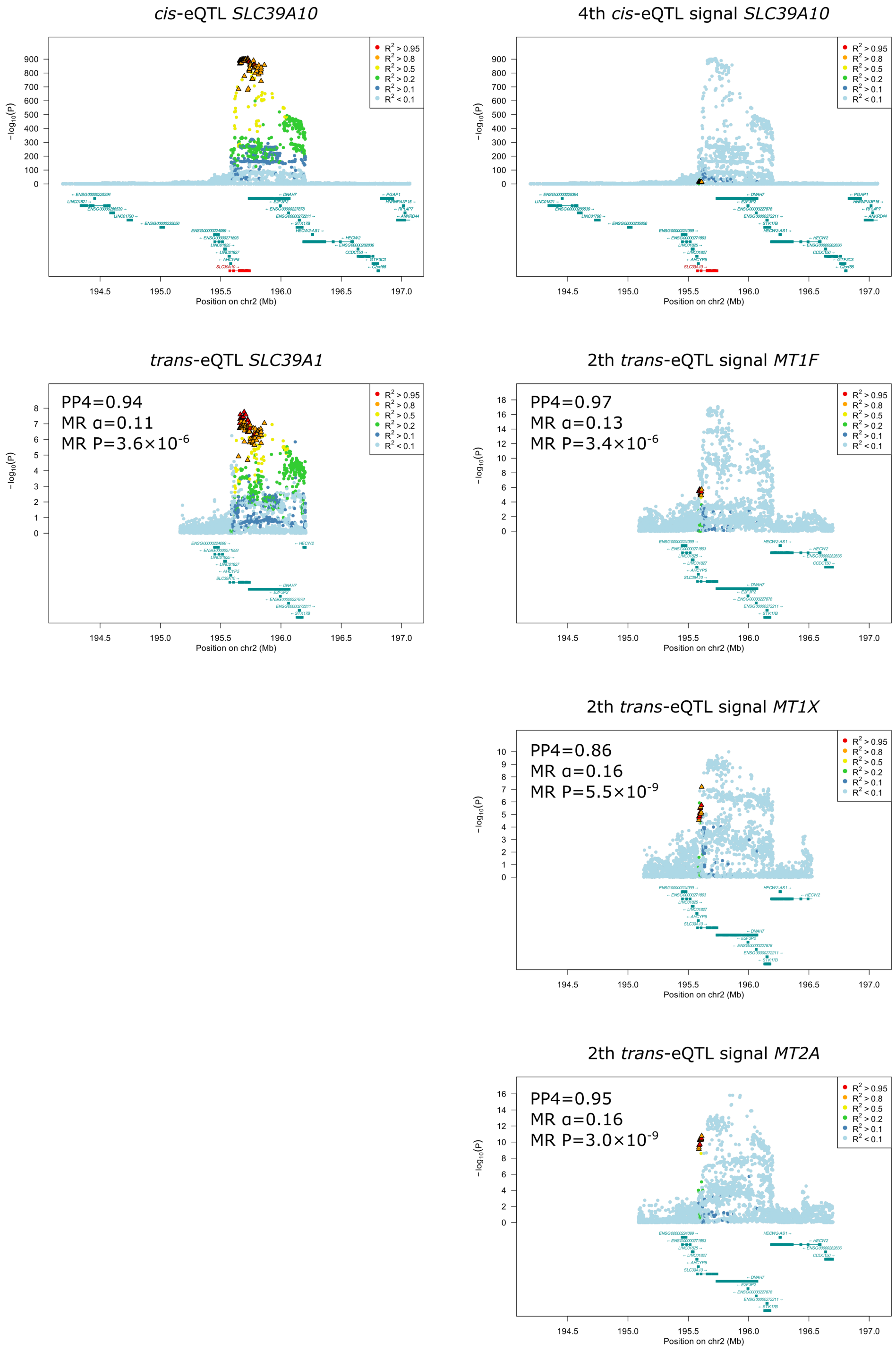


**Figure S17. Regional plots for the *SLC39A10* locus.** Shown are regional association plots for *SLC39A10 cis*-eQTLs and four *trans*-eQTL signals colocalising with *SLC39A10 cis*-eQTLs. Symbol color indicates the linkage disequilibrium (LD) $R^2$ relative to the primary *cis*-eQTL lead variant. Symbols with black outlines are variants in high LD ($R^2 > 0.8$, eQTLGen empirical LD) with the lead *cis*-eQTL variant, and their shape indicates the eQTL effect of the alternative allele. All the *trans*-eQTL signals show colocalization with *cis*-eQTL (colocalization PP4 > 0.8), as well as significant Mendelian randomization effect (MR two-sided $P < 0.05$, **Methods**). Primary *cis*-eQTL for *SLC39A10* colocalizes with overlapping primary *trans*-eQTL signal for *SLC39A1*, and quaternary *cis*-eQTL signal for

*SLC39A10* colocalises with the overlapping secondary *trans*-eQTL signal for *MT1F*, *MT1X*, and *MT2A*.

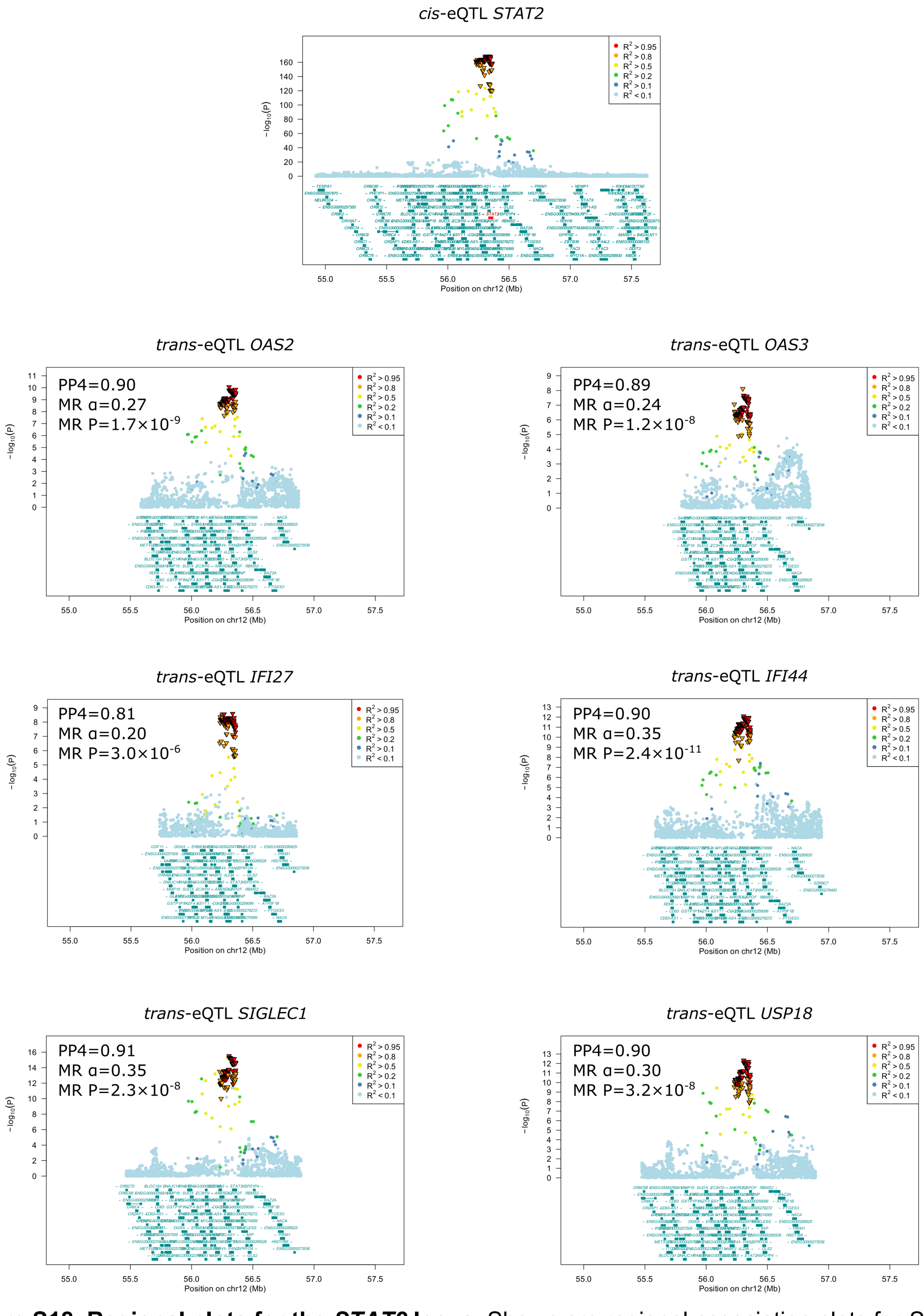


**Figure S18. Regional plots for the *STAT2* locus.** Shown are regional association plots for *STAT2* *cis*-eQTL and six *trans*-eQTL signals for interferon-response genes colocalizing with *STAT2* *cis*-eQTL. Symbol color indicates the linkage disequilibrium (LD) $R^2$ relative to the primary *cis*-eQTL lead variant. Symbols with black outlines are variants in high LD ($R^2 > 0.8$, eQTLGen empirical LD) with the lead *cis*-eQTL variant, and their shape indicates the eQTL effect of the alternative allele. All the *trans*-eQTL signals show colocalization with *cis*-eQTL (colocalization PP4 > 0.8), as well as significant Mendelian randomization effect (MR two-sided $P < 0.05$, **Methods**).

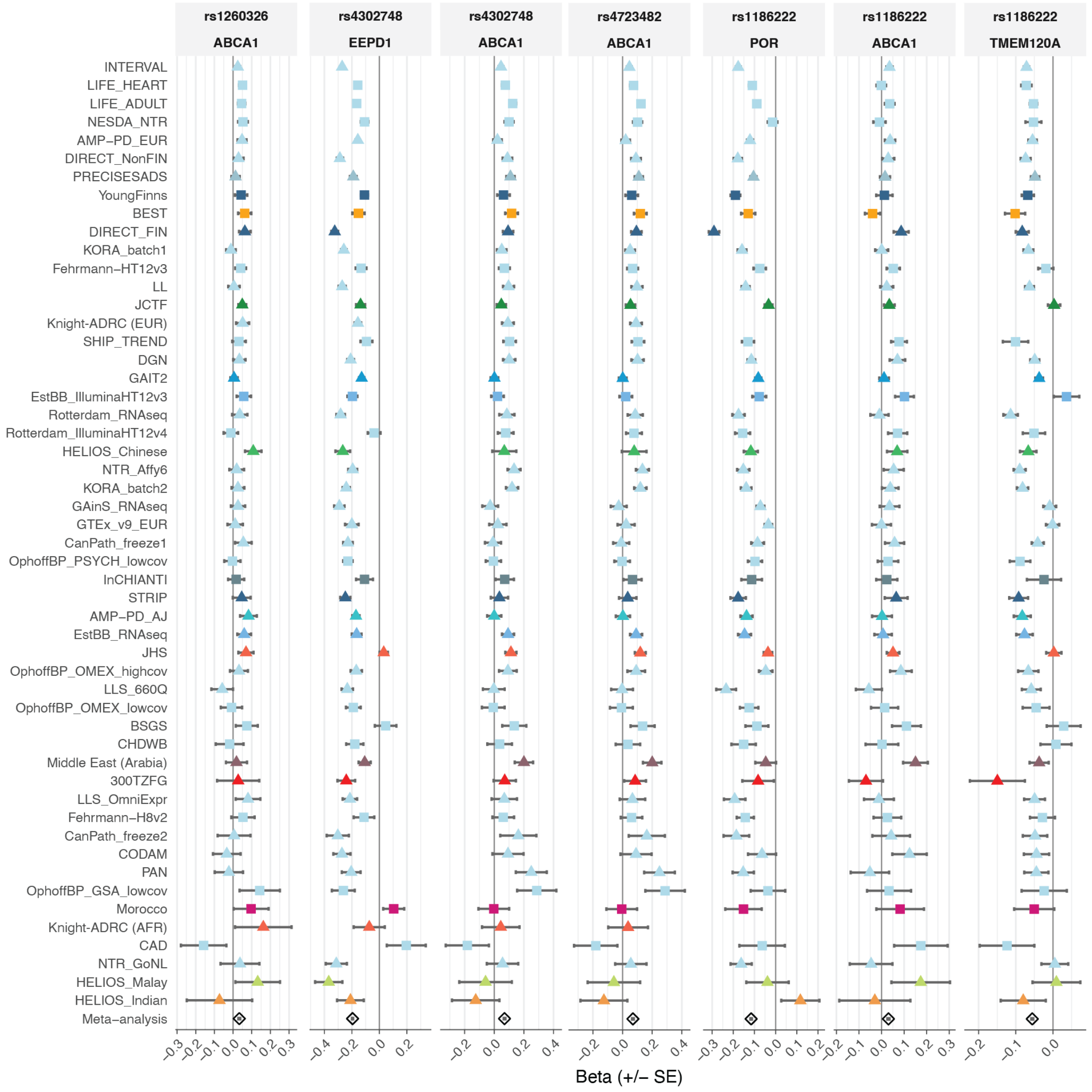
rs1260326
ABCA1
rs4302748
EEPD1
rs4302748
ABCA1
rs4723482
ABCA1
rs1186222
POR
rs1186222
ABCA1
rs1186222
TMEM120A
INTERVAL
LIFE_HEART
LIFE_ADULT
NESDA_NTR
AMP−PD_EUR
DIRECT_NonFIN
PRECISESADS
YoungFinns
BEST
DIRECT_FIN
KORA_batch1
Fehrmann−HT12v3
LL
JCTF
Knight-ADRC (EUR)
SHIP_TREND
DGN
GAIT2
EstBB_IlluminaHT12v3
Rotterdam_RNAseq
Rotterdam_IlluminaHT12v4
HELIOS_Chinese
NTR_Affy6
KORA_batch2
GAinS_RNAseq
GTEx_v9_EUR
CanPath_freeze1
OphoffBP_PSYCH_lowcov
InCHIANTI
STRIP
AMP−PD_AJ
EstBB_RNAseq
JHS
OphoffBP_OMEX_highcov
LLS_660Q
OphoffBP_OMEX_lowcov
BSGS
CHDWB
Middle East (Arabia)
300TZFG
LLS_OmniExpr
Fehrmann−H8v2
CanPath_freeze2
CODAM
PAN
OphoffBP_GSA_lowcov
Morocco
Knight-ADRC (AFR)
CAD
NTR_GoNL
HELIOS_Malay
HELIOS_Indian
Meta−analysis
Beta (+/− SE)

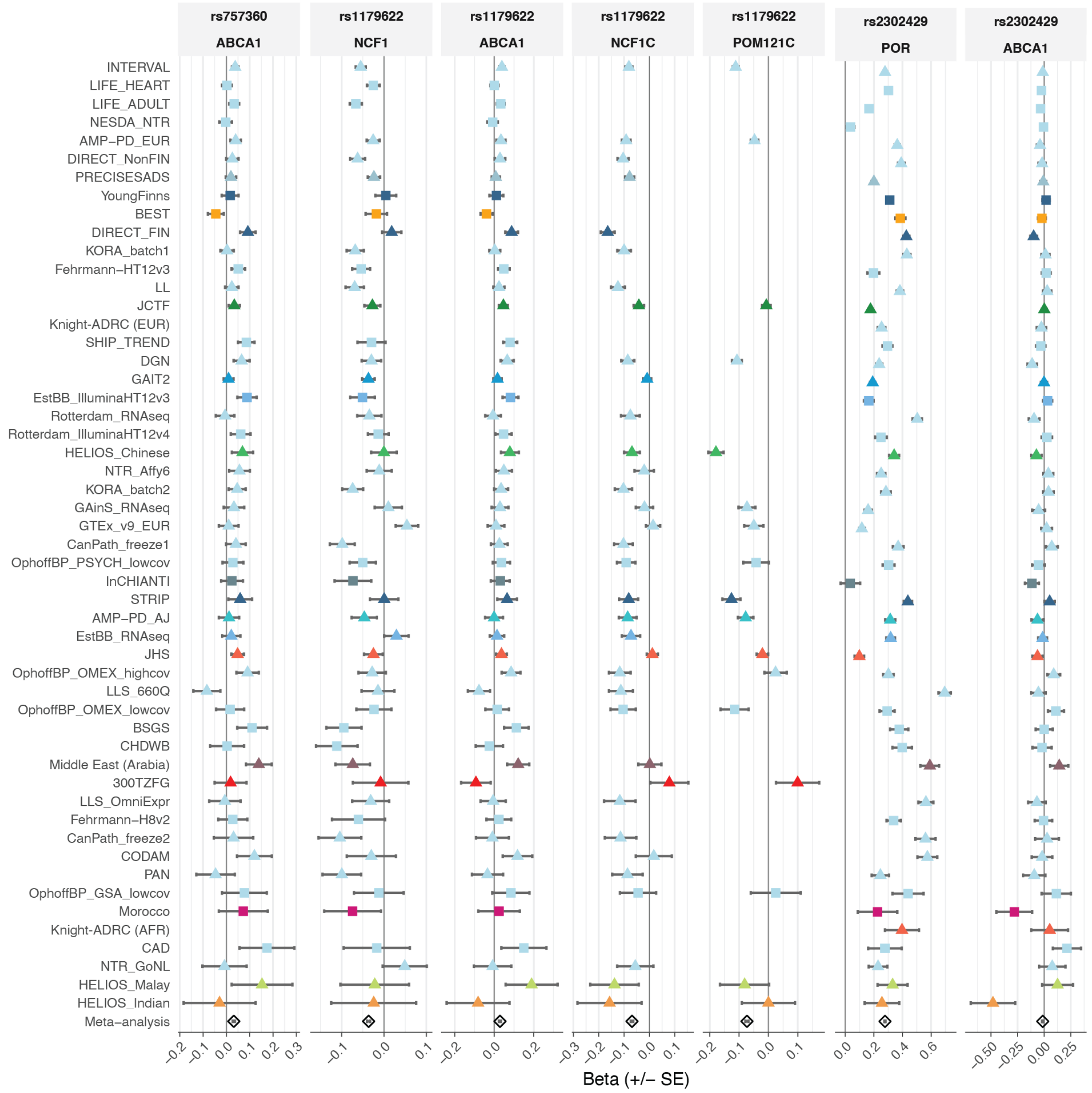
rs757360
ABCA1
rs1179622
NCF1
rs1179622
ABCA1
rs1179622
NCF1C
rs1179622
POM121C
rs2302429
POR
rs2302429
ABCA1
INTERVAL
LIFE_HEART
LIFE_ADULT
NESDA_NTR
AMP−PD_EUR
DIRECT_NonFIN
PRECISESADS
YoungFinns
BEST
DIRECT_FIN
KORA_batch1
Fehrmann−HT12v3
LL
JCTF
Knight-ADRC (EUR)
SHIP_TREND
DGN
GAIT2
EstBB_IlluminaHT12v3
Rotterdam_RNAseq
Rotterdam_IlluminaHT12v4
HELIOS_Chinese
NTR_Affy6
KORA_batch2
GAinS_RNAseq
GTEx_v9_EUR
CanPath_freeze1
OphoffBP_PSYCH_lowcov
InCHIANTI
STRIP
AMP−PD_AJ
EstBB_RNAseq
JHS
OphoffBP_OMEX_highcov
LLS_660Q
OphoffBP_OMEX_lowcov
BSGS
CHDWB
Middle East (Arabia)
300TZFG
LLS_OmniExpr
Fehrmann−H8v2
CanPath_freeze2
CODAM
PAN
OphoffBP_GSA_lowcov
Morocco
Knight-ADRC (AFR)
CAD
NTR_GoNL
HELIOS_Malay
HELIOS_Indian
Meta−analysis
−0.2 −0.1 0.0 0.1 0.2 0.3
−0.1 0.0 0.1
−0.2 0.0 0.2
−0.3 −0.2 −0.1 0.0 0.1
−0.2 −0.1 0.0 0.1
0.0 0.2 0.4 0.6
−0.50 −0.25 0.00 0.25
Beta (+/− SE)

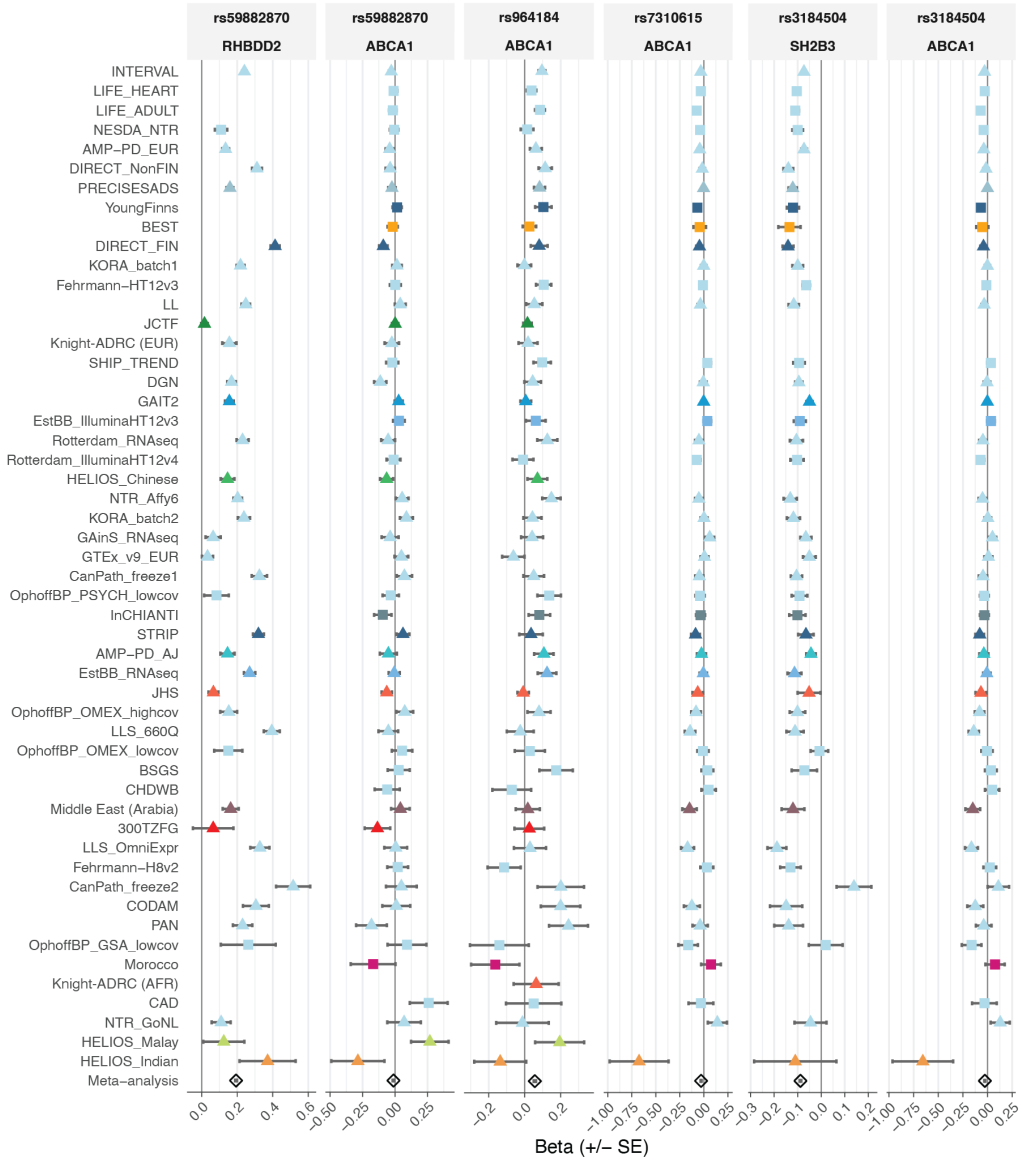

rs59882870
RHBDD2
rs59882870
ABCA1
rs964184
ABCA1
rs7310615
ABCA1
rs3184504
SH2B3
rs3184504
ABCA1
INTERVAL
LIFE_HEART
LIFE_ADULT
NESDA_NTR
AMP–PD_EUR
DIRECT_NonFIN
PRECISESADS
YoungFinns
BEST
DIRECT_FIN
KORA_batch1
Fehrmann–HT12v3
LL
JCTF
Knight-ADRC (EUR)
SHIP_TREND
DGN
GAIT2
EstBB_IlluminaHT12v3
Rotterdam_RNAseq
Rotterdam_IlluminaHT12v4
HELIOS_Chinese
NTR_Affy6
KORA_batch2
GAinS_RNAseq
GTEx_v9_EUR
CanPath_freeze1
OphoffBP_PSYCH_lowcov
InCHIANTI
STRIP
AMP–PD_AJ
EstBB_RNAseq
JHS
OphoffBP_OMEX_highcov
LLS_660Q
OphoffBP_OMEX_lowcov
BSGS
CHDWB
Middle East (Arabia)
300TZFG
LLS_OmniExpr
Fehrmann–H8v2
CanPath_freeze2
CODAM
PAN
OphoffBP_GSA_lowcov
Morocco
Knight-ADRC (AFR)
CAD
NTR_GoNL
HELIOS_Malay
HELIOS_Indian
Meta–analysis
Beta (+/– SE)

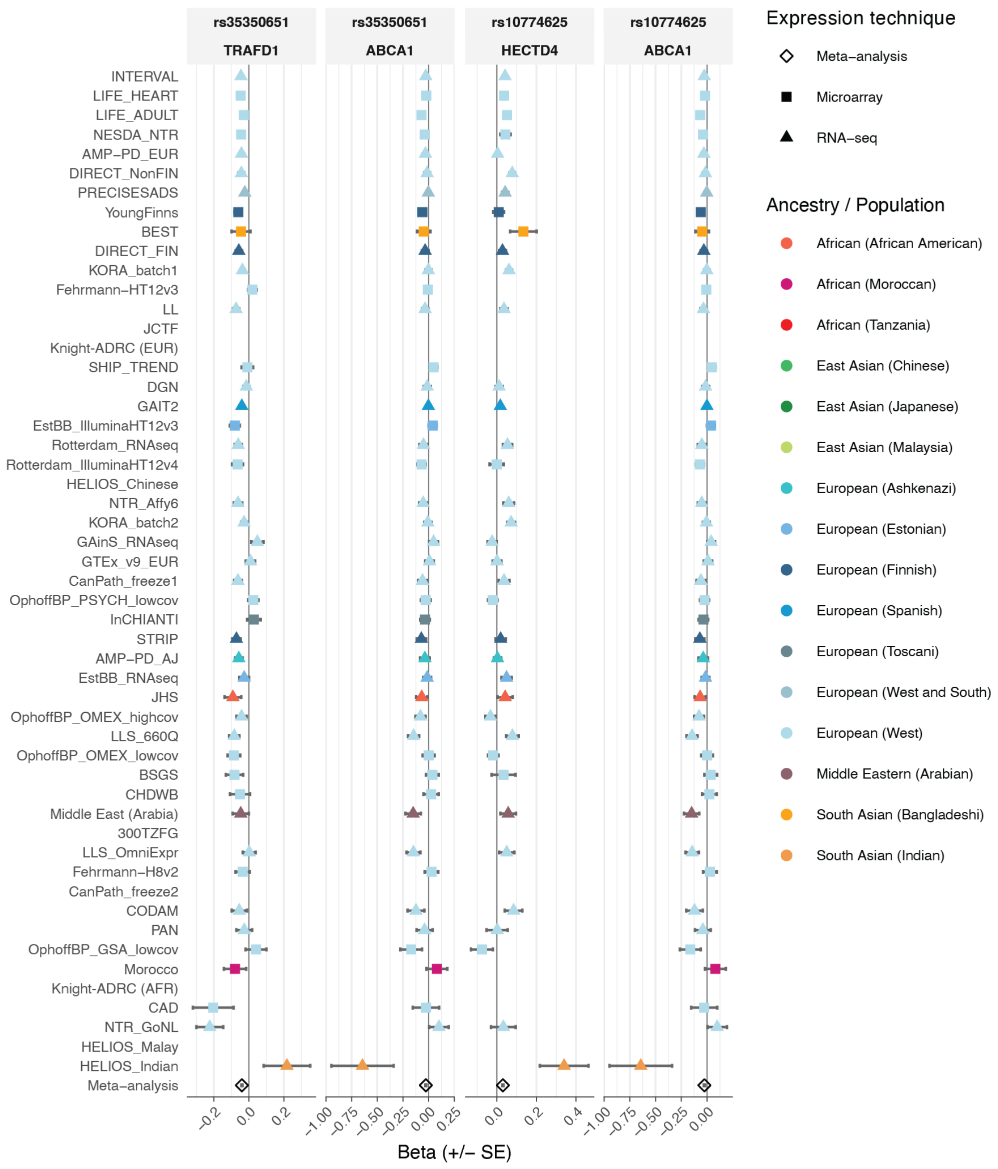


**Figure S19. Forest plots presenting the cross-cohort consistency of eQTL effects that are underlying or implicated by the network in which LDL cholesterol variants are linked to *ABCA1*.** Cohorts are ordered by sample size and colored by ancestry and / or population. Each variant−gene combination represents one of the edges from figure 5f.

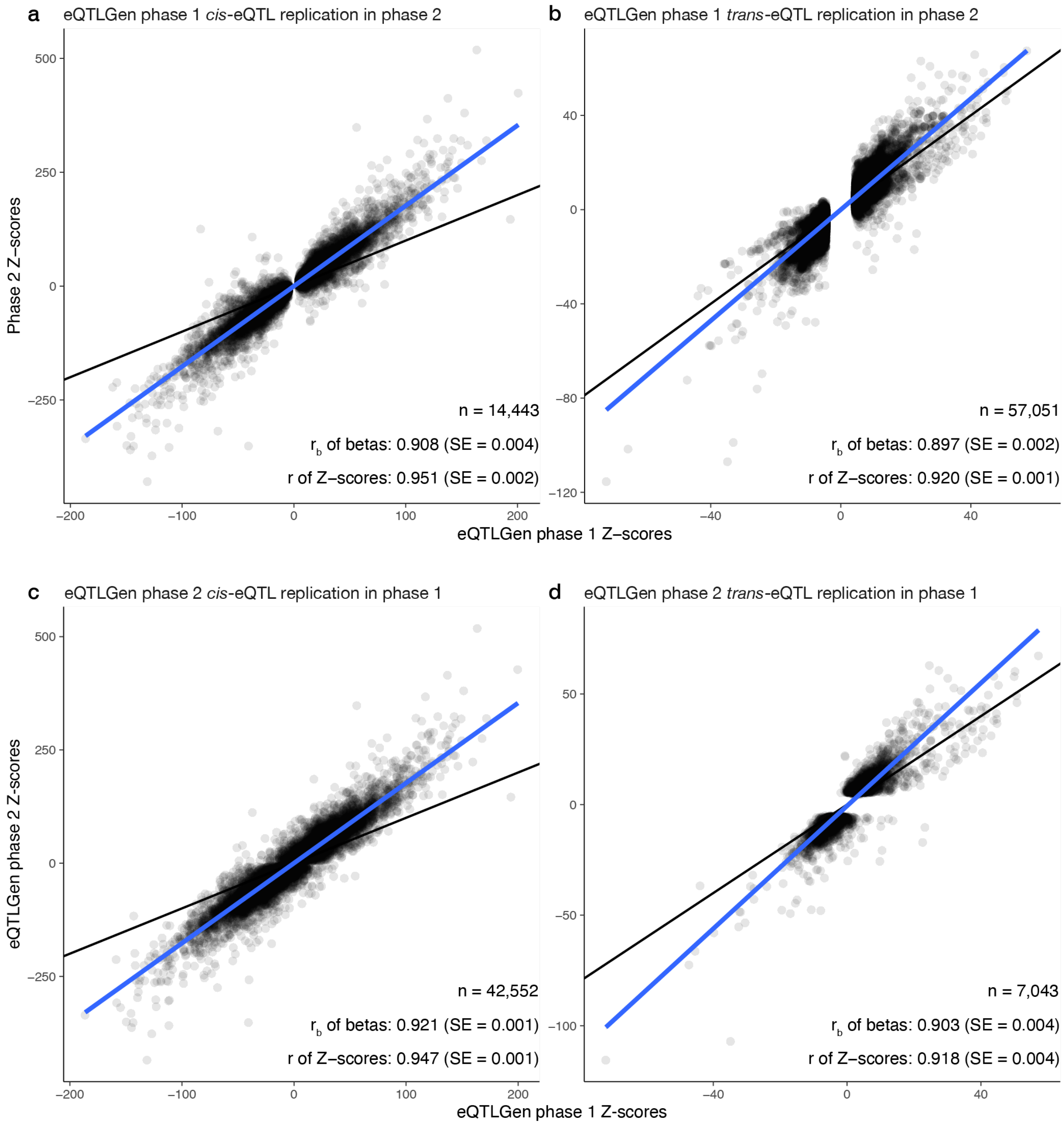


**Figure S20. Comparison of eQTLGen phase 1 to eQTLGen phase 2. a,b** Scatteplots wherein each dot represents a fine-mapped eQTLGen phase 2 eQTL. For these effects, corresponding Z-scores were extracted from eQTLGen phase 1. The blue line indicates the univariate linear regression line through the Z-scores. **c,d** eQTLGen phase 1 eQTLs looked up the phase 2 mata-analysis data. The blue line indicates the univariate linear regression line through the Z-scores. **c** Scatterplots wherein each dot represents the top-associated cis-eQTL per gene from eQTLGen phase 1. **d**. All trans-eQTLs from eQTLGen phase 1 looked up in phase 2. All scatterplots and $r_b$ values indicate an expected level of correspondence between the estimates two studies.

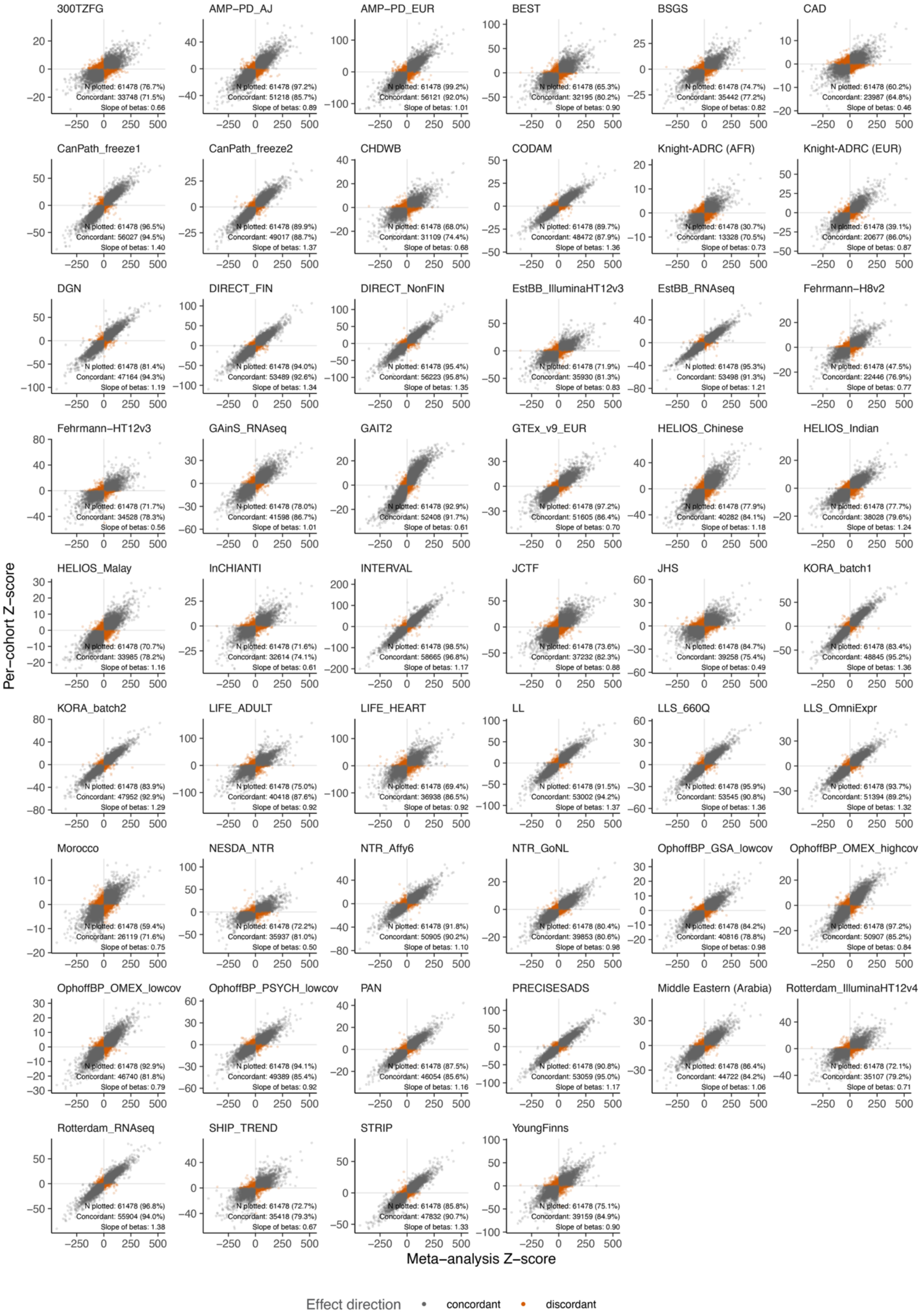


**Figure S21. Replication of fine-mapped *cis*-eQTLs in individual cohorts.** Shown are scatterplots wherein each dot represents a fine-mapped cis-eQTL. The X-axes represent the Z-scores of the cis-eQTL after meta-analysis. The Y-axes represent the Z-scores of the cis-eQTLs for the individual cohorts. Orange dots represent discordant effects in the cohort relative to the meta-analyis. Grey dots represent concordant effects in the cohort relative to the meta-analyis. Taking population background and expression platform into account, *cis*-eQTLs show high concordance between the meta-analysis and the individual cohorts.

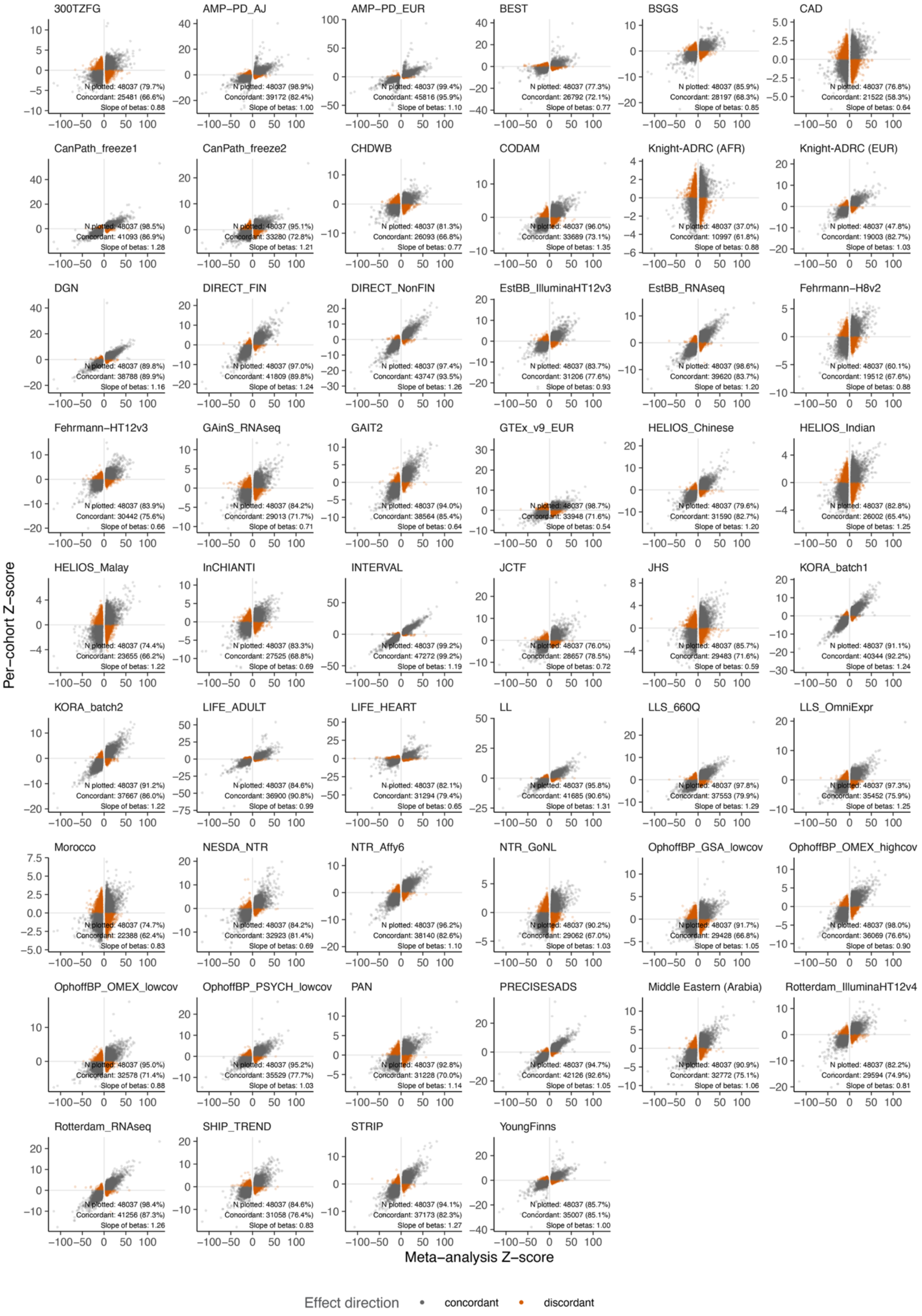


**Figure S22. Replication of fine-mapped *trans*-eQTLs in individual cohorts.** Shown are scatterplots wherein each dot represents a fine-mapped trans-eQTL. The X-axes represent the Z-scores of the trans-eQTL after meta-analysis. The Y-axes represent the Z-scores of the trans-eQTLs for the individual cohorts. Orange dots represent discordant effects in the cohort relative to the meta-analyis. Grey dots represent concordant effects in the cohort relative to the meta-analyis. Taking population background and expression platform into account, trans-eQTLs show high concordance between the meta-analysis and the individual cohorts.

### Supplementary notes

#### Supplementary note 1: Calculation of false discovery rate (FDR) for *cis*- and *trans*-eQTLs:

Given 16,742 genome-wide scans and an effective number of 2,158,129 independent common variants (at MAF > 1%) in the 1000 Genomes 30× WGS reference panel, accounting for correlation between variants[1] we expect ~1,807 false positive *trans*-eQTLs at P-value < $5\times10^{-8}$. Since we identified 42,994 primary *trans*-eQTLs at that significance threshold we estimate the *trans*-eQTL detection FDR to be 4.2%. We estimated the false discovery rate (FDR) for *cis*-eQTLs by noting that, within a ±1 Mb window around each gene (≈$6.5\times10^{-4}$ of the genome), the effective number of independent common variants (MAF > 1%) is ≈1,392, yielding an expected 1.11 false positives across 15,981 genes tested for *cis*-eQTLs at P-value < $5\times10^{-8}$. Given the 15,146 *cis*-eGenes for which we identify a primary eQTL signal at this threshold, we estimate that this threshold corresponds to an FDR < 1%.

### Cohort Information

#### AMP-PD

##### Cohort description

The Accelerating Medicines Partnership – Parkinson's Disease (AMP-PD) is a public–private partnership among the NIH, FDA, foundations, and industry partners, established to accelerate the discovery of biomarkers and therapeutics for Parkinson's disease. AMP-PD integrates multi-modal omics and clinical data from deeply phenotyped Parkinson's disease cohorts, including the Parkinson's Progression Markers Initiative (PPMI), BioFIND, Harvard Biomarkers Study (HBS), and the Parkinson's Disease Biomarkers Program (PDBP). RNA expression data within AMP-PD were generated primarily from whole blood using standardized library preparation and sequencing protocols across contributing cohorts. All AMP-PD data are harmonized and processed using uniform workflows to ensure comparability across studies (see https://amp-pd.org/transcriptomics-data).
All data preprocessing, normalization, and quality control followed AMP-PD pipelines and standard operating procedures, including alignment to GRCh38 and quantification with consistent annotation references.

##### Ethics approval

All AMP-PD participating cohorts obtained written informed consent from participants in accordance with the Declaration of Helsinki and approval by their respective Institutional Review Boards. The use of de-identified data in the AMP-PD Knowledge Platform was reviewed and approved by the appropriate data access committees. Access to AMP-PD data is governed by the AMP-PD Data Use Agreement and complies with NIH and partner institutional ethical guidelines.

##### Methods: expression data

RNA-sequencing (RNA-seq) data were generated from whole-blood samples using Illumina TruSeq Stranded Total RNA or TruSeq Stranded mRNA library preparation kits, following cohort-specific protocols. Sequencing was performed on Illumina HiSeq or NovaSeq platforms, with paired-end reads (75–150 bp). Raw FASTQ files were aligned to the GRCh38 reference genome using STAR aligner (v2.7), and gene-level quantification was performed using featureCounts. Expression matrices were harmonized across studies and normalized using trimmed mean of M values (TMM) or variance-stabilizing transformation as appropriate. Quality control steps included removal of samples with low mapping rates, abnormal library complexity, or outlier expression profiles, as well as filtering of genes with low expression across all samples. Detailed description of the AMP-PD transcriptomic data generation and processing pipeline is available at https://amp-pd.org/transcriptomics-data.

##### Methods: genotyping data

Whole-genome sequencing (WGS) data for the AMP-PD cohort were generated from whole-blood DNA using the Illumina HiSeq X Ten platform, with paired-end 150 bp reads. Sequencing was performed by Macrogen and the Uniformed Services University of the Health Sciences (USUHS) under the coordination of the AMP-PD consortium. Sequencing reads were aligned to the GRCh38 (DH) reference genome using the Broad Institute's Functional Equivalence (FE) pipeline, producing CRAM and gVCF files for each sample. Joint genotyping across all samples was performed using the Broad Joint Genotyping pipeline, and variant annotation was carried out with the Variant Effect Predictor (VEP). Quality-control (QC) procedures followed the AMP-PD WGS v2.5 pipeline and included evaluation of: Mean coverage ≥ 25×, Contamination rate (FREEMIX < 3%),

Transition/transversion (Ti/Tv) ratio ≥ 2.0, Missing genotype rate < 5%, Sex concordance, sample duplication, and population outlier detection by principal component analysis (PCA). The resulting dataset includes per-sample CRAM, gVCF, and QC metric files, as well as joint-genotyped per-chromosome VCFs and PLINK-formatted files. All data were processed and harmonized on the Google Cloud Platform (Terra) to ensure consistency across cohorts and analyses. Detailed information on the AMP-PD WGS processing and quality metrics is publicly available at https://amp-pd.org/whole-genome-data.

#### Data availability statement

AMP-PD data are available through controlled access on the AMP-PD Knowledge Platform (https://amp-pd.org). Access requires registration, an approved Data Use Agreement and project description. Summary-level and processed data are made available under the partnership's data-sharing policies.

#### Acknowledgements

We acknowledge the AMP-PD Consortium and contributing study partners: the Michael J. Fox Foundation for Parkinson's Research, the National Institute of Neurological Disorders and Stroke, the U.S. Food and Drug Administration, and contributing industry and academic partners. We thank all study participants and their families for their valuable contributions to Parkinson's disease research. RNA-seq data generation and harmonization were performed by Verily Life Sciences and the Broad Institute, with coordination and data hosting by the AMP-PD Knowledge Platform team.

#### Funding information

AMP-PD is a collaborative effort supported by the Foundation for the NIH and funded by partners including the Michael J. Fox Foundation for Parkinson's Research, Celgene, Pfizer, Biogen, AbbVie, GSK, Sanofi, Verily Life Sciences, and Takeda. The project is implemented through the Foundation for the NIH's Accelerating Medicines Partnership program.

### LIFE-Heart

#### Cohort description

LIFE-Heart is a cohort study of patients with suspected or confirmed coronary artery disease collected at the Heart-Center of Leipzig. Details of the study can be found elsewhere[1].

#### Ethics approval

LIFE-Heart meets the ethical standards of the Declaration of Helsinki, was approved by the Ethics Committee of the medical Faculty of the University of Leipzig (Reg. No 276/05-ek) and is registered at ClinicalTrials.gov (No NCT00497887). Written informed consent was collected from all patients.

#### Methods: expression data

PBMC-based gene-expression was measured by Illumina HumanHT-12 v4 Expression BeadChip. Pre-processing was performed analogously to LIFE-Adult as described previously by Kirsten et al.[2]. Briefly, PBMC isolation was performed using Cell Preparation Tubes (CPT, Becton Dickinson), total RNA was extracted using TRIzol reagent (Invitrogen) and 500 ng RNA per sample were ethanol precipitated with GlycoBlue (Invitrogen) as carrier and hybridized to Illumina HT-12 v4 Expression BeadChips (Illumina, San Diego, CA, USA). Raw data of all 47,231 gene-expression probes available was extracted by Illumina GenomeStudio without additional background correction.
Two criteria were used to remove samples of low quality: First, the number of detected gene-expression probes of a sample was required to be within ± 3 interquartile ranges (IQR) from the median. Second, the Mahalanobis distance of several quality characteristics of each sample (signal of biotin-control-probes, signal of low-concentration control probes, signal of medium-concentration control probes, signal of mismatch control probes, signal of negative control probes and signal of perfect-match control probes) as described in Cohen Freue et al.[3] had to be within median + 3 x IQR. Overall, of the assayed 4,509 samples, 122 samples were excluded for quality reasons. The raw expression values of 4,322 samples for which genotype data was available were processed for further quality control within the eQTLGen Phase 2 analysis pipelines, as described in the main methods section.

#### Methods: genotyping data

Genotyping was performed using the Affymetrix Axiom Technology with custom option (Axiom-CADLIFE). Genotype calling was performed with Affymetrix Power Tools version 1.12. Sample QC comprised call rate (>97%), hetero- or homozygosity excess (outliers of mean squared differences of observed and expected genotypes), sex-mismatch, cryptic relatedness and outliers of PCA (6SD criterion of Eigenstrat software[4], leaving 5,688 individuals for further analysis. Low quality SNPs defined by low call-rate (<90% plate-wise call rate corresponding to <94.2% overall call- rate), deviation from HWE (P-value $<10^{-6}$) or plate-association (P-value $<10^{-7}$) were filtered. Further quality control and eQTL analysis were performed on the 4,322 samples with gene expression data, using the eQTLGen Phase 2 analysis pipelines, as described in the main methods section.

#### Data availability statement

For access to any data or biosamples, it is mandatory to submit a detailed written proposal describing the background, objectives, methods, timelines, names and affiliations of all researchers involved; the type and scope of requested data and biomaterial; the publication and exploitation strategy; and how results and newly generated data will be returned for further use. Upon review and approval by the Use-

and-Access Committee, data and samples may be provided. Specific contracts and data or material transfer agreements may be necessary, particularly for researchers from countries not bound to the EU General Data Privacy Regulation. All enquiries should be directed via e-mail to Dr. Matthias Nüchter, LIFE Management Cluster, Philipp-Rosenthal-Straße 27, 04103 Leipzig, Tel.: 0049341 - 97 16720 E-Mail:.

#### Acknowledgements

The authors acknowledge all participants of the LIFE-Heart study for spending their time and donating blood. We thank Annegret Unger and Kay Olischer for running the study ambulance, Kerstin Wirkner for running the LIFE study center, and Ronny Dathe for database development, . We thank all study participants, volunteers and study personnel who made this work possible. We thank Lesca Holdt very much for helping to create the LIFE-Heart gene-expression data, and thank Sylvia Henger very much for data quality control.

#### Funding information

This publication is supported by LIFE – Leipzig Research Centre for Civilization Diseases, an organizational unit affiliated to the Medical Faculty of the University of Leipzig. LIFE is funded by means of the European Union, by the European Regional Development Fund (ERDF) and by funds of the Free State of Saxony within the framework of the excellence initiative (project numbers 713-241202, 713-241202, 14505/2470, 14575/2470). Furthermore, this work was funded by the German Federal Ministry of Education and Research (BMBF) within the project "Center for Scalable Data Analytics and Artificial Intelligence (ScaDS.AI) Dresden/Leipzig" (BMBF grant 01IS18026B).

### LIFE-Adult

#### Cohort description

LIFE-Adult is a population-based cohort study that recruited about 10,000 randomly selected inhabitants of the city of Leipzig, Saxony, Germany. Further information can be found elsewhere[1].

#### Ethics approval

The LIFE-Adult-Study is conducted in accordance with the Declaration of Helsinki and was approved by the Ethics Committee of the Medical Faculty of Leipzig University (approval numbers 263–2009-14122009, 263/09-ff, 201/17-ek). Written informed consent was obtained from all participants.

#### Methods: expression data

Whole blood was collected in Tempus Blood RNA Tubes (Life Technologies) and relocated to - 80°C before further processing. Isolated RNA was processed and hybridized to Illumina HT-12 v4 Expression BeadChips (Illumina, San Diego, CA, USA) and measured on the Illumina HiScan. Raw data of all 47,231 probes was extracted by Illumina GenomeStudio in all initially included individuals. Three criteria were used to remove samples of low quality (processed within R / Bioconductor): First, the number of gene-expression probes detected in a sample was required to be within ± 3 interquartile ranges (IQR) from the median. Second, the Mahalanobis distance of several quality characteristics of each sample (signal of Ambion$^{TM}$ ERCC Spike-In control probes, signal of biotin-control-probes, signal of low-concentration control probes, signal of medium-concentration control probes, signal of mismatch control probes, signal of negative control probes and signal of perfect-match control probes) as described by Cohen Freue et al.[2], had to be within median + 3 x IQR. Third, Euclidean distances of expression values as described elsewhere[3], had to be within 4 x IQR from the median. In total, of the assayed 3,526 samples, 107 samples were excluded for quality reasons. The raw expression values of 3,190 samples for which genotype data was available were processed for further quality control within the eQTLGen Phase 2 analysis pipelines, as described in the main methods section.

#### Methods: genotyping data

Genomic DNA was extracted from peripheral blood leukocytes applying an automated protocol on the Autopure LS instrument (Qiagen, Hilden, Germany) as recommended by the manufacturer. Chip-genotyping was done applying Axiom Genome-Wide CEU 1 Array Plate (Affymetrix, Inc., Santa Clara, California, USA) technology including 587,352 Single Nucleotide Polymorphisms (SNPs) according the manufacturer's instructions. Sample quality filtering removed all individuals with dish-QC < 0.82, call rate < 0.97, reported vs. genotype-wise computed sex mismatch and cryptic relatedness. Using about 200,000 high-quality SNPs (call rate > .998), PCA was performed using EIGENSOFT 3.0. Outliers according to the standard-cutoff 6SD were removed, leaving 7,660 individuals for further analysis. SNP quality filtering removed SNPs with call rate < 0.97, Hardy-Weinberg P-value $\leq 10^{-6}$, plate association p-value $\leq 10^{-7}$ and inappropriate cluster-plot quality metrics (Fisher's Linear Discriminant < 3.6, heterozygous cluster strength offset < -0.1 or invalid homozygote ratio offset), leaving 535,632 variants. Further quality control and eQTL analysis were performed on the 3,190 samples with gene expression data, using the eQTLGen Phase 2 analysis pipelines, as described in the main methods section.

#### Data availability statement

For access to any data or biosamples, it is mandatory to submit a detailed written proposal describing the background, objectives, methods, timelines, names and affiliations of all researchers involved; the type and scope of requested data and biomaterial; the publication and exploitation strategy; and how results and newly generated data will be returned for further use. Upon review and approval by the Use-and-Access Committee, data and samples may be provided. Specific contracts and data or material transfer agreements may be necessary, particularly for researchers from countries not bound to the EU General Data Privacy Regulation. All enquiries should be directed via e-mail to Dr. Matthias Nüchter, LIFE Management Cluster, Philipp-Rosenthal-Straße 27, 04103 Leipzig, Tel.: 0049341 - 97 16720 E-Mail:.

#### Acknowledgements

We thank the participants of LIFE-Adult very much for their time and blood samples. Furthermore, we thank the entire LIFE-study team for their commitment.

#### Funding information

This publication is supported by LIFE – Leipzig Research Centre for Civilization Diseases, an organizational unit affiliated to the Medical Faculty of the University of Leipzig. LIFE is funded by means of the European Union, by the European Regional Development Fund (ERDF) and by funds of the Free State of Saxony within the framework of the excellence initiative (project numbers 713-241202, 713-241202, 14505/2470, 14575/2470).
Furthermore, this work was funded by the German Federal Ministry of Education and Research (BMBF) within the project "Center for Scalable Data Analytics and Artificial Intelligence (ScaDS.AI) Dresden/Leipzig" (BMBF grant 01IS18026B).

### Rotterdam Study - RNA-seq

#### Cohort description

The Rotterdam Study[1,2] is a single-center, prospective population-based cohort study conducted in Rotterdam, the Netherlands. Subjects were included in different phases from the start of the study in 1998, with a total of 14,926 men and women aged 45 years and over included as of late 2008. The main objective of the Rotterdam Study is to investigate the prevalence and incidence of risk factors for chronic diseases to contribute to better prevention and treatment of such diseases in the elderly. A subset of the Rotterdam Study is also part of the BIOS Consortium, and these samples have been RNA-sequenced (RNA-seq). We excluded samples that were also measured on the Illumina expression arrays.

#### Ethics approval

The Rotterdam Study has been approved by the institutional review board (IRB) (medical ethics committee) of the Erasmus Medical Center and by the review board of The Netherlands Ministry of Health, Welfare and Sports.

#### Methods: expression data

RNA-seq gene expression data was generated in The Human Genotyping facility (HugeF) (Erasmus MC, Rotterdam, the Netherlands, https://genomicserasmusmc.nl/). RNA-seq extraction and processing has been described before for a subset of the data[3]. Briefly, RNA was extracted from whole blood and paired-end sequenced using Illumina HiSeq 2000. Reads were aligned using STAR 2.3.0e[4] while masking common (minor allele frequency (MAF) > 0.01) single nucleotide polymorphisms (SNPs) from the Genome of the Netherlands (GoNL)[5]. Gene-level expression was quantified using HTSeq[6]. FastQC (http://www.bioinformatics.babraham.ac.uk/projects/fastqc/) was used to check quality metrics, and we removed individuals with < 70% of reads mapping to exons (exon mapped/genome). From the 3,998 BIOS samples for which expression data was available, 723 samples also had filtered quality-controlled genotype data available in the Rotterdam Study. As per the eQTLGen Data QC pipeline, we further filtered this set of overlapping samples by removing outliers (maximum standard deviation (SD) of 3), likely contaminated samples (angle of contamination area = 30°), and likely swapped samples (slope of the line to discriminate between male and female sex expression = 45°). These QC steps resulted in 708 samples with expression data on 19,855 genes.

#### Methods: genotyping data

For genotyping, whole blood was also collected in EDTA tubes and DNA was isolated using a manual salting-out protocol. Genotyping for this sample subset was performed on the Illumina 610K quad beadchip array (Illumina) according to manufacturer's specifications. We filtered variants and samples according to the eQTLGen DataQC pipeline. We used a loss-of-function (LOF) S-threshold of 0.25 and a maximum SD of 3. This resulted in 751 samples with genotype data on 451,967 variants. 723 of these samples also had expression data.

#### Data availability statement

The BIOS RNA data can be obtained from the European Genome-Phenome Archive (EGA) (accession EGAS00001001077). Data-access procedures established for the BIOS Consortium are available at: https://directory.bbmri-eric.eu/ERIC/directory/#/biobank/bbmri-eric:ID:NL_aaaaczxoioaeoacqk2mo6qaaae

#### Acknowledgements

The Rotterdam Study is supported by the Erasmus MC University Medical Center and Erasmus University Rotterdam; the Netherlands Organization for Scientific Research (NWO); the Netherlands Organization for Health Research and Development (ZonMw); the Research Institute for Diseases in the Elderly (RIDE); the Netherlands Genomics Initiative (NGI); the Ministry of Education, Culture and Science; the Ministry of Health, Welfare and Sports; the European Commission (DG XII); and the Municipality of Rotterdam. The contribution of inhabitants, general practitioners, and pharmacists of the Ommoord district to the Rotterdam Study is gratefully acknowledged.

### PAN

#### Cohort description

PAN is a prospective study for patients with amyotrophic lateral sclerosis (ALS)[1]. Since 2006, PAN aims to include all Dutch patients with ALS and similar phenotypes to correlate potential lifestyle, genetic and environmental risk factors with the onset and prognosis of ALS (https://www.als-centrum.nl/kennisplatform/prospectieve-als-studie-nederland-pan/). To date, 3,400 patients have been included, and genotypes and expression data have been generated for a subset of these patients.

#### Ethics approval

All individuals gave written informed consent, and the University Medical Center Utrecht medical ethics committee approved this protocol.

#### Methods: expression data

RNA-seq gene expression data was generated in the HugeF (Erasmus MC, Rotterdam, the Netherlands, https://genomicserasmusmc.nl/). RNA-seq extraction and processing has been described before for a subset of the data[2]. Briefly, RNA was extracted from whole blood and paired-end sequenced using Illumina HiSeq 2000. Reads were aligned using STAR 2.3.0e[3] while masking common (MAF > 0.01) SNPs from GoNL[4]. Gene-level expression was quantified using HTseq[5]. FastQC (http://www.bioinformatics.babraham.ac.uk/projects/fastqc/) was used to check quality metrics, and we removed individuals with < 70% of reads mapping to exons (exon mapped/genome). From the 3,998 BIOS samples for which expression data was available, 164 samples also had filtered, quality-controlled genotype data available in the PAN cohort. As per the eQTLGen Data QC pipeline, we further filtered this set of overlapping samples by removing outliers (maximum SD = 3), likely contaminated samples (angle of contamination area = 30°), and likely swapped samples (slope of the line to discriminate between male and female sex expression = 45°). These QC steps resulted in 163 samples with expression data on 20,162 genes.

#### Methods: genotyping data

To obtain genome-wide genotype data, genomic DNA of patients and controls was hybridized to the Illumina OmniExpress array (Illumina, San Diego, CA, USA). We filtered variants and samples according to the eQTLGen DataQC pipeline. We used a LOF S-threshold of 0.75 and a maximum SD of 3. This resulted in 180 samples with genotype data on 563,539 variants. 164 of these samples also had expression data.

#### Data availability statement

The BIOS RNA data can be obtained from the EGA (accession EGAS00001001077). Data-access procedures established for the BIOS Consortium are available at: https://directory.bbmri-eric.eu/ERIC/directory/#/biobank/bbmri-eric:ID:NL_aaaaczxoioaeoacqk2mo6qaaae

### NTR

#### Cohort description

The Netherlands Twin Register (NTR) was set up in 1987 (https://tweelingenregister.vu.nl) to recruit Dutch mono- and dizygotic twins and their families. The NTR investigates health and lifestyle[1]. Twins and their relatives complete questionnaires and provide clinical measurements. From 2004 onwards, a subset of participants were asked to donate blood in order to create a biobank. Blood samples were used for genotyping, DNA, and RNA isolation and biomarker studies[2,3]. A subset of twins are also part of the BIOS Consortium, and we selected one individual from each twin pair for our study.

#### Ethics approval

The study protocol was approved by Central Ethics Committee on Research Involving Human Subjects of the VU University Medical Center, Amsterdam, an IRB certified by the U.S. Office of Human Research Protections (IRB number IRB-2991 under Federal-wide Assurance-3703; IRB/institute codes, NTR 03-180), and informed consent was obtained from all participants.

#### Methods: expression data

RNA-seq gene expression data was generated in the HugeF (Erasmus MC, Rotterdam, the Netherlands, https://genomicserasmusmc.nl/). RNA-seq extraction and processing has been described before for a subset of the data[4]. Briefly, RNA was extracted from whole blood and paired-end sequenced using Illumina HiSeq 2000. Reads were aligned using STAR 2.3.0e[5] while masking common (MAF > 0.01) SNPs from GoNL[6]. Gene-level expression was quantified using HTSeq[7]. FastQC (http://www.bioinformatics.babraham.ac.uk/projects/fastqc/) was used to check quality metrics, and we removed individuals with < 70% of reads mapping to exons (exon mapped/genome). From the 3,998 BIOS samples for which expression data was available, 131 also had filtered, quality-controlled genotype data available from the Illumina HiSeq 2000 subcohort of NTR. As per the eQTLGen Data QC pipeline, we further filtered this set of overlapping samples by removing outliers (maximum SD = 3), likely contaminated samples (angle of contamination area = 30°), and likely swapped samples (slope of the line to discriminate between male and female sex expression = 45°). These QC steps resulted in 120 samples with expression data on 21,729 genes.
614 BIOS RNA-seq samples also had filtered, quality-controlled genotype data available from the Illumina Affy6 genotyping array in the NTR cohort. For these samples, we performed the same expression data processing. These QC steps resulted in 605 samples with expression data on 19,726 genes.

#### Methods: genotyping data

NTR samples used in eQTLGen were genotyped using both the Affymetrix 6.0 array[1]and an Illumina HiSeq 2000 sequencing instrument as part of the GoNL initiative[6]. We filtered variants and samples according to the eQTLGen DataQC pipeline. For the samples genotyped using the Affymetrix 6.0 array, we used a LOF S-threshold of 0.4 and a maximum SD of 3. This resulted in 1,690 samples with genotype data on 599,850 variants. 614 of these samples also had expression data. For the samples genotyped using the Illumina HiSeq 2000 sequencing machine, we used a LOF S-threshold of 0.4 and a maximum SD of 3. This resulted in 292 samples with genotype data on 9,110,567 variants. 131 of these samples also had expression data available.

#### Data availability statement

NTR data can be requested through the following webpage: https://ntr-data-request.psy.vu.nl/. The BIOS RNA data can be obtained from the EGA (accession EGAS00001001077). Data-access procedures established for the BIOS Consortium are available at: https://directory.bbmri-eric.eu/ERIC/directory/#/biobank/bbmri-eric:ID:NL_aaaaczxoioaeoacqk2mo6qaaae

#### Acknowledgements

We are extremely grateful to the twin families who take part in NTR and to the study team.

#### Funding information

We acknowledge support from BBMRI-NL (Biobanking and Biomolecular Resources Research Infrastructure 184.021.007 and 184.033.111), a Spinoza prize (NWO- 56-464-14192), the European Research Council (ERC Advanced 230374), a KNAW Academy Professor Award (PAH/6635) to DIB, the National Institutes of Health (NIH, Grand Opportunity grants 1RC2 MH089951, and 1RC2 MH089995), the Avera Institute for Human Genetics, Sioux Falls, South Dakota (USA), and NWO-Groot 480-15-001/674: Netherlands Twin Registry Repository.

### Leiden Longevity Study

#### Cohort description

The Leiden Longevity Study (LLS) is a study started to research the mechanisms that contribute to healthy aging and longevity. It consists of 421 long-lived families made up of 944 90-year-old brother-sister pairs, 1,671 of their offspring, and 744 offspring partners. The partners form the control group and represent the general population[1].

#### Ethics approval

The Medical Ethical Committee of the Leiden University Medical Centre approved the study, and informed consent was obtained from all subjects.

#### Methods: expression data

RNA-seq gene expression data was generated in the HugeF (Erasmus MC, Rotterdam, the Netherlands, https://genomicserasmusmc.nl/). RNA-seq extraction and processing has been described before for a subset of the data[2]. Briefly, RNA was extracted from whole blood and paired-end sequenced using Illumina HiSeq 2000. Reads were aligned using STAR 2.3.0e[3] while masking common (MAF > 0.01) SNPs from GoNL[4]. Gene-level expression was quantified using HTSeq[5]. FastQC (http://www.bioinformatics.babraham.ac.uk/projects/fastqc/) was used to check quality metrics, and we removed individuals with < 70% of reads mapping to exons (exon mapped/genome). From the 3,998 BIOS samples for which expression data was available, 248 samples also had filtered Illumina660W-Quad genotype data available after processing in the LLS cohort. As per the eQTLGen Data QC pipeline, we further filtered this set of overlapping samples by removing outliers (maximum SD of 3), likely contaminated samples (angle of contamination area = 30°), and likely swapped samples (slope of the line to discriminate between male and female sex expression = 45°). These QC steps resulted in 355 samples with expression data on 19,823 genes.
248 BIOS RNA-seq samples also had filtered genotype data available after processing from the LLS samples genotyped using the OmniExpress BeadChip array. For these samples, we performed the same expression data processing. These QC steps resulted in 244 samples with expression data on 20,324 genes.

#### Methods: genotyping data

DNA from the LLS was extracted from white blood cells at baseline using conventional methods[6], and genotyping was performed with Illumina Human660W-Quad and OmniExpress BeadChips. This is further described by Deelen et al.[7]. We filtered variants and samples according to the eQTLGen DataQC pipeline. For the samples genotyped using the Illumina Human660W-Quad array, we used a LOF S-threshold of 0.25 and a maximum SD of 3. This resulted in 408 samples with genotype data on 500,411 variants. For the samples genotyped using the OmniExpress BeadChip array, we used a LOF S-threshold of 0.4 and a maximum SD of 3. This resulted in 285 samples with genotype data on 601,855 variants.

#### Data availability statement

Access to the data of the LLS can be requested through the cohort's data-access webpage: https://leidenlangleven.nl/data-access/. BIOS RNA data can be obtained from the EGA (accession EGAS00001001077). Data-access procedures established for the BIOS Consortium are available at: https://directory.bbmri-eric.eu/ERIC/directory/#/biobank/bbmri-eric:ID:NL_aaaaczxoioaeoacqk2mo6qaaae

#### Acknowledgements

We thank all participants of the LLS for their consistent cooperation, as well all participating general practitioners and pharmacists, the secretarial staff (Meriam H. van der Star, Ellen H. Bemer-Oorschot), research nurses (Corrie J. Groenendijk), and data managers (Karin H. Herbschleb) for their expert contributions. We also thank Karin H. Herbschleb for her contribution to the data analysis.

#### Funding information

The LLS has received funding from the European Union's Seventh Framework Programme (FP7/2007-2011) under grant agreement no 259679. The LLS was funded by the Innovation Oriented Research Program on Genomics (SenterNovem; IGE01014 and IGE5007), the Centre for Medical Systems Biology, the NGI/NWO (05040202 and 050-060-810), and the European Union–funded Network of Excellence Lifespan (FP6 036894).

### LifeLines DEEP

#### Cohort description

LifeLines is a population-based longitudinal cohort study that includes questionnaire-based and clinical data of 167,729 individuals living in the three Northernmost provinces of the Netherlands[1]. The study specifically focuses on families and employs a three-generational design. LifeLines DEEP is a subset of 1,500 unrelated Lifelines participants who consented to further investigation of their genetics, gene expression, methylation, gut microbiome, and exhaled breath metabolomics[2].

#### Ethics approval

The LifeLines DEEP study was approved by the ethics committee of the University Medical Center Groningen. All participants signed an informed consent prior to enrollment.

#### Methods: expression data

RNA-seq gene expression data was generated in the HugeF (Erasmus MC, Rotterdam, the Netherlands, https://genomicserasmusmc.nl/). RNA-seq extraction and processing has been described before for a subset of the data[3] (Zhernakova et al., 2017). Briefly, RNA was extracted from whole blood and paired-end sequenced using Illumina HiSeq 2000. Reads were aligned using STAR 2.3.0e[4] while masking common (MAF > 0.01) SNPs from GoNL[5]. Gene-level expression was quantified using HTSeq[6]. FastQC (http://www.bioinformatics.babraham.ac.uk/projects/fastqc/) was used to check quality metrics, and we removed individuals with < 70% of reads mapping to exons (exon mapped/genome).
From the 3,998 BIOS samples for which expression data was available, 1,056 samples also had filtered genotype data available after processing in the LifeLines DEEP cohort. As per the eQTLGen Data QC pipeline, we further filtered this set of overlapping samples by removing outliers (maximum SD of 3), likely contaminated samples (angle of contamination area = 30°) and likely swapped samples (slope of the line to discriminate between male and female sex expression = 45°). These QC steps resulted in 1,029 samples with expression data on 19,765 genes.

#### Methods: genotyping data

Genotyping of genomic DNA was performed using both the HumanCytoSNP-12 BeadChip and the ImmunoChip, a customized Illumina Infinium array. Genotyping was successful for 1,385 samples (CytoSNP) and 1,374 samples (IChip), respectively. First, SNP QC was applied independently for both platforms. Using PLINK, SNPs were filtered on MAF > 0.001, a Hardy-Weinberg equilibrium (HWE) P-value > $1x10^{-4}$, and call rate of 0.98. The genotypes from both platforms were merged into one data set. For genotypes present on both platforms, the genotypes were put on missing in the case of non-concordant calls. After merging, SNPs were filtered again on MAF 0.05 and call rate of 0.98, resulting in 379,885 genotyped SNPs. We further filtered variants and samples according to the eQTLGen DataQC pipeline, using a LOF S-threshold of 0.32 and a maximum SD of 3. This resulted in 1,347 samples with genotype data on 229,835 variants.

#### Data availability statement

The BIOS RNA data can be obtained from the EGA (accession EGAS00001001077). Data-access procedures established for the BIOS Consortium are available at: https://directory.bbmri-eric.eu/ERIC/directory/#/biobank/bbmri-eric:ID:NL_aaaaczxoioaeoacqk2mo6qaaae

#### Acknowledgements

The authors would like to thank the Lifelines participants and the staff of the Lifelines study site, Groningen, for their collaboration. The authors would also like to thank the LifeLines DEEP research assistants, Wilma Westerhuis-van der Tuuk, Marc Jan Bonder, Astrid Maatman, Mathieu Platteel, Kim de Lange, and Debbie van Dussen for their practical and analytical work.

#### Funding information

The LifeLines DEEP project was funded by a Top Institute Food and Nutrition Wageningen grant GH001 to CW, a Biobanking and Biomolecular Research Infrastructure Netherlands (BBMRI-NL) grant RP3 to LF, and an ERC advanced grant ERC-671274 to CW. SZ holds a Rosalind Franklin fellowship (University of Groningen). MCC has a postdoctoral fellowship from the Spanish Fundación Alfonso Martín Escudero.

### CODAM

#### Cohort description

The Cohort on Diabetes and Atherosclerosis Maastricht (CODAM) is group of individuals with a slightly increased risk of cardiometabolic disease selected from a population-based cohort[1]. Individuals in CODAM are of European descent and older than 40 years of age. They have either an increased BMI (>25), a family history of type 2 diabetes, previous gestational diabetes and/or glycosuria, or use medication to treat hypertension.

#### Ethics approval

The study was approved by the Medical Ethical Committee of the Maastricht University, and all subjects gave written informed consent.

#### Methods: expression data

RNA-seq gene expression data was generated in the HugeF (Erasmus MC, Rotterdam, the Netherlands, https://genomicserasmusmc.nl/). RNA-seq extraction and processing has been described before for a subset of the data[2]. Briefly, RNA was extracted from whole blood and paired-end sequenced using Illumina HiSeq 2000. Reads were aligned using STAR 2.3.0e[3] while masking common (MAF > 0.01) SNPs from GoNL[4]. Gene-level expression was quantified using HTSeq[5]. FastQC (http://www.bioinformatics.babraham.ac.uk/projects/fastqc/) was used to check quality metrics, and we removed individuals with < 70% of reads mapping to exons (exon mapped/genome). From the 3,998 BIOS samples for which expression data was available, 179 samples also had filtered, quality-controlled genotype data available in the CODAM cohort. As per the eQTLGen Data QC pipeline, we further filtered this set of overlapping samples by removing outliers (maximum SD of 3), likely contaminated samples (angle of contamination area = 30°), and likely swapped samples (slope of the line to discriminate between male and female sex expression = 45°). These QC steps resulted in 177 samples with expression data on 20,162 genes.

#### Data availability statement

The BIOS RNA data can be obtained from the EGA (accession EGAS00001001077). Data-access procedures established for the BIOS Consortium are available at: https://directory.bbmri-eric.eu/ERIC/directory/#/biobank/bbmri-eric:ID:NL_aaaaczxoioaeoacqk2mo6qaaae. Genotype data are available upon reasonable request.

### Fehrmann

#### Cohort description

The Fehrmann datasets consist of whole-blood samples from the United Kingdom and the Netherlands[1,2]. This dataset consists of blood samples from patients[2] and healthy controls.

#### Ethics approval

All samples were collected after informed consent and approval by local ethical review boards.

#### Methods: expression data

Gene expression levels were measured by Illumina HT-12 v3 and Illumina HumanRef-8 v2.0 arrays. For the samples for which expression was measured using the Illumina HT-12 v3 array, the expression data was processed using the eQTLGen Data QC pipeline. We filtered these samples by removing outliers (maximum SD of 4), likely contaminated samples (angle of contamination area = 30°), and likely swapped samples (slope of the line to discriminate between male and female sex expression = 45°). These QC steps resulted in 206 samples with expression data on 9,059 genes. For the samples for which expression was measured using the HumanRef-8 v2.0 array, expression data was processed with the same settings. These QC steps resulted in 1,071 samples with expression data on 13,285 genes.

#### Methods: genotyping data

Samples were genotyped with Illumina HumanHap300, HumanHap370, or 610 Quad platforms. We filtered variants and samples according to the eQTLGen DataQC pipeline. For the samples for which expression was measured with the Illumina HT-12 v3, we used a LOF S-threshold of 0.4 and a maximum SD of 3. This resulted in 1,208 samples with genotype data on 286,384 variants. 1,206 of these samples also had expression data. For the HumanRef-8 v2.0 samples, we used a LOF S-threshold of 0.4 and a maximum SD of 3. This resulted in 2,204 samples with genotype data on 288,646 variants. 2,037 of these samples also had expression data available.

#### Data availability statement

The expression dataset is available at the Gene Expression Omnibus (GEO) repository (accession numbers GSE20142 and GSE20332).

#### Funding information

Genotype and gene expression generation was funded in part by COPACETIC (EU grant 201379), the Wellcome Trust (084743 to DAvH), and the Coeliac Disease Consortium, an Innovative Cluster approved by the NGI, as well as partially funded by the Dutch Government (BSIK03009 to CW) and the NWO (VICI grant 918.66.620 to CW).

### 300TZFG

#### Cohort description

The Tanzanian 300TFZ cohort included healthy individuals between 18 and 65 years old residing in the Kilimanjaro region of Northern Tanzania. A total of 383 individuals from both rural and urban areas were screened, resulting in the enrollment of 323 participants. All participants tested negative for malaria and HIV and had no signs of acute or chronic illness. Exclusion criteria comprised pregnancy, antibiotic or anti-malarial use within the past three months, a history of tuberculosis in the previous year, blood pressure readings above 140/90 mmHg or below 90/60 mmHg, and random blood glucose levels exceeding 8.0 mmol/L.[1]

#### Ethics approval

The 300TZFG study was approved by the Ethical Committees of the Kilimanjaro Christian Medical University College (CREC) (no. 2443) and the National Institute for Medical Research (NIMR/HQ/R.8a/Vol. IX/2290 and NIMR/HQ/R.8a/Vol.IX/3318) in Tanzania. Written informed consent was obtained from all subjects

#### Methods: expression data

Whole blood was collected in Qiagen PAXgene Blood RNA tubes. Total RNA was extracted using the Qiagen PAXgene RNA Isolation Kit, following the manufacturer's instructions, and eluted in RNase-free water. RNA quality was assessed using the Agilent TapeStation 4200 by evaluating the integrity of the 28S and 18S rRNA bands. cDNA libraries were prepared from total RNA using the TruSeq Stranded Total RNA with Ribo-Zero Globin kit (Illumina). High-throughput sequencing was performed on the Illumina NovaSeq 6000 system using an S4 flow cell with v1.5 chemistry, generating 76 bp paired-end reads. Base calling and demultiplexing were conducted using bcl2fastq2 v2.20 to produce FASTQ files. Raw RNA-seq reads were aligned to the human reference transcriptome (GRCh38/hg38) using STAR aligner (v2.7.3a) to construct the gene counts table. Gene annotation was based on GENCODE v33.

#### Methods: genotyping data

DNA was extracted from whole blood with the DNeasy kit. Genotyping was performed with the Global Screening Array (GSA) SNP chip. We used default settings of Opticall 0.70 to perform genotype calling.[2] Genotype data was further automatically quality-controlled, imputed, and processed by eQTLGen pipelines.

#### Data availability statement

Sequence data have been submitted to the EGA, hosted by the EBI and CRG, under accession number EGAS00001004284. Additional datasets can be accessed through the Human Functional Genomics website upon reasonable request, subject to privacy safeguards for research participants. Requests can be made using the data request form available at: http://www.humanfunctionalgenomics.org/site/?page_id=16.

#### Acknowledgements

300 Tanzania Functioning Genomics Team (300TZFG), Tanzania

Godfrey S. Temba[1],Vesla I. Kullaya[1], Tal Pecht[2],Blandina T. Mmbaga[1], Reginald Kavishe[1], Leo A.B. Joosten[3], Mihai G. Netea[3], Quirijn de Mast[3]

[1]Department of Medical Biochemistry and Molecular Biology, Kilimanjaro Christian Medical University College, (KCMUCo) Moshi, Tanzania
[2]Department for Genomics and Immunoregulation, Life & Medical Sciences (LIMES) Institute, University of Bonn, Bonn, Germany
[3]Department of Internal Medicine, Radboud University Medical Center, Nijmegen, the Netherlands

#### Funding information

This work received funding from the following sources: the European Union's Horizon 2020 Research and Innovation Programme through the ERA-Net Cofund action (grant no. 727565) and the Joint Programming Initiative "A Healthy Diet for a Healthy Life" (JPI-HDHL; project no. 529051018).

### GTEx

#### Cohort description

The Adult Genotype-Tissue Expression (GTEx) Project is a comprehensive public resource to study human gene expression and regulation and its relationship to genetic variation across multiple diverse tissues and individuals. GTEx donor recruitment and molecular data generation are complete for core molecular assays including whole-genome sequencing (WGS), whole-exome sequencing, and RNA-seq[1,2,3].

#### Ethics approval

The GTEx project involves potentially sensitive recruitment, IRB, and consent issues, particularly for deceased donors and their families. The collection of biospecimens from deceased individuals is not legally classified as human subjects research under 45 CFR 46; nonetheless, the depth of the genetic information obtained from the specimens of deceased donors has direct implications for the families of the donors. In recognition of this understanding, sites were required to obtain written or recorded verbal authorization from next of kin for the participation of deceased donors in GTEx, typically through an addendum or modification to an existing authorization form for donation of tissues and organs for research. This authorization included statements common in consent forms, such as the intention to perform genetic analyses, establish cell lines, and share data with the scientific community. Work under way is more closely identifying familial concerns and may result in modifications to authorization procedures. Living surgery donors participate only after full, written informed consent is obtained.

#### Methods: expression data

Expression data was measured and quantified as outlined in The GTEx Consortium (2020)[3] and on the GTEx portal website (https://www.gtexportal.org/home/methods). In short, RNA-seq was performed using the Illumina TruSeq library construction protocol. Alignment to the human reference genome GRCh38/hg38 was performed using STAR v2.5.3a, based on the GENCODE v26 annotation. Of the 756 samples for which expression data was available, 568 samples also had filtered genotype data available after processing. In line with the eQTLGen Data QC pipeline, we further filtered this set of overlapping samples by removing outliers (maximum SD of 4), likely contaminated samples (angle of contamination area = 30°), and likely swapped samples (slope of the line to discriminate between male and female sex expression = 45°). These QC steps resulted in 564 samples with expression data on 20,986 genes.

#### Methods: genotyping data

We used GTEx V9 genotype data for eQTL mapping. We removed genotypes with a genotype quality < 20, an allelic balance < 0.2, an inbreeding coefficient < -0.3, and variant quality score recalibration values < 99.8 for single nucleotide variants (SNVs) and < 99.5 for indels. We further removed multi-allelic variants and variants without the PASS label, with a read-depth < 10, with a MAF < 0.01, and that did not follow HWE (P-value threshold of $1 \times 10^{-6}$). This left 8,495,158 variants and 953 samples for further QC and sample filtering.
The proportion of ancestries within the set of 953 samples was such that no ancestry other than European consisted of more than 100 samples. Therefore, we confined the GTEx dataset to European samples only. We further filtered variants and samples according to the eQTLGen DataQC pipeline, using a LOF S-threshold of 0.5 and a maximum SD of 3. This resulted in 720 samples with genotype data on 6,870,734 variants.

#### Data availability statement

The datasets used for the analyses described in this manuscript were obtained from dbGaP at http://www.ncbi.nlm.nih.gov/gap through dbGaP accession number phs000424.v10.p2.

#### Acknowledgements

The GTEx project was supported by the Common Fund of the Office of the Director of the NIH (commonfund.nih.gov/GTEx). Additional funds were provided by the National Cancer Institute (NCI), National Human Genome Research Institute (NHGRI), National Heart Lung and Blood Institute (NHLBI), National Institute on Drug Abuse (NIDA), National Institute of Mental Health (NIMH), and National Institute of Neurological Disorders and Stroke (NINDS). Donors were enrolled at Biospecimen Source Sites funded by NCI\Leidos Biomedical Research, Inc. subcontracts to the National Disease Research Interchange (10XS170), Roswell Park Cancer Institute (10XS171), and Science Care, Inc. (X10S172). The Laboratory, Data Analysis, and Coordinating Center was funded through a contract (HHSN268201000029C) to The Broad Institute, Inc. Biorepository operations were funded through a Leidos Biomedical Research, Inc. subcontract to Van Andel Research Institute (10ST1035). Additional data repository and project management were provided by Leidos Biomedical Research, Inc. (HHSN261200800001E). The Brain Bank was supported by supplements to University of Miami grant DA006227. Statistical Methods development grants were made to the University of Geneva (MH090941 and MH101814), University of Chicago (MH090951,MH090937, MH101825, and MH101820), University of North Carolina - Chapel Hill (MH090936), North Carolina State University (MH101819), Harvard University (MH090948), Stanford University (MH101782), Washington University (MH101810), and University of Pennsylvania (MH101822).

#### Funding information

The GTEx project was supported by the Common Fund of the Office of the Director of the NIH , and by the NCI , the NHGRI , the National Heart, Lung, and Blood Institute , the NIDA , the NIMH , the NINDS .

### YFS

#### Cohort description

The Cardiovascular Risk in Young Finns Study (YFS) is a population-based, prospective multi-center cohort study being conducted in five university hospital cities in Finland[1]. Of the 3,596 individuals participating at baseline in 1980, 2,050 took part in the YFS follow-up examinations in 2011−2012. As described previously[2], 2,049 gave blood samples for RNA isolation.

#### Ethics approval

The study was approved by the ethical committee of the Hospital District of Southwest Finland on 20 June 2017 (ETMK:68/1801/2017), and all participants have given an informed written consent. Data protection will be handled according to current regulations.

#### Methods: expression data

The whole-genome blood transcriptome was profiled from RNA isolated from whole-blood samples collected from study participants during the 2011 follow-up. Expression levels were analyzed with Illumina HumanHT-12 version 4 Expression BeadChip (Illumina Inc.), containing 47,231 expression and 770 control probes. Samples with fewer than 6,000 significantly detected expression probes (detection P-value < 0.01) were discarded. Raw Illumina summary probe-level data was exported from BeadStudio and processed in R (http://www.r-project.org/) using a nonparametric background correction, followed by quantile normalization with control and expression probes, with the neqc function in the limma package and a log2 transformation. Nine samples were excluded due to sex mismatch between the recorded sex and predicted sex based on RPS4Y1-2 and XIST mRNA levels on the Y and X chromosomes, respectively. After QC, expression data was available for 1,654 samples, including four technical replicates that were used to examine batch effects and subsequently excluded before further analysis.

#### Methods: genotyping data

Genomic DNA was extracted from peripheral blood leukocytes using a commercially available kit and a Qiagen BioRobot M48 Workstation according to the manufacturer's instructions (Qiagen, Hilden, Germany). Genotyping was performed using a custom-built Illumina Human 670 k BeadChip at the Welcome Trust Sanger Institute. Genotypes were called using the Illuminus clustering algorithm. Samples that failed the Sanger genotyping pipeline QC criteria (i.e., duplicated samples, heterozygosity, low call rate, or Sequenom fingerprint discrepancy) were excluded. Similarly, samples with sex discrepancy, a low genotyping call rate (< 0.95), and possible relatedness (pi-hat > 0.2) were excluded. SNPs were filtered on HWE ($P \leq 1x10^{-06}$) and a missingness test (call rate < 0.95). After QC, 2,443 samples and 546,674 SNPs were available for further analysis.

#### Data availability statement

The dataset was obtained from the YFS, which comprises health-related participant data. The use of data is restricted under the regulations on professional secrecy (Act on the Openness of Government Activities, 612/1999) and on sensitive personal data (Personal Data Act, 523/1999, implementing the EU data protection directive 95/46/EC). Due to these restrictions, the data cannot be stored in public repositories or otherwise made publicly available. Data-access may be permitted on a case-by-case basis upon request only. Data-sharing outside the group is done in collaboration with YFS group and requires a data-sharing agreement. Investigators can submit an expression of interest to the chairman of the YFS publication committee Prof. Mika Kähönen (, Tampere University,

Tampere, Finland), coordinator of YFS Prof. Olli T. Raitakari (, University of Turku, Turku, Finland), and responsible investigator of the YFS genetic section Prof. Terho Lehtimäki (, Tampere University, Tampere, Finland).


#### Acknowledgements

We thank the teams that collected data at all measurement time points, the participants who participated in these longitudinal studies as both children and adults, and the biostatisticians Irina Lisinen, Johanna Ikonen, Noora Kartiosuo, Ville Aalto, and Jarno Kankaanranta for data management and statistical advice.

#### Funding information

The YFS has been financially supported by the Academy of Finland: grants 356405, 322098, 286284, 134309 (Eye), 126925, 121584, 124282, 129378 (Salve), 117797 (Gendi), 141071 (Skidi), 349708, 330809, and 338395; the Social Insurance Institution of Finland; Competitive State Research Financing of the Expert Responsibility area of Kuopio and Turku; State funding for university-level health research, Tampere University Hospital (grant X51001); the Juho Vainio Foundation; the Paavo Nurmi Foundation; the Finnish Foundation for Cardiovascular Research; the Finnish Cultural Foundation; the Sigrid Jusélius Foundation; the Tampere Tuberculosis Foundation; the Emil Aaltonen Foundation; the Yrjö Jahnsson Foundation; the Signe and Ane Gyllenberg Foundation; the Diabetes Research Foundation of Finnish Diabetes Association; EU Horizon 2020 (grant 755320 for TAXINOMISIS and grant 848146 for ToAition); the European Research Council (grant 742927 for MULTIEPIGEN project); Tampere University Hospital Supporting Foundation, Finnish Society of Clinical Chemistry, the Cancer Foundation Finland; BETTER4U (EU grant 101080117); CVDLink (EU grant 101137278), and the Jane and Aatos Erkko Foundation.

### STRIP

#### Cohort description

The main purpose of the Special Turku Coronary Risk Factor Intervention Project (STRIP) study is the prevention of atherosclerosis and coronary heart disease through a dietary intervention that began in infancy and has continued to early adulthood. The trial was launched in 1990, when 1,062 7-month-old children and their families were enrolled. Half of the families have received individualized dietary and other lifestyle counseling at least twice a year. The rest of the families have served as a control group. The STRIP study intervention continued until participants reached age 20 years. The first post-intervention follow-up study was conducted six years after the active intervention, at age 26 years[1,2].

#### Ethics approval

First ethics approval was done in 1989 (decision date 7.11.1989) by the joint Committee of Ethics of the University of Turku and the University Central Hospital of Turku and the Ethics Committee, Hospital District of Southwest Finland. The STRIP 26-year follow-up study was approved by the Ethics Committee of Hospital District of Southwest Finland, decision ETMK:51/1801/2014. Written informed consent was received from the participants' parents in the beginning of the study. Later, at ages 15, 18 and 26 years, the participants gave their own informed consent.

#### Methods: expression data

Whole-blood RNA samples (N = 539) were collected in PAXgene Blood RNA tubes (Qiagen) during the 26-year follow-up study. Total RNA was isolated manually with MagMAX™ for Stabilized Blood Tubes RNA Isolation Kit (Thermo Fisher Scientific) according to manufacturer's instructions and eluted in RNase-free water. The quality of the RNA was ensured using Bioanalyzer 2100 or Advanced Analytical Fragment Analyzer (Agilent). RNA concentration was measured with Nanodrop ND-2000 (Thermo Fisher Scientific) and/or Qubit®/Quant-IT® Fluorometric Quantitation (Life Technologies). One sample had low RNA quality and was discarded from the sequencing steps. Sequencing libraries were prepared from total RNA following the TruSeq® Stranded mRNA RNA Sample Preparation protocol (Illumina). The quality of the sample libraries were ensured using Advanced Analytical Fragment Analyzer. Library concentration was measured with Qubit®/Quant-IT® Fluorometric Quantitation. Next-generation 2x100 bp RNA-sequencing was performed for 538 samples in three runs using Illumina NovaSeq 6000 instrument using S4 flow cell with v1.5 chemistry. Some of the samples (17) were sequenced twice to get at least 20 million reads per sample. The raw RNA-seq data for each run was preprocessed with bcl2fastq2 conversion software (Illumina) including automatic adapter trimming. The quality of the RNA-seq reads was assessed with the FastQC and MultiQC tools. RNA-seq reads were aligned to the human reference genome GRCh38 using the Subread aligner (Rsubread v.2.12.2). Gene-level read counts were obtained by running featureCounts, a read count summarization program within the Rsubread package and the inbuilt Rsubread annotation hg38. Expression data and genotype data were available for 515 samples. Expression levels were available for 28,395 genes.

#### Methods: genotyping data

Genomic DNA was extracted from peripheral blood leukocytes using QIAamp DNA Blood Mini kit and an automated biorobot M48 extraction (Qiagen, Hilden, Germany). Genotyping was performed using Illumina Infinium™ GSA-24 DNA Analysis Beadchip version 3.0, according to the manufacturer's recommendation, at Helmholtz Zentrum, München, Germany. The following QC filters were applied: GenCall score < 0.15, GenTrain score < 0.20, sample and SNP call rate < 0.95, HWE P-value <

10x10−6, excess heterozygosity, cryptic relatedness (pi-hat > 0.2), sex check, and multidimensional scaling. After QC, 780 samples and 640,640 SNPs were available for further analysis.

#### Data availability statement

The use of data is restricted under the regulations on professional secrecy (Act on the Openness of Government Activities, 612/1999) and on sensitive personal data (Personal Data Act, 523/1999, implementing the EU data protection directive 95/46/EC). Due to these restrictions, the data cannot be stored in public repositories or otherwise made publicly available. Selected variables and their descriptions without personal identification codes are distributed to investigators and collaborators working on specific projects. The rights to the data belong to the STRIP research group. Data-sharing outside the STRIP group requires a data-sharing agreement. Investigators can submit an expression of interest to the STRIP Steering Committee (https://stripstudy.utu.fi/en/strip-study/).

#### Acknowledgements

We thank the study participants and their families, as well as the research group who collected the data.

#### Funding information

STRIP has been supported by the Academy of Finland (grants 206374, 294834, 251360, 275595, 307996, 322112, and 357183), the Juho Vainio Foundation, the Finnish Foundation for Cardiovascular Research, the Finnish Ministry of Education and Culture, the Finnish Cultural Foundation, the Sigrid Jusélius Foundation, Special Governmental grants for Health Sciences Research (Turku University Hospital), the Yrjö Jahnsson Foundation, the Finnish Medical Foundation, and the Turku University Foundation. This study was supported by the Finnish Functional Genomics Centre, University of Turku and Åbo Akademi and Biocenter Finland.

### SHIP-TREND

#### Cohort description

The Study of Health in Pomerania (SHIP) is a population-based survey consisting of three independent cohorts in the North-East of Germany: SHIP-START, SHIP-TREND, and SHIP-NEXT. The study design and sampling methods were described previously[1,2]. For this eQTL analysis, a subset of the SHIP-TREND cohort (N = 986) with gene expression levels measured was used.

#### Ethics approval

The study followed the recommendations of the Declaration of Helsinki. The medical ethics committee of the University of Greifswald approved the study protocol. Oral and written informed consents were obtained from each of the study participants.

#### Methods: expression data

Blood sample collection as well as RNA preparation are described in detail elsewhere[3]. Briefly, RNA was prepared from whole blood under fasting conditions in PAXgene tubes (BD) using the PAXgene Blood miRNA Kit (Qiagen, Hilden, Germany) on a QIAcube according to the protocols provided by the manufacturer (Qiagen). RNA was amplified (Ambion TotalPrep RNA) and hybridized to the Illumina whole-genome Expression BeadChips (HT-12v3).

#### Methods: genotyping data

Serum aliquots were prepared for immediate analysis and for storage at -80°C in the Integrated Research Biobank (Liconic, Liechtenstein). Genotyping was performed using the Illumina HumanOmni2.5 BeadChip. Genotype calling was done with GenomeStudio Genotyping Module v1.0 (GenCall). Samples with a call rate < 94%, reported vs. genotyped sex mismatch, and duplicate samples (by estimated Identity-By-Descent (IBD)) were excluded. Monomorphic SNVs, SNVs with a call rate ≤ 90%, and SNVs out of HWE (P ≤ 0.0001) were excluded.

#### Data availability statement

The genotype data of the SHIP study cannot be made publicly available due to the informed consent of the study participants, but it can be accessed through a data application form available at https://fvcm.med.uni-greifswald.de for researchers who meet the criteria for access to confidential data. The SHIP-TREND expression dataset is available at GEO public repository under the accession GSE 36382.

#### Funding information

SHIP is part of the Community Medicine Research net of the University of Greifswald, Germany, which is funded by the Federal Ministry of Education and Research (grants no. 01ZZ9603, 01ZZ0103, and 01ZZ0403), the Ministry of Cultural Affairs and the Social Ministry of the Federal State of Mecklenburg-West Pomerania, and the network ‘Greifswald Approach to Individualized Medicine (GANI_MED)’ funded by the Federal Ministry of Education and Research (grant 03IS2061A).

### Rotterdam Study (HT12v4)

#### Cohort description

The Rotterdam Study[1,2,3] is a single-center, prospective population-based cohort study conducted in Rotterdam, the Netherlands. Subjects were included in different phases from the start of the study in 1998, with a total of 14,926 men and women aged 45 years and over included as of late 2008. The main objective of the Rotterdam Study is to investigate the prevalence and incidence of risk factors for chronic diseases to contribute to better prevention and treatment of such diseases in the elderly.

#### Ethics approval

The Rotterdam Study has been approved by the IRB (medical ethics committee) of the Erasmus Medical Center and by the review board of The Netherlands Ministry of Health, Welfare and Sports.

#### Methods: expression data

Whole blood was collected in PAXgene tubes (Becton Dickinson), and total RNA was isolated using PAXgene Blood RNA kits (Qiagen). To ensure constant high quality of RNA preparation, all RNA samples were analyzed using the Labchip GX (Calliper) according to manufacturer's instructions. Samples with an RNA Quality Score > 7 were amplified and labeled (Ambion TotalPrep RNA) and hybridized to the Illumina HumanHT-12 v4 Expression BeadChips (Illumina) as described by the manufacturer's protocol. Processing of the Rotterdam Study RNA samples was performed at the Genetic Laboratory of Internal Medicine, Erasmus University Medical Center Rotterdam.
From 763 samples with expression data, 710 also had filtered quality-controlled genotype data available. As per the eQTLGen Data QC pipeline, we further filtered this set of overlapping samples by removing outliers (maximum SD of 4), likely contaminated samples (angle of contamination area = 15°), and likely swapped samples (slope of the line to discriminate between male and female sex expression = 25°). These QC steps resulted in 700 samples with expression data on 13,348 genes.

#### Methods: genotyping data

For genotyping, whole blood was also collected in EDTA tubes and DNA was isolated using a manual salting-out protocol. Genotyping for this sample subset was performed on the Illumina 610K quad beadchip array (Illumina) according to manufacturer's specifications. We filtered variants and samples according to the eQTLGen DataQC pipeline. We used a LOF S-threshold of 0.4 and a maximum SD of 3. This resulted in 710 samples with genotype data on 505,829 variants.

#### Data availability statement

Data can be obtained upon request. Requests should be directed towards the management team of the Rotterdam Study, which has a protocol for approving data requests. Because of restrictions based on privacy regulations and informed consent of the participants, data cannot be made freely available in a public repository.

#### Acknowledgements

The contribution of inhabitants, general practitioners, and pharmacists of the Ommoord district to the Rotterdam Study is gratefully acknowledged.

#### Funding

The Rotterdam Study is supported by the Erasmus MC University Medical Center and Erasmus University Rotterdam; NWO; ZonMw; the Research Institute for Diseases in the Elderly (RIDE); NGI; the Ministry of Education, Culture and Science; the Ministry of Health, Welfare and Sports; the European Commission (DG XII); and the Municipality of Rotterdam.

### PRECISEADS

#### Cohort description

PRECISEADS is a systemic autoimmune disease cohort including healthy controls and participants with seven pathological conditions: systemic lupus erythematosus, systemic sclerosis, primary Sjögren's syndrome, rheumatoid arthritis, primary antiphospholipid syndrome, mixed connective tissue disease, and undifferentiated connective tissue disease[1].

#### Ethics approval

The ethical review boards of the 18 participating institutions approved the protocol of the cross-sectional study. The studies adhered to the standards set by the International Conference on Harmonization and Good Clinical Practice (ICH-GCP) and to the ethical principles that have their origin in the Declaration of Helsinki (2013). The protection of the confidentiality of records that could identify the included subjects is ensured as defined by the EU Directive 2001/20/EC and the applicable national and international requirements relating to data protection in each participating country. The study is registered with the number NCT02890121 in ClinicalTrials.gov.

#### Methods: expression data

Total RNA was extracted from whole-blood samples collected in Tempus tubes using Tempus Spin technology (Applied Biosystems). Samples were processed in batches of 384, randomized to four 96-well plates with respect to patient diagnosis, recruitment center, and RNA extraction date. The samples were depleted of alpha- and beta-globin mRNAs using GLOBINclear protocol (Ambion) and 1 µg of total RNA as input. Subsequently, 400 ng of globin-depleted total RNA was used for library synthesis with TruSeq Stranded mRNA HT kit (Illumina). The libraries were quantified using qPCR with the PerfeCTa NGS kit (Quanta Biosciences), and equimolar amounts of samples from the same 96-well plate were pooled. Four pools were clustered on a high output flow cell (two lanes per pool) using HiSeq SR Cluster kit v4 and the cBot instrument (Illumina). Subsequently, 50 cycles of single-read sequencing were performed on a HiSeq2500 instrument using HiSeq SBS kit v4 (Illumina). The clustering and sequencing steps were repeated for a total of three runs to generate a sufficient number of reads per sample. The raw sequencing data for each run were preprocessed using bcl2fastq software, and the quality was assessed using FastQC tools. Cutadapt was used to remove 3' end nucleotides below the 20 Phred quality score and Illumina adapters. Additionally, reads below 25 nucleotides after trimming were discarded. Reads were then processed and aligned to the UCSC *Homo sapiens* reference genome (build hg19) using STAR v2.5.2b. 2-pass mapping with default alignment parameters. To produce the quantification data, we used RSEM v1.2.31, resulting in gene-level expression estimates.

#### Methods: genotyping data

DNA was extracted using a magnetic bead nucleic acid isolation protocol (Chemagic DNA Blood Kit special, CHEMAGEN) automated with the Chemagic Magnetic Separation Module I (PerkinElmer) from a 10 ml K2EDTA BLOOD TUBE (lavender cap, BD Vacutainer) (extractions were performed on 3 ml). 2 µg of DNA were normalized to 100 ng/µg and sent for genotype analysis. DNA was genotyped using the Illumina HumanCore-24 v1.0 and the Infinium CoreExome-24 v1.2 genome-wide SNP genotyping platform (Illumina). Genotypes from both versions were merged based on genetic variant identity.

#### Data availability statement

Data is hosted by ELIXIR Luxembourg. Data is available upon request, and the access procedure is described on the data landing page (doi.org/10.17881/th9v-xt85).


#### Acknowledgements

The authors would like to particularly express their gratitude to the patients, nurses, and many others who helped directly or indirectly in the conduct of this study. This work is supported by ELIXIR Luxembourg via its data hosting service.

#### Funding information

The research leading to these results has received support from the Innovative Medicines Initiative Joint Undertaking under the Grant Agreement Number 115565 (PRECISESADS project), resources of which are composed of financial contribution from the European Union's Seventh Framework Program (FP7/2007–2013) and EFPIA companies' in-kind contribution. G.B. is supported by "Ministerio de Ciencia, Innovación y Universidades" (Spanish Government) through the program "Juan de la Cierva-Incorporación" (IJC2020-043364-I).

### OphoffBP

#### Cohort description

The samples included are from a study with individuals ascertained for bipolar disorder. The cohort consists of 1,045 individuals with bipolar disorder and 601 controls, with whole-blood RNA-seq and corresponding genotypes included for all individuals[1]. All data preprocessing was conducted using eQTLGen pipelines.

#### Ethics approval

Data was generated according to the protocols approved by the respective local ethics committee: the Medical Ethical Review Board at the University Medical Center Utrecht and the IRB at the University of California, Los Angeles. Information consent was obtained from all subjects.

#### Methods: expression data

Whole peripheral blood RNA samples were collected at the UCLA Neurogenomics Core using the TruSeq Stranded plus rRNA and GlobinZero library preparation method. About 600 samples were sequenced to an average of 13.9 million reads per sample, while about 2,000 samples were sequenced to an average of 5.9 million mapped reads per sample. We used FastQC to visually inspect the read quality from the low-coverage whole-blood RNA-seq and the high-coverage whole-blood RNA-seq. We then used kallisto to pseudoalign reads to the GRCh37 gencode transcriptome (v.33) and quantify estimates for transcript expression. We aggregated transcript counts to obtain gene-level read counts using scripts from the GTEx consortium (https://github.com/broadinstitute/gtex-pipeline). Raw expression data was further processed automatically with eQTLGen pipelines.

#### Methods: genotyping data

The genotypes are derived from multiple project cohorts, each using different genotyping platforms including OmniExpressExome (OMEX), GSA, COEX, and Psych Chip (PSYCH). Genotypes for the low-coverage whole-blood RNA-seq were obtained from the following platforms: OMEX (N = 816), GSA (N = 211), COEX (N = 162), and PSYCH (N = 522). Genotypes for the high-coverage whole-blood RNA-seq were obtained from the OMEX platform. Genotype data was further automatically quality-controlled, imputed, and processed by eQTLGen pipelines.

#### Data availability statement

The low-coverage RNA-seq and the corresponding genotypes have been deposited in dbGAP (accession number phs002856.v1).

#### Acknowledgements

We thank the study subjects for their willingness to provide specimens and clinical data. We thank the participating clinicians for their support in recruitment of subjects.

#### Funding information

This research was supported by the NIMH of the NIH under award no. 5R01MH115676. Additional support came from the NINDS under award no. T32 NS048004 and from the NHGRI under award no. T32 HG002536.

### NTR NESDA

#### Cohort description

This cohort consists of two subcohorts from different projects: the Netherlands Study of Depression and Anxiety (NESDA)[1] and the NTR[2]. Additional details can be found in Võsa *et al.*[3].

#### Ethics approval

The NESDA and NTR were both approved by the Central Ethics Committee on Research Involving Human Subjects of the VU University Medical Center, Amsterdam (IRB number IRB-2991 under Federal-wide Assurance 3703; IRB/institute codes: NESDA 03-183 and NTR 03-180). All participants provided written informed consent.

#### Genotyping and sample QC

Genotyping of NTR/NESDA DNA samples was performed on the Affymetrix 6.0 SNP array platform. Genotype calling was executed following the manufacturer's protocols and white papers. The resulting genotypes were based on build 37. For each individual platform, DNA samples were checked for sex mismatches, heterozygosity (PLINK F value between -0.10 to 0.10), and PLINK-estimated IBD mismatches in comparison to the known family structure. For the call rate in the samples, each sample needed to have at least 90% genotyped and at least 80% of the genotypes needed to be present on each separate chromosome 1−22 plus X for each person.

### Morocco

#### Cohort description

The Morocco cohort includes 188 healthy Arab and Amazigh individuals from a city and two villages in Morocco[1,2]. Sampling was designed so that four localities representing two main lifestyles, urban and rural, and both genders were sampled. All study participants were between the ages of 18 and 50 years, and the mean age of the three locations was similar (31–34 yr). After the QC steps described in the link below, 136 samples were kept. All data preprocessing and analyses were conducted using eQTLGen pipelines, according to eQTLGen cookbook (https://eqtlgen.github.io/eqtlgen-web-site/eQTLGen-p2-cookbook.html).

#### Ethics approval

The study was approved by the ethical review committees of the Moroccan Ministry of Health, North Carolina State University and the University of Queensland.

#### Methods: expression data

Peripheral blood samples (~8 ml) were collected over the course of 6 days during June and July 2008. The total leukocyte population was isolated from ~6 ml, and its total RNA was stabilized within minutes using a LeukoLOCK Total RNA Isolation System5 (Ambion). HumanHT-12 beadchips (Illumina) were used to generate expression profiles of >48,000 transcripts using 500 ng of labeled cRNA for each sample. We received the log 2 transformed expression data and converted it to raw counts prior to running the eQTLgen QC pipeline.

#### Methods: genotyping data

The genotype data were imputed to the 1000 Genomes Phase 1 Version 3 reference panel with IMPUTE v2, imputation QC threshold $R^2 \geq 0.3$. Variants were filtered by HWE $P < 10^{-3}$, missingness per individual < 10%, and missingness per marker < 2%. To confine the imputed data, we extracted the directly typed variants from the imputed genotype data. The genotype data were then processed by the eQTLgen pipeline.

#### Data availability statement

The expression data are available at NCBI GEO available under accession number GSE17065.

### Jackson Heart Study

#### Cohort description

The Jackson Heart Study (JHS) is a community-based cohort study investigating causes of cardiovascular and related diseases among African American adults, with the aim of more effective prevention and treatment. JHS recruited 5,306 participants from the Jackson, Mississippi metropolitan area[1]. Peripheral blood mononuclear cells (PBMCs) from a subset of consenting participants were cryopreserved, as described in Wilson et al. [1]

#### Ethics approval

The JHS was approved by the University of Mississippi Medical Center IRB, approval 1998-6004. All participants provided written consent for study participation.

#### Methods: expression data

PBMCs from a subset of consenting JHS participants were cryopreserved, as described.[1] RNA-seq for participants with adequate remaining cryopreserved PBMCs and genetic analysis consent (n = 1,027) was performed at the University of Washington Northwest Genomics Center, with an average read-depth of 50M. For library construction, total RNA was scaled to 7.5 ng/µl (total volume 50µl) on a Perkin Elmer Janus II Workstation. Poly-A selection and cDNA synthesis were performed using the TruSeq Stranded mRNA kit (Illumina, cat#RS-122-2103). All steps were automated on the Perkin Elmer Sciclone NGSx Workstation. Final RNA-seq libraries were quantified using the Quant-it dsDNA High Sensitivity assay, and library insert size distribution was checked using a fragment analyzer (Advanced Analytical, kit ID DNF474). Samples where adapter dimers constituted > 4% of the electropherogram area were excluded. Successful libraries were normalized and pooled prior to sequencing. For read processing, base call generation was conducted on the NovaSeq6000 instrument (RTA 3.1.5). Next, demultiplexed, unaligned BAM files produced by Picard ExtractIlluminaBarcodes and IlluminaBasecallsToSam were converted to FASTQ format using SamTools bam2fq (v1.4). Sequence read and base quality checking was performed using the FASTX toolkit (v0.0.13), and sequence alignment to GRCh38 with reference transcriptome GENCODE release 30 was performed using STAR (v2.6.1d)[2].
The gene-level expression quantification was estimated as transcripts per million for each gene, which was quantitated and released on the merged-level data across individuals with RSEM (v1.3.1). We then performed sample-, variant-, and gene-level QC for the RNA-seq data. We removed five individuals due to low RNA-seq quality and two individuals due to mismatched sex compared to the recorded sex from JHS. We then used VerifyBamID[3] to check the individual consistency between the RNA-seq and WGS data. Eight individuals who failed VerifyBamID (inconsistency rate ≥ 2%), likely due to potential mixing or contamination of the samples, were also removed. After sample-level QC, 1,012 participants remained. Further details have been published previously.[4]

#### Methods: genotyping data

WGS data from 3,406 JHS participants are available from the NHLBI Trans-Omics for Precision Medicine (TOPMed) Program[5]. We here used WGS data with minimum depth 0 (DP0) from freeze 8 (unphased), with methods described at https://topmed.nhlbi.nih.gov/topmed-whole-genome-sequencing-methods-freeze-8.

#### Data availability statement

JHS RNA-seq data is being submitted to phs000286 in dbGaP. Genome sequencing data is available at phs000964.


#### Acknowledgements

The JHS is supported by contracts HHSN268201800010I, HHSN268201800011I, HHSN268201800012I, HHSN268201800013I, HHSN268201800014I, and HHSN268201800015I from the NHLBI, with additional support from the National Institute of Minority Health and Health Disparities (NIMHD).
The views expressed in this manuscript are those of the authors and do not necessarily represent the views of the NHLBI, the NIMHD, the NIH, or the U.S. Department of Health and Human Services. Molecular data for the TOPMed program was supported by the NHLBI. Genome sequencing for "NHLBI TOPMed: The Jackson Heart Study" (phs000964.v1.p1) was performed at the Northwest Genomics Center (HHSN268201100037C). Core support including centralized genomic read-mapping and genotype calling, along with variant quality metrics and filtering, were provided by the TOPMed Informatics Research Center (3R01HL-117626-02S1; contract HHSN268201800002I). Core support including phenotype harmonization, data management, sample-identity QC, and general program coordination were provided by the TOPMed Data Coordinating Center (R01HL-120393; U01HL-120393; contract HHSN268201800001I). We gratefully acknowledge the studies and participants who provided biological samples and data for TOPMed. The TOPMed Banner Authorship List can be found at: https://www.nhlbiwgs.org/topmed-banner-authorship.

#### Funding information

The project is supported by funding from the NIH through R01 HL129132 (to YL), R01AG075884 (to LMR), and R01HL146500 (to APR). YL is also partially supported by U01HG011720 and R01HL163972. J.W. was supported by NIH grant 5T32ES007018. The content is solely the responsibility of the authors and does not necessarily represent the official views of the NIH.

### Japan COVID-19 Task Force

#### Cohort description

The study participants were recruited through Japan COVID-19 Task Force (JCTF), which is described in detail in previous manuscripts[1,2]. In brief, hospitalized patients diagnosed with COVID-19 by physicians using the clinical manifestation and PCR test results were recruited at more than 100 affiliated hospitals across Japan. The samples were annotated with four levels of phenotype severity: “Most severe” for patients in intensive care units (ICU) or requiring intubation and ventilation, “Severe” for others requiring oxygen support, “Mild” for other symptomatic patients (e.g. shortness of breath), and “Asymptomatic” for those without COVID-19 related symptoms.

#### Ethics approval

This study was approved by the ethical committees of Keio University School of Medicine, Osaka University Graduate School of Medicine, and affiliated institutes (Keio IRB approval 20200061, Osaka University IRB approval 734-14, University of Tsukuba IRB approval H29-294). Informed consent was obtained from all participants.

#### Methods: expression data

RNA-seq was performed using the NovaSeq6000 platform (Illumina) with paired-end reads (read length of 100bp), using S4 Reagent kit (200 cycles). Read count quantification was based on the analysis pipeline provided by the GTEx (https://github.com/broadinstitute/gtex-pipeline), with minimal changes. Specifically, RNA-seq data was first aligned to hg38 human reference genome (excluding ALT, HLA, and decoy contigs) using STAR v2.5.3, with parameter ‘--sjdbOverhang 100’ instead of 75. Transcript amounts were quantified using RSEM v1.3.0.

#### Methods: genotyping data

The study samples were genotyped using Infinium Asian Screening Array (Illumina). Stringent sample and variant level QC filters were applied (e.g., sample call rate > 0.97, variant call rate > 0.99).

#### Data availability statement

The summary statistics of QTL analyses, as well as the RNA-seq expression matrix, are available at the National Bioscience Database Center Human Database (accession code: hum0343; https://humandbs.biosciencedbc.jp/en/hum0343). The QTL summary statistics are available in an interactive browser (https://japan-omics.jp). The individual genotype data is available at the EGA (accession code: EGAS00001006284 (https://ega-archive.org/studies/EGAS00001006284).

#### Acknowledgements

We would like to sincerely thank all the participants involved in this study and all the members of Japan COVID-19 Task Force for their support. We thank Mr. Johji Kitano, e-Parcel Corporation, and Ascend Corporation for voluntarily supporting Japan COVID-19 Task Force.

#### Funding information

This study was supported by AMED (JP23kk0305022, JP22ek0410075, JP23km0405211, JP23km0405217, JP23ek0109594, JP23ek0410113, JP223fa627002, JP223fa627010, JP233fa627011, JP23zf0127008, JP22fk0108510, JP21fk0108553, JP21fk0108431, JP20fk0108415,

JP20fk0108452), JST CREST (JPMJCR20H2), JST FOREST (JPMJFR225Y), JST PRESTO (JPMJPR21R7), JST Moonshot R&D (JPMJMS2021, JPMJMS2024), MHLW (20CA2054), JSPS KAKENHI (22H00476, 23K14233), the Nakajima Foundation, the Uehara Memorial Foundation, Takeda Science Foundation, the Mitsubishi Foundation, and the Bioinformatics Initiative of Osaka University Graduate School of Medicine, Institute for Open and Transdisciplinary Research Initiatives, Center for Infectious Disease Education and Research (CiDER), and the Center for Advanced Modality and DDS (CAMaD), Osaka University. The supercomputing resource was provided by the Human Genome Center (the Univ. of Tokyo).

### INTERVAL

#### Cohort description

The INTERVAL study is a prospective cohort study of approximately 50,000 participants nested within a randomized trial of varying blood donation intervals.[1,2] Between 2012 and 2014, blood donors aged 18 years and older were recruited at 25 centers of England's National Health Service Blood and Transplant. Participants were generally in good health as blood donation criteria exclude individuals with a history of major diseases (e.g. myocardial infarction, stroke, cancer, HIV, and hepatitis B or C) and those who have had a recent illness or infection. Participants completed an online questionnaire comprising questions on demographic characteristics (e.g. age, sex, ethnicity), lifestyle (e.g. alcohol and tobacco consumption), self-reported height and weight, diet, and use of medications. The INTERVAL data that is used for this study has been presented before by Tokolyi et al.[3].

#### Ethics approval

All participants gave informed consent before joining the study and the National Research Ethics Service approved this study (11/EE/0538).

#### Methods: expression data

*Blood collection.* Blood samples were collected from all INTERVAL participants at baseline and from ~60% of participants approximately 24 months after baseline. For a subset of ~5,000 participants at the 24-month time point, an aliquot of 3 ml of whole blood was collected in Tempus Blood RNA Tubes (Thermo Fisher Scientific) following the manufacturer's instructions, and then transferred at ambient temperature to the UK Biocentre (Stockport, UK). Samples were stored at -80°C until use.
*RNA extraction.* RNA extraction was performed by Qiagen Genomic Services using Qiagen's proprietary silica technology. QC of the extracted RNA was performed by spectrophotometric measurement on an Infinite 200 Microplate Reader (Tecan). RNA Integrity Number (RIN) values were determined using a TapeStation 4200 system (Agilent), following the manufacturer's protocol. Samples with a concentration < 20 ng/µl and a RIN < 4 were excluded from further analyses.
*Automated RNA-seq library preparation.* Samples were quantified with a QuantiFluor RNA System (Promega) using a Mosquito LV liquid handling platform (SPT Labtech), Bravo automation system (Agilent), and FLUOstar Omega plate reader (BMG Labtech), and then cherry-picked to 200 ng in 50 µl (= 4 ng/µl) using a liquid handling platform (Tecan Freedom EVO). Next, mRNA was isolated using a NEBNext Poly(A) mRNA Magnetic Isolation Module (NEB) and then re-suspended in nuclease-free water. Globin-depletion was performed using a KAPA RiboErase Globin Kit (Roche). RNA library preparation was done using a NEBNext Ultra II RNA Library Prep Kit for Illumina (NEB) on a Bravo NGS workstation automation system (Agilent). PCR was performed using a KapaHiFi HotStart ReadyMix (Roche) and unique dual-indexed tag barcodes on a Bravo NGS workstation automation system (Agilent). We applied the following PCR program: 45 sec at 98°C, 14 cycles of 15 sec at 98°C, 30 sec at 65°C and 30 sec at 72°C, followed by 60 sec at 72°C. Using a Zephyr liquid handling platform (PerkinElmer), PCR products were purified using AMPure XP SPRI beads (Agencourt) at a 0.8:1 bead:sample ratio and then eluted in 20 µl of Elution Buffer (Qiagen). RNA-seq libraries were quantified with an AccuClear Ultra High Sensitivity dsDNA Quantitation Kit (Biotium) using a Mosquito LV liquid handling platform (SPT Labtech), Bravo automation system (Agilent), and FLUOstar Omega plate reader (BMG Labtech). Then, libraries were pooled up to 95-plex in equimolar amounts on a Biomek NX-8 liquid handling platform (Beckman Coulter), quantified using a High Sensitivity DNA Kit on a 2100 Bioanalyzer (Agilent), and normalized to 2.8 nM prior to sequencing.
*RNA-sequencing and data preprocessing.* Samples were sequenced using 75bp paired-end sequencing reads (reverse stranded) on a NovaSeq 6000 system (S4 flow cell, Xp workflow; Illumina). The sequencing data were deplexed into separate CRAM files for each library in a lane. Adapters that

had been hard-clipped prior to alignment were reinserted as soft-clipped post-alignment, and duplicated fragments were marked in the CRAM files. The data preprocessing, including sequence QC and STAR, and alignments were performed with the Nextflow pipeline publicly available at https://www.intervalstudy.org.uk/wp-content/uploads/2024/02/Data-Access-Policy-v1.0-14Apr2020.pdf, including the specific aligner parameters. We assessed the sequence data quality using FastQC v0.11.8. Samples mismatched between RNA-seq and genotyping data within the cohort were identified using QTLtools MBV v1.2[4]. Reads were aligned to the GRCh38 human reference genome (Ensembl GTF annotation v99) using STAR v2.7.3a[5]. The STAR index was built against GRCh38 Ensembl GTF v99 using the option -jdbOverhang 75. STAR was run in a two-pass setup with standard ENCODE options to increase mapping accuracy, as follows: (i) a first alignment step of all samples was used to discover novel splice junctions, (ii) splice junctions of all samples from the first step were collected and merged into a single list, and (iii) a second step realigned all samples using the merged splice junctions list as input. We used featureCounts v2.0.0[6] to obtain a count matrix.

*Gene expression quantification.* Sequencing was performed across 15 batches. The raw gene-level count data contained 60,676 genes across 4,778 individuals. On average, each sample had 25.3 million unique reads (interquartile range = 21.5−26.9, including batches 1 and 15, for which libraries were sequenced twice).

*QC of gene expression data.* We filtered samples of poor quality by removing samples with a read-depth below 10 million uniquely mapped reads. A relatedness matrix was obtained using the PLINK v1.9[7] -make-rel 'square' command on pruned genotype data, and a cut-off threshold of 0.1 was used to define related individuals. For each pair of related individuals, one individual was arbitrarily removed. After filtering, the gene expression dataset included 4,731 individuals. We retained 60,580 genes located on autosomal and sex chromosomes. The raw expression matrix was then automatically processed with eQTLGen pipelines.

#### Methods: genotyping data

In brief, DNA extracted from buffy coat samples collected from INTERVAL participants at the study baseline was used to assay approximately 830,000 variants on the Affymetrix Axiom UK Biobank genotyping array.[8] Genotyping and sample QC were performed as previously described.[8] Next, genotype data was automatically quality-controlled, pre-phased, and imputed by eQTLGen pipelines.

#### Data availability statement

The INTERVAL study data used in this paper are available to bona fide researchers from. The data-access policy for the data has been approved by the ethics committee and is available at https://www.donorhealth-btru.nihr.ac.uk/wp-content/uploads/2020/04/Data-Access-Policy-v1.0-14Apr2020.pdf. The generated RNA-sequencing data have been deposited at the EGA under the accession number EGAD00001008015.

#### Acknowledgements

Participants in the INTERVAL randomized controlled trial were recruited with the active collaboration of NHS Blood and Transplant England (https://www.nhsbt.nhs.uk/), which has supported field work and other elements of the trial. DNA extraction and genotyping were co-funded by the National Institute for Health and Care Research (NIHR), the NIHR BioResource (https://bioresource.nihr.ac.uk/), and the NIHR Cambridge Biomedical Research Centre (BRC-1215-20014) [*]. RNA-seq was funded as part of an alliance between the University of Cambridge and the AstraZeneca Centre for Genomics Research and by the NIHR Cambridge Biomedical Research Centre (BRC-1215-20014) [*]. The academic coordinating center for INTERVAL was supported by core funding from the NIHR Blood and Transplant Research Unit (BTRU) in Donor Health and Genomics (NIHR BTRU-2014-10024), NIHR BTRU in Donor Health and Behaviour (NIHR203337),

UK Medical Research Council (MR/L003120/1), British Heart Foundation (SP/09/002; RG/13/13/30194; RG/18/13/33946 and RG/F/23/110103), BHF Chair Award (CH/12/2/29428), and NIHR Cambridge BRC (BRC-1215-20014; NIHR203312) [*]. A complete list of the investigators and contributors to the INTERVAL trial is provided in Di Angelantonio et al.[2]
The academic coordinating center would like to thank blood donor center staff and blood donors for participating in the INTERVAL trial. This work was supported by Health Data Research UK, which is funded by the UK Medical Research Council, Engineering and Physical Sciences Research Council, Economic and Social Research Council, Department of Health and Social Care (England), Chief Scientist Office of the Scottish Government Health and Social Care Directorates, Health and Social Care Research and Development Division (Welsh Government), Public Health Agency (Northern Ireland), British Heart Foundation, and Wellcome.
The Wellcome Sanger Institute is supported by core funding from the Wellcome Trust (206194 and 220540/Z/20/A). We thank the Wellcome Sanger Institute's Scientific Operations team for their contribution to sequencing data generation. For the purpose of Open Access, the authors have applied a CC BY public copyright license to any Author Accepted Manuscript version arising from this submission. This work was supported by the Cambridge Service for Data Driven Discovery (CSD3) operated by the University of Cambridge Research Computing Service (https://www.csd3.cam.ac.uk/), provided by Dell EMC and Intel using Tier-2 funding from the Engineering and Physical Sciences Research Council (capital grant EP/P020259/1), and DiRAC funding from the Science and Technology Facilities Council (https://dirac.ac.uk/).

#### Funding information

E.P. was funded by the EU/EFPIA Innovative Medicines Initiative Joint Undertaking BigData@Heart (grant 116074) and by the NIHR BTRU in Donor Health and Behaviour (NIHR203337) [*]. A.T. is supported by the Wellcome Trust (PhD studentship 222548/Z/21/Z). M.I. is supported by the Munz Chair of Cardiovascular Prediction and Prevention and the NIHR Cambridge Biomedical Research Centre (BRC-1215-20014; NIHR203312) [*]. M.I. is also supported by the UK Economic and Social Research Council (ES/T013192/1). M.I. is a trustee of the Public Health Genomics Foundation, a member of the Scientific Advisory Board of Open Targets, and has a research collaboration with AstraZeneca that is unrelated to this study. D.S.P. is an employee and stockholder of AstraZeneca.

*The views expressed are those of the authors and not necessarily those of the NIHR or the Department of Health and Social Care.

### HELIOS

#### Cohort description

The Health for Life in Singapore (HELIOS) study is a longitudinal population resource that aimed to understand the diseases and health states that are important to Asian populations. A total of 10,004 Asian men and women aged 30 to 84 years were recruited between 2018 and 2022 (www.healthforlife.sg). Participants were recruited from the Singapore general population. The cohort includes 6,784 people who identified as Chinese or other East Asian background, 1,807 people of Indian or other South Asian background, and 1,354 people of Malay or other South-East Asian heritage. There were 59 participants from other backgrounds. Data collected include demographic, clinical, molecular, and genetic epidemiological information.

#### Ethics approval

HELIOS asks permission from participants to use the data and samples they contribute for clinical and molecular epidemiological research focused on improving human health. This includes the application of 'untargeted' molecular profiling techniques that assess genomic, proteomic, transcriptional, metabolomic, and other 'omic' variation in the biological samples collected. Participant consent also includes permission for linkage to disease registers, medical records, social care datasets, and other health-related datasets held by Singapore's public bodies. Linkage is enabled by the Singapore NRIC, a unique national identifier allocated at birth, with universal coverage. Consent provides permission for use of the data and samples from participants by both academic and industry researchers and for recontact of participants, including recontact based on phenotypic or genotypic characteristics. The HELIOS study operates under the governance framework of the Nanyang Technological University, with IRB approval (Ref: 2016-11-030).

#### Methods: expression data

Blood collection

Blood was collected from all participants at a single time point by a certified phlebotomist during the baseline assessment visit and processed according to well established protocols, complying with international best practice. The blood samples were either analyzed immediately for hematology, coagulation, and biochemistry tests or stored at -80°C for future research use.

##### Library preparation

RNA-seq libraries were prepared using samples of whole blood (n = 1,234) collected in PaxGene RNA tubes at enrollment. RNA-seq libraries were prepared from at least 1 mg of total RNA using NEBNext® Ultra™ II Directional RNA Library Prep (New England Biolabs, Inc.), with GLOBINclear (Thermo Fisher Scientific) for depletion of globin gene RNA and rRNA. The libraries were sequenced on a NovaSeq6000, using a paired-end run of 2 x 150bp.

##### Data processing

Sequenced data with at least 30M aligned reads per library (~9 Gb of data) were considered for this study. Adapter and quality trimming were performed using TrimGalore[1], and SortMeRNA[2] was used for the removal of rRNA. Alignment to the GRCh38 reference genome was conducted using STAR version 2.7.9a[3], followed by quantification of reads with RSEM version 1.3.3[4], which identified a total of 60,708 genes. Sex mismatch check was performed by checking for anomalies across five genes: *XIST*, *RPS4Y1*, *EIF1AY*, *DDX3Y*, and *KDM5D*. A total of six samples failed this check, resulting in 1,228 samples for downstream analysis.

##### Common sequenced samples

A total of 1,168 samples have both RNA-seq and WGS data. This results in 60,664 genes detected.

#### Methods: genotyping data

##### Data processing

WGS was carried using the NovaSeq platform, with data processing using DRAGEN v3.7.8. Individual sample VCF files were transformed into HAIL matrix tables.[5] Multi-allelic sites were efficiently split into multiple rows of bi-allelic sites, ensuring a comprehensive representation of the genetic variation. Samples were merged in batches of 1,000 to create a unified HAIL matrix table representing the sample cohort, with 258,062,302 genetic variants. Stringent variant and sample QC parameters were employed to ensure the accuracy and reliability of the genomic data. These included a number of q30_bases (threshold 77.5GB high quality bases), as well as ratios for transition/transversion, heterozygous/homozygous variation, and insertion/deletion, applying a threshold of 6x median absolute deviation for each. Samples exhibiting > 1% cross-contamination, a call rate < 95%, autosomal coverage < 95% at 15X, or discordant sex information (reported vs genetically determined) were also flagged. The QC metrics were added as annotations to the HAIL matrix table, which was then converted to a merged VCF file of 10,000 samples. The VCF file was also converted and stored as PLINK2[6] binary files to perform downstream analysis.

#### Data availability statement

The HELIOS data are protected and are not publicly available due to data privacy regulations. Data-access request can be submitted to the HELIOS Data Access Committee by emailing for details.

#### Acknowledgements

We thank all participants and research staff who made the study possible. The computational work for this study was partially performed on resources of the National Supercomputing Centre, Singapore (https://www.nscc.sg).

#### Funding information

This study is supported by Singapore Ministry of Health's (MOH) National Medical Research Council (NMRC) under its OF-LCG funding scheme (MOH-000271-00); the Singapore Translational Research (StaR) funding scheme (NMRC/StaR/0028/2017); the National Research Foundation, Singapore through the Singapore MOH NMRC and Precision Health Research, Singapore (PRECISE) under the National Precision Medicine programme (NMRC/PRECISE/2020); and intramural funding from Nanyang Technological University, Lee Kong Chian School of Medicine, and the National Healthcare Group. RNA-seq was partially funded by a Ministry of Education Academic Research Fund Tier 1 Grant (RS09/20), a A*STAR-National Health and Medical Research Council (NHMRC) Joint Grant Call (A20PRb0138), a Start-Up Grant (awarded to M. Loh [PI]) from Lee Kong Chian School of Medicine, Nanyang Technological University, Singapore, and the Imperial - Nanyang Technological University Collaboration Fund (awarded to M. Loh [PI]).

### GAIT-2

#### Cohort description

The Genetic Analysis of Idiopathic Thrombophilia 2 (GAIT-2) cohort has been described previously[1,2]. Detailed criteria for the recruitment of these 915 individuals from 35 Spanish families, with an average of 27 individuals per pedigree and a total of 8654 related pairs, have been described in the original papers. In short, families were recruited with a proband with idiopathic thrombophilia, and all families have at least 10 living individuals over at least three generations, with a maximum of five generations. The mean age at study enrollment was 39.5, with ages ranging from 2.6 to 101.1. There were 462 men and 454 women included in the study. Additional information about the composition of the families and the collection of lifestyle, medical, and family history data is included in Souto et al.[1].

#### Ethics approval

The study was performed according to the Declaration of Helsinki. All procedures of the study were reviewed by the IRB of the Hospital de la Santa Creu i Sant Pau, Barcelona, Spain. Adult subjects gave informed consent for themselves and for their minor children.

#### Methods: blood collection

Blood was collected by vein puncture following a 12-hour fast. Samples were collected in 1/10 volume containing 0.129 mol/L sodium citrate. None of the participants were using oral anticoagulants or heparins at the time of blood collection. Platelet-poor plasma was obtained by centrifugation at 2000 g for 20 minutes at room temperature (22 ± 2°C) and stored at -80°C before performing the thrombin generation assay. DNA was extracted from whole-blood samples using a standard salting-out procedure[3].

#### Methods: expression data

In addition to gene expression, imputed genotypes were available. The 49-bp sequenced paired-end reads were mapped to the GRCh37 reference genome[4] with GEM[5]. Samples with fewer than 5 million exonic reads were excluded.

#### Methods: genotyping data

Individuals were genotyped using two different strategies. A set of 324 individuals were genotyped with the HumanOmniExpressExome-8v1.2 chip. These 324 individuals were the founder members of the families included in the study and individuals who were not related to these founders. The remaining 610 individuals were genotyped with the HumanCoreExome-12v1.1 chip. Genotype data was further processed by eQTLGen pipelines.

#### Methods: correction of the relatedness of the samples

With the GAIT2 cohort consisting of families, we adjusted the gene expression QC to account for sample-relatedness. After gene expression levels were inverse normal transformed, we regressed them on the cohort's kinship matrix using lme4qtl's `relmatLmer` function[6]. The residuals were then used to calculate principal components and used for downstream analyses.

#### Data availability statement

Data is currently being deposited in EGA.

#### Acknowledgements

#### Funding information

This work was funded by the Regional Government of Catalonia under grants 2021_SGR_00830 and CERCA Program, the Spanish Ministry of Health under grants FIS_PI12_0612 and FIS_PI20_00325, and the non-profit association Activa´TT por la Salud. Genotyping was performed at the Spanish National Cancer Research Centre at the Human Genotyping Laboratory, supported by ISCIII and ERDF under grant PT17/0019. Center for Biomedical Network Research on Rare Diseases (CIBERER).

### GAinS

#### Cohort description

The UK Genomic Advances in Sepsis (GAinS) study (NCT00121196) is a biobank of samples from >1,000 patients, established to characterize genetic variants associated with susceptibility to and outcomes from sepsis.[1,2] Adult patients (>18 years old) were recruited from 34 ICUs between 16/11/2005 and 30/05/2018. Inclusion criteria were sepsis diagnosed according to ACCP/SCCM guidelines due to community acquired pneumonia (CAP) or fecal peritonitis (FP). CAP was defined as febrile illness associated with cough, sputum production, breathlessness, leukocytosis, and radiological features of pneumonia acquired in the community or within two days of admission to hospital. FP was defined as inflammation of the peritoneal membrane secondary to fecal contamination, diagnosed by laparotomy. Exclusion criteria were immunosuppression, admission for palliative care only, and pregnancy.

#### Ethics approval

Ethics approval was granted nationally and locally for individual participating centers, and informed consent was obtained from the patient or their legal representative.

#### Methods: expression data

Samples were taken for RNA extraction on the first, third, and/or fifth day after ICU admission. The total leukocyte population was isolated at the bedside using the LeukoLOCK filter system (Life Technologies), and RNA was extracted using the Total RNA Isolation Protocol.
As described in Cano-Gamez et al (2022)[3], stranded libraries were prepared for 909 samples from 695 sepsis patients using NEB Ultra II Library Prep kits (Illumina) with NEBNext Poly(A) mRNA Magnetic isolation, and sequenced using an Illumina NovaSeq 6000 system. Reads were aligned to the reference genome (GRCh38.99) using STAR v2.7.3a[4], and gene expression was quantified using featureCounts v2.0.0[5] using an in-house Nextflow pipeline (https://github.com/wtsi-hgi/nextflow-pipelines/tree/2e5ac3cee33ca2a1ced2943bb7e366a7771a4d3c). To improve the accuracy of HLA gene expression quantification, HLA alleles were imputed using arcasHLA 0.2.0 and HIBAG v.1.4[6,7] from RNA-seq and genotyping data, respectively. These imputed alleles were then used to define personalized reference sequences for HLA gene expression re-quantification. MBV[8] was used to identify and resolve mismatches between genotyping and RNA-seq gene expression data. 45 samples were excluded following QC based on mapping rate, PCA outliers, non-resolution of sample mix-ups, and detection of contamination using VerifyBamID.[9] This resulted in a dataset of 864 samples from 667 patients. For use in eQTLGen II, the cohort was restricted to the first available sample for each patient.

#### Methods: genotyping data

DNA was extracted from buffy coat or whole-blood samples using the Qiagen DNA extraction protocol, the automated Maxwell Blood purification kit (Promega), or the QIAamp Blood Midi kit protocol (Qiagen). Genotyping data were generated in three batches: 295 patients using the Illumina HumanOmniExpress BeadChip (730,525 variants), 655 patients using the Infinium CoreExome BeadChip (551,839 variants), and 307 patients (including reanalysis of 38 samples that previously failed QC) using the Infinium GSA BeadChip (654,027 SNPs). Given the low number of variants genotyped in all three batches, each batch was imputed separately and stringently filtered prior to being processed with the eQTLgen pipeline.
Genotyping QC was performed within each batch in PLINK 1.9[10] according to the methods described in Anderson et al (2010).[11] Samples were excluded on the basis of discordant sex information,

proportion of missing genotypes > 0.02, outlying heterozygosity rate, IBD (Pi_hat ≥ 0.1875), and detection of sample mix-ups through comparison to RNA-seq on the same patient.[8] Variants with a missing data proportion > 0.05, MAF < 0.01, and HWE P < $1x10^{-5}$ were excluded. Each of the three genotyping data sets were imputed using the Haplotype Reference Consortium (HRC) release 1.1 panel and the Sanger Imputation Service[12], following checks for strand, alleles, position, ref/alt assignments, and frequency differences versus the HRC (http://www.well.ox.ac.uk/~wrayner/tools/). Genotypes were phased using Eagle2[13] and imputed using Positional Burrows-Wheeler Transform[14]. Only SNPs with an imputation info score of 1 were retained before the datasets were combined and used as input for the eQTLgen imputation pipeline.

#### Data availability statement

The raw RNA-seq data are available on the EGA via a data-access committee (Accession ID: EGAD00001008730). The genotyping data will be available on the EGA via a data-access committee upon publication. Further information and requests for resources should be directed to and will be fulfilled by Emma Davenport and Julian Knight.


#### Acknowledgements

We thank all the patients, patient families, nurses, and clinicians who participated in the UK GAinS study. We thank Iaroslav Popov and the Wellcome Sanger Institute's Human Genetics Informatics team for facilitating application of the eQTLGen pipelines to the cohort.

#### Funding information

This work was funded by the Wellcome Trust Investigator Award (204969/Z/16/Z) (to JCK) and Wellcome Trust core funding to the Wellcome Sanger Institute (Grant numbers 206194 and 108413/A/15/D) and the Medical Research Council (MR/V002503/1) (to JCK and EED).

### EstBB (RNA-seq)

#### Cohort description

The Estonian Biobank (EstBB) is a population-based cohort of approximately 200,000 biobank participants (~20% of the Estonian adult population), with a rich variety of phenotypic and health-related information collected for each individual[1,2]. At recruitment, participants signed a consent to allow follow-up linkage of their electronic health records (EHR), including lab measurements, thereby providing a longitudinal collection of phenotypic information. EstBB links regularly with the National Health Insurance Fund (from 2004) and other relevant registries. For every participant, there is information on diagnoses in International Classification of Disease 10 (ICD-10) codes (version 2019). All data preprocessing and analyses were conducted using eQTLGen pipelines, according to eQTLGen cookbook (https://eqtlgen.github.io/eqtlgen-web-site/eQTLGen-p2-cookbook.html).

#### Ethics approval

The activities of the EstBB are regulated by the Human Genes Research Act, which was adopted in 2000 specifically for the operations of the EstBB. The research activities involving biobank participant data have been carried out under the ethical approval nr. 1.1-12/655 (24.03.2020) and its extensions 1.1-12/490 (26.01.2023) and 1.1-12/2573 (13.10.2025) by the Estonian Committee on Bioethics and Human Research (Estonian Ministry of Social Affairs), using data according to release application 6-7/GI/29457 from the Estonian Biobank. The study was conducted in accordance with the Declaration of Helsinki.

#### Methods: expression data

The EstBB RNA-seq dataset has previously been described elsewhere[3]. RNA was extracted from thawed Tempus tubes using TRIzol Reagent (Invitrogen) and further purified using RNeasy Mini Kit (Qiagen). Globin mRNA was depleted using GLOBINclear Kit (Invitrogen). RNA quality was checked using an Agilent 2200 TapeStation (Agilent Technologies). Sequencing libraries were prepared using 200 ng of RNA according to the Illumina TruSeq stranded mRNA protocol. RNA-sequencing was performed at the Estonian Genome Center Core Facility using Illumina paired-end 50bp sequencing technology, according to manufacturer specifications. We used FastQC v.0.11.3 for raw data QC and Trimmomatic (version 0.36)[4] to remove three leading and three trailing bases and to remove adapters. For adapter removal, we used the adapter file provided with fastQC. Additional read quality filtering was done using the FASTX toolkit v.0.0.13 fastq_quality_filter script with minimum quality score 30 and minimum 50% of base pairs with required quality.
QC was done by FastQC (version 0.11.2)[5]. The quality-filtered data were mapped on genome hg19, position sorted, and indexed with STAR v. 2.5.2 (STAR index files were previously provided by eQTLGen)[6]. The mapped data quality statistics were collected with Picardtools v.1.130 CollectRnaSeqMetrics. Read counts were obtained using HTSeq-count script v. 0.6.1 and the GRCh37.v71 annotation file.

#### Methods: genotyping data

Genotyping was performed by the Estonian Genome Center Core Facility according to manufacturer specifications using Illumina Omni Express genotyping arrays. Genotype data was further automatically quality-controlled, imputed, and processed by eQTLGen pipelines.

#### Data availability statement

The Estonian Biobank data is governed by the Estonian Biobank, and the controlled access application procedure is documented on its website (https://genomics.ut.ee/en/content/estonian-biobank; link Data Access).


#### Acknowledgements

This work was carried out in the High Performance Computing Center of University of Tartu (https://hpc.ut.ee/)[7].
Estonian Biobank research team
Andres Metspalu[1], Lili Milani[1], Tõnu Esko[1], Reedik Mägi1, Mari Nelis[1] and Georgi Hudjashov[1]
1. Estonian Genome Centre, Institute of Genomics, University of Tartu

#### Funding information

Urmo Võsa was supported by the European Regional Development Fund, the Mobilitas Pluss program (MOBTP108), and through the Estonian Research Council grant PUT (PRG1291). Tõnu Esko was supported through the Estonian Research Council grant PUT (PRG1291).

### EstBB (Illumina HT12v3)

#### Cohort description

The Estonian Biobank (EstBB) is a population-based cohort of approximately 200,000 biobank participants (~20% of the Estonian adult population) with a rich variety of phenotypic and health-related information collected for each individual[1,2]. At recruitment, participants signed a consent to allow follow-up linkage of their EHR, including lab measurements, thereby providing a longitudinal collection of phenotypic information. The EstBB links regularly with the National Health Insurance Fund (from 2004) and other relevant registries. For every participant, there is information on diagnoses in ICD-10 codes (version 2019). The EstBB also includes subset of individuals for whom llumina HT12v3 array whole-blood gene expression data is available. Samples overlapping with the EstBB RNA-seq dataset were removed, and, after eQTLGen QC, 789 individuals were included to the analyses.
All data preprocessing and analyses were conducted using eQTLGen pipelines, according to the eQTLGen cookbook (https://eqtlgen.github.io/eqtlgen-web-site/eQTLGen-p2-cookbook.html).

#### Ethics approval

The activities of the EstBB are regulated by the Human Genes Research Act, which was adopted in 2000 specifically for the operations of the EstBB. The research activities involving biobank participant data have been carried out under the ethical approval nr. 1.1-12/655 (24.03.2020) and its extensions 1.1-12/490 (26.01.2023) and 1.1-12/2573 (13.10.2025) by the Estonian Committee on Bioethics and Human Research (Estonian Ministry of Social Affairs), using data according to release application 6-7/GI/29457 from the Estonian Biobank. The study was conducted in accordance with the Declaration of Helsinki.

#### Methods: expression data

Whole peripheral blood RNA samples were collected using Tempus Blood RNA Tubes (Life Technologies, NY, USA), and RNA was extracted using Tempus Spin RNA Isolation Kit (Life Technologies). Quality was measured by NanoDrop 1000 Spectrophotometer (Thermo Fisher Scientific, DE, USA) and Agilent 2100 Bioanalyzer (Agilent Technologies, CA, USA). Gene expression levels were obtained by Illumina Human HT12v3 arrays (Illumina Inc, San Diego, US) according manufacturer's protocols. Raw expression data was exported from GenomeStudio and further processed automatically with eQTLGen pipelines.

#### Methods: genotyping data

Genotyping of DNA samples from the EstBB was done at the Core Genotyping Lab of the Institute of Genomics, University of Tartu using the Illumina Global Screening Arrays (GSAv1.0, GSAv2.0, and GSAv2.0_EST). Altogether, 200,000 samples were genotyped, and PLINK format files were created using Illumina GenomeStudio v2.0.4. During QC, all individuals with a call rate < 95% or mismatching sex (defined based on the heterozygosity of X chromosome and sex in the phenotype data) were excluded from the analysis. Variants were filtered by call rate < 95% and HWE P-value < $1\times10^{-4}$ (autosomal variants only). Variant positions were in Genome Reference Consortium Human Build 37, and all variants were changed to be from TOP strand using reference information provided by Dr. Will Rayner from the University of Oxford (https://www.well.ox.ac.uk/~wrayner/strand/). Genotype data was further automatically quality-controlled, imputed, and processed by eQTLGen pipelines.

#### Data availability statement

The expression dataset is available at the GEO public repository under the accession GSE48348. Estonian Biobank genotype data is governed by the Estonian Biobank and the controlled access application procedure is documented on its web site (https://genomics.ut.ee/en/content/estonian-biobank; link Data Access).


#### Acknowledgements

This work was carried out in the High Performance Computing Center of the University of Tartu (https://hpc.ut.ee/)[3].
Estonian Biobank research team
Andres Metspalu[1], Lili Milani[1], Tõnu Esko[1], Reedik Mägi1, Mari Nelis[1] and Georgi Hudjashov[1]
1. Estonian Genome Centre, Institute of Genomics, University of Tartu

#### Funding information

Urmo Võsa was supported by the European Regional Development Fund, the Mobilitas Pluss program (MOBTP108), and through the Estonian Research Council grant PUT (PRG1291). Tõnu Esko was supported through the Estonian Research Council grant PUT (PRG1291).

### IMI DIRECT

#### Cohort description

The IMI DIRECT (Diabetes Research on Patient Stratification) consortium includes pre-diabetic participants (target sample size 2,200–2,700) and patients with newly diagnosed type 2 diabetes (target sample size ~1,000), with detailed metabolic phenotyping. Characteristics of the cohort as well as inclusion/exclusion criteria have been described elsewhere.[1] Fasting blood samples from venous blood were collected, and DNA extractions and other biochemical analyses were carried out.

#### Ethics approval

Approval for the study protocol was obtained from each of the regional research ethics review boards separately (Lund, Sweden: 20130312105459927; Copenhagen, Denmark: H-1-2012-166 and H-1-2012-100; Amsterdam, Netherlands: NL40099.029.12; Newcastle, Dundee and Exeter, UK: 12/NE/0132), and all participants provided written informed consent at enrollment. The research conformed to the ethical principles for medical research involving human participants outlined in the Declaration of Helsinki.

#### Methods: expression data

Details on the RNA-seq analysis have been described elsewhere.[2] Briefly, the mRNA sample quality check was assessed using the TapeStation Software (A.01.04) with an RNA Screen Tape from Agilent. Quality of the libraries was evaluated using Qubit and TapeStation using DNA1000 Screen Tape. The samples were then sequenced on the Illumina HiSec2000 platform using 49bp paired-end reads. For each sample, we evaluated possible samples mix-ups using the function match from the suite QTLtools[3]. To confirm correct assignment of matched DNA/RNA samples and recovered failed genotypes during QC, we re-genotyped samples from 96 individuals. Further validation compared the sex information provided by clinical reports with both genotype data and RNA-seq data. After QC, the total number of samples with paired RNA-seq−genotype data of both RNA-seq and imputed genotypes was 3,029.

#### Methods: genotyping data

Genotyping was conducted using the Illumina HumanCore array (HCE24 v1.0), and genotypes were called using Illumina's GenCall algorithm. Samples were excluded for any of the following reasons: call rate < 97%, low or excess mean heterozygosity, sex discordance, or monozygosity. To identify possible population outliers in the DIRECT data, we performed a PCA using the genotype data from our studied population (3,102 samples, 547,644 markers) using the following cut-offs: MAF > 0.01, HWE > $10^{-4}$, and call rate > 90%. A total of 3,033 samples and 517,958 markers across the two studies passed QC procedures. Only individuals with both expression and genotypes were included in further analyses.

#### Data availability statement

Due to the type of consent provided by study participants and the ethical approvals for this study, individual-level clinical and omics data from IMI-DIRECT cohorts cannot be exported from the centralized IMI-DIRECT repository. Requests for access to IMI-DIRECT data, including for data presented here, can be made to. Requesters will be provided with information and assistance on how data can be accessed via the DIRECT Computerome secure analysis platform following submission of appropriate documentation. The IMI-DIRECT data-access policy is available from www.direct-diabetes.org.

#### Funding information

The DIRECT consortium has received support from the Innovative Medicines Initiative Joint Undertaking under grant agreement n°115317 (DIRECT, http://www.direct-diabetes.org/), resources of which are composed of financial contributions from the European Union's Seventh Framework Programme (FP7/2007-2013) and EFPIA companies' in-kind contribution. Ana Viñuela has received additional support from the AMS Springboard Award (SBF007\100033).

### Knight-ADRC cohort

#### Cohort description

Individuals in this cohort were recruited from four separate studies: the Knight-Alzheimer's Disease Research Center for the Memory and Aging Project, the Dominantly Inherited Alzheimer's Network, the Molecular Genetic Studies of Parkinsonism, and the Dystonia Coalition Projects-3.
The Knight-Alzheimer Disease Research Center (Knight-ADRC) at Washington University in St. Louis has pioneered and led worldwide seminal studies that have expanded our clinical, social, pathological, and molecular understanding of Alzheimer Disease. Over more than 40 years, research volunteers have been recruited to participate in cognitive, neuropsychologic, imaging, fluid biomarkers, genomic, and multi-omic studies. Tissue and longitudinal data are collected to foster, facilitate, and support research on dementia and aging. The Genetics and High-Throughput-Omics core (GHTO) have collected more than 26,000 biological samples from 6,625 Knight-ADRC participants. Samples available include longitudinal DNA, RNA, non-fasted plasma, cerebrospinal fluid (CSF) pellets, and PBMCs. The GHTO has performed deep molecular profiling (genomic, transcriptomic, epigenomic, proteomic, and metabolomic) from a large number of brain (n = 2,117), CSF (n = 2,012), and blood/plasma (n=8,265) samples, with the goal of identifying novel risk and protective variants, novel molecular biomarkers, and causal and druggable targets. Overall, the resources available at the GHTO aid in the exponential increase of our understanding of Alzheimer Disease.
All data generated from Knight-ADRC participants have been and will be deposited at NIAGADS under the Knight ADRC collection (https://www.niagads.org/knight-adrc-collection). All data analyses were run using eQTLGen pipelines, according to eQTLGen cookbook and discussions regarding any adjustments (https://eqtlgen.github.io/eqtlgen-web-site/eQTLGen-p2-cookbook.html).

#### Ethics approval

The IRB of Washington University School of Medicine in St. Louis approved the study with the IRB number 201109148, and the research was performed in accordance with the approved protocols.

#### Methods: expression data

The information discussed in both methods sections has been copied or adapted from Fernandez *et al.*[1]. The blood samples were collected in PAXgene tubes with all RNA-seq undergoing ribo-depletion and globin-depletion, with 150bp reads. Samples were collected as longitudinal blood samples with a variety of phenotypes and backgrounds. The eQTLGen DataQC stage reduced the set to just baseline European samples.
Samples were stored at -80°C until extraction. For extraction, samples were removed from the freezer to thaw overnight. The PAXgene Blood (~2.5 mL blood in a preservative solution) was then processed to be run on the Maxwell RSC 48 using the Maxwell RSC simplyRNA Blood kit. Total RNA integrity was determined using Agilent Bioanalyzer or 4200 Tapestation. Library preparation was performed with 500 ng to 1 µg total RNA. Ribosomal RNA was blocked and globin was depleted using FastSelect reagents (Qiagen) during cDNA synthesis. RNA was fragmented in reverse transcriptase buffer with FastSelect reagent and heating to 94° for 5 minutes, 75° for 2 minutes, 70° for 2 minutes, 65° for 2 minutes, 60° for 2 minutes, 55° for 2 minutes, 37° for 5 minutes, 25° for 5 minutes. mRNA was reverse transcribed to yield cDNA using SuperScript III RT enzyme (Life Technologies, per manufacturer's instructions) and random hexamers. A second strand reaction was performed to yield ds-cDNA. cDNA was blunt ended, an A base was added to the 3' ends, and Illumina sequencing adapters were ligated to the ends. Ligated fragments were then amplified for 15 cycles using primers incorporating unique dual index tags. Fragments were sequenced on an Illumina NovaSeq-6000 using paired-end reads extending 150 bases.

FastQC (v0.11.9) was run on each fastq file to determine quality of data, and data were then aligned with STAR[2] (v2.7.8a) to GRCh38. Transcript counts then were quantified using Salmon[3] (v1.7.0). Post-alignment QC metrics were collected with Picard (http://broadinstitute.github.io/picard) CollectRNAseqMetrics, MarkDuplicates, and CollectAlignmentSummaryMetrics (v2.25.0). Transcript Integrity Numbers (TIN) were calculated with the RSeQC[4] (v5.0.1) package. All QC data was aggregated with multiqc[5] (v1.7.0). QC was performed to remove samples with poor quality sequencing. Samples with low % mapped reads from STAR or Salmon, multiple FastQC failures, or low TIN scores were excluded. Additionally, PCA was performed on vst() normalized counts from DESeq2(v1.38.1) in R (v4.2.2), and the top PCs were plotted. PCA outliers were considered for exclusion.
GATK(v4.2.6.1) was used to perform joint calling by GenomicsDBImport and GenotypeGVCFs on the full dataset[6]. These joint call results generated variant call files that were then sorted with GATK. PLINK (v1.9) was used to run IBD analysis comparing longitudinal RNA samples with each other as well as with genotyping data for matches[7]. This process was run to correct for any mislabels or contaminations at the sample level and to ensure the best accuracy of phenotypes available. Our complete pipelines for RNA-seq processing, QC, and VCF preparation can be found at the GitHub NGI repository. The complete pipeline for brain and blood transcriptomics can be found at: https://github.com/NeuroGenomicsAndInformatics/RNAseq_pipeline. Expression data was further processed with the eQTLGen pipelines.

#### Methods: genotyping data

The Hope Center DNA/RNA Purification Core at Washington University in Saint Louis (associated to the GHTO) uses the Autogen FlexSTAR+ salt precipitation to isolate pure DNA. FlexSTAR is fully automated to perform high quality DNA isolation from large volumes (5−10ml) of whole blood and buffy coat samples. To isolate DNA from small volume samples (< 1 ml) from a range or sources (e.g., blood, buffy coat, saliva, blood cards, tissue, buccal swab, plasma, and CSF) the Core uses a bead-based automatized purification system, the Maxwell 48 automated workstation. After extraction, DNA quality and quantity are assessed with the TapeStation 4200, a high-throughput automated electrophoresis platform that runs up to 96 samples in one run. All samples are then normalized to 100 ng/µL and stored in 2D barcoded tubes at -80°C
Array-based genotyping data generation, QC, and imputation is handled separately for each genotyping round. Once the data reaches the GHTO standard of quality, the genotyping rounds are merged. Briefly, after genotyping, all SNPs are called using Genome Studio. A two-step QC pipeline is implemented to ensure maximum retention of individuals and variants. First, the GHTO uses a loose QC in which all variants and individuals with call rate < 80% are removed, in that order. This is followed by a more stringent second pass using 98% for both parameters, prior to data export into plink format. The PLINK files are used to feed the TOPMed Imputation Server pipeline using the reference genome GRCh38. After imputation, any variant with an imputation quality $R^2 > 0.30$ is retained. For the remaining variants, standard QC is applied: retention of variants and individuals with 98% call rate, removal of SNPs not in HWE, concordance check between reported sex and genetic sex, and concordance with expected IBD estimates for technical replicates and family members (if present). Once the different data generation rounds are merged, a final round of stringent QC is applied. Briefly, variants and individuals are filtered by 98% call rate, and autosomal SNPs not in HWE ($P < 10^{-6}$) are filtered out. The concordance between reported and genetic sex is assessed a second time, and any additional discordances are removed. Finally, IBD estimates allow confirmation of expected duplicated samples, familial relatedness, and removal of any potential sample swaps. All the QC procedures are performed using PLINK1.9 or PLINK2.0[7].
The genotype data is built from six separate arrays. Lists of SNPs from each of these batches' imputed files were combined with their imputation $R^2$ value and with MAF > 0.02. Variants were selected based on being common in all batches, with a minimum $R^2$ across batches of 0.99171, which

created a list of 300,000 SNPs. These SNPs were extracted from a PLINK file prepared with standard QC and used in the eQTLGen pipeline.
This data was further quality-controlled and processed by eQTLGen pipelines.

#### Data availability statement

All data generated from Knight-ADRC participants have been and will be deposited at NIAGADS under the Knight ADRC collection (https://www.niagads.org/knight-adrc-collection).

#### Acknowledgements

We thank the research volunteers and their families, as well as the nurses, physicians, and all personnel involved in participant recruitment and sample collection for the Knight-ADRC, whose help and participation made these studies possible. This work was supported by access to equipment made possible by the Hope Center for Neurological Disorders, the Neurogenomics and Informatics Center (NGI: https://neurogenomics.wustl.edu/), and the Departments of Neurology and Psychiatry at Washington University School of Medicine.

#### Funding information

The recruitment and clinical characterization of research participants at Washington University were supported by the NIH (P30AG066444, P01AG026276, U19AG0032438, P01AG003991). Data generation was supported by NIH grants ((R01AG044546 (to CC), P01AG003991(to CC and JCM), RF1AG053303 (to CC), RF1AG058501 (to CC), U01AG058922 (to CC), RF1AG074007 (to YJS), K99/R00AG062723 (to LI), P30AG066444 (to JCM)), the Chan Zuckerberg Initiative, the Michael J. Fox Foundation (awarded to LI and CC), the Department of Defense (W81XWH2010849 awarded to LI), an Alzheimer's Association Zenith Fellows Award (ZEN-22-848604, awarded to CC), the Bright Focus Foundation (2021033S, awarded to LI), the Alzheimer Drug Discovery Foundation (GDAPB-201807-2015632, awarded to LI), and an Anonymous foundation.

### CHDWB

#### Cohort description

The Emory-Georgia Tech Center for Health Discovery and Well-Being (CHDWB) cohort includes 439 healthy individuals of mixed ethnicity and sex, between the ages of 26 and 79. These participants were recruited as a part of the Emory-Georgia Tech Predictive Health Initiative at the CHDWB, located at Emory University Midtown Hospital in Atlanta, GA. They are broadly representative of Emory employees, covering occupations from janitorial staff to upper administration with roles in healthcare or general academic services, and were free of any known acute illness at the time of recruitment. Participants are taking a wide diversity of medications, but no attempt was made in this study to monitor changes in medication usage or the effect of medication on outcomes. Subjects with uncontrolled or poorly controlled acute or chronic diseases including cardiovascular, endocrine, autoimmune, inflammatory, gastro-intestinal, psychiatric, neurological, musculo-skeletal, infectious, or respiratory disease were excluded. Among 439 individuals, 214 individuals are male and 215 individuals are female. After the QC steps described on the link below, 285 samples were kept.
All data preprocessing and analyses were conducted using eQTLGen pipelines, according to eQTLGen cookbook (https://eqtlgen.github.io/eqtlgen-web-site/eQTLGen-p2-cookbook.html).

#### Ethics approval

Informed consent was obtained from each participant following protocols approved by the IRBs of the two participating institutions: Emory University and the Georgia Institute of Technology.

#### Methods: expression data

The whole-blood samples were stored frozen in Tempus tubes, and RNA was extracted and hybridized at the Emory Biomarker Service Center. We received the log 2 transformed expression data and converted it to the raw counts prior to running the eQTLgen pipelines.

#### Methods: genotyping data

The genotype data were imputed to the 1000 Genomes Phase 1 Version 3 reference panel with IMPUTE v2, imputation quality threshold $R^2 \geq 0.3$. Variants were filtered by HWE $P < 10^{-03}$, missingness per individual < 10%, and missingness per marker < 2%. The total number of SNPs with MAF > 0.01 post QC was 8,131,242. To confine the imputed data, we extracted the directly typed variants from the imputed genotype data. Genotype data were then processed by the eQTLgen pipeline.

#### Data availability statement

The expression data are available at NCBI GEO under accession number GSE35846.

### CanPath

#### Cohort description

The Canadian Partnership for Tomorrow's Health (CanPath) is a population-based cohort providing comprehensive genomic, clinical, behavioral, and environmental data on 330,000 Canadians for the global research community to produce evidence to establish health-related policies. CanPath has collected data from approximately 330,000 volunteer Canadians, including information about health, lifestyle, environment, and behavior. The size of the cohort and the richness of its epidemiological, clinical and biological data positions Canada amongst the world's leaders in longitudinal cancer and chronic disease research. The CanPath subcohort CARTaGENE (CaG) is a regional cohort from Québec province. CaG targeted the segment of the population that is most at risk of developing chronic diseases, with participants' ages ranging from 40 to 60 years old. Health and social-demographic information, such as disease history, physiological measures, lifestyle, and environmental factors, were collected for each individual, along with biological samples[1]

#### Ethics approval

This project is approved by the Research Ethics Board (REB) of the University of Toronto.

#### Methods: expression data

We selected 708 samples (Set 1) from the CaG biobank based on the availability of Tempus Blood RNA Tubes and on Framingham risk scores to ensure an equal distribution of ages and gender. In a second phase, 292 samples (Set 2) were included based on availability of RNA and arterial stiffness measures. These samples, provided by participants with high and average values of arterial stiffness, were chosen to achieve a uniform range of arterial stiffness values.

##### Gene expression

Whole-blood samples from participants included in Sets 1 and 2 were collected in 2010. Total RNA was isolated using Tempus Spin RNA Isolation Kit (Thermo Fisher Scientific), and the GLOBINclear-Human kit (Thermo Fisher Scientific) was used to perform globin mRNA-depletion. All samples displayed high quality and minimal degradation of the RNA, based on a RIN > 7.5. Participants' transcriptomes were obtained by RNA-seq, for which we used paired-end libraries constructed with TruSeq RNA Sample Prep kit v2 (Illumina) with 500 ng of globin-depleted total RNA. Paired-end RNA-seq libraries were inspected before sequencing according to Illumina protocols, and the sequencing was performed on a HiSeq 2000 platform at the Genome Quebec Innovation Center (Montreal, Canada). Set 1 (708 samples) and Set 2 (292 samples) were sequenced by using three and six samples per lane, respectively.

#### Methods: genotyping data

High-density SNP genotyping data for 928 samples with RNA-seq profiles passing QC thresholds were obtained by using the Illumina Omni2.5 array. Variant imputation was conducted on 968 individuals. We pre-phased the genotypes with SHAPEIT (v2.r64410)[2], using the default parameters, on both the autosomes and the chromosome X. We filtered variants for MAF > 1% and HWE P-value > 0.0001 and used the haplotypes within IMPUTE2 (v2.2.2)[3] to perform the imputation using the 1000 Genomes Phase I integrated haplotypes (Dec 2013). We used the parameters Ne = 11418 and call thresh = 0.9. We removed variants with a call rate < 90%, MAF > 1%, and HWE P-value > 0.0001. A total of 9,157,622 variants passed the filters. Of these, 8,877,297 variants were found on the autosomes and included 779,579 indels (8.78%) and 8,097,718 SNPs (91.22%). 280,325 variants were found on the X chromosome, which included 28,504 indels (10.16%) and 251,821 SNPs

(89.84%). After sample preprocessing and QC, 634 samples from Set 1 and 191 samples from Set 2 were included to eQTLGen meta-analyses.

#### Data availability statement

Raw data (.fasta,.bam,.IDAT) or processed data files (gene expression counts, VCF files) can be requested by researchers following REB approval through the CanPath portal at: https://canpath.ca/.

#### Acknowledgements

We thank the participants of CanPath and the research team, including each of the contributing subcohort teams.

#### Funding information

This work is supported by a Genome Canada Grant, Genomics in Application Partnership, awarded to Philip Awadalla, Principal Investigator.

### CAD

#### Cohort description

The Coronary Artery Disease (CAD) cohort is part of the Emory Cardiovascular Biobank, which includes 147 individuals of European ancestry with suspected or confirmed CAD, with age range 48−85. Patients were designated as having non-significant CAD (visible plaque resulting in < 50% luminal stenosis) or significant CAD (at least one major epicardial vessel with ≥ 50% stenosis). Subjects with congenital heart disease, severe valvular heart disease, history of orthotopic heart transplant, severe anemia, recent blood transfusion, active inflammatory diseases, or cancer were excluded. Among 147 individuals, 93 individuals are male and 54 individuals are female. After the QC steps described on the link below, 120 samples were kept.
All data preprocessing and analyses were conducted using eQTLGen pipelines, according to eQTLGen cookbook (https://eqtlgen.github.io/eqtlgen-web-site/eQTLGen-p2-cookbook.html).

#### Ethics approval

The study was approved by the IRBs at Emory University and the Georgia Institute of Technology, Atlanta, GA, USA and was conducted in accordance with the principles of the Declaration of Helsinki. All subjects provided written informed consent.

#### Methods: expression data

Peripheral blood samples were collected immediately prior to angiography and after overnight fasting and stored in PAXgene tubes (Qiagen, San Diego, CA, USA) at -80°C. Microarray analysis of transcript abundance was performed by hybridization of dye-labeled RNA to Illumina HT-12 bead arrays containing probes for all human reference genes. We received the log 2 transformed expression data and converted it to raw counts prior to running the eQTLGen QC pipelines.

#### Methods: genotyping data

The genotype data were imputed to the 1000 Genomes Phase 1 Version 3 reference panel with IMPUTE v2, imputation QC threshold $R^2 \geq 0.3$. Variants were filtered by HWE $P < 10^{-03}$, missingness per individual < 10%, and missingness per marker < 2%. To confine the imputed data, we extracted the directly typed variants from the imputed genotype data. Genotype data were then processed by the eQTLgen pipeline.

#### Data availability statement

The expression data are available at NCBI GEO under accession number GSE49925.

### BEST

#### Cohort description

The BEST (Bangladesh Vitamin E and Selenium Trial) study is a randomized chemoprevention trial evaluating the long-term effects of vitamin E and selenium supplementation on nonmelanoma skin cancer risk among 7,000 individuals with arsenic-related skin lesions living in seven sub-districts in Bangladesh (Argos et al., 2013). Participants included in this work are a subset of BEST participants for whom data is available on genome-wide SNPs and array-based expression.

#### Ethics approval

The study protocol was approved by the Ethical Review Committee of International Center for Diarrheal Disease Research, Bangladesh, the Bangladesh Medical Research Council, and the IRBs of the University of Chicago and Columbia University. Informed consent was obtained from all participants.

#### Methods: expression data

Concentration and quality of RNA samples were assessed on Nanodrop 1000. cRNA synthesis was done from 250 ng of RNA using the Illumina TotalPrep 96 RNA Amplification kit, and 750 ng of cRNA was applied to the Illumina Human HT-12 v4 expression array. Individuals having < 30% of probes with detection P-value < 0.05 were excluded from the analysis. We also excluded 1$^{st}$ degree relatives using GCTA software (–grm cut point of 0.3).

#### Methods: genotyping data

DNA was extracted from whole blood using the QIAamp 96 DNA Blood Kit (cat # 51161; Qiagen, Valencia, USA). Concentration and quality of extracted DNA were assessed using Nanodrop 1000. Genotyping was conducted using Illumina HumanCytoSNP-12 v2.1 chips according to Illumina's protocol, and chips were read on the BeadArray Reader. Image data was processed in BeadStudio software to generate genotype calls. QC was conducted as described previously (Pierce et al., 2012; Pierce et al., 2013). RNA was extracted from PBMCs, preserved in buffer RLT, and stored at −86°C using RNeasy Micro Kit (cat# 74004) from Qiagen.

#### Data availability statement

The data that support the findings of this study are available from the corresponding author upon reasonable request.

#### Acknowledgements

We would like to thank all BEST study participants and research staff.

#### Funding information

This study was funded by NIH grant number R01CA107431 (to HA), NIH grant number R01GM108711 (to LC), NIH grant numbers R35ES028379, R01ES023834, and R01ES020506 (to BP), and NIH grant number R21ES024834 (to BP and MA).

### KORA - batch 1

#### Cohort description

The KORA (Cooperative Health Research in the Region of Augsburg) study is a prospective population-based adult cohort study in the southeast of Germany. The study area covers the city of Augsburg and the two bordering districts of Augsburg and Aichach-Friedberg. Inclusion criteria were 25 to 74 years of age with main place of residence in the study area and German nationality. The data originates from the KORA FF4 study. The KORA FF4 study is the second follow-up of the KORA S4 study conducted between 1999 and 2001. A total sample of 6,640 individuals was drawn from the target population. Of all 4,261 participants in the S4 baseline study, 2,279 participated in the 14-year follow-up FF4 study. Participants were ineligible for FF4 if they had died in the meantime (n = 455, 10.7%), lived too far outside the study region or were completely lost to follow-up (n = 296, 6.9%), or had demanded deletion of their address data (n = 191, 4.5%). Of the remaining 3,319 eligible participants, 157 could not be contacted, 504 were unable to participate because they were too ill or had no time, and 379 were unwilling to participate in the follow-up, giving a response rate of 68.7%.

#### Ethics approval

The investigations were carried out in accordance with the Declaration of Helsinki, including written informed consent of all participants. All study methods of KORA FF4 were approved by the ethics committee of the Bavarian Chamber of Physicians, Munich (EC No. 06068).

#### Methods: expression data

RNA isolation was done using PAXgene Blood_RNA_Kit. RIN was measured using the Agilent 2100 Bioanalyzer system. RNA samples with RIN values of approximately 6 or more were selected for mRNA sequencing (poly-A selected). The libraries were prepared using the Illumina stranded mRNA prep ligation kit (Illumina), following the kit's instructions. The libraries were sequenced in a paired-end mode (2 x 100 bases) in the Novaseq6000 sequencer (Illumina) with a depth of ≥ 40 Million reads per sample. After demultiplexing, FASTQ files from each sample are processed using standard tools as part of an in-house pipeline.[1] Alignment to UCSC Genome Browser hg19 human reference genome was done using STAR v2.4.2a.[2] Unaligned reads were discarded. Sequencing QC was done using RNASeQC v1.1.8.1.[3] Properly aligned reads were then processed with HTSeq-count v0.6.185 to generate read counts. We used a count matrix of 22,073 genes and 1,232 participants.

#### Methods: genotyping data

Genome-wide SNP data were measured using an Affymetrix Axiom array. HapMap CEU build 37 was used for calling the SNPs. Samples with high levels of missing SNPs (individuals not having genotyping calls for at least 97% of the SNPs) were removed. Sex checks were made to correct for sample swaps by observing mismatches of phenotypic and genetic sex. These samples were removed. Samples with high or low heterozygosity rates were excluded (5*SD of mean heterozygosity rate). SNPs with low genotyping calls (SNPs not present in at least 97% of the samples) were removed. SNPs that deviated from HWE were excluded (P-value < $5x10^{-10}$). SNPs were imputed using the Michigan Imputation Server and minimac3 software. The HRC was used as a Reference panel. To reduce the burden of multiple testing and select only SNPs with high statistical power in the analysis, only SNPs with a MAF > 1% were considered.

#### Data availability statement

Data and biosamples can be requested by scientists for research projects by means of a project agreement via the KORA.PASST use and access hub (https://helmholtz-muenchen.managed-otrs.com/external). The KORA Board is responsible for review and approval of the requests. The rights of study participants, adherence to Good Scientific Practice, and the goals of the KORA study guide the decision. The European General Data Protection Regulation (GDPR) applies to all applicants.

#### Acknowledgements

We thank all participants for their long-term commitment to the KORA study, the staff for data collection and research data management, the members of the KORA Study Group who are responsible for the design and conduct of the study, and all the co-workers at Helmholtz Munich in the participating institutes and in the administration for their contributions and support.

#### Funding information

The KORA study is financed by the Helmholtz Zentrum München – German Research Center for Environmental Health, which is funded by the German Federal Ministry of Education and Research (BMBF) and by the State of Bavaria. Data collection in the KORA study is done in cooperation with the University Hospital of Augsburg. The expression data were funded by the Bavarian State Ministry of Health, Care and Prevention through the research project DigiMed Bayern (www.digimed-bayern.de).

### KORA - Batch 2

#### Cohort description

The KORA (Cooperative Health Research in the Region of Augsburg) study is a prospective population-based adult cohort study in the southeast of Germany. The study area covers the city of Augsburg and the two bordering districts of Augsburg and Aichach-Friedberg. Inclusion criteria were 25 to 74 years of age with main place of residence in the study area and German nationality. The data originates from the KORA FF4 study. The KORA FF4 study is the second follow-up of the KORA S4 study, conducted between 1999 and 2001. A total of 6,640 individuals were drawn from the target population. Of all 4,261 participants in the S4 baseline study, 2,279 participated in the 14-year follow-up FF4 study. Participants were considered ineligible for FF4 if they had died in the meantime (n = 455, 10.7%), lived too far outside the study region or were completely lost to follow-up (n = 296, 6.9%), or had demanded deletion of their address data (n = 191, 4.5%). Of the remaining 3,319 eligible participants, 157 could not be contacted, 504 were unable to participate because they were too ill or had no time, and 379 were unwilling to participate in the follow-up, giving a response rate of 68.7%.

#### Ethics approval

The investigations were carried out in accordance with the Declaration of Helsinki, including written informed consent of all participants. All study methods of KORA FF4 were approved by the ethics committee of the Bavarian Chamber of Physicians, Munich (EC No. 06068).

#### Methods: expression data

RNA isolation was done using PAXgene Blood_RNA_Kit. RIN was measured using the Agilent 2100 Bioanalyzer system. RNA samples with RIN values of approximately 6 or more were selected for mRNA sequencing (poly-A selected). The libraries were prepared using the Illumina stranded mRNA prep ligation kit (Illumina), following the kit's instructions. The libraries were sequenced in a paired-end mode (2x100 bases) in the Novaseq6000 sequencer (Illumina) with a depth of ≥ 40 Million reads per sample. After demultiplexing, FASTQ files from each sample are processed using standard tools as part of an in-house pipeline.[1] Alignment to UCSC Genome Browser hg19 human reference genome using STAR v2.4.2a.[2] Unaligned reads were discarded. Sequencing QC was done using RNASeQC v1.1.8.1.[3] Properly aligned reads were then processed with HTSeq-count v0.6.185 to generate read counts. We used a count matrix of 22,073 genes and 635 participants.

#### Methods: genotyping data

Genome-wide SNP data were measured using an Affymetrix Axiom array. HapMap CEU build 37 was used for calling the SNPs. Samples with high levels of missing SNPs (individuals not having genotyping calls for at least 97% of the SNPs) were removed. Sex checks were made to correct for sample swaps by observing mismatches of phenotypic and genetic sex. These samples were removed. Samples with high or low heterozygosity rates were excluded (5*SD of mean heterozygosity rate). SNPs with low genotyping calls (SNPs not present in at least 97% of the samples) were removed. SNPs that deviated from HWE were excluded (P-value $< 5x10^{-10}$). SNPs were imputed using the Michigan Imputation Server and minimac3 software. The HRC was used as a Reference panel. To reduce the burden of multiple testing and select only SNPs with high statistical power in the analysis, only SNPs with a MAF > 1% were considered.

#### Data availability statement

Data and biosamples can be requested by scientists for research projects by means of a project agreement via the KORA.PASST use and access hub (https://helmholtz-muenchen.managed-otrs.com/external). The KORA Board is responsible for review and approval of the requests. The rights of study participants, adherence to Good Scientific Practice, and the goals of the KORA study guide the decision. The European GDPR applies to all applicants.


#### Acknowledgements

We thank all participants for their long-term commitment to the KORA study, the staff for data collection and research data management, the members of the KORA Study Group who are responsible for the design and conduct of the study, and all the co-workers at Helmholtz Munich in the participating institutes and in the administration for their contributions and support.

#### Funding information

The KORA study is financed by the Helmholtz Zentrum München – German Research Center for Environmental Health, which is funded by the German Federal Ministry of Education and Research (BMBF) and by the State of Bavaria. Data collection in the KORA study is done in cooperation with the University Hospital of Augsburg. The expression data were funded by the Bavarian State Ministry of Health, Care and Prevention through the research project DigiMed Bayern (www.digimed-bayern.de).

### InCHIANTI

#### Cohort description

We used individuals from the InCHIANTI study, a study of aging from the Chianti region in Tuscany, Italy.

#### Ethics approval

InCHIANTI data collection was approved by the Ethical Committee at INRCA, Ancona (protocol 14/CE, 28 February 2000) and FU1 (protocol 45/01, 16 January 2001), and participants provided written informed consent. Secondary analysis was approved by the University of Sherbrooke Ethics Board (ref. 2019-2657) and Columbia University Medical Center’s IRB (no. AAAU5622).

#### Methods: expression data

Peripheral blood specimens were collected from 712 individuals using the PAXgene tube technology to preserve levels of mRNA transcripts. RNA was extracted from peripheral blood samples using the PAXgene Blood mRNA kit (Qiagen, Crawley, UK), according to the manufacturer's instructions. RNA was biotinylated and amplified using the Illumina® TotalPrep(tm) -96 RNA Amplification Kit and directly hybed with HumanHT-12_v3 Expression BeadChips that include 48,803 probes. Image data were collected on an Illumina iScan and analyzed using Illumina GenomeStudio software.

#### Methods: genotyping data

Genome-wide genotyping was performed using the Illumina Infinium HumanHap550 genotyping chip. Standard QC procedures were used to filter out SNPs with a MAF <1%, HWE $P < 1 \times 10^{-4}$, and a call rate < 99%.

#### Data availability statement

Data from the InCHIANTI study are available from the GEO database (GSE 48152).


#### Acknowledgements

#### Funding information

The InCHIANTI study baseline (1998-2000) was supported as a "targeted project" (ICS110.1/RF97.71) by the Italian Ministry of Health and in part by the U.S. National Institute on Aging (Contracts: 263 MD 9164 and 263 MD 821336). The InCHIANTI Follow-up 1 (2001-2003) was funded by the U.S. National Institute on Aging (Contracts: N.1-AG-1-1 and N.1-AG-1-2111)

### DGN

#### Cohort description

The Depression Genes and Networks (DGN) study includes genotyping and gene expression data from 922 European individuals (463 cases of Major Depressive Disorder (MDD) and 459 controls) aged 21 to 60 years, recruited through a survey company called Knowledge Networks Inc (KN) for the Depression Susceptibility Genes and Networks Project (NIMH Grant 5RC2MH089916). Through a process involving an online questionnaire (relevant sections of CIDI-SF), KN identified potential candidates for this study. From this pool, 1,259 individuals went on to have their blood drawn, filled out consent forms, and were telephone interviewed (SCID interview − Structured Clinical Interview for DSM-IV covering depressive, bipolar, psychotic, alcohol, substance, and anxiety disorders, as well as family history of mood disorders). After excluding some eligible individuals for reported non-European ancestry or medical comorbidities, which were thought to be too unusual and significant for the gene expression analysis, and performing QC, 463 cases of MDD and 459 control individuals were analyzed. Sample collection, QC, and data processing are described in detail in Battle et al.[1].

#### Ethics approval

The research activities involving participant data and recruitment for the DGN study were conducted under a protocol approved by the Stanford IRB.

#### Methods: expression data

RNA was extracted from whole blood, and hemoglobin RNA was removed from each sample using GLOBINclearTMKit (Invitrogen). RNA-sequencing was performed using Illumina HiSeq 2000 (50 bp single-ended reads) following the Illumina TruSeq RNA protocol. Reads were aligned to the NCBI v37 human reference genome using TopHat. Gene expression was quantified by HTSeq using uniquely mapped reads. Sample collection, QC, and data processing are described in detail in Battle et al. (2014).

#### Methods: genotyping data

DNA was extracted and genotyped on the Illumina HumanOmni1-Quad BeadChip. QC was performed to identify samples with elevated heterozygosity, unexpected ancestry or pairwise IBD, and potential mislabeling.
Genotype data was filtered for genotype quality as follows. Pairwise estimates of IBD were computed in PLINK, and any sample duplicates were excluded. PCA was carried out for all individuals using every fifth autosomal SNP (to reduce the influence of LD among SNPs), and the principal component scores were examined for relationship to the self reported geographical and ancestral origins. Additionally, samples were excluded for elevated rates of heterozygosity (> 34.5% of SNPs) or if genotypes could not be called for > 1.4% of SNPs. For SNPs, we evaluated QC metrics in each study and retained SNPs with a missingness rate < 0.012, a 10% Gencall score > 0.55, and HWE < 0.0001. Genotype data was further quality-controlled, imputed, and processed by eQTLGen pipelines in accordance with the eQTLGen cookbook. After preprocessing and QC, 865 samples were added to eQTLGen analyses.

#### Data availability statement

Genotype, raw RNA-seq, quantified expression, and covariate data are available by application through the NIMH Center for Collaborative Genomic Studies on Mental Disorders. Instructions for requesting access to data can be found at:

https://www.nimhgenetics.org/access_data_biomaterial.php, and inquiries should reference the "Depression Genes and Networks study (D. Levinson, PI)."

#### Acknowledgements

For the DGN data, we gratefully acknowledge the resources supported by NIH/NIMH grants 5RC2MH089916 (PI: Douglas F. Levinson, M.D.; Co-investigators: Myrna M. Weissman, Ph.D., James B. Potash, M.D., MPH, Daphne Koller, Ph.D., and Alexander E. Urban, Ph.D.) and 3R01MH090941 (Co-investigator: Daphne Koller, Ph.D.)

#### Funding information

This work was originally supported by the NIMH Grants RC2MH089916 and R01MH090941 for the initial DGN work and is currently funded by the NIH/NIGMS Award R35GM139580 Grant for the ongoing eQTLGen analyses of DGN data.

### BSGS

#### Cohort description

The Brisbane Systems Genetics Study (BSGS) cohort was previously described in detail in (Powell et al., 2012; Powell et al., 2013)[1,2]. Briefly, BSGS is comprised of 862 individuals of Northern European origin from 274 families consisting of either monozygotic or dizygotic twin pairs, along with their siblings and parents. Here, we selected only unrelated individuals, leaving 299 for analysis by the eQTLGen analysis plan for Illumina arrays.

#### Ethics approval

All participants gave informed consent, and the study protocol was approved by the appropriate IRBs.

#### Methods: expression data

Expression levels for each individual were measured from whole blood using Illumina HT-12 v4.0 microarray chips.

#### Methods: genotyping data

Whole-genome SNP genotypes were generated using Illumina 610 Quad-Beadchips and, after QC, were imputed to the 1000 Genomes Release.

#### Data availability statement

The expression dataset is available from the GEO public repository under the accession GSE 33321.


#### Acknowledgements

We gratefully acknowledge the participation of the individuals sampled in this work.

#### Funding information

This research was supported by Australian NHMRC grants 389892, 496667, 613601, 1010374, and 1046880 and by NIH grant GM057091.

### Consortium Banner Authors

#### Estonian Biobank research team

Andres Metspalu[1], Lili Milani[1], Tõnu Esko[1], Reedik Mägi[1], Mait Metspalu[1], Mari Nelis[1], Georgi Hudjashov[1]

1. Estonian Genome Centre, Institute of Genomics, University of Tartu

#### PRECISEADS Clinical Consortium

Lorenzo Beretta[1], Barbara Vigone[1], Jacques-Olivier Pers[2], Alain Saraux[2], Valérie Devauchelle-Pensec[2], Divi Cornec[2], Sandrine Jousse-Joulin[2], Bernard Lauwerys[3], Julie Ducreux[3], Anne-Lise Maudoux[3], Carlos Vasconcelos[4], Ana Tavares[4], Esmeralda Neves[4], Raquel Faria[4], Mariana Brandão[4], Ana Campar[4], António Marinho[4], Fátima Farinha[4], Isabel Almeida[4], Miguel Angel Gonzalez-Gay[5], Ricardo Blanco Alonso[5], Alfonso Corrales Martínez[5],Ricard Cervera[6], Ignasi Rodríguez-Pintó[6], Gerard Espinosa[6],Rik Lories[7], Ellen De Langhe[7],Nicolas Hunzelmann[8], Doreen Belz[8],Torsten Witte[9], Niklas Baerlecken[9],Georg Stummvoll[10], Michael Zauner[10], Michaela Lehner[10],Eduardo Collantes[11], Rafaela Ortega-Castro[11], Mª Angeles Aguirre-Zamorano[11], Alejandro Escudero-Contreras[11], Mª Carmen Castro-Villegas[11], Yolanda Jiménez Gómez[11], Norberto Ortego[12], María Concepción Fernández Roldán[12], Enrique Raya[13], Inmaculada Jiménez Moleón[13], Enrique de Ramon[14], Isabel Díaz Quintero, Pier Luigi Meroni[15], Maria Gerosa[15], Tommaso Schioppo[15], Carolina Artusi[15],Carlo Chizzolini[16], Aleksandra Dufour[16], Donatienne Wynar ,Laszló Kovács[17], Attila Balog[17], Magdolna Deák[17], Márta Bocskai[17], Sonja Dulic[17], Gabriella Kádár[17],Falk Hiepe[18], Velia Gerl[18], Silvia Thiel[18],Manuel Rodriguez Maresca[19], Antonio López-Berrio[19], Rocío Aguilar-Quesada[19], Héctor Navarro-Linares[19], Yiannis Ioannou[20], Chris Chamberlain[20], Jacqueline Marovac[20], Marta Alarcón Riquelme[21], Lorena Jiménez Jaén[21].

Funding: This work has received support from the EU/EFPIA Innovative Medicines Initiative Joint Undertaking (PRECISESADS, grant n. 115565) including in-kind contributions from the EFPIA members involved.

1. Referral Center for Systemic Autoimmune Diseases, Fondazione IRCCS Ca' Granda Ospedale Maggiore Policlinico di Milano, Italy.
2. Centre Hospitalier Universitaire de Brest, Hospital de la Cavale Blanche, Brest, France.
3. Pôle de pathologies rhumatismales systémiques et inflammatoires, Institut de Recherche Expérimentale et Clinique, Université catholique de Louvain, Brussels, Belgium.
4. Centro Hospitalar do Porto, Portugal.
5. Hospital Universitario Marqués de Valdecilla, IDIVAL, Universidad de Cantabria, Santander, Spain.
6. Hospital Clinic I Provicia, Institut d'Investigacions Biomèdiques August Pi i Sunyer, Barcelona, Spain.
7. Katholieke Universiteit Leuven, Belgium.
8. Klinikum der Universitaet zu Koeln, Cologne, Germany.
9. Medizinische Hochschule Hannover, Germany.
10. Medical University Vienna, Vienna, Austria.
11. Servicio Andaluz de Salud, Hospital Universitario Reina Sofía Córdoba, Spain.
12. Servicio Andaluz de Salud, Complejo hospitalario Universitario de Granada (Hospital Universitario San Cecilio), Spain.
13. Servicio Andaluz de Salud, Complejo hospitalario Universitario de Granada (Hospital Virgen de las Nieves), Spain.

14. Servicio Andaluz de Salud, Hospital Regional Universitario de Málaga, Spain
15. Università degli studi di Milano, Milan, Italy.
16. Hospitaux Universitaires de Genève, Switzerland.
17. University of Szeged, Szeged, Hungary.
18. Charite, Berlin, Germany.
19. Andalusian Public Health System Biobank, Granada, Spain
20. UCB Pharma, Slough, United Kingdom (PRECISESADS Project office)
21. Department of Medical Genomics, Center for Genomics and Oncological Research (GENYO), Granada, Spain (PRECISESADS Project Office)

### IMI DIRECT Consortium

Leen 't Hart[1,2,3], Jonathan Adam[4,5], Jerzy Adamski[6,7,8], Kristine H. Allin[9], Anna A. Artati[5], Naeimeh Atabaki-Pasdar[10], Karina Banasik[11], Jimmy D. Bell[12], Joline W. Beulens[1], Susanna B. Bianzano[13], Roberto Bizzotto[14], Marieke Blom[15], Amelie Bonnefond[16], Caroline A. Brorsson[11,17], Andrew A. Brown[18], Søren Brunak[11,17], Marc Clos-Garcia[19], David Davtian[18], Federico De Masi[11], Emmanouil T. Dermitzakis[20,21,22], Christiane Dings[23], Théo Dupuis[18], Petra J.M. Elders[15], Line Engelbrechtsen[9], Rebeca Eriksen[24], Massimo Faggiani[14], Yong Fan[9], Juan J. Fernandez-Tajes[25], Jorge Ferrer[26,27], Ian M. Forgie[18], Paul W. Franks[28], Tim Frayling[20], Andreas Fritsche[29], Philippe Froguel[16], Gary Frost[24], Johann Gassenhuber[30], Giuseppe N. Giordano[28], Harald Grallert[4,5], Lenka Groeneveld[1], Valborg Gudmundsdóttir[11], Ramneek Gupta[31], Mark Haid[32], Torben Hansen[9], Tue H. Hansen[9], Peter Harms[15], Andrew T. Hattersley[33], Anita M.H. Hennige[34], Anita V. Hill[35], Reinhard W. Holl[36], Mun-gwan Hong[37], Michelle Hudson[35], Bernd Jablonka[30], Ulrik Plesner Jacobsen[11], Christopher Jennison[38], Angus G. Jones[33], Tugce Karaderi[11], Jane Kaye[39], Robert W. Koivula[10], Tarja Kokkola[40], Teemu Kuulasmaa[41], Markku Laakso[40], Thorsten Lehr[23], Agnete Troen T. Lundgaard[11], Liwei Lyu[31], Anubha Mahajan[25], Andrea Mari[14], Gianluca Mazzoni[11], Mark I McCarthy[25,42], Timothy J. McDonald[43], Nicky McRobert[10], Theodora-Dafni Michalettou[44], Petra B. Musholt[45], Rachel Nice[43], Colin N. Palmer[18], Francois Pattou[16], Imre Pavo[46], Ewan R. Pearson[47], Oluf Pedersen[19,31], Helle K Pedersen[11], Cornelia P Prehn[32], Anna Ramisch[20], Simon Rasmussen[11], Violeta Raverdy[16], Carlo Alberto Rossi[14], Hartmut Ruetten[30], Femke Rutters[1], Jochen M. Schwenk[37], Sapna Sharma[5,48], Iryna Sihinevich[23], Melissa K Thomas[49], Cecilia Engel E Thomas[31], Elizabeth Louise L Thomas[12], Barbara Thorand[4,5], Andrea Tura[14], Mathias Uhlen[50], Jagadish Vangipurapu[40], Henrik Vestergaard[9,51], Ana Viñuela[18], Josef Vogt[9], Mark Walker[52], Agata Wesolowska-Andersen[10]

[1]Epidemiology and Data Science, VUMC, Amsterdam, The Netherlands, [2]Department of Cell and Chemical Biology, Leiden University Medical Center, Leiden, The Netherlands, [3]Department of Biomedical Data Sciences, Molecular Epidemiology section, Leiden University Medical Center, Leiden, The Netherlands, [4]German Center for Diabetes Research (DZD), Neuherberg, 85764, Germany, [5]Research Unit of Molecular Epidemiology, Institute of Epidemiology, German Research Center for Environmental Health, Helmholtz Zentrum München, Neuherberg, 85764, Germany, [6]Department of Biochemistry, Yong Loo Lin School of Medicine, National University of Singapore, Singapore, 117597, Singapore, [7]Institute of Experimental Genetics, German Research Center for Environmental Health, Helmholtz Zentrum München, Neuherberg, 85764, Germany, [8]Institute of Biochemistry, Faculty of Medicine, University of Ljubljana, Ljubljana, Slovenia, [9]The Novo Nordisk Center for Basic Metabolic Research, Faculty of Health and Medical Science, University of Copenhagen, Copenhagen, DK-2100, Denmark, [10]Oxford Centre for Diabetes Endocrinology and Metabolism, University of Oxford, Oxford, OX3 7LJ, United Kingdom, [11]Department of Health Technology, Technical University of Denmark, Kongens Lyngby, Denmark, [12]Research Centre for Optimal Health, School of Life Sciences, University of Westminster, London, United Kingdom, [13]Therapeutic Area CardioMetabolism and Respiratory Medicine, Boehringer Ingelheim International GmbH, Biberach an der Riss, 88397, Germany, [14]Institute of Neuroscience, National Research Council, Padova, 35127, Italy, [15]Department of General Practice, Amsterdam UMC- location Vumc,

Amsterdam Public Health research institute, Amsterdam, The Netherlands, [16]University of Lille, Inserm, Lille Pasteur Institute, Lille, France, [17]Novo Nordisk Foundation Center for Protein Research, Faculty of Health and Medical Sciences, University of Copenhagen, Copenhagen, DK-2100, Denmark, [18]Population Health and Genomics, Ninewells Hospital and Medical School, University of Dundee, Dundee, DD1 9SY, United Kingdom, [19]Center for Clinical Metabolic Research, Herlev and Gentofte University Hospital, Copenhagen, Denmark, [20]Department of Genetic Medicine and Development, University of Geneva Medical School, Geneva, 1211, Switzerland, [21]Institute for Genetics and Genomics in Geneva (iGE3), University of Geneva, Geneva, 1211, Switzerland, [22]Swiss Institute of Bioinformatics, Geneva, 1211, Switzerland, [23]Clinical Pharmacy, Saarland University, Saarbrücken, 66123, Germany, [24]Nutrition and Dietetics Research Group, Imperial College London, London, SW7 2AZ, United Kingdom, [25]Wellcome Trust Centre for Human Genetics, University of Oxford, Oxford, OX3 7BN, United Kingdom, [26]Department of Metabolism, Digestion and Reproduction, Imperial College London, London, United Kingdom, [27]Regulatory genomics and diabetes, Centre for Genomic Regulation, Barcelona, Spain, [28]Department of Clinical Science, Genetic and Molecular Epidemiology, Lund University Diabetes Centre, Malmö, Sweden, [29]Medizinische Universitätsklinik Tübingen, Eberhard Karls Universität Tübingen, Tübingen, Germany, [30]Sanofi Partnering, Sanofi-Aventis Deutschland GmbH, Frankfurt am Main, 65926, Germany, [31]Novo Nordisk Foundation Center for Basic Metabolic Research, Faculty of Health and Medical Sciences, University of Copenhagen, Copenhagen, DK-2100, Denmark, [32]Metabolomics and Proteomics Core, German Research Center for Environmental Health, Helmholtz Zentrum München, Neuherberg, 85764, Germany, [33]Department of Clinical and Biomedical Sciences, University of Exeter College of Medicine & Health, Exeter, EX25DW, United Kingdom, [34]Boehringer Ingelheim International GmbH, Biberach an der Riss, 88397, Germany, [35]NIHR Exeter Clinical Research Facility, Royal Devon and Exeter NHS Foundation Trust, Exeter, United Kingdom, [36]Institute for Epidemiology and medical Biometry, University of Ulm, Ulm, Germany, [37]Science for Life Laboratory, School of Biotechnology, KTH - Royal Institute of Technology, Solna, SE-171 21, Sweden, [38]Department of Mathematical Sciences, University of Bath, Bath, United Kingdom, [39]Nuffield Department of Population Health, Centre for Health, Law and Emerging Technologies (HeLEX), University of Oxford, Oxford, OX2 7DD, United Kingdom, [40]Internal Medicine, Institute of Clinical Medicine, University of Eastern Finland, Kuopio, Finland, [41]Institute of Biomedicine, Bioinformatics Center, University of Eastern Finland, Kuopio, Finland, [42]Current address:, GENENTECH, 1 DNA Way, San Francisco, CA 94080, United States, [43]Blood Sciences, Royal Devon and Exeter NHS Foundation Trust, Exeter, EX2 5DW, United Kingdom, [44]Biosciences Institute, Faculty of Medical Sciences, University of Newcastle, Newcastle upon Tyne, NE1 4EP, United Kingdom, [45]Global Development, Sanofi-Aventis Deutschland GmbH, Hoechst Industrial Park, Frankfurt am Main, 65926, Germany, [46]Eli Lilly Regional Operations Ges.m.b.H., Vienna, 1030, Austria, [47]Diabetes Endocrinology and Reproductive Biology, Ninewells Hospital and Medical School, University of Dundee, Dundee, DD1 9SY, United Kingdom, [48]Food Chemistry and Molecular and Sensory Science, Technical University of Munich, München, Germany, [49]Lilly Research Laboratories, Eli Lilly and Company, Indianapolis, USA, [50]Science for Life Laboratory, School of Biotechnology, KTH - Royal Institute of Technology, Solna, [51]Steno Diabetes Center Copenhagen, Copenhagen, Denmark, [52]Translational and Clinical Research Institute, Faculty of Medical Sciences, University of Newcastle, Newcastle upon Tyne, United Kingdom

### sc-eQTLGen Consortium

Daniel Kaptijn[1,2], Lieke Michielsen[3,4], Drew Neavin[5,6,7], Aida Ripoll-Cladellas[8,9], José Alquicira-Hernández[5,10], Maryna Korshevniuk[1,2], Jimmy Tsz Hang Lee[11], Roy Oelen[1,2], Martijn Vochteloo[1,2], Robert Warmerdam[1,2], Yoshinari Ando[12], Odmaa Bayaraa[13], Maria Ban[14], Marijn Berg[15,16], Irene van Blokland[1,17], Daniel Considine[11,18], Mame M. Dieng[13], Ryuya Edahiro[19,20], M. Grace Gordon[21,22,23,24], Hilde E. Groot[17], Pim van der Harst[25], Matthias Heinig[26,27], Chung-Chau Hon[12], Youssef Idaghdour[13], Pooja Kathail[28], Niek de Klein[11], Wenchao Li[29], Yang Li[29,30], Corinna Losert[26,27], Vinu Manikanda[13], Jonathan Moody[12], Martijn C. Nawijn[15,16], Mihai Netea[30], Jelmer Niewold[1,2], Yukinori Okada[24,25],

Stephen Sawcer[14], Issiaka Soulama[31], Oliver Stegle[11,32,33], Yakov Tsepilov[11,18], Woong-Yang Park[34], Deepa Rajagopalan[35], Tala Shahin[13], Jay W. Shin[35], Gosia Trynka[11,18], Harm-Jan Westra[1,2], Seyhan Yazar[8], Jimmie Ye[22,23,36,37,38,39], zhenhua zhang[29], Martin Hemberg[40], Ahmed Mahfouz[3,4], Marta Melé[10], Joseph E. Powell[5,6,41], Lude Franke[1,2], Monique G.P. van der Wijst[1,2], Marc Jan Bonder[1,2]

1. Department of Genetics, University of Groningen, University Medical Center Groningen.
2. Oncode Institute, Groningen, the Netherlands.
3. Department of Human Genetics, Leiden University Medical Center, Leiden, The Netherlands.
4. Delft Bioinformatics Laboratory, Delft University of Technology, Delft, The Netherlands
5. Translational Genomics Program, Garvan Institute of Medical Research, Darlinghurst, Sydney, NSW, Australia
6. University of New South Wales, Kensington, Sydney, NSW, Australia
7. Current affiliation: Institute for Molecular Bioscience, University of Queensland, Brisbane, QLD, Australia
8. Life Sciences Department, Barcelona Supercomputing Center, Barcelona, Catalonia, Spain
9. Universitat de Barcelona, Barcelona, Spain
10. Computational Genomics, Institute for Molecular Bioscience, University of Queensland, Brisbane, Australia
11. Wellcome Sanger Institute, Wellcome Genome Campus, Cambridge, UK.
12. Laboratory for Genome Information Analysis, RIKEN Center for Integrative Medical Sciences, Japan.
13. Biology Program, New York University Abu Dhabi, United Arab Emirates.
14. University of Cambridge, Department of Clinical Neurosciences, Cambridge Biomedical Campus, Cambridge, CB2 0QQ, UK
15. Department of Pathology and Medical Biology, University of Groningen, University Medical Center Groningen, Groningen, the Netherlands.
16. GRIAC research institute, University Medical Center Groningen, Groningen, the Netherlands.
17. Department of Cardiology, University of Groningen, University Medical Center Groningen, Groningen, the Netherlands.
18. Open Targets, Wellcome Genome Campus, Hinxton, Cambridgeshire
19. Department of Statistical Genetics, Osaka University Graduate School of Medicine.
20. Laboratory for Systems Genetics, RIKEN Center for Integrative Medical Sciences.
21. Biological and Medical Informatics Graduate Program, University of California San Francisco, San Francisco, USA.
22. UCSF Division of Rheumatology, Department of Medicine, University of California San Francisco, San Francisco, CA, USA.
23. Institute for Human Genetics, University of California San Francisco, USA.
24. Department of Bioengineering and Therapeutic Sciences, University of California San Francisco, San Francisco, USA.
25. Department of Cardiology, University Medical Center Utrecht, Utrecht, the Netherlands.
26. Institute of Computational Biology, German Research Center for Environmental Health, Helmholtz Zentrum München, Neuherberg, Germany
27. Department of Computer Science, TUM School of Computation, Information and Technology, Technical University of Munich, Garching, Germany 5. Open Targets, Wellcome Genome Campus, Hinxton, Cambridges
28. Department of Electrical Engineering and Computer Science, Center for Computational Biology, University of California Berkeley, Berkeley, CA, USA
29. Centre for Individualised Infection Medicine (CiiM) & TWINCORE, joint ventures between the Helmholtz-Centre for Infection Research (HZI) and the Hannover Medical School (MHH), Hannover, Germany
30. Department of Internal Medicine and Radboud Center for Infectious Diseases, Radboud University Nijmegen Medical Center, Nijmegen, Netherlands

31. Institut de Recherche en Sciences de la Santé, Ouagadougou, Burkina Faso
32. Division of Computational Genomics and Systems Genetics, German Cancer Research Center (DKFZ), Heidelberg, Germany.
33. European Molecular Biology Laboratory (EMBL), Genome Biology Unit, Heidelberg, Germany.
34. Samsung Genome Institute, Samsung Medical Center, Seoul, Korea.
35. A*STAR Genome Institute of Singapore, Singapore.
36. Bakar Computational Health Sciences Institute, University of California San Francisco, San Francisco, CA, USA.
37. Department of Epidemiology and Biostatistics, University of California San Francisco, San Francisco, CA, USA.
38. Parker Institute for Cancer Immunotherapy, San Francisco, CA, USA.
39. Chan Zuckerberg Biohub, San Francisco, CA, USA
40. The Gene Lay Institute of Immunology and Inflammation, Brigham and Women's Hospital, Massachusetts General Hospital, and Harvard Medical School, Boston, USA
41. UNSW Cellular Genomics Futures Institute, University of New South Wales, Sydney, Australia.

### The HELIOS Study Team

John C. Chambers[1,2,3], Marie Loh[1,4,2,5], Paul Eillott[2], Eng Sing Lee[6,1], Jimmy Lee[7,8,1], Joanne Ngeow[1,9], Sabrina Wong[10,11], Elio Riboli[2], Tricia Chang[10], Rinkoo Dalan[12,1], Wai Kee Kok[10], Benjamin Lam[13,1,14], Kelvin Li[12], Tock Han Lim[12], Pritesh R. Jain[1], Hong Kiat Ng[1], Theresia Mina[1], Nilanjana Sadhu[1], Akash Bahai[1], Dorrain Low[1], Xiaoyan Wang[1], Harinakshi Sanikini[2], Darwin Tay[1], Terry Tong[1], Kostas Tsilidis[2], Wansaicheong Khin-lin, Gervais Tsilidis[12,1], Yik Weng Yew[5,1]

1. Nanyang Technological University, Lee Kong Chian School of Medicine, Singapore
2. Imperial College London, School of Public Health, London, UK
3. Precision Health Research, Singapore
4. Genome Institute of Singapore, Agency for Science, Technology and Research, Singapore
5. National Skin Centre, Singapore
6. Ministry of Health Office for Healthcare Transformation, Singapore
7. National Healthcare Group, Singapore
8. Institute of Mental Health, Singapore
9. National Cancer Centre, Singapore
10. National Healthcare Group Polyclinics, Singapore
11. Nanyang Technological University, Singapore
12. Tan Tock Seng Hospital, Singapore
13. Khoo Teck Puat Hospital, Singapore
14. Changi General Hospital, Singapore
